# Effects of online professional learning on healthcare professionals’ knowledge and skill acquisition: A systematic review and meta-analysis

**DOI:** 10.64898/2026.04.27.26351794

**Authors:** Shantell Griffith, Ghassani Swaryandini, Lachlan McKee, Kate Oxnard, Liana S. Cahill, Hannah Forbes, Kathy Rees, Jade Davis, Taren Sanders, James A. Coleman, Jessica Graham, Sandy Middleton, Dominique A. Cadilhac, Simeon Dale, Oyebola Fasugba, Michael Noetel

## Abstract

**Background:** Online professional learning offers a scalable alternative to traditional face-to-face learning, but there are doubts regarding how well it works and when it works best. This review assessed the effectiveness of online professional learning interventions on healthcare professionals’ knowledge and skill acquisition.

**Methods:** We conducted a systematic review and meta-analysis of randomised controlled trials that compared online professional learning against static controls or face-to-face controls. We searched MEDLINE Complete, Scopus, Embase, CENTRAL, and PsycINFO from inception to January 31, 2025. Eligible studies included practising healthcare professionals in any clinical setting that measured knowledge or skill acquisition related to patient care. Data was extracted in duplicate, with disagreements resolved through discussion or by a third reviewer. We used multilevel meta-analyses to estimate the overall effect size and conducted moderation analyses for pre-specified factors. The study protocol was pre-registered on the Open Science Framework (OSF; https://osf.io/46zav).

**Findings:** Of 55,376 records; 171 studies (391 effects, 25,412 participants) met the inclusion criteria. Online learning significantly improved knowledge and skill acquisition compared to static controls (*g* = 0.93, 95% CI [0.78,1.07], p < 0.001; *I*² = 89.8%), with larger effects in lower-middle income countries (*g* = 1.30, 95% CI [0.88, 1.72]) than in high income (*g* = 0.75, 95% CI [0.63, 0.86]). Online learning also significantly improves outcomes compared to face-to-face instruction (*g* = 0.45, 95% CI [0.31,0.59], *p* < 0.001; *I²* = 85.92%), with larger effects for knowledge outcomes (*g* = 0.46, 95% CI [0.33, 0.59]) than skills outcomes (*g* = 0.20, 95% CI [0.04, 0.36]). Effects did not differ significantly by profession, clinician experience, clinical setting, intervention characteristics or the learning design features (all *p* > 0.05). No studies had low overall risk of bias, and some evidence of publication bias was found.

**Interpretation:** From this body of evidence, we identified that online learning appears to improve healthcare professionals’ knowledge and skill acquisition, exceeding traditional teaching methods. Healthcare organisations can be confident implementing or expanding online professional learning to improve knowledge and skill acquisition.

**Funding:** No funding

---

Knowledge translation aims to accelerate the implementation of research into clinical practice. It can be a slow process; research can take an average of 17 years to reach widespread clinical practice^1–3^, leading to preventable deaths^4,5^ and disability^6,7^. To close this research-practice gap, continuing professional learning aims to provide healthcare professionals with up-to-date knowledge and skills^8–10^. Traditionally, face-to-face education has been shown to develop practitioners’ knowledge and skills^11,12^. However, face-to-face programs often require significant resources, incur high costs, and present scalability challenges^2,13,14^. As a result, many healthcare professionals, especially those in remote or low-resource areas, struggle to access professional learning opportunities^15–19^. Online learning has emerged as a flexible and cost-effective alternative that allows professionals to access materials at their own pace, regardless of time or location^20–23^.

Despite widespread adoption^2,24^, evidence regarding the effectiveness of online learning remains mixed. For instance, Ding et al.^25^ conducted a meta-analysis of e-learning programs for nurse education on pressure injury management, reporting significant knowledge gains compared to traditional approaches. In contrast, Vaona et al.’s^14^ meta-analysis involving licensed healthcare professionals found online learning made little to no difference in knowledge acquisition compared to traditional professional development methods. Rouleau et al.’s^26^ overview of systematic reviews of nursing continuing education found that while online learning was effective when compared to no intervention, traditional classroom learning was superior in certain clinical skills, particularly drug dose practices. Overall, the variability in findings underscores the need to better understand when and how online learning can be most effective compared to traditional methods.

Whether online learning is effective or not may depend on the delivery format and design features, such as the level of interactivity^27^, blended learning, collaboration^28,29^, or feedback^21,30^. Yet few reviews have examined how these principles influence knowledge and skills outcomes across healthcare professionals. The aim of our systematic review was to assess the effectiveness of online professional learning interventions on healthcare professionals’ knowledge and skill acquisition, and investigate a broad range of online learning design features to determine which, if any, influence knowledge and skills outcomes. Understanding these elements will enable development of evidence-based interventions to effectively bridge the research-practice gap.

## Methods

We conducted a systematic review and meta-analysis following Preferred Reporting Items for Systematic Review and Meta-Analysis guidelines (see Supplementary File 1 for PRISMA 2020 checklist)^31^, the Assessing the Methodological Quality of Systematic Review (AMSTAR) 2 tool^32^, and pre-registered our protocol on the Open Science Framework (OSF; https://osf.io/46zav). This report focuses on aims two and three of the protocol, examining outcomes related to knowledge and skill acquisition, while a complementary paper addresses aims one and four^33^.

## Eligibility Criteria

We included randomised controlled trials of practising healthcare professionals comparing online professional learning with any other professional learning or a control condition whereby the outcomes reported were specific to knowledge or skill development as relevant to patient care. Studies in any language were eligible; non-English reports were translated using Google Translate. Studies needed to present new data (e.g., protocols and reviews were excluded) in a form that could be used to calculate an effect size (e.g., means, standard deviations, number of events/successes, precise p values). Where data required for effect-size calculation were missing, we contacted study authors and excluded studies if data could not be obtained. Full eligibility criteria are provided in the supplementary materials (see Supplementary File 2).

### Search strategy and selection criteria

Our search strategy development began with analysing key terms from an initial sample of relevant papers, including systematic reviews ^e.g.,14,34^ and primary studies ^e.g.,35,36^. We compiled a list of terms to accurately identify target papers^37^, achieving high sensitivity and specificity by successfully capturing all 28 identified primary studies.

We used those terms to search MEDLINE Complete, Scopus, Embase (Ovid), CENTRAL, and PsycINFO from inception to Jan 31, 2025; full strategies are provided in Supplementary File 3. After de-duplication in EndNote, title/abstract records were processed using RobotSearch^38^ to assist identification of randomised controlled trials^39^ and then imported into Covidence for screening. Two reviewers screened records independently at title/abstract and full-text stages, resolving disagreements through discussion or adjudication by a third reviewer (GS, SG, or MN). Forward and backward citation searches were conducted in Scopus on July 19, 2025 (by SG) and screened in duplicate (SG, GS, and KR), with disagreements resolved by discussion.

### Data Analysis

#### Data Items

We extracted study characteristics and outcome data required to calculate effect sizes (as defined in the protocol), independently and in duplicate, with disagreements resolved through discussion or by a third reviewer (SG, GS or MN). When studies presented alternative data in formats that did not directly provide effect sizes (e.g., interquartile range instead of standard deviations), we calculated the necessary values following guidelines from the Cochrane Handbook for Systematic Reviews of Interventions^40^. When studies provided only figures, we extracted the relevant data using WebPlotDigitizer^41^.

#### Risk of Bias

We a priori planned that risk of bias assessments would be conducted independently and in duplicate. Given the high volume of included studies, we modified this process following guidance from AMSTAR 2^32^ and the Cochrane Handbook^40^ to maintain reliability. We assessed methodological quality of the included randomised studies using the Cochrane Risk of Bias 2 (RoB2), we calibrated reviewers on 29% of studies. Reviewers independently assessed these articles, then met to resolve disagreements and establish decision rules. Once we achieved 80% agreement on all RoB2 domains, reviewers proceeded with individual assessments of remaining studies.

#### Certainty Assessment

We assessed the certainty of the evidence for each outcome using the Grading of Recommendations Assessment, Development, and Evaluation (GRADE) framework, evaluating risk of bias, inconsistency, indirectness, imprecision, and publication bias^40,42^. Assessments were conducted in duplicate (SG, MN), with disagreements resolved through discussion.

#### Statistical Analysis

We extracted all eligible effect sizes from each study, using the post-test, between-groups standardised mean difference as the primary summary measure. All metrics were converted to Hedges’ *g*^43^. Our multilevel meta-analyses examined the effect of online professional learning on knowledge and skill acquisition through two distinct comparisons: online learning versus static controls (e.g., waitlists, electronic or print materials, self-directed textbook reading), and online learning versus face-to-face instruction. For each comparison, we quantified heterogeneity using *I²*. We tested the influence of this heterogeneity following Mathur and VanderWeele^44^, estimating the proportion of true effects (not due to sampling error) likely to be helpful (interpreted as a small, positive effect; Hedges’ g > 0.2) or harmful (Hedges’ g < –0.2).

To explore sources of heterogeneity, we pre-specified a range of moderators (see protocol https://osf.io/46zav). To avoid generalising from small samples, we only included moderator levels with at least three studies. For learning design principles, we planned three approaches to moderation analyses, given these design principles may co-occur and interact. First, we modelled the effect of all learning design principles simultaneously (akin to multiple regression). Then, we modelled the effect of each learning design principle, one at a time (akin to running multiple simple regressions), using 99% confidence interval (CI) to mitigate false positives from multiple comparisons. Finally, we used meta-analytic classification and regression trees (meta-CART) to explore possible interactions between moderator effects. The meta-CART analysis was performed using the *metacart* package in R (version 4.3.1)^45^ with default parameters.

#### Sensitivity and Publication Bias Analysis

We conducted a sensitivity analysis to assess the influence of extreme outliers, testing whether removal of effect sizes exceeding *g* > 2.5 meaningfully altered the pooled effect size estimates. To balance methodological rigour with analytic stability, outliers were retained in the main analysis but excluded from moderator analyses, as extreme effect sizes can disproportionately influence subgroup estimates and produce unstable or misleading moderator effect estimates^46,47^.

We conducted sensitivity analyses to assess whether the risk of bias in the included studies explained the variance observed in pooled effect size estimates. For each bias domain and for overall risk of bias, we modelled the effects of studies rated as low risk and compared effects against our overall analysis (including all studies).

We assessed publication bias using a multi-level meta-analytic Egger’s test to determine whether smaller studies reported disproportionately larger effect sizes, suggesting potential small-study effects^40,48^. This approach assessed whether effect sizes and their standard errors indicated publication bias while controlling for clustering, including a random effect for studies and incorporated delivery type as a categorical moderator to explore whether publication bias varied across control types. Standard error was modelled as a predictor to detect funnel plot asymmetry, an indicator of publication bias.

## Results

Our systematic search identified 55,376 titles and abstracts. Given the extensive literature identified, this paper addresses knowledge and skill acquisition, while a complementary paper reports effects on clinical practice^33^. After deduplication and screening (detailed in Figure 1; see Supplementary File 1 for PRISMA 2020 checklist^31^; see Supplementary File 4 for excluded full-text studies with reasons), we included 171 studies reporting 391 effect sizes from 25,412 participants. Sex or gender was reported in 123 studies (16,567 participants), of whom 74% were female. Table 1 provides the full characteristics of each included study.

**Figure 1.**
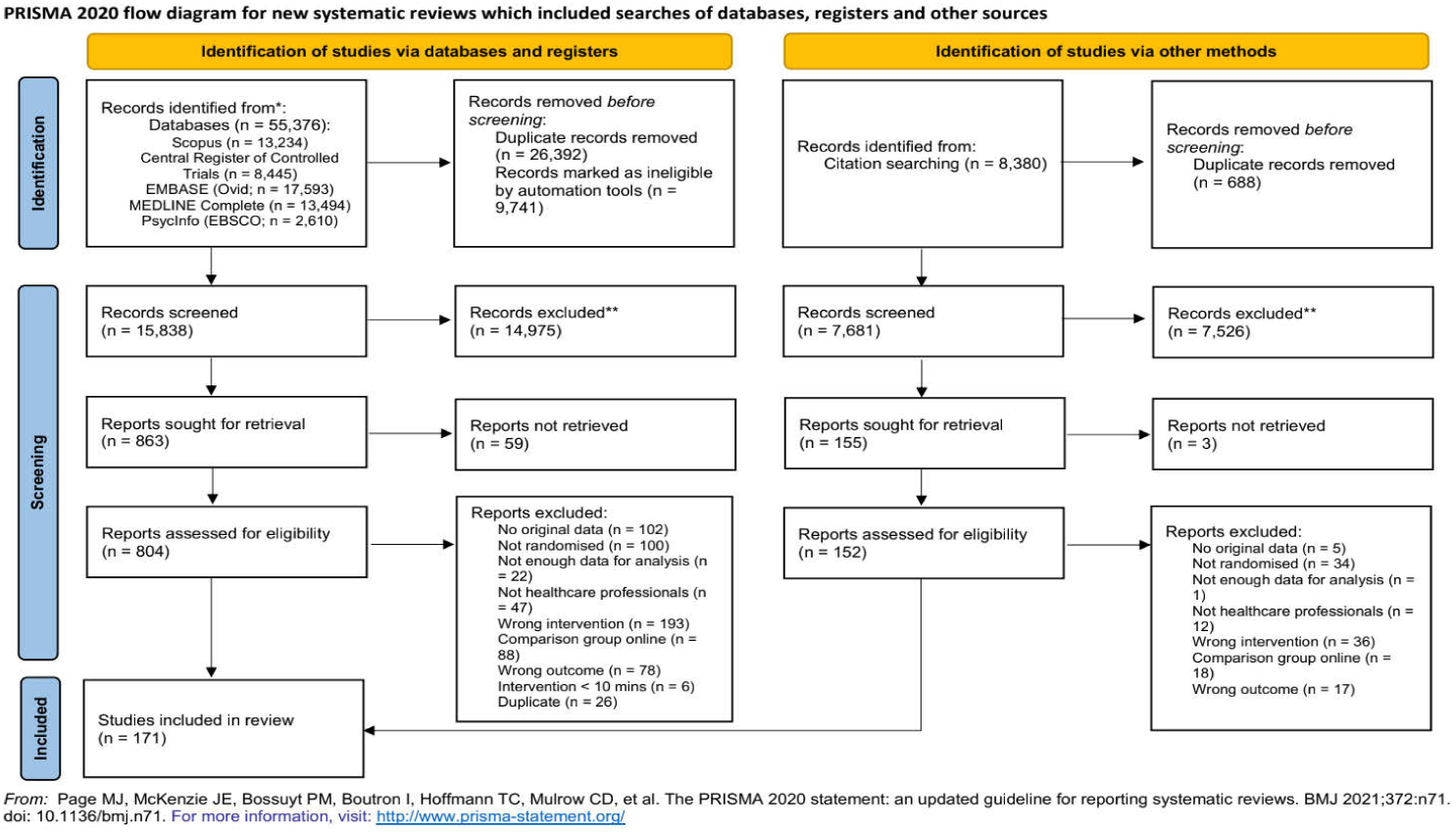
PRISMA flow diagram for inclusion in this systematic review.

**Table 1.**
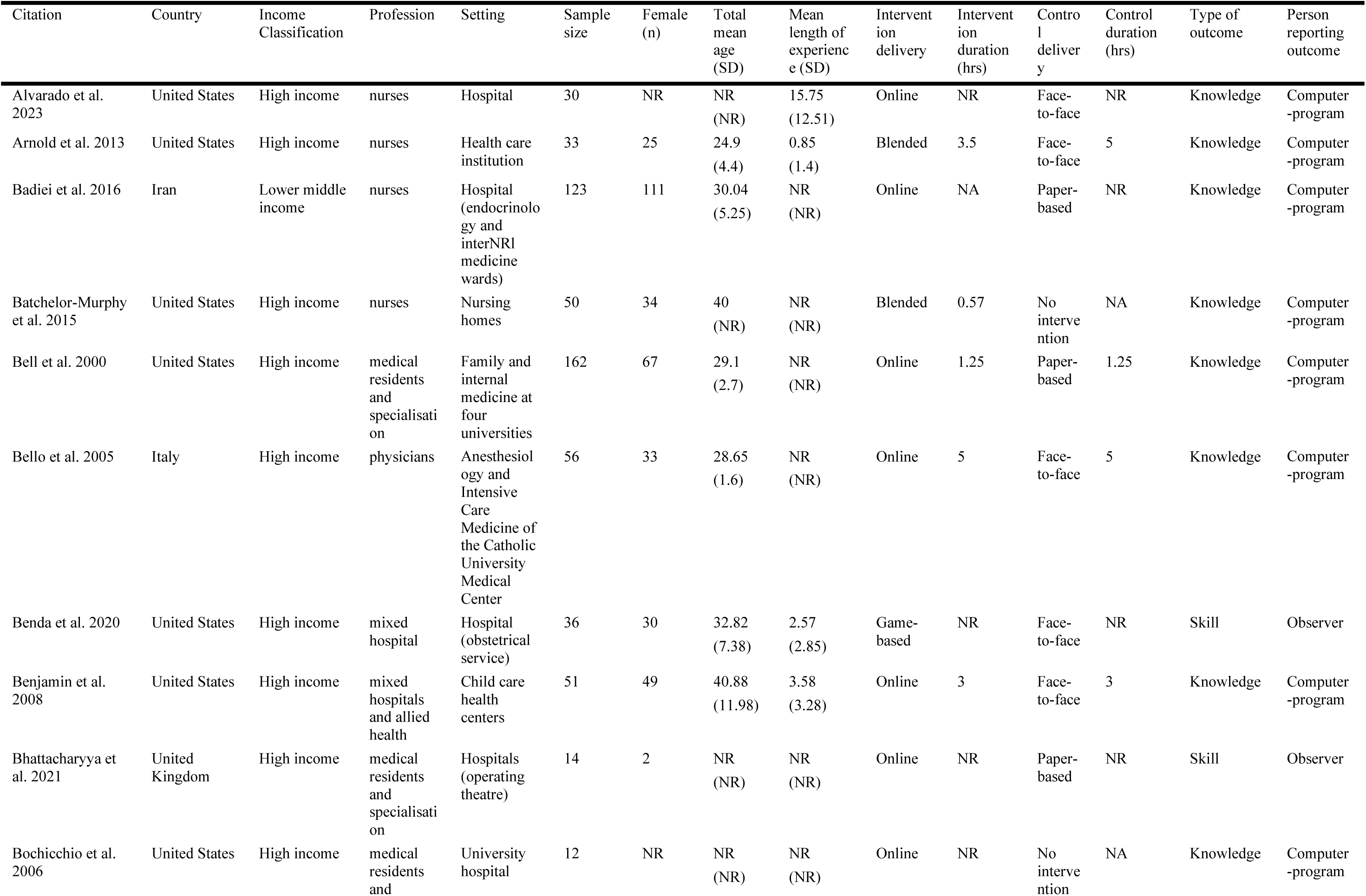
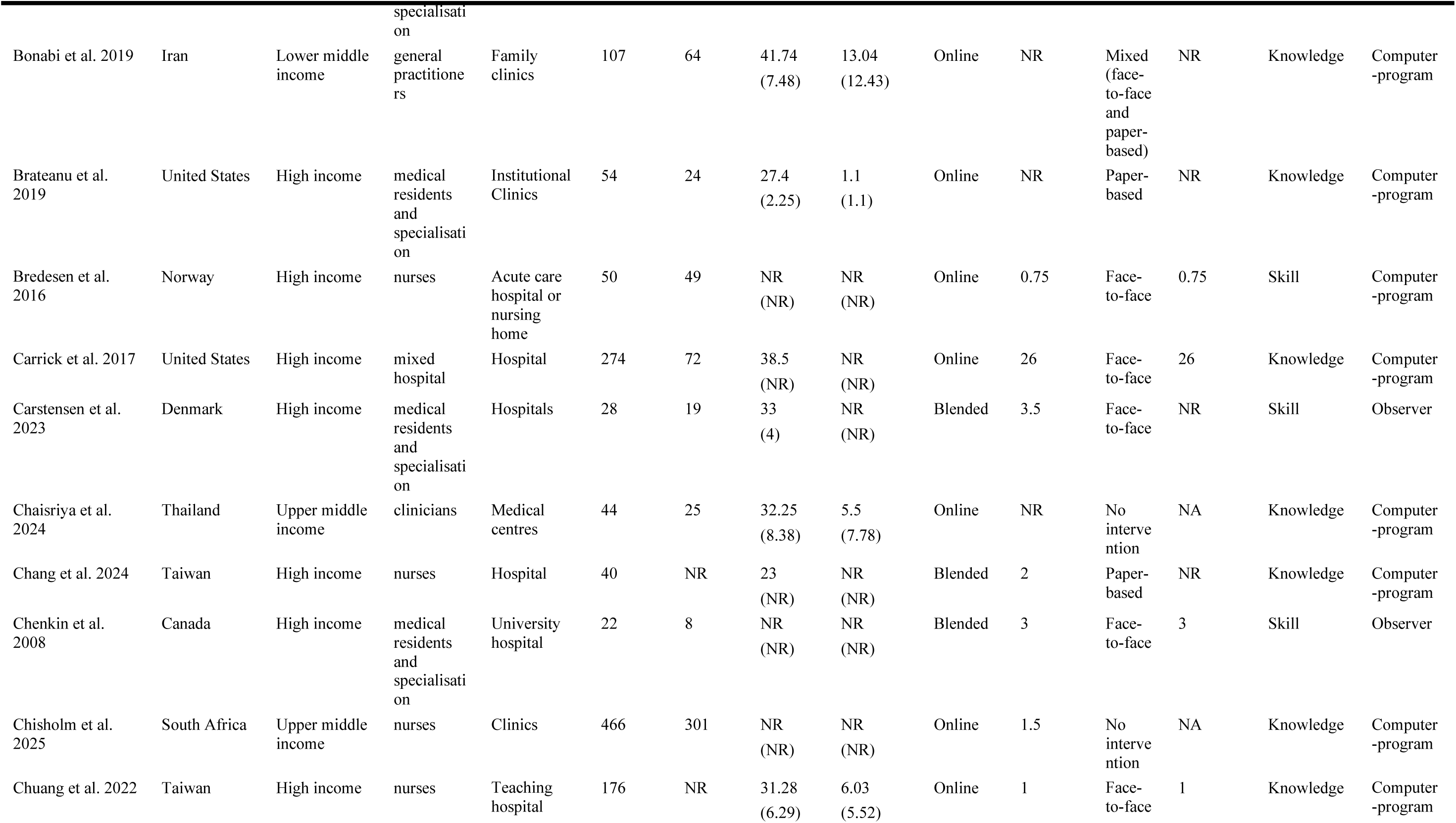
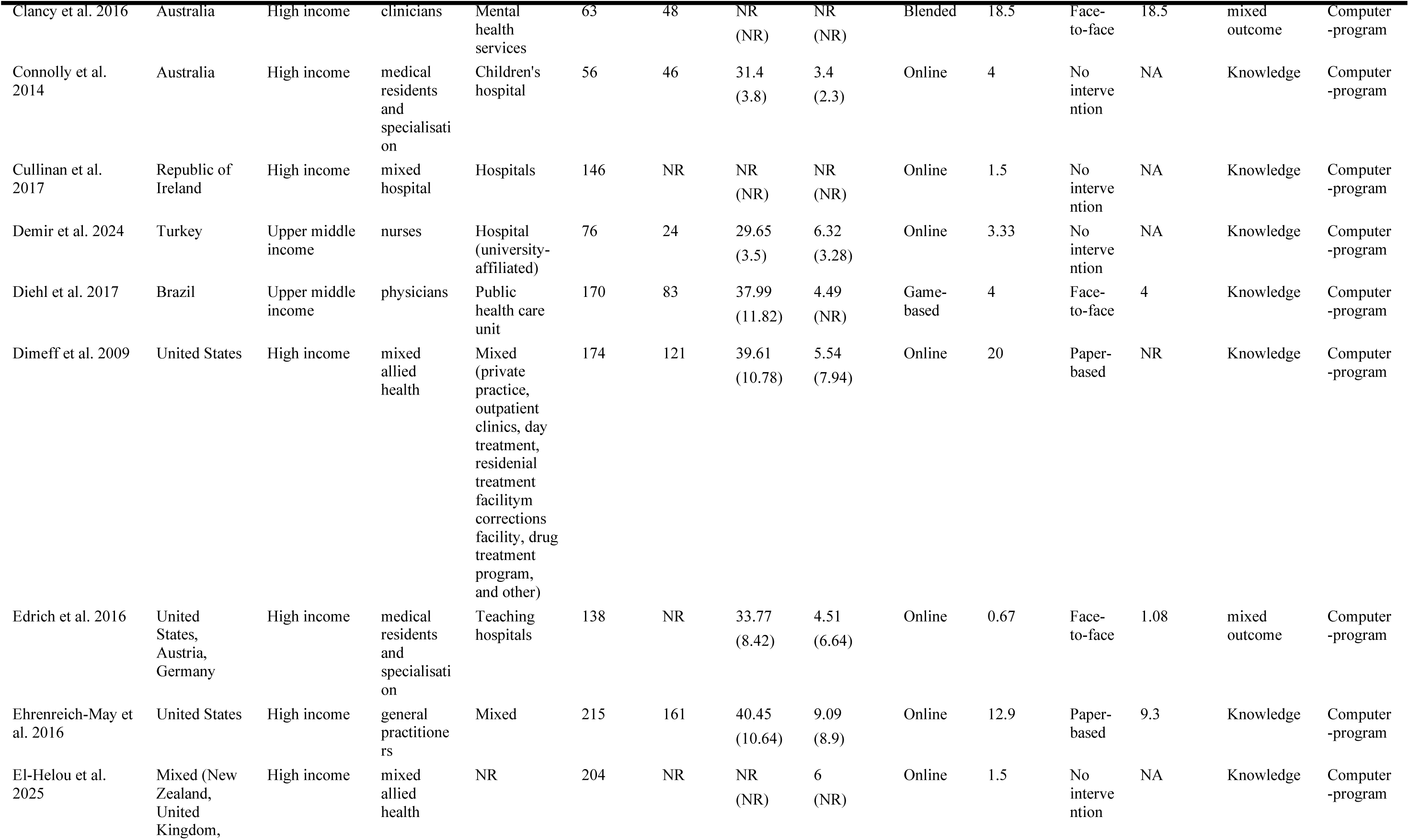
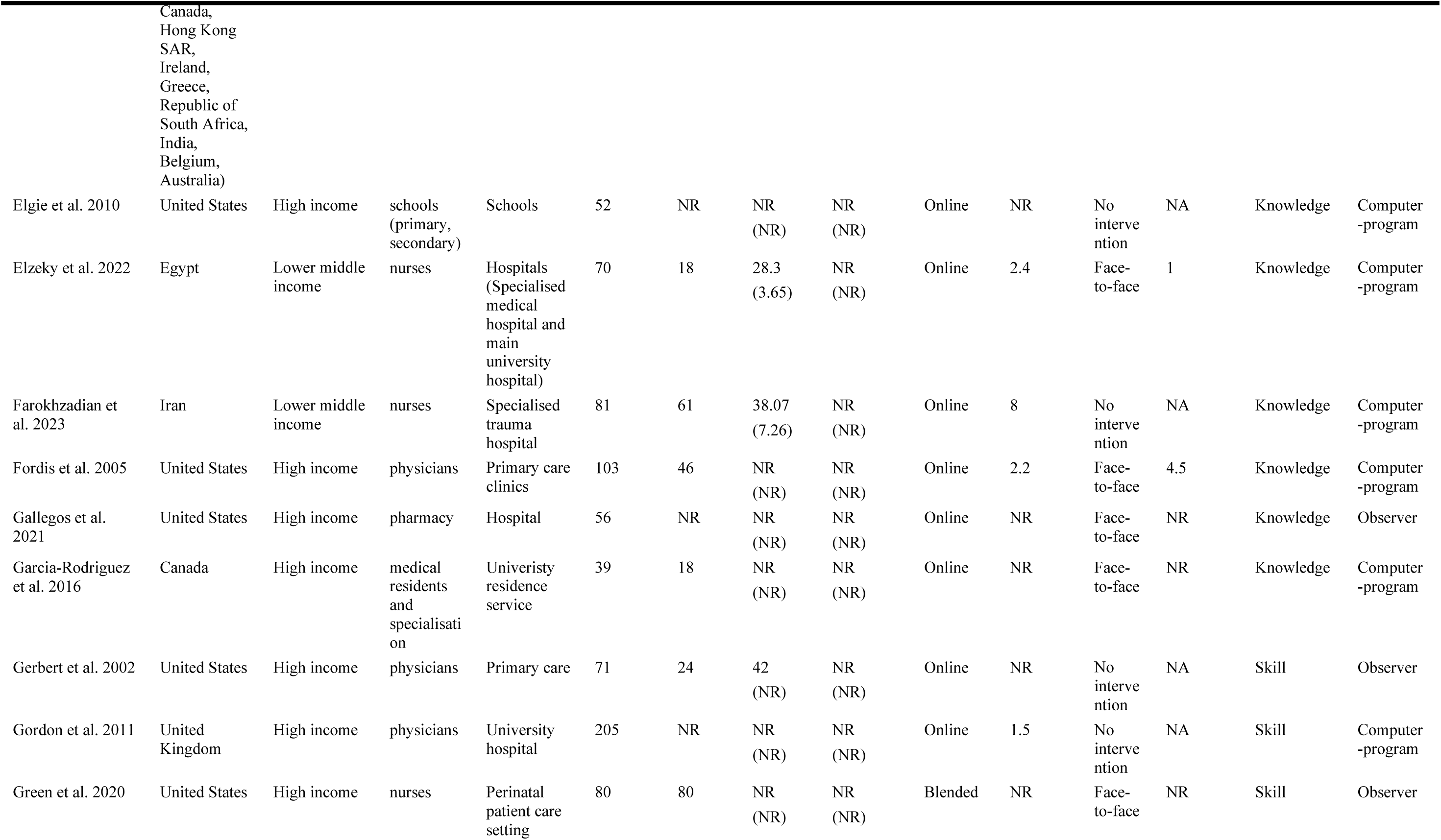
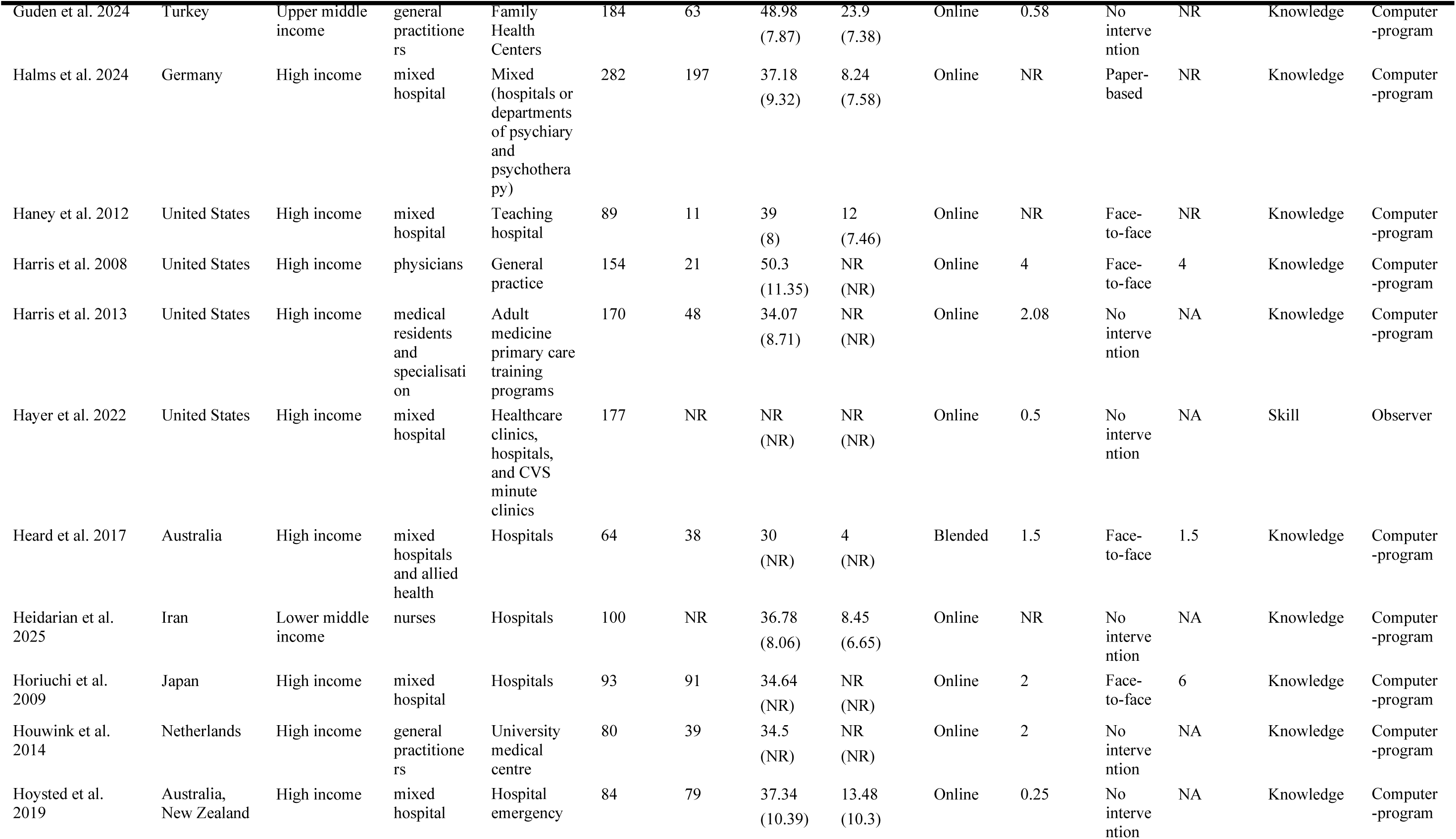
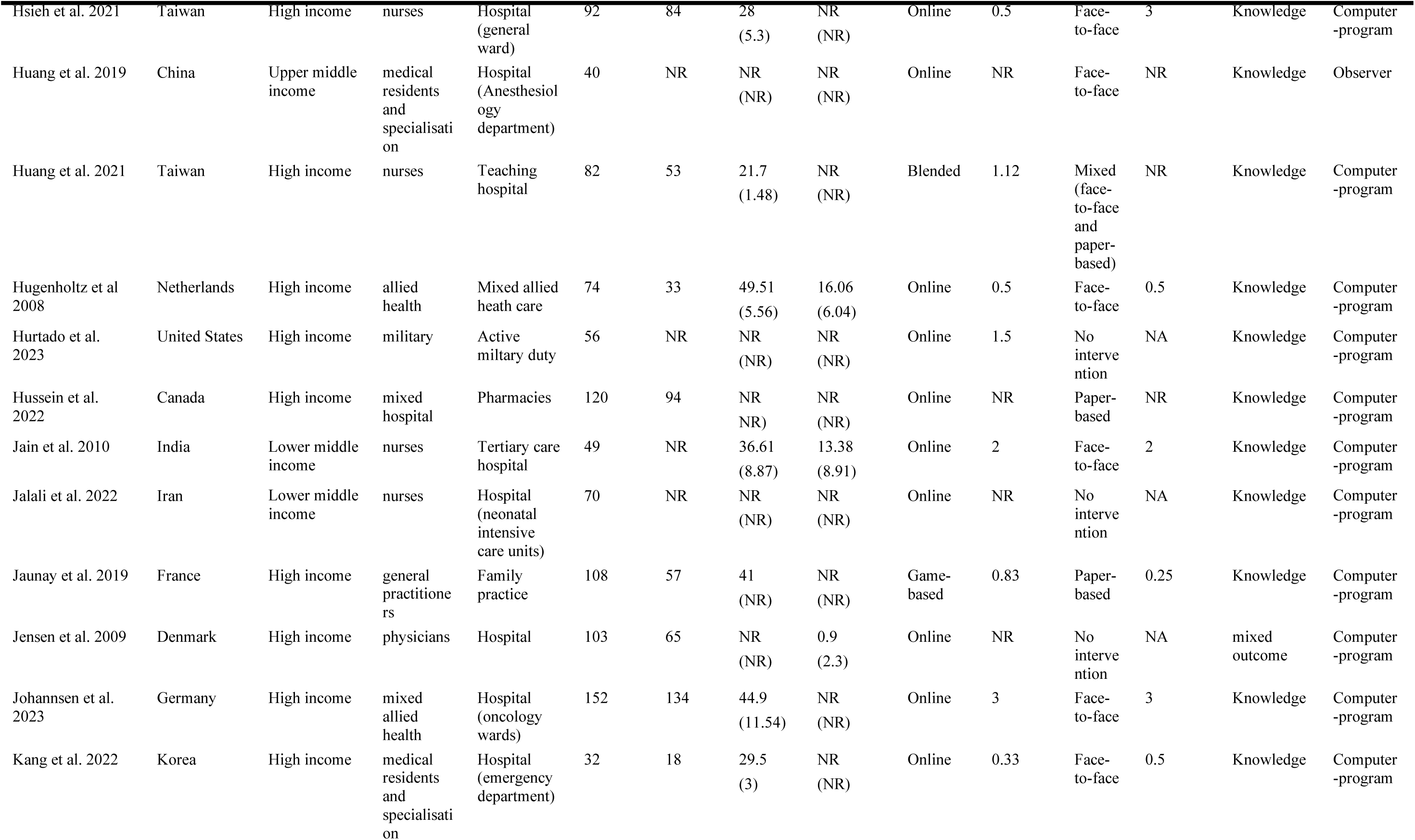
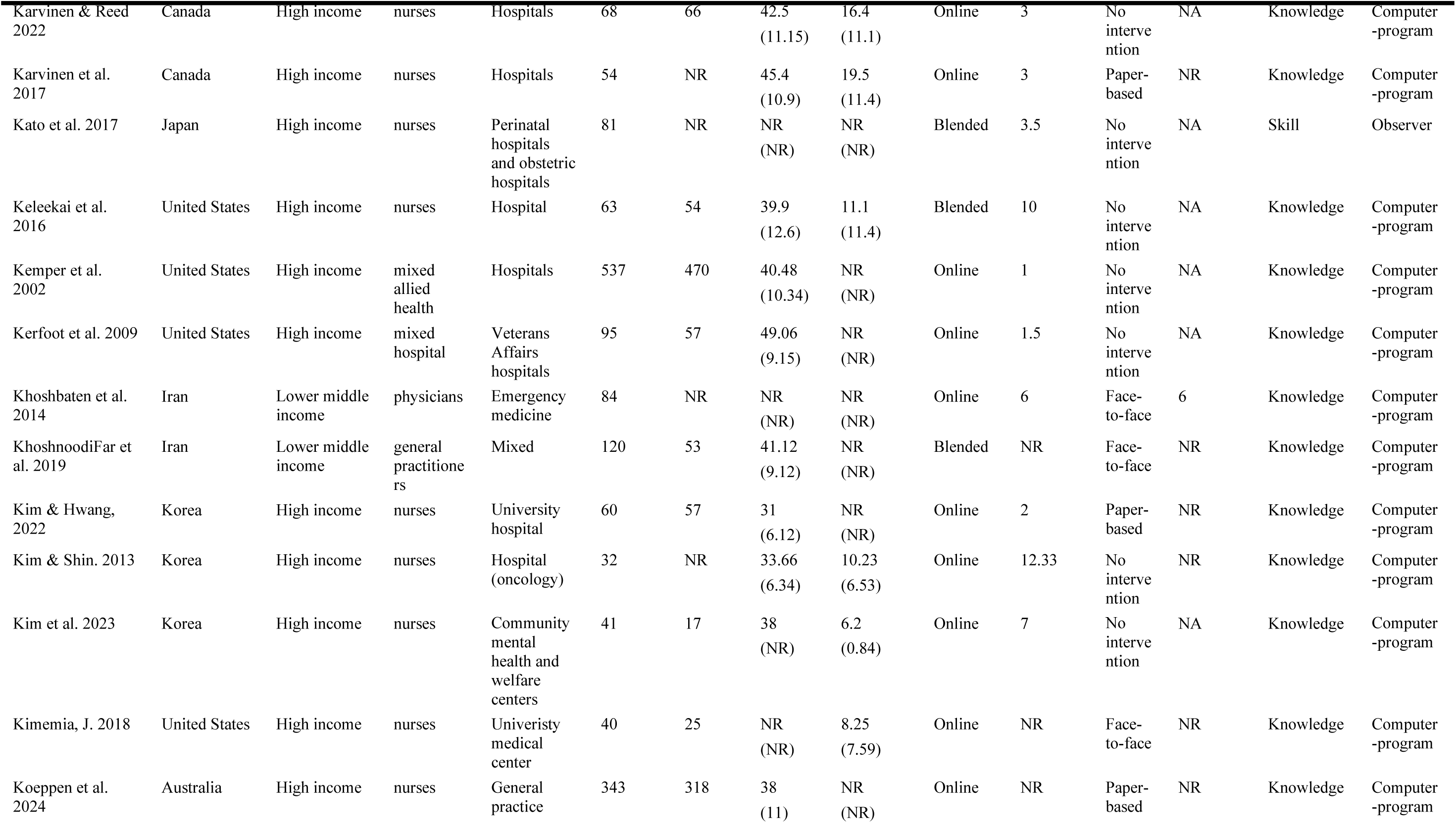
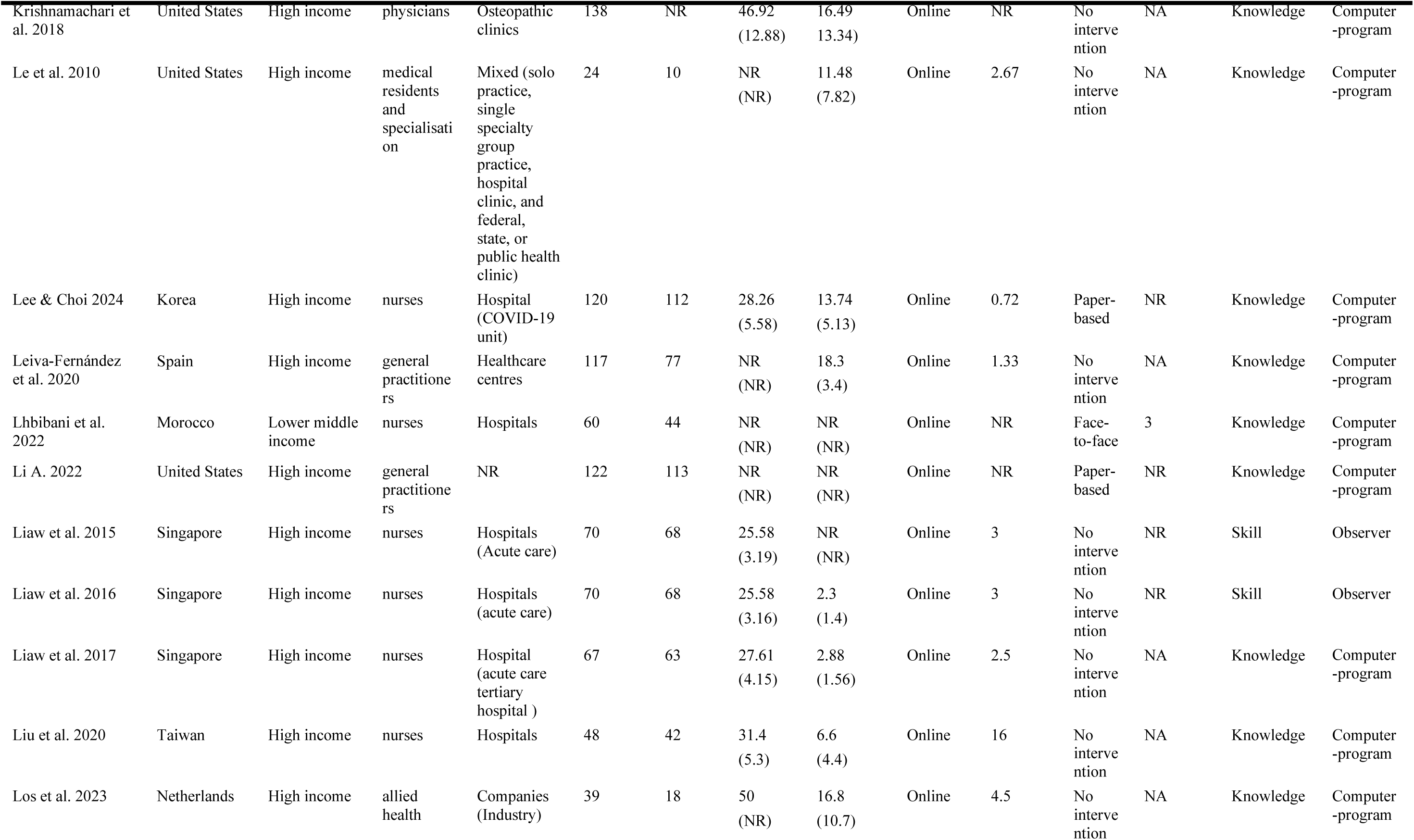
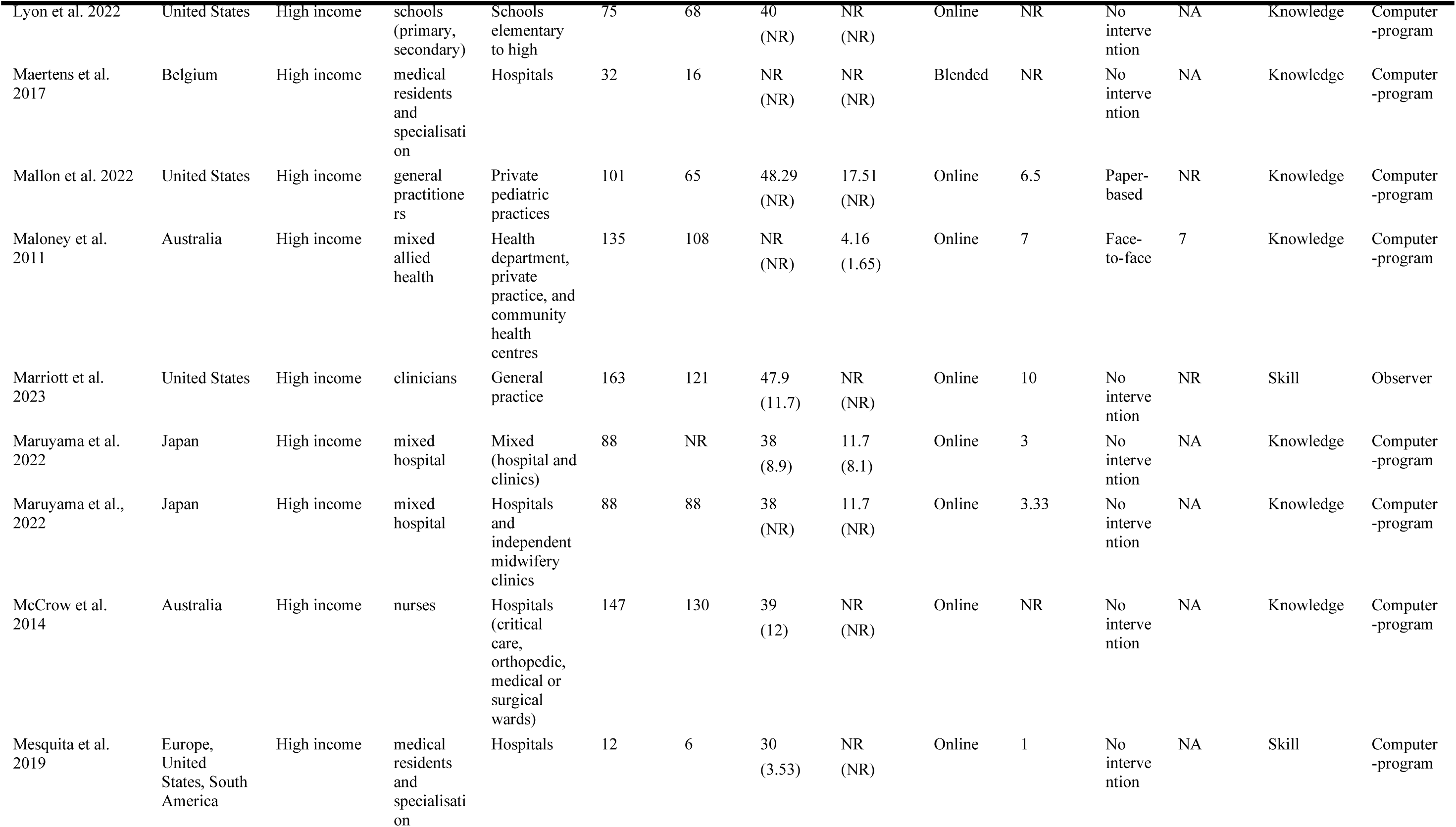
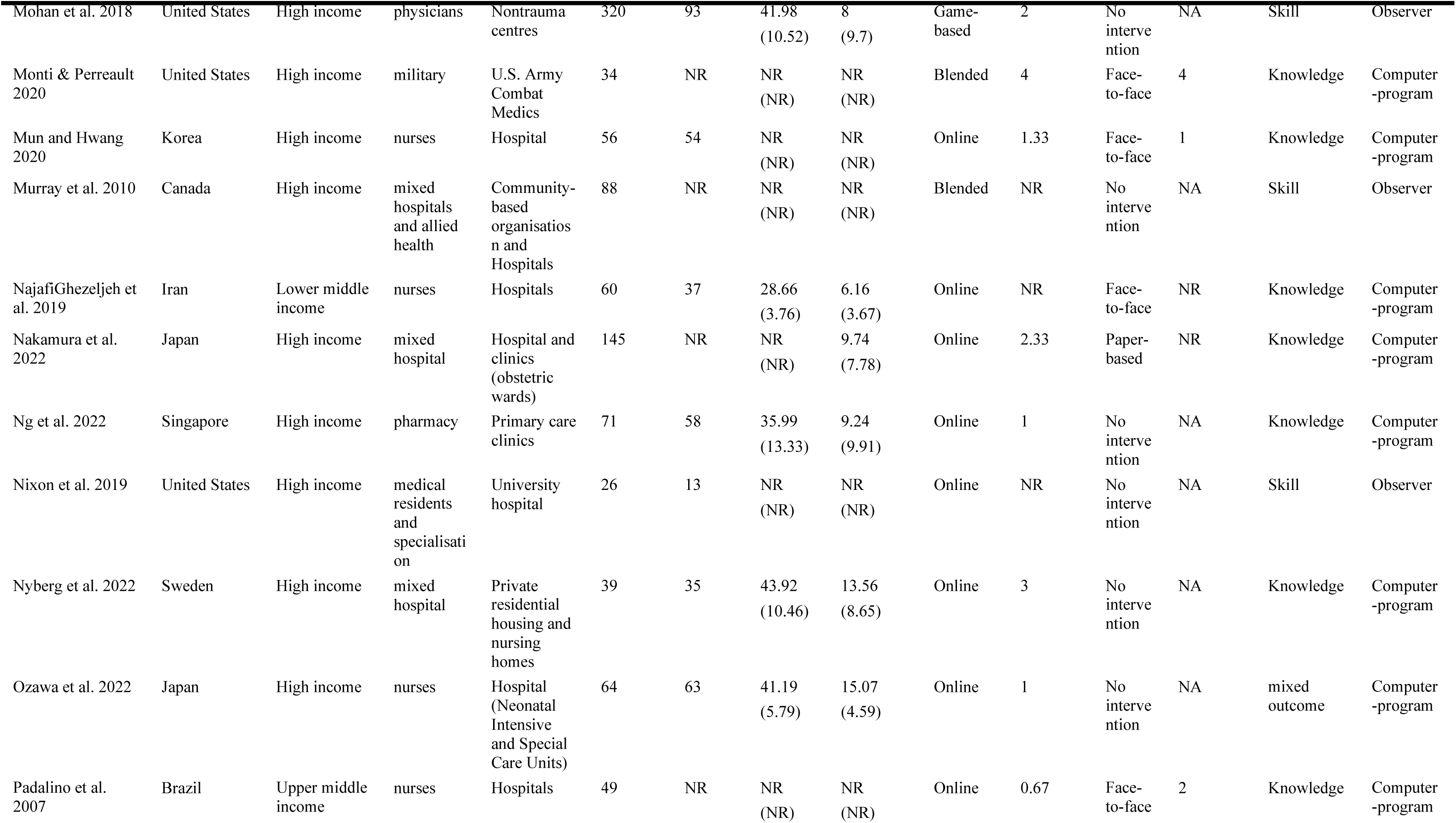
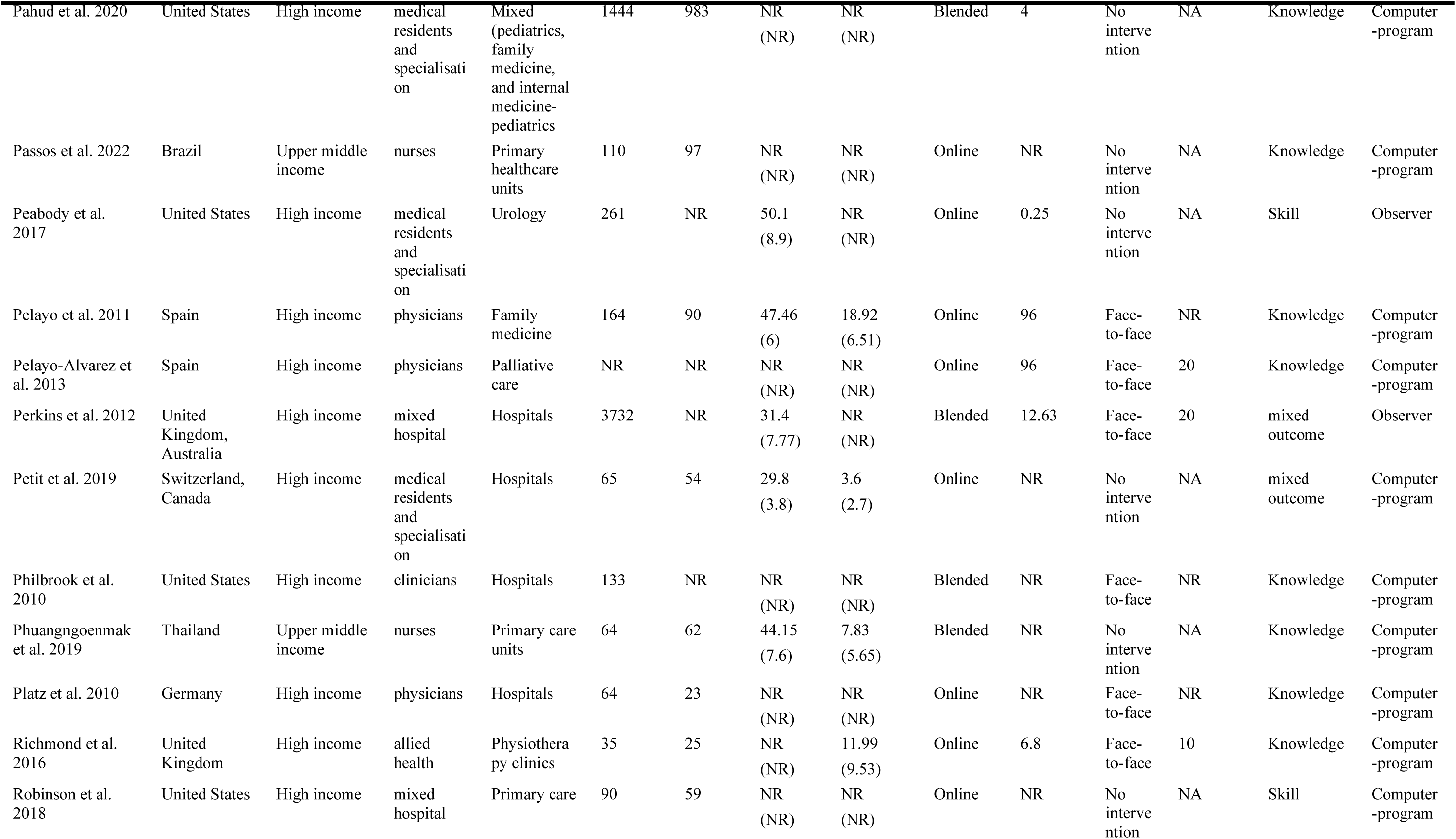
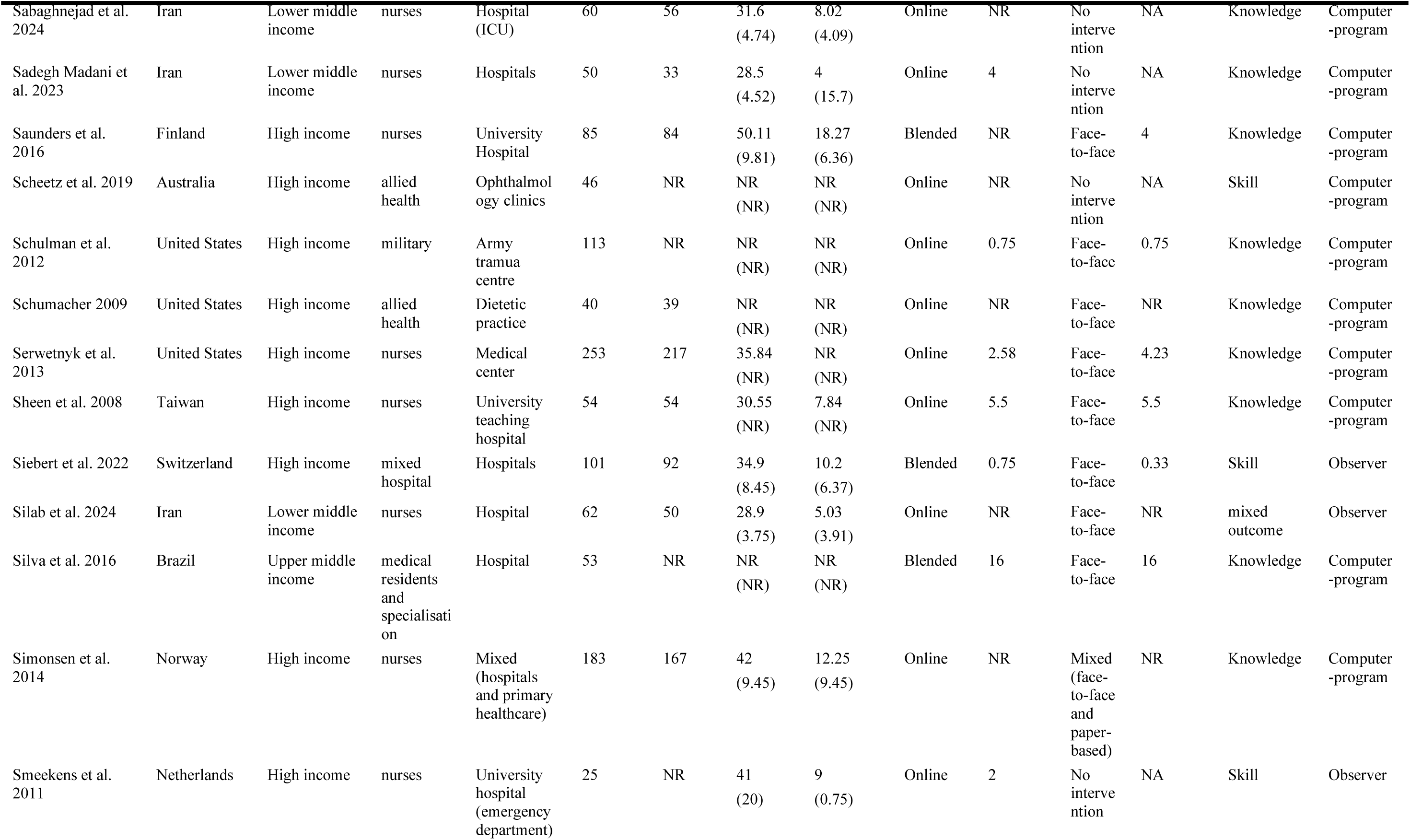
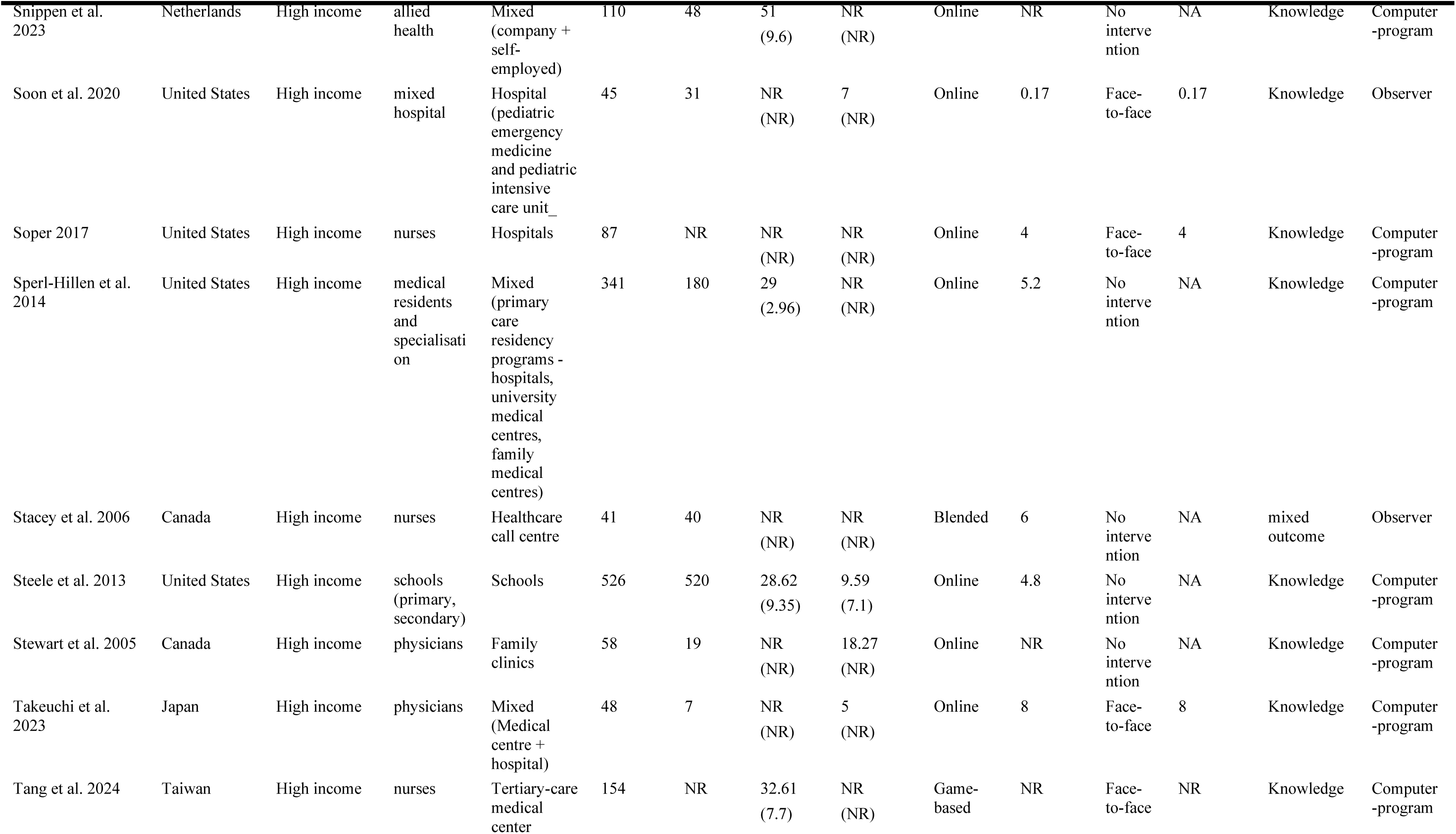
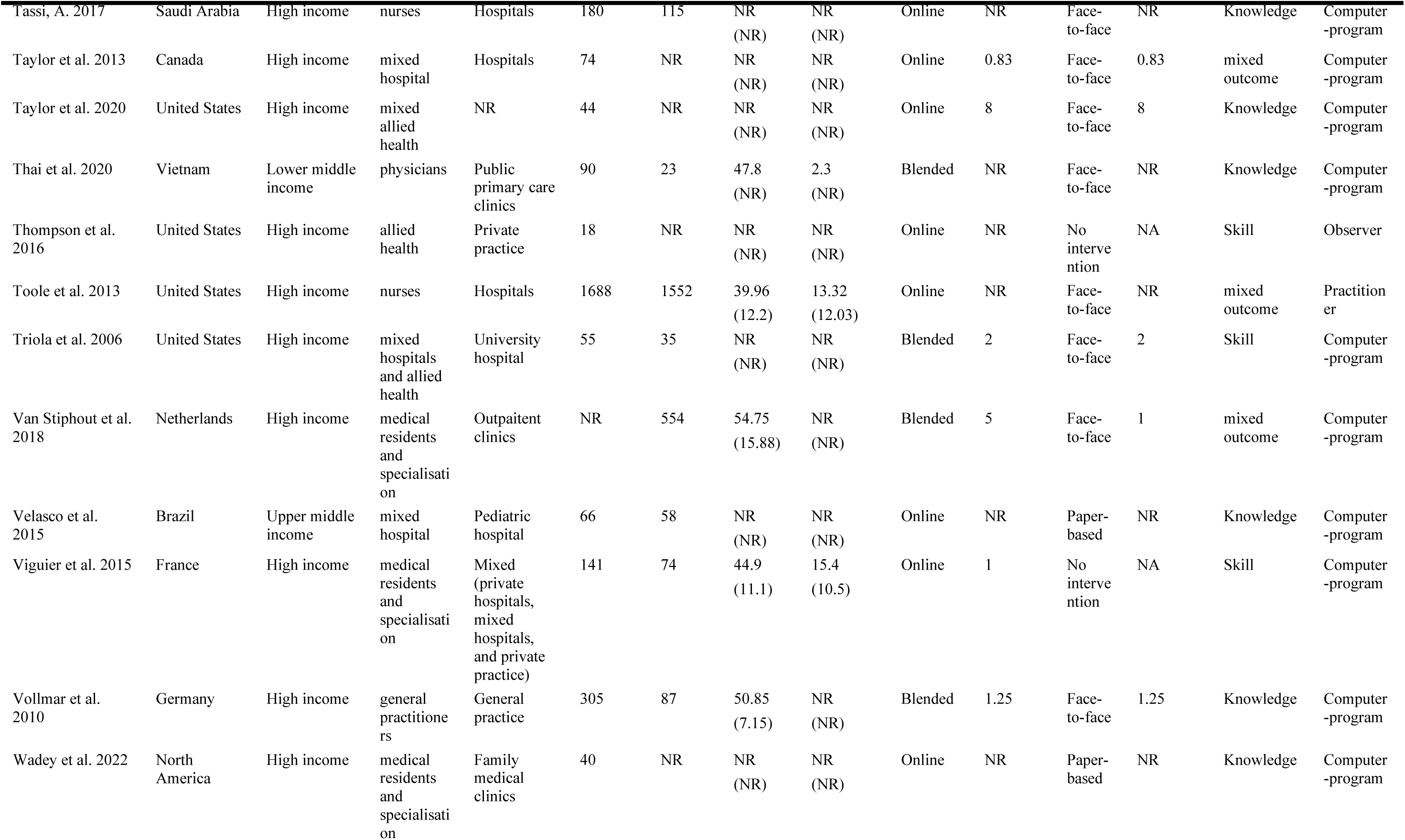
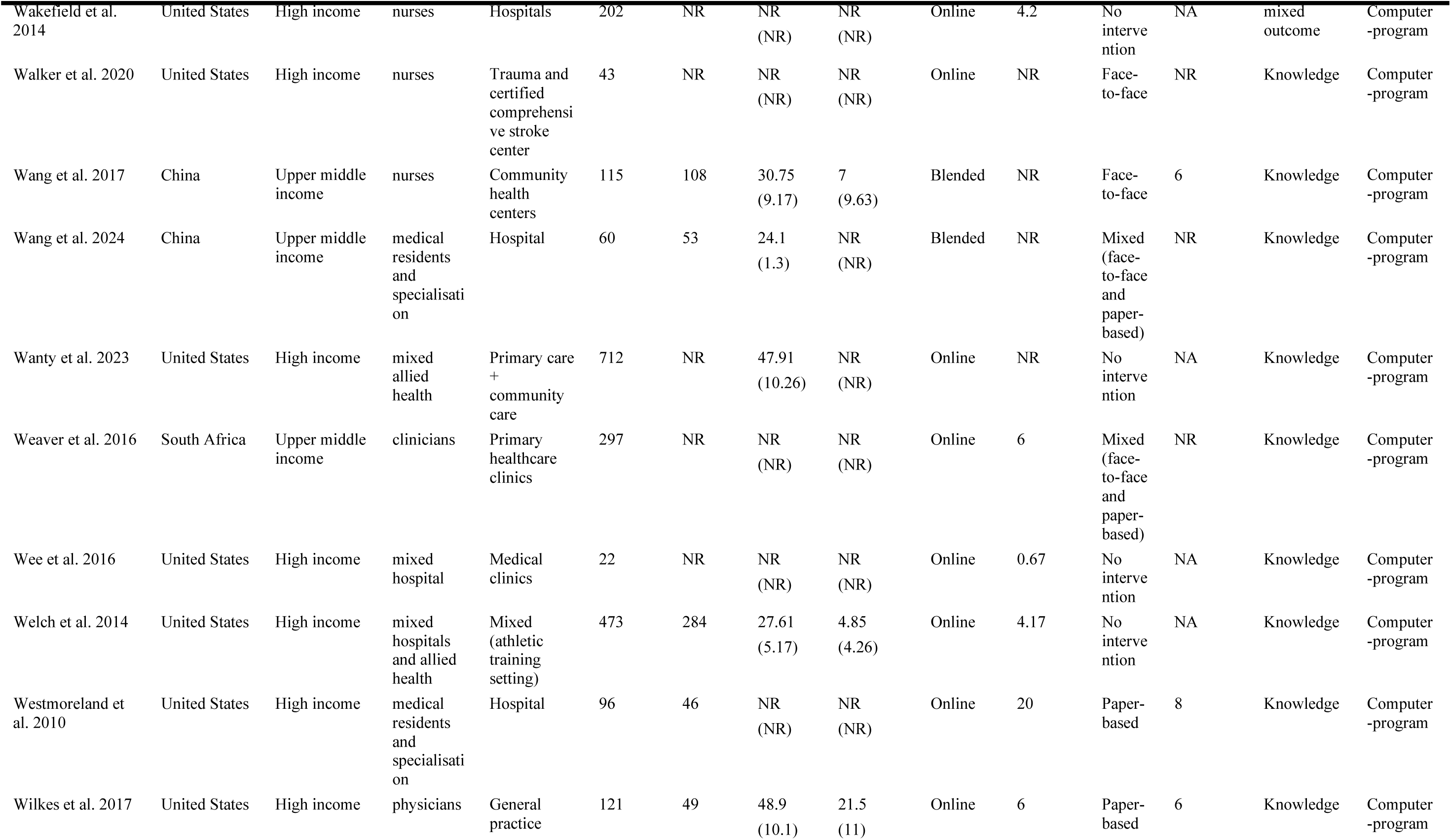
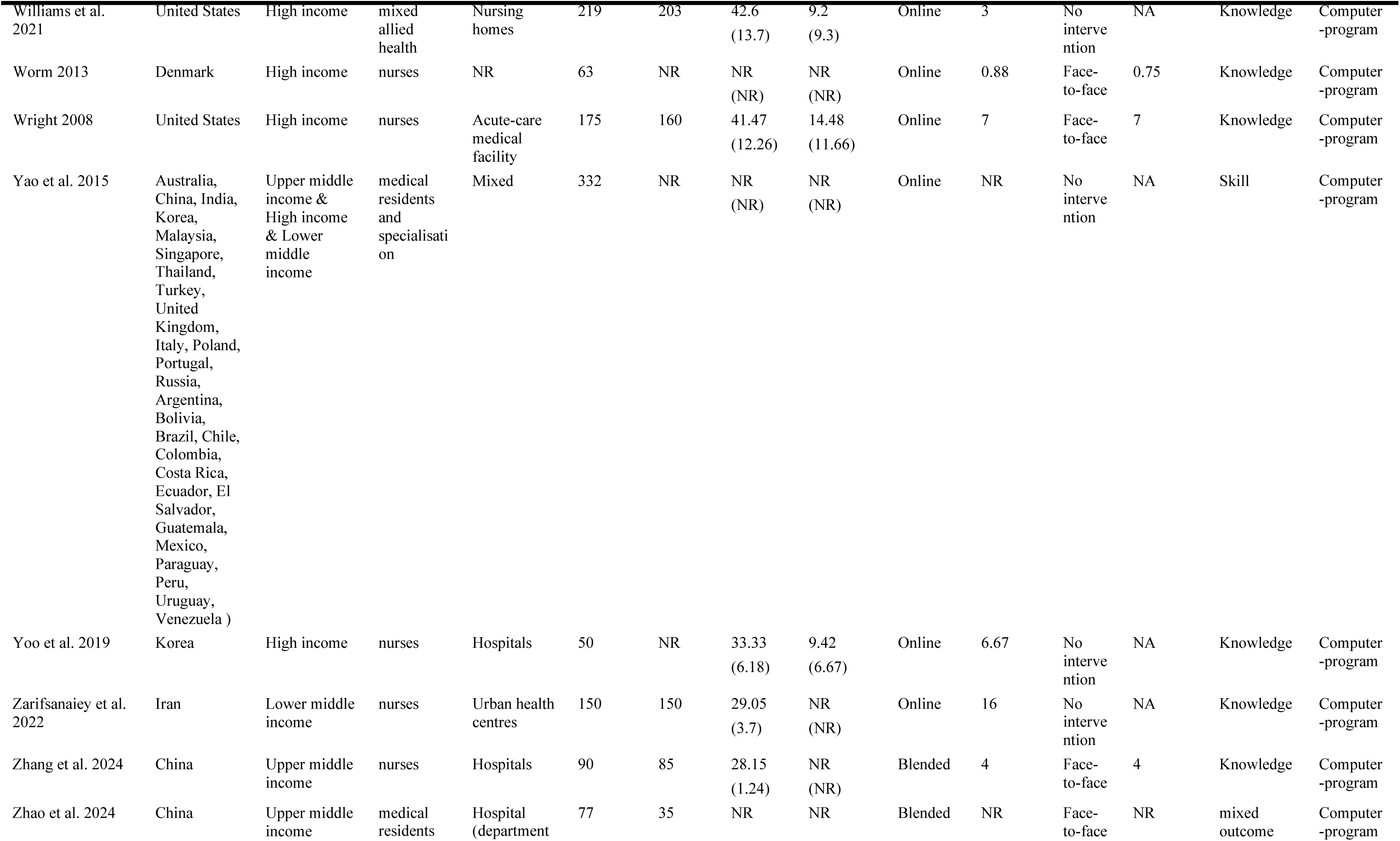
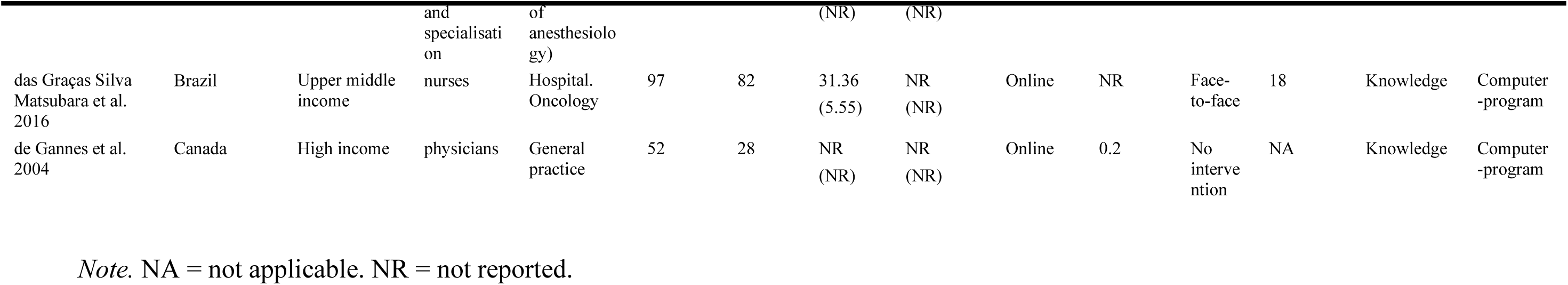
Study Characteristics Table of the Included Studies (N =171)

### Effect of Online Learning Versus Static Controls

The effect sizes of learning extracted from individual studies are shown in Figure 2. Overall, online interactive professional learning significantly improved knowledge and skill acquisition compared against static controls such as waitlist or static dissemination (*g* = 0.93, 95% CI [0.78,1.07], *n* = 103, *k* = 230, *p* < 0.001). This point estimate demonstrated a high level of heterogeneity that was not explained by sampling error alone (*I*^2^= 89.8). Our models estimated that online learning was likely to be meaningfully better than controls (i.e., *g* > 0.2) for 89% of true effects (95% CI [82%, 93%]). Models suggested online learning would be meaningfully worse (i.e., *g* < –0.2) than controls in 0% of true effects (95% CI [0%, 2%]) of cases.

**Figure 2.**
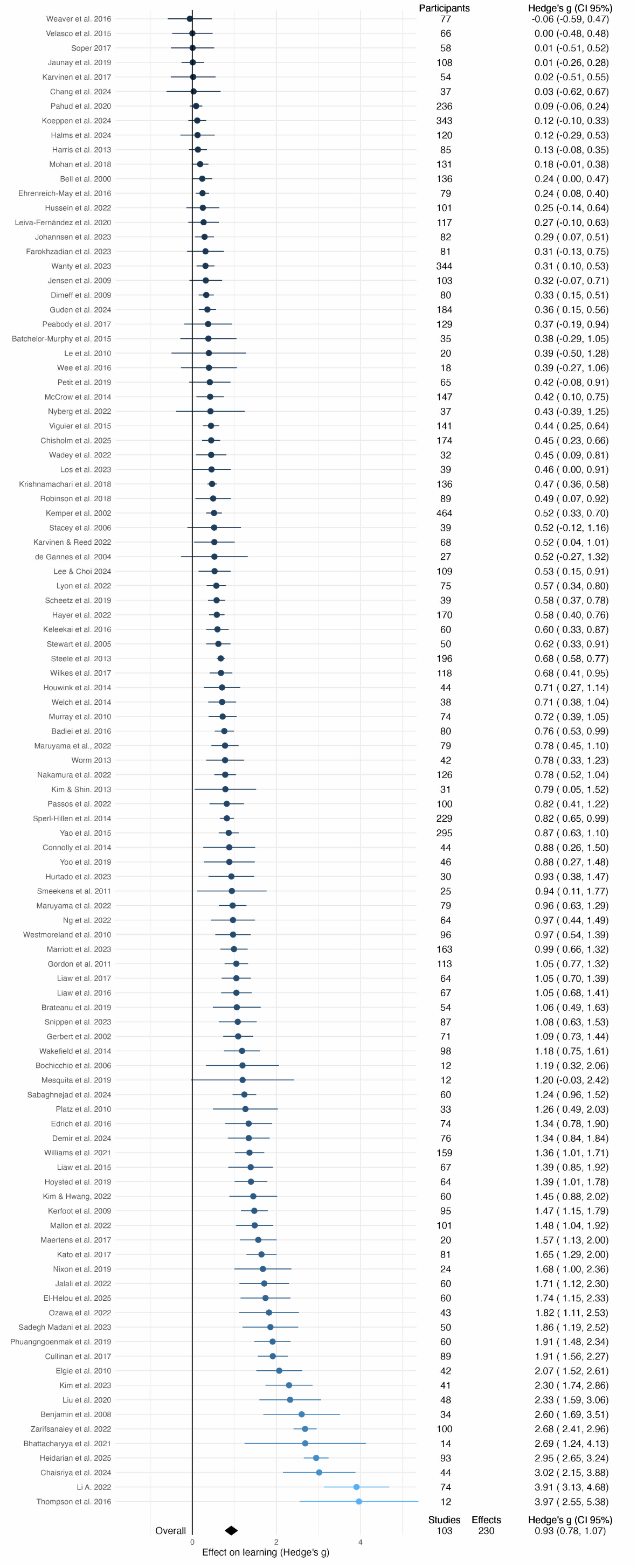
Forest plot of pooled result for online learning compared against static controls.

#### Outlier Consideration

Visual inspection of effect size distributions identified 18 extreme outliers with effect sizes *g* > 2.5 from 12 studies. Sensitivity analysis confirmed that removing these outliers had minimal impact on the pooled effect size (Δ*g* = 0.15), with overlapping 95% confidence intervals, demonstrating that the main effect estimate was robust to the inclusion of extreme values. All moderator analyses reported below were conducted using the filtered dataset without outliers (*n* = 97, *k* = 212).

#### Moderation Analysis for Online Learning Versus Static Controls

##### Moderation Analysis by Outcome

The type of learning outcome (i.e., knowledge vs skill) assessed did not significantly moderate the effectiveness of online learning in promoting learning (*F*(2, 209) = 0.44, *p* = 0.64). There was minimal variation in effect sizes for different outcome types (see Figure 3).

**Figure 3.**
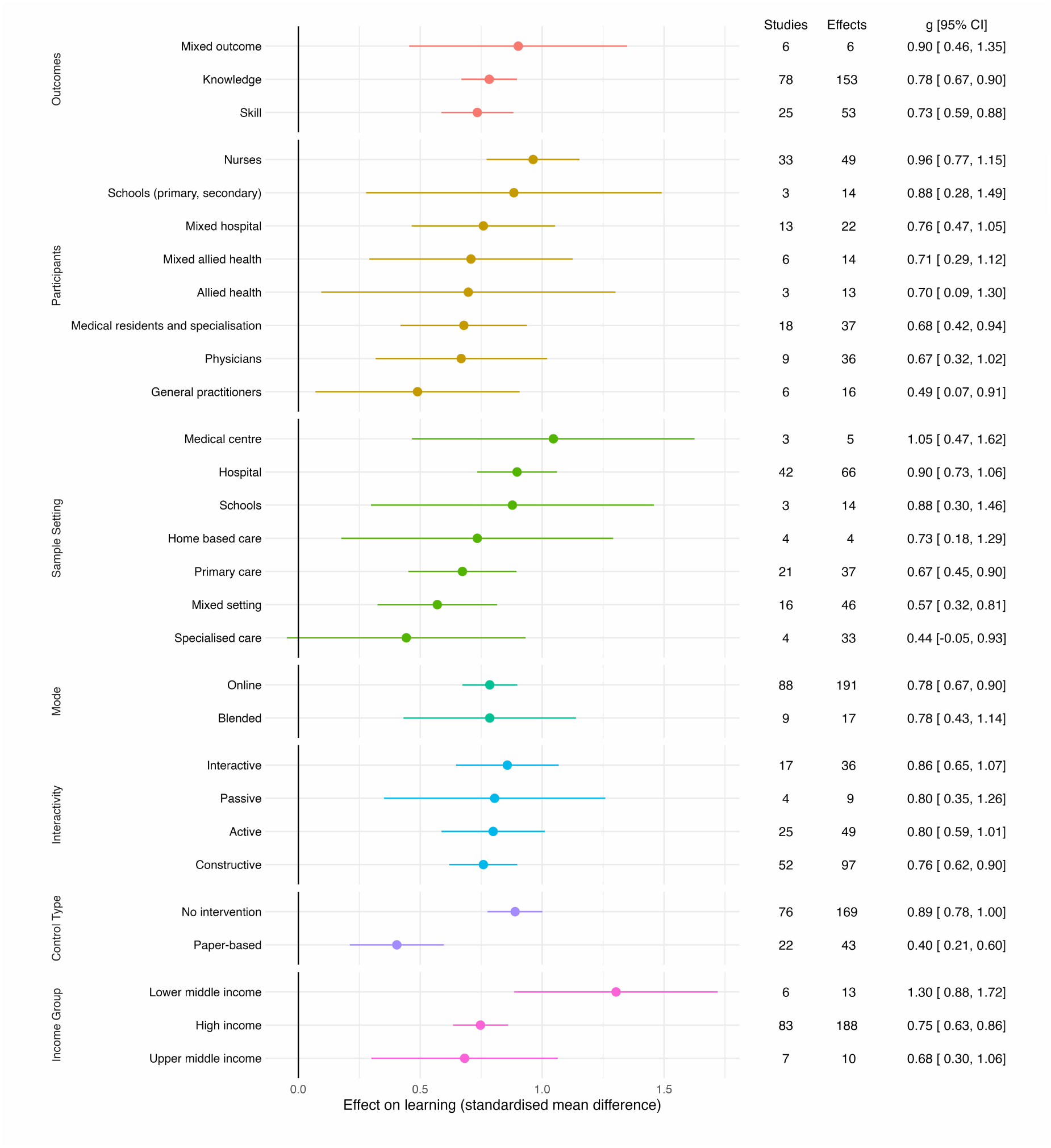
Summary of moderation analyses for online learning versus static controls by outcomes, participant, intervention, and comparison.

##### Moderation Analysis by Follow-up

The time delay between completion of the intervention and follow-up post-test measure was not a significant moderator of the effect of online learning on knowledge and skills acquisition (*F*(1,210) = 0.11, *p* = 0.74).

##### Moderation Analysis by Participant

The type of professional (*F*(7, 193) = 0.97, *p* = 0.46) and length of professional experience was not a significant moderator (*F*(1, 107) = 2.54, *p* = 0.11).

##### Moderation Analysis by Sample Setting

The setting (e.g., hospital, medical centre) in which a healthcare professional worked was not a significant moderator of effectiveness (*F*(6, 198) = 1.40, *p* = 0.21; see Figure 3 for all settings).

##### Moderation Analysis by Intervention

The length of an online learning program (in hours) was not a significant moderator (*F*(1, 130) = 1.36, *p* = 0.25.

The type of online learning (blended, or online only) was not a significant moderator (*F*(1, 206) = 0.00, *p* = 1). As shown in Figure 3, both methods showed significant positive effects on knowledge and skills acquisition.

The degree of interactivity (i.e., passive, active, interactive, constructive) in the online learning intervention did not significantly moderate its overall impact on knowledge and skills acquisition (*F*(3,187) = 0.27, *p* = 0.85).

##### Moderation Analysis by Control Type

The type of comparison group was a significant moderator of the effect of online learning on knowledge and skills acquisition (*F*(1, 210) = 19.89, *p* < 0.001). As shown in Figure 3, online learning demonstrated a higher effect size compared to no intervention (*g* = 0.89, 95% CI [0.78, 1.00], *n* = 76, *k* = 169, *p* <0.001) and a smaller but still positive effect compared to paper-based learning (*g* = 0.40, 95% CI [0.21, 0.60], *n* = 22, *k* = 43, *p* <0.001). These results indicate that online learning consistently outperformed static controls such as waitlist or paper-based educational materials.

##### Moderation Analysis by Income Groups

The income level of the country where the study was conducted significantly moderated effects (*F*(2, 208) = 3.34, *p* = 0.04). Lower-middle income countries showed substantially larger effects (*g* = 1.30, 95% CI [0.88, 1.72], *n* = 6, *k* = 13, *p* <0.001) compared to high income (*g* = 0.75, 95% CI [0.63, 0.86], *n* = 83, *k* = 188, *p* <0.001) and upper middle income countries (*g* = 0.68, 95% CI [0.30, 1.06], *n* = 7, *k* = 10, *p* = 0.001; see Figure 3).

##### Moderator Analysis by Other Design Features

When considering all learning design components together, there was no significant moderation effect on knowledge and skills acquisition (*F*(10, 219) = 1.45, *p* = 0.16; see Supplementary File 5). When separately modelling the effect of each learning design feature in isolation (see Supplementary File 5), none showed significant effects on knowledge and skill outcomes. Planned analysis using classification and regression trees (meta-CART) did not identify any significant moderators.

### Effect of Online Learning Versus Face-to-Face

The effect sizes of learning extracted from individual studies are shown in Figure 4. When compared to face-to-face learning, online learning maintained a significant positive effect (*g* = 0.45, 95% CI [0.31,0.59], *n* = 76, *k* = 162, *p* < 0.001). The analysis indicated a high level of heterogeneity (*I*^2^ = 85.92), which suggests that the variability in effect sizes across studies is not solely due to sampling error. Accounting for this heterogeneity, approximately 60% of true effects (95% CI [47%, 69%]) were likely to be beneficial (i.e., *g* > 0.2), and only 6% of true effects (95% CI [2%, 15%]) of interventions would be detrimental (i.e., *g* < –0.2).

**Figure 4.**
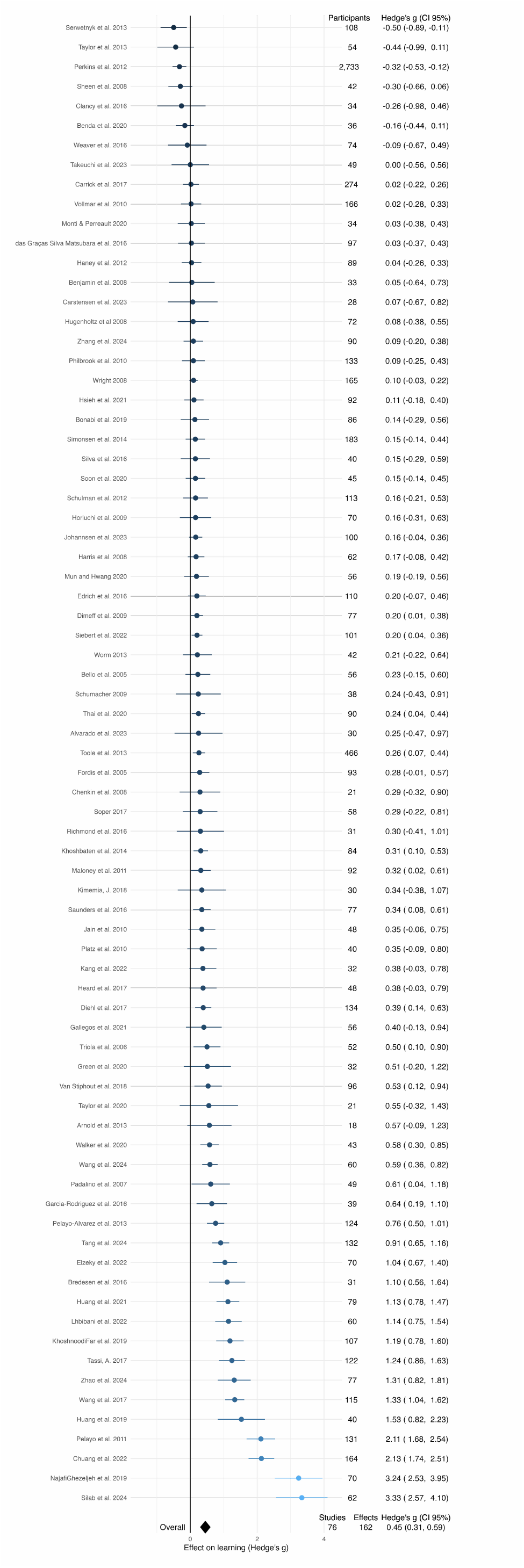
Forest plot of pooled result for online learning compared against face-to-face controls.

#### Outlier Considerations

Consistent with the analytical approach for static controls, visual inspection identified 3 extreme outliers (*g* > 2.5) from three studies. Sensitivity analysis confirmed that removing these outliers had minimal impact on the pooled effect size (Δ*g* = 0.07), with overlapping 95% confidence intervals, demonstrating that the main effect estimate was robust to the inclusion of extreme values. All moderator analyses reported below were conducted using the filtered dataset without outliers (*n* = 74, *k* = 159).

#### Moderation Analysis for Online Learning Versus Face-to-Face

##### Moderation Analysis by Outcome

The type of learning outcome assessed (i.e., knowledge, skills, or mixed) significantly moderated the effectiveness of online learning in promoting knowledge and skills acquisition (*F*(2, 156) = 7.03, *p* = 0.001; See Figure 5). Online learning showed a significant positive effect for knowledge acquisition (*g* = 0.46, 95% CI [0.33, 0.59], *n* = 62, *k* = 94, *p* <0.001) and skills acquisition (*g* = 0.20, 95% CI [0.04, 0.36], *n* = 25, *k* = 58, *p* = 0.017), while mixed outcomes (*g* = 0.28, 95% CI [-0.13, 0.69], *n* = 6, *k* = 7, *p* = 0.179) were positive but not statistically significant.

**Figure 5.**
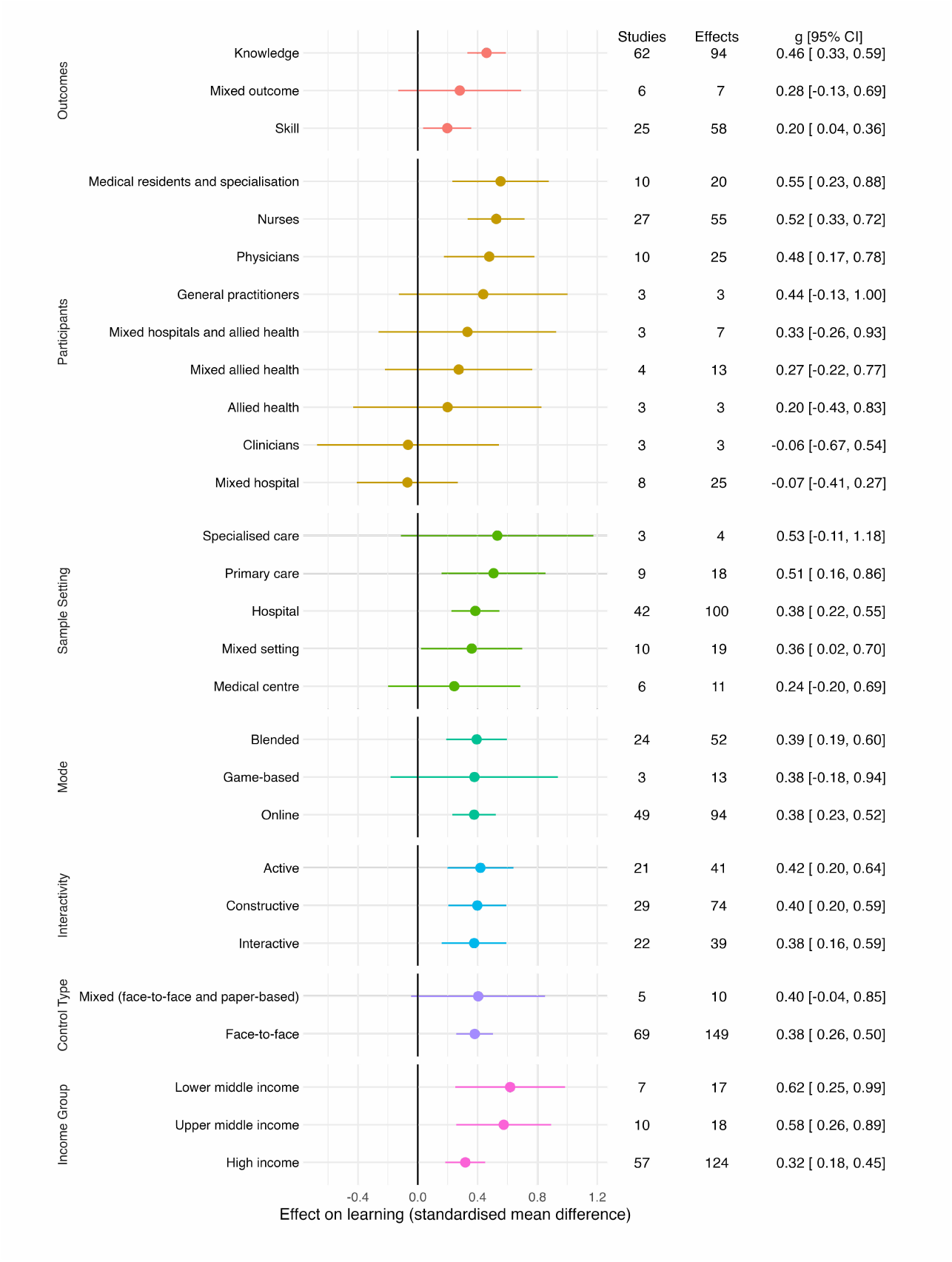
Summary of moderation analyses for online learning versus face-to-face by outcomes, participant, intervention, and comparison.

##### Moderation Analysis by Follow-up

The length of follow-up was not a significant moderator of the effect of online learning on knowledge and skill outcomes (*F*(1, 157) = 0.41, *p* = 0.52).

##### Moderation Analysis by Participant

The type of professional (*F*(8, 145) = 1.67, *p* = 0.11) and length of professional experience was not a significant moderator of effects (*F*(1, 67) = 0.44, *p* = 0.51).

##### Moderation Analysis by Sample Setting

The setting (e.g., hospital, medical centre) in which the online learning took place did not significantly moderate its effectiveness in improving knowledge and skill acquisition (*F*(4, 147) = 0.27, *p* = 0.9; See Figure 5).

##### Moderation Analysis by Intervention

The longer online learning programs (i.e., > 80 hours) appeared to lead to smaller effects (*F*(1, 104) = 7.78, *p* = 0.01). However, after removing two outliers where the online learning intervention exceeded 80 hours, moderation by length was no longer significant (*F*(1, 101) = 2.77, *p* = 0.1). The next longest intervention in the dataset was 20 hours (spread over days or months), suggesting that within this more typical range, intervention length does not substantially influence effectiveness.

The type of online learning (i.e., blended, game-based, or online only) did not significantly influence its effectiveness compared to face-to-face learning (*F*(2, 156) = 0.01, *p* = 0.99). Similarly, the degree of interactivity (i.e., passive, active, interactive, constructive) in the online learning intervention did not significantly moderate its impact on knowledge and skills acquisition (*F*(2,151) = 0.04, *p* = 0.96).

##### Moderation Analysis by Control Type

The type of comparison group (i.e., face-to-face or face-to-face with paper-based) was not a significant moderator on knowledge and skill outcomes (*F*(1,157) = 0.01, *p* = 0.92).

##### Moderation Analysis by Income Groups

The income-level of the country where the study was conducted did not significantly moderate the effect of online learning on knowledge and skill outcomes (*F*(2, 156) = 1.98, *p* = 0.14).

##### Moderator Analysis by Learning Design Features

When considering all learning design components together, there was no significant moderation effect on knowledge and skill outcomes (see Supplementary File 6; *F*(7, 151) = 0.86, *p* = 0.54). When separately modelling the effect of each learning design feature in isolation (see Supplementary File 6), none showed significant effects on knowledge and skill outcomes. Planned analysis using classification and regression trees (meta-CART) did not identify any significant moderators.

### Sensitivity Analyses

Sensitivity analysis was conducted to evaluate the impact of Cochrane’s Risk of Bias domains on the observed effect sizes. Full consensus ratings for each included study are presented in Supplementary File 7. As illustrated in Figure 6, no studies were categorised as having a low overall risk of bias. Therefore, we ran separate sensitivity analyses for each Risk of Bias criterion, when at least three studies met the low-risk threshold. Effect estimates were smaller in studies at low risk of selective reporting for both comparisons (Figure 7 and 8), and smaller in studies at low risk of deviations from intended interventions for the face-to-face comparison.

**Figure 6.**
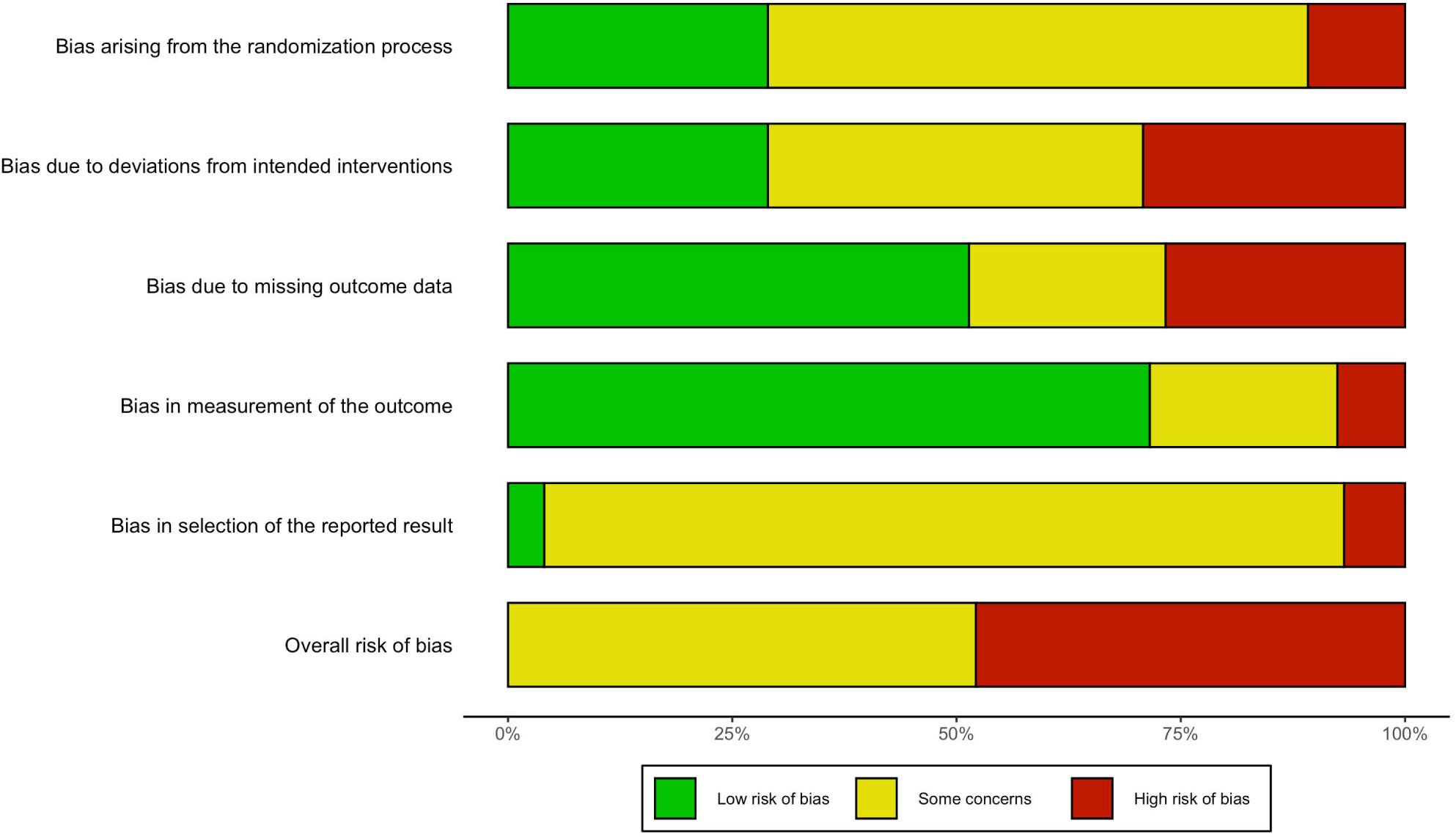
Risk of bias summary plot.

**Figure 7.**
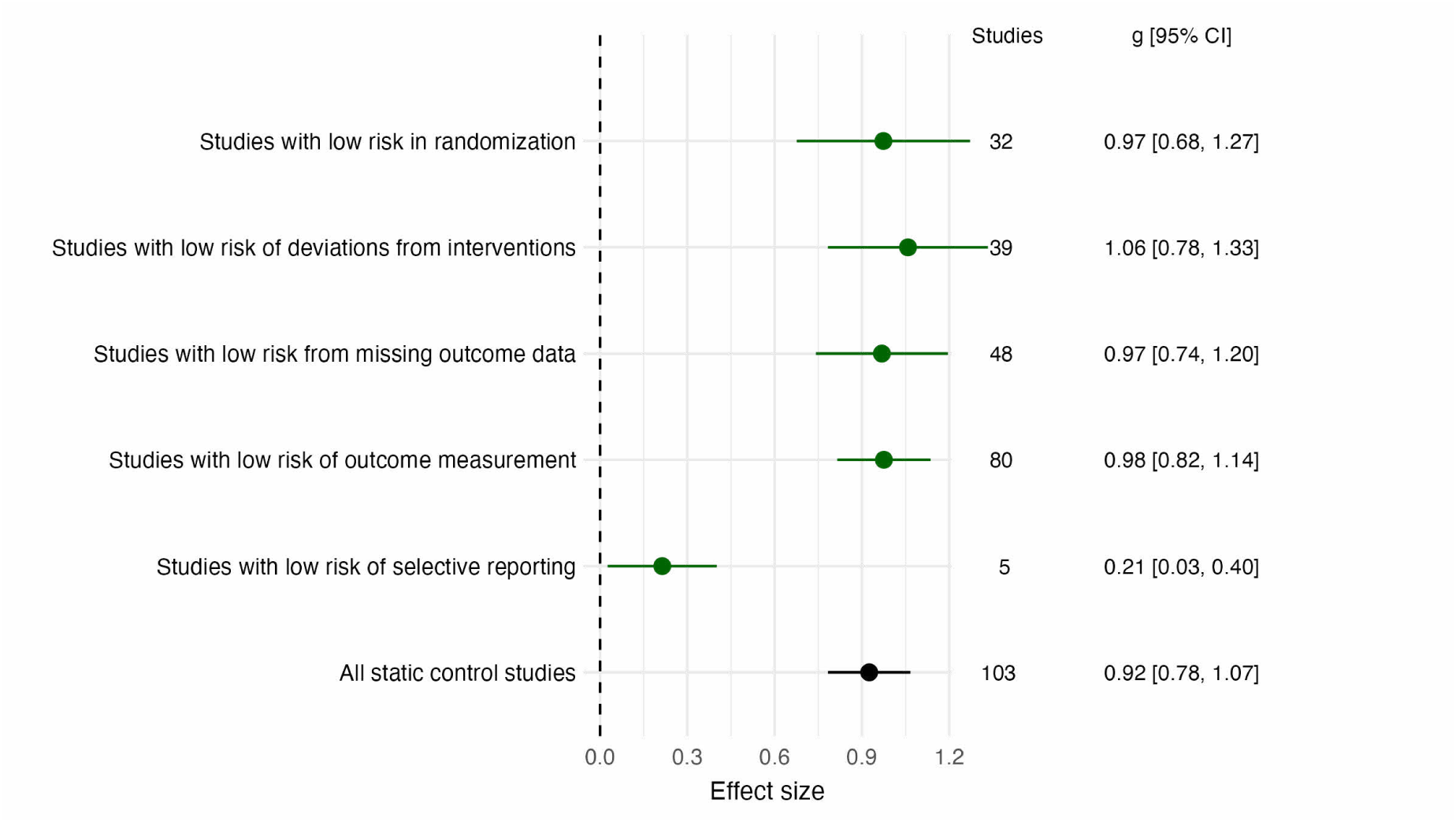
Sensitivity analysis for static comparison studies with low risk of bias vs main analysis.

**Figure 8.**
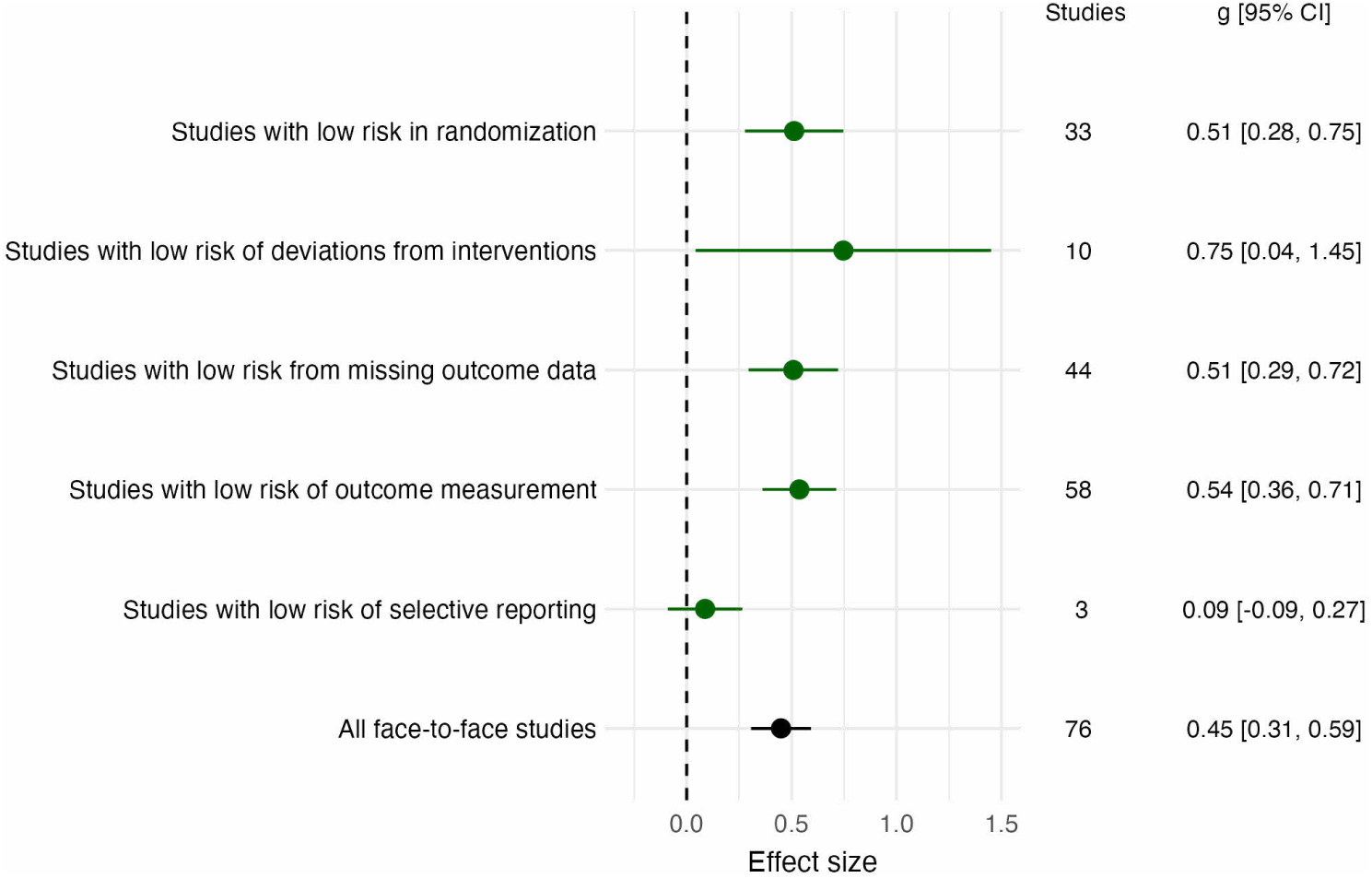
Sensitivity analysis for face-to-face comparison studies with low risk of bias vs main analysis.

#### Publication Bias

We plotted effect sizes against standard errors using a funnel plot (Figure 9). These plots show some asymmetry driven by the outliers described above (*g* > 2.5). The multi-level meta-analytic Egger’s test detected the presence of small-study effects for online versus static controls (*QM*(df = 1) = 22.47, *p* < 0.001) and online versus face to face (*QM*(df = 1) = 7.23, *p* = 0.007). Removing the outliers (*g* > 2.5), there was lower asymmetry detected by Egger’s test for comparisons with static controls (*QM*(df = 1) = 12.91, *p* < 0.001) and non-significant asymmetry for comparisons with face-to-face instruction (*QM*(df = 1) = 1.36, *p* = 0.243).

**Figure 9.**
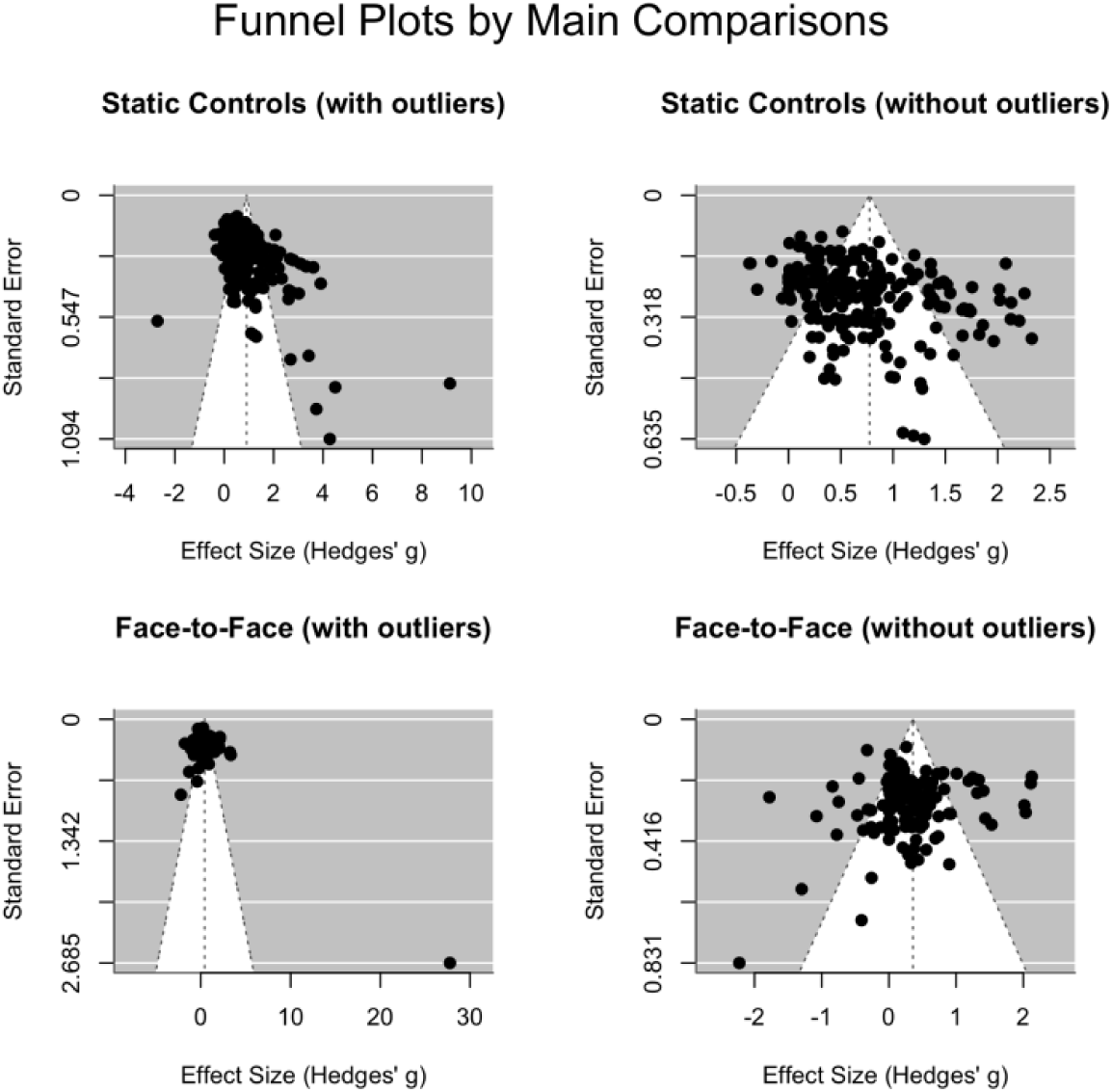
Funnel plots moderated by main effects analyses.

#### Certainty Assessment

We assessed the certainty of the evidence for each outcome using the GRADE framework (Table 2). Overall, comparisons against face-to-face controls was ‘low’ certainty for knowledge acquisition. Skill acquisition was ‘very low’, as were both knowledge and skills outcomes comparing online learning to static controls. Evidence was generally downgraded for not having studies of low-risk of bias, and high levels of unexplained heterogeneity.

**Table 2.** Summary of Findings for Certainty Assessment for the Effects of Online Professional Learning on Healthcare Professionals’ Knowledge and Skill Acquisition. **Population**: Practising healthcare professionals in any clinical setting, **Intervention:** Online professional learning, **Comparison:** Static controls (waitlist, no intervention, paper-based materials) or face-to-face controls

| GRADE domain assessment |  |  |  |  |  |  |  |  |
| --- | --- | --- | --- | --- | --- | --- | --- | --- |
| Outcome | Studies (effects) | Hedges' g (95% CI) | Risk of bias | Inconsistency | Indirectness | Imprecision | Publication bias | Certainty of evidence |
| Static Controls (waitlist, no intervention paper-based materials) |  |  |  |  |  |  |  |  |
| Knowledge acquisition | 78 (153) | 0.78 (0.67, 0.90) | Serious<sup>a</sup> | Serious<sup>b</sup> | Not serious | Not serious | Serious<sup>c</sup> | ⊕○○○<br>Very low |
| Skill acquisition | 25 (53) | 0.73 (0.59, 0.88) | Serious<sup>a</sup> | Serious<sup>b</sup> | Not serious | Not serious | Serious<sup>c</sup> | ⊕○○○<br>Very low |
| Face-to-Face Controls |  |  |  |  |  |  |  |  |
| Knowledge acquisition | 62 (94) | 0.46 (0.33, 0.59) | Serious<sup>a</sup> | Serious<sup>b</sup> | Not serious | Not serious | Not serious | ⊕⊕○○<br>Low |
| Skill acquisition | 25 (58) | 0.20 (0.04, 0.36) | Serious<sup>a</sup> | Serious<sup>b</sup> | Not serious | Some concerns<sup>d</sup> | Not serious | ⊕○○○<br>Very low |

*Note.* All included studies were randomised controlled trials; evidence therefore begins at High certainty and is downgraded for concerns in each domain. A positive Hedges’ g indicates online learning outperforms the comparator. Certainty ratings: ⊕⊕○○ Low; ⊕○○○ Very low. <sup>a</sup> Downgraded one level due to risk of bias.
<sup>b</sup> Downgraded one level due to inconsistency. <sup>c</sup> Downgraded one level due to publication bias. <sup>d</sup> Downgraded one level due to imprecision.

## Discussion

We found 171 randomised controlled trials meeting our criteria for examining online learning for healthcare professionals’ knowledge and skills acquisition. Overall, online learning demonstrated large positive effects when compared to static controls (e.g., waitlists, self-directed textbook reading, electronic or print materials) and small but significant positive effects compared to face-to-face instruction. Results seemed consistent regardless of the learning design, intervention characteristics, professional group, clinician experience, or clinical setting. Overall, the findings suggest that online programmes may be more effective than traditional methods for improving healthcare professionals’ knowledge and skills, with the added advantages of scalability and flexibility for healthcare professional development^20–23^.

Prior reviews of online learning were small ^e.g.,14^ or focused on a single skill or context^25,26,30^. In contrast, our findings show that online learning is effective across diverse professional groups, experience levels, practice settings, and income classifications (Figures 3 and 5). These consistent findings across different contexts establish online learning as a reliable approach for healthcare professional development. Our broad inclusion criteria also gave us the power to test a large number of potential moderators. Few of these moderators demonstrated consistent effects. Given the heterogeneity we observed, this means differences between effect sizes are driven by differences we did not or could not assess from the study’s report (e.g., the institutional support for online learning^49^, or the degree of end-user co-design^50^).

A few significant moderators emerged. Benefits of online learning were particularly pronounced in lower-middle income countries when compared against static controls, likely because there were fewer high-quality alternatives to the training provided by the trials in these settings^18,19^. In these contexts, geography and limited infrastructure are key barriers to professional development^22,26^, and online learning’s digital delivery, flexible access, and low cost^21,23^ directly address these constraints. In high-income countries, where professionals have greater resources and access to professional learning, these advantages are less distinctive and the marginal benefit of any single programme is smaller by comparison. Even so, online learning improved knowledge and skill acquisition compared to static control resources and face-to-face delivery across all income settings, suggesting its value is universal but its potential to reduce global health inequalities is greatest where access to professional development is most limited^22^.

Another moderator of note was the type of outcome assessed, observed only when comparing online learning to face-to-face instruction. Online learning produced larger effects for knowledge acquisition than for skills acquisition, which aligns with evidence that structured digital content is well suited to transferring declarative knowledge. More notably, online learning still outperformed face-to-face instruction for skills acquisition, challenging the assumption that physical presence is the primary driver of procedural development. Other systematic reviews have shown online video leads to bigger improvements in skill acquisition than knowledge development^51^, arguably because video can easily show authentic skill demonstrations. Benefits of online learning may reflect the inherent variability of face-to-face clinical training, where the learning experience depends on available cases, supervisors, and setting, meaning practitioners may develop inconsistent levels of competence. Online learning, by contrast, delivers standardised instruction consistently across learners regardless of context, which can mitigate the inconsistencies of traditional professional learning.

Interactivity level did not moderate online learning effectiveness in either comparison, contrary to the Interactive, Constructive, Active, and Passive (ICAP) framework’s prediction that constructive and interactive engagement produce greater learning gains than passive modes^27^. Two explanations are plausible. Conceptually, the lack of a moderating effect may stem from the fact that all engagement levels (i.e., active, constructive, interactive, and passive) produced significant positive effects with tightly clustered effect sizes (Figure 3 & 5). When every delivery format consistently outperforms control conditions, specific differences between categories become difficult to isolate, suggesting that the primary driver of benefit is the online learning itself rather than the specific level of interactivity. Methodologically, interactivity was categorised from published intervention descriptions rather than direct measurement of learner experience, which may have reduced sensitivity to detect meaningful differences. Overall, the findings suggest that even less resource-intensive, passive online learning formats provide a robust and scalable alternative to traditional professional development for advancing professional knowledge and skills.

It remains unclear whether any learning design principles such as feedback, scaffolding, simulation, and group collaboration, enhance the effects of online learning on knowledge and skill acquisition. This may be an artefact of our design’s sensitivity to detect between-study moderators. Studies in our review covered different content for different audiences, measuring different outcomes. It might therefore be difficult to detect effects of studies using specific learning design principles. Robust assessments of these learning design principles exist, for example, where the same training is conducted with and without online videos^51^. Learning designers should use design principles supported by existing systematic reviews, such as letting learners collaborate^28,29^, use quizzes^52,53^, design clear multimedia^54^, scaffold the training^55^ or provide authentic simulations^56^.

### Limitations of the included research

Several limitations of the included studies and the available evidence should be considered when interpreting these findings. When we conducted sensitivity analyses for each risk of bias domain, effect sizes appeared robust. However, a major limitation of this review is that none of the included studies were classified as having a low overall risk of bias (see Figure 6), primarily because most studies did not have clear blinding of participants, did not follow intention-to-treat principles, incompletely concealed allocation, or lacked pre-registration. While sensitivity analyses indicated no significant differences in effect sizes between studies rated as ‘low’ risk and the main analysis for most domains (see Figure 7 & 8), effects were smaller when controlling for selective reporting. Few studies pre-registered their protocols, and when they did, effects were lower. This effect is not uncommon, which is why pre-registration is becoming the norm^57^. In combination with funnel plot asymmetry and significant Eggers test, the data raises concerns of selective reporting and publication bias. Authors of future trials should be expected to prospectively register their protocols prior to data collection.

We initially planned to conduct a Two-Stage Structural Equation Model (TSSEM) to assess whether knowledge and skill acquisition mediate the relationship between online learning and clinical practice. However, our analysis was limited by available data: none reported the necessary correlation between participant’s learning and behaviour change. Due to the insufficient number of studies reporting this correlation, we were unable to proceed with the planned analysis. Reporting this correlation in the future would allow for useful tests of the hypothesised causal structure.

Our analysis did not include studies conducted in low-income settings because no eligible studies from these contexts were identified, despite studies in any language being eligible. While conclusions could be drawn for studies of other income levels, the absence of data from low-income settings leaves a gap in our understanding for the most resource-constrained contexts. Challenges unique to these settings, such as limited technology infrastructure and internet accessibility, are likely to influence outcomes and must be considered for future research. Addressing this gap is essential, as online learning has high potential to overcome traditional barriers to education and professional development in low-income countries. Future studies should prioritise evaluating online learning in these environments to better inform scalable and inclusive strategies.

### Limitations of this review

Our review provides strong evidence for the efficacy of online learning, yet policymakers require more than effectiveness evidence to justify adoption. Online learning is assumed to be cost-efficient at scale, as once developed, programmes can reach large numbers of learners at low marginal cost. In practice, however, upfront investment in content development, platform infrastructure, and facilitation can be substantial, and these costs may only be recovered when programmes are delivered at sufficient scale. Future reviews should examine cost alongside effectiveness through formal economic evaluations to better inform investment decisions across diverse healthcare settings.

Finally, we used intention-to-treat effect sizes where available, but we did not assess course completion rates, which serve as an important indicator of the overall success and sustainability of online learning programmes^22^. Barriers to engagement (e.g., time constraints, technical difficulties, or lack of motivation) were not examined but remain critical for understanding how to optimise online learning experiences^22^. Addressing these factors in future research will help identify strategies to improve course completion and engagement rates, particularly in diverse and resource-limited healthcare environments.

### Conclusion

Our findings support online learning as an effective tool for professional learning, delivering strong positive effects on knowledge and skills acquisition for all health professional groups. Online learning not only outperforms static controls, but also proves more effective than traditional face-to-face learning. Results suggest that modern online platforms can replicate many benefits of face-to-face learning by incorporating features like virtual simulations, real-time feedback, and interactive case studies. Yet, we found that no single design element guarantees greater knowledge and skill acquisition outcomes.

Healthcare organisations can use online learning to deliver high-quality training across diverse settings, including resource-limited environments. Its scalability, accessibility, and cost-efficiency make it a practical solution for professional development, ensuring healthcare professionals can acquire essential knowledge and skills without relying on face-to-face instruction. Despite challenges such as publication bias, the evidence supports online learning as a reliable and effective method for enhancing healthcare professionals’ knowledge and skill acquisition.

## Contributors statement

Conceputalisation: SG, MN, SM, DC, SD, OF; Methodology: SG, MN, GS, LM, KO, LC, HF, KH, JD, TS, JC, JG, SM, DC, OF, SD; Analysis: SG, MN; Writing – original draft preparations: SG, MN; Writing – review and editing; SG, MN, GS, KO, LC, HF, KH, JD, TS, JG, SM, DC, OF, SD, JC, LM; Supervision: MN, SM, DC, SD, OF.

## Data Availability

Data and code for reproducing analyses are available at

https://osf.io/46zav/?view_only=b1b46e241a9d4ee691d587c6c74a1ebe

## Acknowledgements

During the preparation of this manuscript, the authors used ChatGPT (OpenAI) and Claude (Anthropic) to support language editing and improve readability. The authors also used Claude Code and Codex to support drafting and checking code. All AI-assisted outputs were reviewed and edited by the authors, who take full responsibility for the content of the publication.

## Data sharing statement

Data and code for reproducing analyses are available at https://osf.io/46zav/?view_only=b1b46e241a9d4ee691d587c6c74a1ebe

