## Supplementary Files 1-7 for "Effects of online professional learning on healthcare professionals’ knowledge and skill acquisition: A systematic review and meta-analysis": Supplementary 4_Exclusion_Table.pdf

### Supplementary File 3

*Consensus Reasons for Exclusion*

| Title | Authors | Year | Journal | Volume | Issue | Pages | DOI | Exclusion Reason |
|---|---|---|---|---|---|---|---|---|
| 27. Strengthening Parent - Physician Communication: a Randomized Delayed Start Crossover Study on Empathic Communication Training | Taff, Heather T.; Gilkerson, Linda; Osta, Amanda; Seo-Lee, Alisa; Schwartz, Alan; Barnes, Michelle | 2020 | Academic pediatrics | 20 | 7 | e15-e15 | 10.1016/j.acap.2020.06.048 | Not randomised |
| 322 A randomized trial of a simulation-based resuscitation curriculum for maternal cardiac arrest | Shields, A.; Battistelli, J.; Annis, K.; Minard, C.; Thomson, B.; Nielsen, P. D. | 2024 | American Journal of Obstetrics and Gynecology | 230 | 1 Supplement | S183 | https://dx.doi.org/10.1016/j.ajog.2023.11.344 | Wrong intervention (no online learning) |
| 382. A Multifaceted Intervention to Reduce Blood Culture Contamination in Medical and Surgical Wards: a Cluster Randomized Controlled Trial | Sayabovorn, N.; Chongtrakool, P.; Rachakhom, S.; Kaewsomnuck, S.; Wejsuwanmanee, R.; Khamphimabood, C.; Jitmuang, A. | 2023 | Open Forum Infectious Diseases | 10 | | S237 | 10.1093/ofid/ofad500.452 | Not enough data for analysis |
| 3D Virtual Reality Smartphone Training for Chemotherapy Drug Administration by Non-oncology Nurses: A Randomized Controlled Trial | Wang, Chin-Yun; Lu, Chi-Yu; Yang, Su-Yueh; Tsai, Shu-Chun; Huang, Tsai-Wei | 2022 | Frontiers in medicine | 9 | | 889125 | 10.3389/fmed.2022.889125 | Wrong intervention (no online learning) |
| A Blended Gaming COVID-19 Training System (BGCTS) With WHO Guidelines for Staff in Residential Care Homes | Nct | 2021 | https://clinicaltrials.gov/show/NCT04783025 | | | | | No data in abstract, and no PDF available |
| A Clinical Process Support System for Primary Care to Address Family Stress | Nct | 2018 | https://clinicaltrials.gov/show/NCT03700697 | | | | | No data in abstract, and no PDF available |
| A Comparison of Educational Interventions for the Asthma Guidelines | Lee, J.; Baptist, A.; Sales, A.; Sabhapathy, G. | 2024 | Journal of Allergy and Clinical Immunology | 153 | 2 Supplement | AB262 | https://dx.doi.org/10.1016/j.jaci.2023.11.839 | Intervention <10 mins |
| A Heart Healthy Intervention Improved Tobacco Screening Rates and Cessation Support in Primary Care Practices | Kowitt, Sarah D.; Goldstein, Adam O.; Cykert, Samuel | 2022 | Journal of prevention (2022) | 43 | 3 | 375-386 | 10.1007/s10935-022-00672-5 | Not randomised |
| A KT intervention including the evidence alert system to improve clinician's evidence-based practice behavior--a cluster randomized controlled trial | Campbell, Lanie; Novak, Iona; McIntyre, Sarah; Lord, Sarah | 2013 | Implementation science : IS | 8 | | 132 | 10.1186/1748-5908-8-132 | Wrong intervention (no online learning) |

| Title | Authors | Year | Journal | Volume | Issue | Pages | DOI | Exclusion Reason |
|---|---|---|---|---|---|---|---|---|
| A Multi-centered Feedback and Education Intervention Designed to Reduce Inappropriate Transthoracic Echocardiograms | Nct | 2014 | https://clinicaltrials.gov/show/NCT02038101 | | | | | Duplicate |
| A Novel Web-Based Substance Use Disorder Curriculum for Hospice Interdisciplinary Teams (TH106) | Langmann, G.; Rothenberger, S.; Sheehan, L.; Merlin, J.; Childers, J. | 2022 | Journal of pain and symptom management | 63 | 5 | 781- | 10.1016/j.jpainsymman.2022.02.199 | No knowledge, skills, or clinical outcome |
| A Provider-based Approach to Address Racial Disparities in Lupus Clinical Trial Participation | Wanty, Nicole I.; Cooper, Dexter L.; Simkus, Andrew; Twombly, Eric C.; McCalla, Sheryl; Holtz, S. Kristen D.; Langford, Aisha T.; Cozart, Thometta; Gorlitsky, Barry; Moore, Catherine; Culton, Donna; Richardson, Christopher T.; Wardrop, Richard M., 3rd; Newcomb, Jeff; Aranow, Cynthia; Lim, Sam; Anandarajah, Allen; Sheikh, Saira Z. | 2022 | Arthritis care & research | | | | 10.1002/acr.25054 | Not healthcare clinical practice |
| A Randomised Controlled Trial of an online mental health education program for physician supervisors | Actrn | 2019 | https://trialsearch.who.int/Trial2.aspx?TrialID=ACTRN12619001496101 | | | | | No original data |
| A Randomized Controlled Trial of the Effects of Online Pain Management Education on Primary Care Providers | Trudeau, Kimberlee J.; Hildebrand, Cristina; Garg, Priyanka; Chiauzzi, Emil; Zacharoff, Kevin L. | 2017 | Pain medicine (Malden, Mass.) | 18 | 4 | 680-692 | 10.1093/pm/pnw271 | Comparison group online |
| A Randomized Educational Intervention Trial to Determine the Effect of Online Education on the Quality of Resident-Delivered Care | Dolan, Brigid M.; Yialamas, Maria A.; McMahon, Graham T. | 2015 | Journal of graduate medical education | 7 | 3 | 376-381 | 10.4300/JGME-D-14-00571.1 | Wrong intervention (no online learning) |
| A Randomized Trial of External Practice Support to Improve Cardiovascular Risk Factors in Primary Care | Parchman, Michael L.; Anderson, Melissa L.; Dorr, David A.; Fagnan, Lyle J.; O'Meara, Ellen S.; Tuzzio, Leah; Penfold, Robert B.; Cook, Andrea J.; Hummel, Jeffrey; Conway, Cullen; Cholan, Raja; Baldwin, Laura-Mae | 2019 | Annals of family medicine | 17 | Suppl 1 | S40-S49 | 10.1370/afm.2407 | Wrong intervention (no online learning) |

| Title | Authors | Year | Journal | Volume | Issue | Pages | DOI | Exclusion Reason |
|---|---|---|---|---|---|---|---|---|
| A Statewide Randomized Controlled Trial to Compare Three Models for Implementing Parent Child Interaction Therapy | Herschell, Amy D.; Kolko, David J.; Scudder, Ashley T.; Taber-Thomas, Sarah M.; Schaffner, Kristen F.; Hart, Jonathan A.; Mrozowski, Stanley J.; Hiegel, Shelley A.; Iyengar, Satish; Metzger, Aaron; Jackson, Carrie B. | 2021 | Journal of clinical child and adolescent psychology : the official journal for the Society of Clinical Child and Adolescent Psychology, American Psychological Association, Division 53 | | | Jan-17 | 10.1080/15374416.2021.2001745 | Not healthcare professionals |
| A Statewide Randomized Controlled Trial to Compare Three Models for Implementing Parent Child Interaction Therapy | Herschell, Amy D.; Kolko, David J.; Scudder, Ashley T.; Taber-Thomas, Sarah M.; Schaffner, Kristen F.; Hart, Jonathan A.; Mrozowski, Stanley J.; Hiegel, Shelley A.; Iyengar, Satish; Metzger, Aaron; Jackson, Carrie B. | 2023 | Journal of clinical child and adolescent psychology : the official journal for the Society of Clinical Child and Adolescent Psychology, American Psychological Association, Division 53 | 52 | 6 | 780-796 | https://dx.doi.org/10.1080/15374416.2021.2001745 | Not healthcare professionals |
| A Statewide Trial to Compare Three Training Models for Implementing an Evidence-based Treatment (EBT) | Nct | 2015 | https://clinicaltrials.gov/show/NCT02543359 | | | | | Duplicate |
| A Teachable Moment Communication Process for smoking cessation talk: description of a group randomized clinician-focused intervention | Flocke, Susan A.; Antognoli, Elizabeth; Step, Mary M.; Marsh, Sybil; Parran, Theodore; Mason, Mary Jane | 2012 | BMC health services research | 12 | | 109 | 10.1186/1472-6963-12-109 | Wrong intervention (no online learning) |
| A Trial of a Video Game Intervention to Recalibrate Physician Heuristics | Nct | 2016 | https://clinicaltrials.gov/show/NCT02857348 | | | | | Comparison group online |
| A Trial of the Implementation of iFOBT in General Practice | Nct | 2014 | https://clinicaltrials.gov/show/NCT02308384 | | | | | No original data |
| A Web-Based Education Program About Primary Palliative Care for Heart Failure: A Study Protocol of Wait-Listed Randomized Controlled Trial | Togashi, S.; Wakabayashi, R.; Takehara, A.; Higashitsuji, A.; Ikarashi, A.; Nakashima, N.; Tanaka, N.; Nakano, N.; Shibata, T.; Oishi, S.; Sakashita, A. | 2025 | The Journal of cardiovascular nursing | 40 | 1 | 31-38 | 10.1097/JCN.0000000000001120 | No data in abstract, and no PDF available |
| A brief educational intervention to increase communication about complementary and alternative medicine (CAM) in community oncology settings | Cohen, L.; Urbauer, D.; Fisch, M.; Fellman, B.; Hough, H.; Miller, J.; Lanzotti, V. J.; Whisnant, M.; Weiss, M.; Fellenz, L.; Bury, M. J.; Kokx, P.; Finn, K. T.; Daily, M.; Parker, P. A. | 2012 | Journal of Clinical Oncology | 30 | 15 SUPPL. 1 | | | No data in abstract, and no PDF available |

| Title | Authors | Year | Journal | Volume | Issue | Pages | DOI | Exclusion Reason |
|---|---|---|---|---|---|---|---|---|
| A brief evidence-based curriculum can improve internal medicine residents' ability to successfully manage irritable bowel syndrome and reduce unnecessary diagnostic testing | Yeboah-Korang, A.; Kumar, S.; Gandhi, S.; Jedel, S.; Brown, M. | 2014 | American Journal of Gastroenterology | 109 | SUPPL. 2 | S540 | https://dx.doi.org/10.1038/ajg.2014.282 | Wrong intervention (no online learning) |
| A chlamydia education and training program for general practice nurses: reporting the effect on chlamydia testing uptake | Wood, Anna; Braat, Sabine; Temple-Smith, Meredith; Lorch, Rebecca; Vaisey, Alaina; Guy, Rebecca; Hocking, Jane | 2021 | Australian journal of primary health | 27 | 1 | 36-42 | 10.1071/PY20056 | Wrong intervention (no online learning) |
| A cluster randomized trial on improving nurses' detection and management of elder abuse and neglect (I-NEED): study protocol | Loh, Debbie Ann; Choo, Wan Yuen; Hairi, Noran Naqiah; Othman, Sajaratulnisah; Mohd Hairi, Farizah; Mohd Mydin, Fadzilah Hanum; Jaafar, Siti Nur Illiani; Tan, Maw Pin; Mohd Ali, Zainudin; Abdul Aziz, Suriyati; Ramli, Rohaya; Mohamad, Rosmala; Lal Mohammad, Zaiton; Hassan, Norlela; Brownell, Patricia; Bulgiba, Awang | 2015 | Journal of advanced nursing | 71 | 11 | 2661-2672 | 10.1111/jan.12699 | No original data |
| A cluster-randomised trial of staff education to improve the quality of life of people with dementia living in residential care: the DIRECT study | Beer, Christopher; Horner, Barbara; Flicker, Leon; Scherer, Samuel; Lautenschlager, Nicola T.; Bretland, Nick; Flett, Penelope; Schaper, Frank; Almeida, Osvaldo P. | 2011 | PloS one | 6 | 11 | e28155 | 10.1371/journal.pone.0028155 | Wrong intervention (no online learning) |
| A community-based trial of an online intimate partner violence CME program | Short, Lynn M.; Surprenant, Zita J.; Harris, John M., Jr. | 2006 | American journal of preventive medicine | 30 | 2 | 181-185 | 10.1016/j.amepre.2005.10.012 | Not enough data for analysis |
| A comparative study of the effect of triage training by role-playing and educational video on the knowledge and performance of emergency medical service staffs in Iran | Aghababaeian, H.; Sedaghat, S.; Tahery, N.; Moghaddam, A. S.; Maniei, M.; Bahrami, N.; Ahvazi, L. A. | 2013 | Prehospital and Disaster Medicine | 28 | 6 | 605-609 | 10.1017/S1049023X13008911 | Wrong intervention (no online learning) |
| A comparison between lecture and videotape inservice for certified nursing assistants in skilled nursing facilities | Brooks, P. A.; Renvall, M. J.; Bulow, K. B.; Ramsdell, J. W. | 2000 | Journal of the American Medical Directors Association | 1 | 5 | 191-196 | | Wrong intervention (no online learning) |
| A comparison between the impacts of lecturing and flipped classrooms in virtual learning on triage nurses' knowledge and professional capability: an experimental study | Javadi, Mostafa; Gheshlaghi, Majid; Bijani, Mostafa | 2023 | BMC nursing | 22 | 1 | 205 | https://dx.doi.org/10.1186/s12912-023-01353-2 | Comparison group online |

| Title | Authors | Year | Journal | Volume | Issue | Pages | DOI | Exclusion Reason |
|---|---|---|---|---|---|---|---|---|
| A comparison of educational interventions to impact behavioral intent toward pressure ulcer prevention among nurses on medical surgical units | Russell-Babin, Kathleen | 2015 | | 76 | | | | Wrong intervention (no online learning) |
| A comparison of rapid cycle deliberate practice and traditional reflective debriefing on interprofessional team performance | Colman, N.; Wiltrakis, S. M.; Holmes, S.; Hwu, R.; Iyer, S.; Goodwin, N.; Mathai, C.; Gillespie, S.; Hebbar, K. B. | 2024 | BMC medical education | 24 | 1 | 122 | 10.1186/s12909-024-05101-1 | Wrong intervention (no online learning) |
| A complex intervention to reduce avoidable hospital admissions in nursing homes: a research programme including the BHiRCH-NH pilot cluster RCT | Downs, Murna; Blighe, Alan; Carpenter, Robin; Feast, Alexandra; Froggatt, Katherine; Gordon, Sally; Hunter, Rachael; Jones, Liz; Lago, Natalia; McCormack, Brendan; Marston, Louise; Nurock, Shirley; Panca, Monica; Permain, Helen; Powell, Catherine; Rait, Greta; Robinson, Louise; Woodward-Carlton, Barbara; Wood, John; Young, John; Sampson, Elizabeth | 2021 | | | | | 10.3310/pgfar09020 | Not healthcare professionals |
| A controlled quality improvement trial to reduce the use of physical restraints in older hospitalized adults | Enns, Echo; Rhemtulla, Rishma; Ewa, Vivian; Fruetel, Karen; Holroyd-Leduc, Jayna M. | 2014 | Journal of the American Geriatrics Society | 62 | 3 | 541-545 | 10.1111/jgs.12710 | Wrong intervention (no online learning) |
| A dementia care training using mobile e-learning with mentoring support for home care workers: a controlled study | Su, Hsin-Feng; Koo, Malcolm; Lee, Wen-Li; Sung, Huei-Chuan; Lee, Ru-Ping; Liu, Wen- I. | 2021 | BMC geriatrics | 21 | 1 | 126 | 10.1186/s12877-021-02075-3 | Not healthcare professionals |
| A driving in dementia decision tool: preliminary analysis | Rapoport, M. J.; Sarracini, C. Z.; Rozmovits, L.; Kiss, A.; Grigoriev, I.; Taylor, R.; Herrmann, N.; Mulsant, B. H.; Cameron, D.; Frank, C.; et al. | 2016 | Alzheimer's & dementia | 12 | 7 | P789- | | Wrong intervention (no online learning) |
| A geographical cluster randomised stepped wedge study of continuing medical education and cancer diagnosis in general practice | Toftegaard, Berit Skjødeberg; Bro, Flemming; Vedsted, Peter | 2014 | Implementation science : IS | 9 | | 159 | 10.1186/s13012-014-0159-z | Wrong intervention (no online learning) |

| Title | Authors | Year | Journal | Volume | Issue | Pages | DOI | Exclusion Reason |
|---|---|---|---|---|---|---|---|---|
| A multifaceted intervention to improve blood pressure control: The Guideline Adherence for Heart Health (GLAD) study | Bonds, Denise E.; Hogan, Patricia E.; Bertoni, Alain G.; Chen, Haiying; Clinch, C. Randall; Hiott, Ann E.; Rosenberger, Erica L.; Goff, David C. | 2009 | American heart journal | 157 | 2 | 278-284 | 10.1016/j.ahj.2008.09.021 | Wrong intervention (no online learning) |
| A multifaceted intervention to increase chlamydia testing in australian general practice | Hocking, J.; Poznanski, S.; Vaisey, A.; Walker, J.; Wood, A.; Lewis, D.; Guy, R.; Temple-Smith, M. | 2011 | Sexually transmitted infections | 87 | | A199- | 10.1136/sextrans-2011-050108.232 | No original data |
| A novel educational approach for improving medication-related problems in community pharmacies | Al Mazrouei, Nadia; Ibrahim, Rana M.; Al Meslamani, Ahmad Z.; Mohamed Ibrahim, Osama | 2022 | Research in social & administrative pharmacy : RSAP | 18 | 3 | 2510-2516 | 10.1016/j.sapharm.2021.04.017 | Wrong intervention (no online learning) |
| A novel interactive online tool | Amell, F.; Solomon, J.; Zikmund-Fisher, B.; Boroda, K.; Zisman Ilani, Y.; Park, C.; Thibaud, S. M.; Jagannath, A. D.; Yick, F.; Shah, T.; Miller, T.; Reddy, R.; Cho, J.; Pong, M.; Mukundan, S.; E, M. Izrachi; Ceniceros, A.; Assafin, M.; Lari, H.; Ye, K.; Le Francois, D. | 2016 | Journal of General Internal Medicine | 31 | 2 SUPPL. 1 | S916 | | No original data |
| A novel resident clinical dashboard highlights significant trends in clinical performance that can enhance resident feedback and create targeted educational interventions | Sun, J.; Chung, A. S.; Genes, N.; Li, K.; Peng, P.; Apakama, D.; Loo, G.; Shah, K.; Shearer, P.; Richardson, L. D. | 2018 | Academic emergency medicine | 25 | | S292- | 10.1111/acem.13433 | Not randomised |
| A phase III wait-listed rct of a novel targeted inter-professional clinical education iintervention to improve cancer patients' reported pain outcomes: protocol | Phillips, J. L.; Lovell, M.; Davidson, P.; Boyle, F.; Lam, L.; McCaffrey, N.; Heneka, N.; Shaw, T. | 2018 | Palliative medicine | 32 | 1 | 130- | 10.1177/0269216318769196 | No data in abstract, and no PDF available |
| A pragmatic trial seeking to implement an improved model of care for people with insomnia and obstructive sleep apnoea (OSA) within an Australian primary care setting, in order to increase access to evidence-based therapies | Actrn | 2020 | https://trialsearch.who.int/Trial2.aspx?TrialID=ACTRN12620001075976 | | | | | No original data |
| A preliminary efficacy and feasibility of an obstructive sleep apnea educational intervention in Oman | Al Mezeini, Khamis Abdallah | 2018 | | 79 | | | | Comparison group online |

| Title | Authors | Year | Journal | Volume | Issue | Pages | DOI | Exclusion Reason |
|---|---|---|---|---|---|---|---|---|
| A primary care Web-based Intervention Modeling Experiment replicated behavior changes seen in earlier paper-based experiment | Treweek, Shaun; Francis, Jill J.; Bonetti, Debbie; Barnett, Karen; Eccles, Martin P.; Hudson, Jemma; Jones, Claire; Pitts, Nigel B.; Ricketts, Ian W.; Sullivan, Frank; Weal, Mark; MacLennan, Graeme | 2016 | Journal of clinical epidemiology | 80 | | 116-122 | 10.1016/j.jclinepi.2016.07.008 | Intervention <10 mins |
| A randomised control trial to test an educational innovation of diabetes care in general practice | Actrn | 2012 | https://trialsearch.who.int/Trial2.aspx?TrialID=ACTRN12612000747820 | | | | | No original data |
| A randomised controlled trial investigating the efficacy of online continuing education for health professionals to improve the management of Chronic Fatigue Syndrome (CFS) | Actrn | 2016 | https://trialsearch.who.int/Trial2.aspx?TrialID=ACTRN12616000296437 | | | | | No original data |
| A randomized clinical trial to test the effectiveness of a multifaceted intervention to increase HPV vaccination rates | Isrctn | 2021 | https://trialsearch.who.int/Trial2.aspx?TrialID=ISRCTN55473884 | | | | | No original data |
| A randomized controlled study of an e-learning program (YURAIKU-PRO) for public health nurses to support parents with severe and persistent mental illness and their family members | Kageyama, Masako; Koide, Keiko; Saita, Ryotaro; Iwasaki-Motegi, Riho; Ichihashi, Kayo; Nemoto, Kiyotaka; Sakae, Setsuko; Yokoyama, Keiko | 2022 | BMC nursing | 21 | 1 | 342 | 10.1186/s12912-022-01129-0 | No knowledge, skills, or clinical outcome |
| A randomized controlled trial of a multilevel intervention to increase colorectal cancer screening among Latino immigrants in a primary care facility | Aragones, Abraham; Schwartz, Mark D.; Shah, Nirav R.; Gany, Francesca M. | 2010 | Journal of general internal medicine | 25 | 6 | 564-567 | 10.1007/s11606-010-1266-4 | Not healthcare professionals |
| A randomized controlled trial of an educational intervention on Hellenic nursing staff's knowledge and attitudes on cancer pain management | Patiraki, Elisabeth I.; Papathanassoglou, Elizabeth D. E.; Tafas, Cheryl; Akarepi, Vasiliki; Katsaragakis, Stelios G.; Kampitsi, Anjuleta; Lemonidou, Chrysoula | 2006 | European journal of oncology nursing : the official journal of European Oncology Nursing Society | 10 | 5 | 337-352 | 10.1016/j.ejon.2005.07.006 | Wrong intervention (no online learning) |
| A randomized trial comparing two models of web-based training in cognitive-behavioral therapy for substance abuse counselors | Weingardt, Kenneth R.; Cucciare, Michael A.; Bellotti, Christine; Lai, Wen Pin | 2009 | Journal of substance abuse treatment | 37 | 3 | 219-227 | 10.1016/j.jsat.2009.01.002 | Not healthcare professionals |

| Title | Authors | Year | Journal | Volume | Issue | Pages | DOI | Exclusion Reason |
|---|---|---|---|---|---|---|---|---|
| A randomized trial to reduce the prevalence of depression and self-harm behavior in older primary care patients | Almeida, Osvaldo P.; Pirkis, Jane; Kerse, Ngaire; Sim, Moira; Flicker, Leon; Snowdon, John; Draper, Brian; Byrne, Gerard; Goldney, Robert; Lautenschlager, Nicola T.; Stocks, Nigel; Alfonso, Helman; Pfaff, Jon J. | 2012 | Annals of family medicine | 10 | 4 | 347-356 | 10.1370/afm.1368 | Wrong intervention (no online learning) |
| A randomized, controlled, investigator-blinded, multicenter, single-cross-over interventional pilot study to investigate the effect of virtual reality enhanced education on rheumatoid arthritis for healthcare professionals on knowledge, illness perception | Drks | 2020 | https://trialsearch.who.int/Trial2.aspx?TrialID=DRKS00021507 | | | | | No data in abstract, and no PDF available |
| A serious game for education of primary care physicians on insulin therapy: early results of a randomized controlled trial | Diehl, L. A.; Ferreira, V. H.; Souza, R. M.; Gordan, P. A.; Esteves, R. Z.; Coelho, I. C. M. | 2015 | Diabetes | 64 | | A188- | 10.2337/db15742931 | No original data |
| A simple heuristic for Internet-based evidence search in primary care: a randomized controlled trial | Eberbach, Andreas; Becker, Annette; Rochon, Justine; Finkemeler, Holger; Wagner, Achim; Donner-Banzhoff, Norbert | 2016 | Advances in medical education and practice | 7 | | 433-441 | 10.2147/AMEP.S78385 | Not healthcare clinical practice |
| A simulation-based nursing education of psychological first aid for adolescents exposed to hazardous chemical disasters | Kim, Hye-Won; Choi, Yun-Jung | 2022 | BMC medical education | 22 | 1 | 93 | 10.1186/s12909-022-03164-6 | Wrong intervention (no online learning) |
| A spaced education curriculum to improve bone health care by internal medicine residents | Dolan, B. M.; McMahon, G. T.; Yialamas, M. | 2012 | Journal of General Internal Medicine | 27 | SUPPL. 2 | S545 | | No data in abstract, and no PDF available |
| A technology-enhanced intervention to reduce the duration of untreated psychosis through rapid identification and engagement | Niendam, T.; Loewy, R.; Ragland, D.; Lesh, T.; Skymba, H.; Savill, M.; Pierce, K.; Fedechko, T.; Delucchi, K.; Goldman, H.; et al. | 2017 | Schizophrenia bulletin | 43 | | S98- | | Not healthcare professionals |
| A theory-based adaptive E-learning program aimed at increasing intentions to provide brief behavior change counseling: Randomized controlled trial | Fontaine, Guillaume; Cossette, Sylvie | 2021 | Nurse education today | 107 | | 105112 | 10.1016/j.nedt.2021.105112 | Not healthcare professionals |
| A training intervention to improve information management in primary care | Schifferdecker, Karen E.; Reed, Virginia A.; Homa, Karen | 2008 | Family medicine | 40 | 6 | 423-432 | | No knowledge, skills, or clinical outcome |

| Title | Authors | Year | Journal | Volume | Issue | Pages | DOI | Exclusion Reason |
|---|---|---|---|---|---|---|---|---|
| A trial of education, prompts, and opinion leaders to improve prescription of lipid modifying therapy by primary care physicians for patients with ischemic heart disease | Bloomfield, H. E.; Nelson, D. B.; van Ryn, M.; Neil, B. J.; Koets, N. J.; Basile, J. N.; Samaha, F. F.; Kaul, R.; Mehta, J. L.; Bouland, D. | 2005 | Quality & safety in health care | 14 | 4 | 258-263 | 10.1136/qshc.2004.012617 | Wrong intervention (no online learning) |
| A ward-based trial of an online learning resource to reduce medication dose omissions | Bourke, D.; Black, P. | 2010 | Internal Medicine Journal | 40 | SUPPL. 1 | 79 | https://dx.doi.org/10.1111/j.1445-5994.2010.02187.x | Not randomised |
| A web-based course on complementary medicine for medical students and residents improves knowledge and changes attitudes | Cook, David A.; Gelula, Mark H.; Lee, Mark C.; Bauer, Brent A.; Dupras, Denise M.; Schwartz, Alan | 2007 | Teaching and learning in medicine | 19 | 3 | 230-238 | 10.1080/10401330701366325 | Not randomised |
| ADHD awareness in primary care | Isrctn | 2019 | https://trialsearch.who.int/Trial2.aspx?TrialID=ISRCTN45400501 | | | | | No data in abstract, and no PDF available |
| Achieving Lumbar Epidural Block Competency in Inexperienced Trainees after a Structured Epidural Teaching Model: A Randomized, Single Blind, Prospective Comparison of CUSUM Learning Curves | Scorzoni, Marco; Gonnella, Gian Luigi; Capogna, Emanuele; Velardo, Matteo; Giuri, Pietro Paolo; Ciancia, Mariano; Capogna, Giorgio; Draisci, Gaetano | 2022 | Anesthesiology research and practice | 2022 | | 1738783 | 10.1155/2022/1738783 | Wrong intervention (no online learning) |
| Active implementation of a computerized cpg for major depression in primary care: 18-month follow-up | Cavero, M.; Monreal, J. A.; Cardoner, N.; Moreno, M. D.; Bellerino, E.; Perez-Sola, V.; Palao, D. | 2018 | European psychiatry | 48 | | S415-S416 | 10.1016/j.eurpsy.2017.12.023 | No knowledge, skills, or clinical outcome |
| Adapting the design of a Web-based decision support clinical trial during the COVID-19 pandemic | Meline, Jessica; Prigge, Jason M.; Dye, Debbie; Rieder, Julie; Asan, Onur; Chhatre, Sumedha; Fraenkel, Liana; Kravetz, Jeffrey D.; Rodriguez, Keri L.; Whittle, Jeff; Kaminstein, Dana; Schapira, Marilyn M. | 2021 | Trials | 22 | 1 | 734 | 10.1186/s13063-021-05700-z | No original data |
| Adaptive instruction and learner interactivity in online learning: A randomized trial | Warner, David O.; Nolan, Margaret; Garcia-Marcinkiewicz, Annery; Schultz, Caleb; Warner, Matthew A.; Schroeder, Darrell R.; Cook, David A. | 2020 | Advances in Health Sciences Education | 25 | 1 | 95-109 | 10.1007/s10459-019-09907-3 | Comparison group online |

| Title | Authors | Year | Journal | Volume | Issue | Pages | DOI | Exclusion Reason |
|---|---|---|---|---|---|---|---|---|
| Addressing cardiovascular complications and heart rate in HF: Effect of online CME | Spyropoulos, J.; Healy, C. S. | 2016 | Journal of Cardiac Failure | 22 | Supplement 8 | S102 | | Not randomised |
| Addressing gaps in care for patients with rare cancers and blood disorders: The impact of a collaborative digital education initiative | Turell, W.; Ackbarali, T.; Del Nido, E. L.; Pucci, K.; O'Hara, R.; Kowalski, K.; Hopper, J.; Palma, J. | 2021 | Journal of Clinical Oncology | 39 | 15 SUPPL | | https://dx.doi.org/10.1200/JCO.2021.39.15_suppl.e23005 | Not randomised |
| Adequacy of antidepressant treatment in primary care: preliminary results after implementing a computerised guide for major depression | Cavero, M.; Moreno, M.; Monreal, J.; Labad, J.; Cardoner, N.; Palao, D. | 2015 | European neuropsychopharmacology | 25 | | S448 | | Wrong intervention (no online learning) |
| Adjunctive Team Enhanced Intervention to Improve Suicide Prevention Evidence-Based Practices in Primary Care | Nct | 2020 | https://clinicaltrials.gov/show/NCT04606173 | | | | | No data in abstract, and no PDF available |
| Advance Care Planning in Primary Care: a Cluster Randomized Clinical Trial | Nct | 2021 | https://clinicaltrials.gov/show/NCT04867005 | | | | | No data in abstract, and no PDF available |
| An Educational Intervention for Acute Dizziness Care: A Randomized, Vignette-based Study | Meurer, William J.; Johnson, Patricia; Brown, Devin; Tsodikov, Alexander; Rowell, Brigid; Fagerlin, Angela; Telian, Steven A.; Damschroder, Laura; An, Lawrence C.; Morgenstern, Lewis B.; Kerber, Kevin A. | 2019 | Otology & neurotology : official publication of the American Otological Society, American Neurotology Society [and] European Academy of Otology and Neurotology | 40 | 8 | e830-e838 | 10.1097/MAO.0000000000002338 | Wrong intervention (no online learning) |
| An Educational Intervention to Improve Statin Use: Cluster RCT at the Primary Care Level in Argentina | Gulayin, Pablo E.; Lozada, Alfredo; Beratarrechea, Andrea; Gutierrez, Laura; Poggio, Rosana; Chaparro, Raúl Martín; Santero, Marilina; Masson, Walter; Rubinstein, Adolfo; Irazola, Vilma | 2019 | American journal of preventive medicine | 57 | 1 | 95-105 | 10.1016/j.amepre.2019.02.018 | Wrong intervention (no online learning) |
| An Evaluation of A Web-Based Crisis Management Training Program for Nurse Managers: the Case of The COVID-19 Crisis | Alan, H.; Harmanci Seren, A. K.; Eskin Bacaksiz, F.; Gungor, S.; Bilgin, O.; Baykal, U. | 2023 | Disaster medicine and public health preparedness | 17 | | e358 | 10.1017/dmp.2023.8 | Not healthcare clinical practice |
| An educational intervention on drug use in nursing homes improves health outcomes resource utilization and reduces inappropriate drug prescription | García-Gollarte, Fermín; Baleriola-Júlvez, José; Ferrero-López, Isabel; Cuenllas-Díaz, Álvaro; Cruz-Jentoft, Alfonso J. | 2014 | Journal of the American Medical Directors Association | 15 | 12 | 885-891 | 10.1016/j.jamda.2014.04.010 | Wrong intervention (no online learning) |

| Title | Authors | Year | Journal | Volume | Issue | Pages | DOI | Exclusion Reason |
|---|---|---|---|---|---|---|---|---|
| An external pilot cluster randomised controlled trial of a theory-based intervention to improve appropriate polypharmacy in older people in primary care (PolyPrime): study protocol | Rankin, Audrey; Cadogan, Cathal A.; Barry, Heather E.; Gardner, Evie; Agus, Ashley; Molloy, Gerard J.; Gorman, Ashleigh; Ryan, Cristín; Leathem, Claire; Maxwell, Marina; Gormley, Gerard J.; Ferrett, Alan; McCarthy, Pat; Fahey, Tom; Hughes, Carmel M. | 2021 | Pilot and feasibility studies | 7 | 1 | 77 | 10.1186/s40814-021-00822-2 | Wrong intervention (no online learning) |
| An implementation science approach to intervention fidelity in the SPEACS-2 clinical trial | Tate, J. A.; Sereika, S. M.; Thompson, D.; Devido, J.; Sawicki, J.; Angus, D. C.; Barnato, A. E.; Happ, M. B. | 2013 | American Journal of Respiratory and Critical Care Medicine | 187 | Meeting Abstracts | | | Not randomised |
| An innovative programme for training in maternal and newborn care | Woods, D. L. | 1999 | Seminars in Neonatology | 4 | 3 | 209-216 | https://dx.doi.org/10.1016/S1084-2756%2899%2990090-8 | Wrong intervention (no online learning) |
| An interactive online education program improves knowledge of ECG monitoring: findings from the pulse trial | Funk, M.; May, J. L.; Stephens, K.; Fennie, K. P.; Chang, P. S.; Winkler, C.; Feder, S. L.; Drew, B. J. | 2013 | Circulation | 128 | 22 SUPPL. 1 | | | No data in abstract, and no PDF available |
| An interdisciplinary knowledge translation intervention in long-term care: study protocol for the vitamin D and osteoporosis study (ViDOS) pilot cluster randomized controlled trial | Kennedy, Courtney C.; Ioannidis, George; Giangregorio, Lora M.; Adachi, Jonathan D.; Thabane, Lehana; Morin, Suzanne N.; Crilly, Richard G.; Marr, Sharon; Josse, Robert G.; Lohfeld, Lynne; Pickard, Laura E.; King, Susanne; van der Horst, Mary-Lou; Campbell, Glenda; Stroud, Jackie; Dolovich, Lisa; Sawka, Anna M.; Jain, Ravi; Nash, Lynn; Papaioannou, Alexandra | 2012 | Implementation science : IS | 7 | | 48 | 10.1186/1748-5908-7-48 | Duplicate |
| An internet training to reduce assaults in long-term care | Irvine, Blair; Billow, Molly B.; Gates, Donna M.; Fitzwater, Evelyn L.; Seeley, John R.; Bourgeois, Michelle | 2012 | Geriatric nursing (New York, N.Y.) | 33 | 1 | 28-40 | 10.1016/j.gerinurse.2011.10.004 | Not healthcare clinical practice |

| Title | Authors | Year | Journal | Volume | Issue | Pages | DOI | Exclusion Reason |
|---|---|---|---|---|---|---|---|---|
| An intervention to increase physicians' use of adherence-enhancing strategies in managing hypercholesterolemic patients | Casebeer, L. L.; Klapow, J. C.; Centor, R. M.; Stafford, M. A.; Renkl, L. A.; Mallinger, A. P.; Kristofco, R. E. | 1999 | Academic medicine : journal of the Association of American Medical Colleges | 74 | 12 | 1334-1339 | 10.1097/00001888-199912000-00018 | Wrong intervention (no online learning) |
| An online insulin pump and CGM curriculum for trainees: Tekno T1D | Marks, B.; Waldman, G.; Stafford, D. E.; Garvey, K.; Nagler, J.; Wolfsdorf, J. I. | 2019 | Diabetes | 68 | Supplement 1 | | https://dx.doi.org/10.2337/db19-336-OR | Comparison group online |
| An online intervention for carers to manage behavioral symptoms in motor neuron disease (MiNDToolkit): a randomized parallel multi-center feasibility trial | Mioshi, E.; Grant, K.; Flanagan, E.; Heal, S.; Copsey, H.; Gould, R. L.; Hammond, M.; Shepstone, L.; Ashford, P. A. | 2024 | Amyotrophic lateral sclerosis & frontotemporal degeneration | 25 | 5‐6 | 506‐516 | 10.1080/21678421.2024.2350658 | Not healthcare professionals |
| An online spaced-education game for global continuing medical education: a randomized trial | Kerfoot, B. Price; Baker, Harley | 2012 | Annals of surgery | 256 | 1 | 33-38 | 10.1097/SLA.0b013e31825b3912 | Comparison group online |
| Assessing physician knowledge on optimal management of pulmonary arterial hypertension: effectiveness of on-demand education | Negron, J.; Sendaydiego, A.; Hoeper, M. | 2018 | Journal of the American College of Cardiology | 71 | 11 | | 10.1016/S0735-1097(18)33191-7 | Not randomised |
| Assessing the effectiveness of a LGBT cultural competency training for oncologists: study protocol for a randomized pragmatic trial | Seay, Julia; Hernandez, Eryk N.; Pérez-Morales, Jaileene; Quinn, Gwendolyn P.; Schabath, Matthew B. | 2022 | Trials | 23 | 1 | 314 | 10.1186/s13063-022-06274-0 | No original data |
| Assessing the impact of an eHealthResp intervention on antibiotics use | Isrctn | 2022 | https://trialsearch.who.int/Trial2.aspx?TrialID=ISRCTN57694130 | | | | | No data in abstract, and no PDF available |
| Assessing the impact of immersive simulation on clinical performance during actual in-hospital cardiac arrest with CPR-sensing technology: A randomized feasibility study | Weidman, Elizabeth K.; Bell, George; Walsh, Deborah; Small, Stephen; Edelson, Dana P. | 2010 | Resuscitation | 81 | 11 | 1556-1561 | 10.1016/j.resuscitation.2010.05.021 | Wrong intervention (no online learning) |
| Assessment of education and computerized decision support interventions for improving transfusion practice | Rothschild, Jeffrey M.; McGurk, Siobhan; Honour, Melissa; Lu, Linh; McClendon, Aubre A.; Srivastava, Priya; Churchill, W. Hallowell; Kaufman, Richard M.; Avorn, Jerry; Cook, E. Francis; Bates, David W. | 2007 | Transfusion | 47 | 2 | 228-239 | 10.1111/j.1537-2995.2007.01093.x | Wrong intervention (no online learning) |

| Title | Authors | Year | Journal | Volume | Issue | Pages | DOI | Exclusion Reason |
|---|---|---|---|---|---|---|---|---|
| Audit-based education to implement NICE clinical recommendations of things 'not to do' in people with cardiometabolic diseases: a cluster randomised trial | Isrctn | 2022 | https://trialsearch.who.int/Trial2.aspx?TrialID=ISRCTN16678243 | | | | | No original data |
| Barriers and facilitators to implementing a continuing medical education intervention in a primary health care setting | Reis, Teresa; Faria, Inês; Serra, Helena; Xavier, Miguel | 2022 | BMC health services research | 22 | 1 | 638 | 10.1186/s12913-022-08019-w | Wrong intervention (no online learning) |
| Barriers to care linkage and educational impact on unnecessary MASLD referrals | Lee, J. H.; Yoon, E. L.; Oh, J. H.; Kim, K.; Ahn, S. B.; Jun, D. W. | 2024 | Frontiers in Medicine | 11 | | | 10.3389/fmed.2024.1407389 | Not randomised |
| Bcma-Directed Therapy for Multiple Myeloma: Effectiveness of Online Education in Improving Knowledge and Competence in Practical Application | Harvey-Jones, V.; Koekkoek, S.; Vandenbroucque, J.; Mateos, M. V. | 2022 | HemaSphere | 6 | Supplement 3 | 3470 | https://dx.doi.org/10.1097/01.HS9.0000852292.38263.b8 | Not randomised |
| Bcma-directed therapy in multiple myeloma: Effect of online education on clinician knowledge and confidence | Taylor, A.; Koekkoek, S.; Vandenbroucque, J.; Popat, R. | 2021 | HemaSphere | 5 | SUPPL 2 | 786 | https://dx.doi.org/10.1097/HS9.0000000000000566 | Not randomised |
| Benefits of simulation as a teaching tool in postgraduate education in emergency medicine “cardiac arrest scenario” | Khrouf, M.; Sandid, H.; Hraiech, K.; Boukadida, F.; Bel haj, Y.; Mezgar, Z.; Methamem, M. | 2019 | Critical care (London, England) | 23 | | | 10.1186/s13054-019-2358-0 | Wrong intervention (no online learning) |
| Blood Pressure Control in African Americans | Nct | 2005 | https://clinicaltrials.gov/show/NCT00233220 | | | | | No original data |
| Blood pressure screening in pediatric primary care settings | Wall, T. C. | 2010 | Journal of investigative medicine | 58 | 2 | 439‑440 | 10.231/JIM.0b013e3182820c55 | Comparison group online |
| Brief Online Training (BOLT) for Routine Outcome Monitoring | Nct | 2021 | https://clinicaltrials.gov/show/NCT05041517 | | | | | No original data |
| Brief intervention and FASD training for physicians: Results of a randomized trial | Balachova, T. N.; Bonner, B. L.; Isurina, G. L.; Tsvetkova, L. A. | 2010 | Alcoholism: Clinical and Experimental Research | 34 | SUPPL. 3 | 87A | https://dx.doi.org/10.1111/j.1530-0277.2010.01292-3.x | Wrong intervention (no online learning) |
| Building CAPACITI for Palliative Care in Primary Care: early Results From a Randomized Controlled Trial of an Educational Intervention in Canada | Seow, H.; Burge, F.; Winemaker, S.; Pond, G.; Marshall, D.; Kilbertus, F.; Kortez-Miller, K.; Kelley, M. L.; Stajduhar, K. | 2022 | Palliative medicine | 36 | 1 SUPPL | 38‑ | 10.1177/02692163221093145 | No data in abstract, and no PDF available |
| CONNECT for Better Falls Prevention in VA Community Living Centers | Nct | 2009 | https://clinicaltrials.gov/show/NCT00836433 | | | | | No original data |

| Title | Authors | Year | Journal | Volume | Issue | Pages | DOI | Exclusion Reason |
|---|---|---|---|---|---|---|---|---|
| CONNECT for better fall prevention in nursing homes: results from a pilot intervention study | Colon-Emeric, Cathleen S.; McConnell, Eleanor; Pinheiro, Sandro O.; Corazzini, Kirsten; Porter, Kristie; Earp, Kelly M.; Landerman, Lawrence; Beales, Julie; Lipscomb, Jeffrey; Hancock, Kathryn; Anderson, Ruth A. | 2013 | Journal of the American Geriatrics Society | 61 | 12 | 2150-2159 | 10.1111/jgs.12550 | Not healthcare professionals |
| Can Internet-based education improve physician confidence in dealing with domestic violence? | Harris, John M., Jr.; Kutob, Randa M.; Surprenant, Zita J.; Maiuro, Roland D.; Delate, Thomas A. | 2002 | Family medicine | 34 | 4 | 287-292 | | Not enough data for analysis |
| Can an EASYcare based dementia training programme improve diagnostic assessment and management of dementia by general practitioners and primary care nurses? The design of a randomised controlled trial | Perry, M.; Drasković, I.; van Achterberg, T.; Borm, G. F.; van Eijken, M. I. J.; Lucassen, Pl; Vernooij-Dassen, M. J. F. J.; Olde Rikkert, M. G. M. | 2008 | BMC health services research | 8 | | 71 | 10.1186/1472-6963-8-71 | No original data |
| Can dialectical behavior therapy be learned in highly structured learning environments? Results from a randomized controlled dissemination trial | Dimeff, Linda A.; Woodcock, Eric A.; Harned, Melanie S.; Beadnell, Blair | 2011 | Behavior therapy | 42 | 2 | 263-275 | 10.1016/j.beth.2010.06.004 | Not healthcare professionals |
| Can distance learning improve smoking cessation advice in family practice? A randomized trial | Young, Jane M.; Ward, Jeanette | 2002 | The Journal of continuing education in the health professions | 22 | 2 | 84-93 | 10.1002/chp.1340220204 | Wrong intervention (no online learning) |
| Can emergency nurses' triage skills be improved by online learning? Results of an experiment | Rankin, James A.; Then, Karen L.; Atack, Lynda | 2013 | Journal of emergency nursing | 39 | 1 | 20-26 | 10.1016/j.jen.2011.07.004 | Comparison group online |
| Cancer risk communication with low health literacy patients: a continuing medical education program | Price-Haywood, Eboni G.; Roth, Katherine G.; Shelby, Kit; Cooper, Lisa A. | 2010 | Journal of general internal medicine | 25 Suppl 2 | | S126-S129 | 10.1007/s11606-009-1211-6 | Not enough data for analysis |
| Cardiometabolic management-success of online CME at expanding diabetes management past glycemic control | Larkin, A.; Lacouture, M.; Le, A. | 2018 | Diabetes | 67 | Supplement 1 | A327 | | Not randomised |
| Challenges in teaching clinical trials: The experience of teaching biostatistics in online post-graduate academic courses, target to industrial biometrician | Cavaliere, L.; Perissinotto, E.; Baldi, I.; Barbetta, B.; Gregori, D. | 2017 | Trials | 18 | Supplement 1 | | https://dx.doi.org/10.1186/s13063-017-1902-y | Not randomised |
| Changing Talk Online (CHATO) Study | Nct | 2019 | https://clinicaltrials.gov/show/NCT03849937 | | | | | No original data |

| Title | Authors | Year | Journal | Volume | Issue | Pages | DOI | Exclusion Reason |
|---|---|---|---|---|---|---|---|---|
| Changing provider behaviour to increase nurse visits for obesity in family practice: the 5As Team randomized controlled trial | Campbell-Scherer, Denise L.; Asselin, Jodie; Osunlana, Adedayo M.; Ogunleye, Ayodele A.; Fielding, Sheri; Anderson, Robin; Cave, Andrew; Johnson, Jeffrey A.; Sharma, Arya M. | 2019 | CMAJ open | 7 | 2 | E371-E378 | 10.9778/cmajo.20180165 | Wrong intervention (no online learning) |
| Choosing Words Wisely: a Pilot Study to Effect Communication Change | Groninger, H.; Lott, A. | 2018 | Journal of pain and symptom management | 56 | 6 | e86‑e87 | 10.1016/j.jpainsymman.2018.10.305 | No data in abstract, and no PDF available |
| Clinical Decision Support to Implement ED-initiated Buprenorphine for OUD | Nct | 2018 | https://clinicaltrials.gov/show/NCT03658642 | | | | | Wrong intervention (no online learning) |
| Clinical Simulation Scenario for the acquisition of competence in the management of Postpartum Hemorrhage | hk9vgs, R. B. R. | 2023 | https://trialsearch.who.int/Trial2.aspx?TrialID=RBR-6hk9vgs | | | | | Comparison group online |
| Clinical and Healthcare Improvement through My Health Record usage and Education in General Practice â€“ The CHIME-GP Study | Actrn | 2020 | https://trialsearch.who.int/Trial2.aspx?TrialID=ACTRN12620000010998 | | | | | No original data |
| Clinical evidence continuous medical education: a randomised educational trial of an open access e-learning program for transferring evidence-based information - ICEKUBE (Italian Clinical Evidence Knowledge Utilization Behaviour Evaluation) - study protoc | Moja, Lorenzo; Moschetti, Ivan; Cinquini, Michela; Sala, Valeria; Compagnoni, Anna; Duca, Piergiorgio; Deligant, Christian; Manfrini, Roberto; Clivio, Luca; Satolli, Roberto; Addis, Antonio; Grimshaw, Jeremy M.; Dri, Pietro; Liberati, Alessandro | 2008 | Implementation science : IS | 3 | | 37 | 10.1186/1748-5908-3-37 | No original data |
| Clinical implementation and evaluation of three implementation interventions for a family-oriented care for children of mentally ill parents (ci-CHIMPS) | Drks | 2021 | https://trialsearch.who.int/Trial2.aspx?TrialID=DRKS00026217 | | | | | No data in abstract, and no PDF available |
| Clinical utility of a blood-based protein assay to increase screening of elevated-risk patients for colorectal cancer in the primary care setting | Peabody, John; Paculdo, David; Swagel, Eric; Fugaro, Steven; Tran, Mary | 2017 | Journal of cancer research and clinical oncology | 143 | 11 | 2301-2307 | 10.1007/s00432-017-2469-4 | Intervention <10 mins |
| Cluster randomised controlled trial of a theory-based multiple behaviour change intervention aimed at healthcare professionals to improve their management of type 2 diabetes in primary care | Presseau, Justin; Mackintosh, Joan; Hawthorne, Gillian; Francis, Jill J.; Johnston, Marie; Grimshaw, Jeremy M.; Steen, Nick; Coulthard, Tom; Brown, Heather; Kaner, Eileen; Elovainio, Marko; Sniehotta, Falko F. | 2018 | Implementation science : IS | 13 | 1 | 65 | 10.1186/s13012-018-0754-5 | Wrong intervention (no online learning) |

| Title | Authors | Year | Journal | Volume | Issue | Pages | DOI | Exclusion Reason |
|---|---|---|---|---|---|---|---|---|
| Cluster-randomized controlled intervention study to evaluate the implementation of an evidence-based and patient-centered care concept for the treatment of venous leg ulcers in primary care ("Ulcus Cruris Care") | Drks | 2021 | https://trialsearch.who.int/Trial2.aspx?TrialID=DRKS00026126 | | | | | No data in abstract, and no PDF available |
| Coaching leadership development program for nurses | mpn8qk, R. B. R. | 2024 | https://trialsearch.who.int/Trial2.aspx?TrialID=RBR-2mpn8qk | | | | | No data in abstract, and no PDF available |
| Cognitive De-Biasing and the Assessment of Pediatric Bipolar Disorder | Nct | 2013 | https://clinicaltrials.gov/show/NCT01799291 | | | | | Not healthcare professionals |
| ComOn Coaching: Study protocol of a randomized controlled trial to assess the effect of a varied number of coaching sessions on transfer into clinical practice following communication skills training | Niglio de Figueiredo, Marcelo; Rudolph, Bärbel; Bylund, Carma L.; Goelz, Tanja; Heußner, Pia; Sattel, Heribert; Fritzsche, Kurt; Wuensch, Alexander | 2015 | BMC cancer | 15 | | 503 | 10.1186/s12885-015-1454-z | Wrong intervention (no online learning) |
| Communication Skills Training for Oncology Health Professionals working with Culturally and Linguistically Diverse Patients | Actrn | 2013 | http://www.who.int/trialsearch/Trial2.aspx?TrialID=ACTRN12613000560796 | | | | | Wrong intervention (no online learning) |
| Comparative effectiveness of audit-feedback versus additional physician communication training to improve cancer screening for patients with limited health literacy | Price-Haywood, Eboni G.; Harden-Barrios, Jewel; Cooper, Lisa A. | 2014 | Journal of general internal medicine | 29 | 8 | 1113-1121 | 10.1007/s11606-014-2782-4 | Wrong intervention (no online learning) |
| Comparing Effectiveness of Remote-learning Formats for Resident Physician Didactics | Shinnick, Julia K.; Narvaez, Jennifer L.; Raker, Christina; Brousseau, E. Christine | 2023 | Rhode Island medical journal (2013) | 106 | 1 | 52-57 | | Comparison group online |
| Comparing the effect of electronic and lecture education of pain management on the knowledge, attitude, and practice of nurses: A randomized-controlled trial | Farshbaf-Khalili, Azizeh; Jasemi, Madine; Seyyedzavvar, Atefe | 2021 | Journal of education and health promotion | 10 | | 374 | 10.4103/jehp.jehp_918_20 | Wrong intervention (no online learning) |
| Comparing the effectiveness of different training packages at preparing clinical staff to deploy mechanical chest compression devices | Isrctn | 2016 | https://trialsearch.who.int/Trial2.aspx?TrialID=ISRCTN43049287 | | | | | Wrong intervention (no online learning) |
| Comparing two different schedules of online learning for updated cardiopulmonary resuscitation guidelines in Covid-19 patients: A randomized study | Joshi, Poonam; Das, Smita; Mawar, Shashi; Gopichandran, Lakshmanan; Naik, Nitish; Shariff, Ahamadulla; Garg, Rakesh | 2022 | The National medical journal of India | 35 | 3 | 168-171 | 10.25259/NMJI-35-3-168 | Comparison group online |

| Title | Authors | Year | Journal | Volume | Issue | Pages | DOI | Exclusion Reason |
|---|---|---|---|---|---|---|---|---|
| Comparison of Two Learning Modalities on Continuing Medical Education Consumption and Knowledge Acquisition: A Pilot Randomized Controlled Trial | McEvoy, Matthew D.; Fowler, Leslie C.; Robertson, Amy; Gelfand, Brian J.; Fleming, Geoffrey M.; Miller, Bonnie; Moore, Donald | 2021 | The journal of education in perioperative medicine : JEPM | 23 | 3 | E668 | 10.46374/volxxiii_issue3_mcevoy | Comparison group online |
| Comparison of Web- Versus Classroom-based Basic Ultrasound and Extended Focused Assessment With Sonography for Trauma (EFAST) Training in Two European Hospitals | Nct | 2009 | https://clinicaltrials.gov/show/NCT01040767 | | | | | Duplicate |
| Comparison of Web- versus classroom-based basic ultrasound and extended focused assessment with sonography for trauma training in two European hospitals | Platz, E.; Goldflam, K.; Mennicke, M.; Parisini, E.; Christ, M.; Hohenstein, C. | 2009 | Annals of Emergency Medicine | 54 | 3 SUPPL. 1 | S88 | | Duplicate |
| Comparison of an interactive with a didactic educational intervention for improving the evidence-based practice knowledge of occupational therapists in the public health sector in South Africa: a randomised controlled trial | Buchanan, Helen; Siegfried, Nandi; Jelsma, Jennifer; Lombard, Carl | 2014 | Trials | 15 | | 216 | 10.1186/1745-6215-15-216 | Wrong intervention (no online learning) |
| Comparison of nursing care learning in air evacuation and transport by lecture and e-learning methods | Farshi, M.; Babatabar Darzi, H.; Mahmoudi, H.; Mokhtari Nouri, J. | 2012 | Journal of Military Medicine | 14 | 1 | 27-31 | | Not randomised |
| Comparison of the effect of scenario-based training in face-to-face and virtual methods on nurses` skills in advanced cardiopulmonary resuscitation | Irct20230222057504N, | 2023 | https://trialsearch.who.int/Trial2.aspx?TrialID=IRCT20230222057504N1 | | | | | Not enough data for analysis |
| Comparison of two methods for teaching advanced arrhythmias to nurses | Lamb, M. J.; Henderson, M. C. | 1993 | Journal of continuing education in nursing | 24 | 5 | 221-226 | 10.3928/0022-0124-19930901-08 | Wrong intervention (no online learning) |
| Comparison of two new educational techniques on knowledge of nurses about cerebrovascular accident nursing care in emergency department | Dehghan, Zahra; Alimohammadi, Nasrollah; Mohamadirizi, Shahla | 2022 | Journal of education and health promotion | 11 | | 60 | 10.4103/jehp.jehp_985_20 | Comparison group online |
| Comparison the effect of lecturing method and e-learning program | Irct2013102815203N | 2013 | https://trialsearch.who.int/Trial2.aspx?TrialID=IRCT2013102815203N1 | | | | | No data in abstract, and no PDF available |
| Comparison the effects of two educational methods on knowledge, attitude and practices of Arak physicians about breast cancer | Moshfeghi, K.; Mohammadbeigi, A. | 2010 | Pakistan journal of biological sciences : PJBS | 13 | 18 | 901-905 | 10.3923/pjbs.2010.901.905 | Wrong intervention (no online learning) |

| Title | Authors | Year | Journal | Volume | Issue | Pages | DOI | Exclusion Reason |
|---|---|---|---|---|---|---|---|---|
| Comprehensive approach for hypertension control in low-income populations: rationale and study design for the hypertension control program in Argentina | Mills, Katherine T.; Rubinstein, Adolfo; Irazola, Vilma; Chen, Jing; Beratarrechea, Andrea; Poggio, Rosana; Dolan, Jacquelyn; Augustovski, Federico; Shi, Lizheng; Krousel-Wood, Marie; Bazzano, Lydia A.; He, Jiang | 2014 | The American journal of the medical sciences | 348 | 2 | 139-145 | 10.1097/MAJ.0000000000000298 | No knowledge, skills, or clinical outcome |
| Computer-assisted instruction in AIDS infection control for physicians | Garrett, T. J.; Selnow, Gary; Dobkin, Jay F.; Healton, Cheryl | 1990 | Teaching and Learning in Medicine | 2 | 4 | 215-218 | 10.1080/10401339009539463 | Wrong intervention (no online learning) |
| Constipation challenge: randomized controlled trial of interactive spaced education to improve primary care management of pediatric constipation | Mallon, D.; Fei, L.; Farrell, M.; Iyer, S.; Klein, M. | 2018 | Journal of pediatric gastroenterology and nutrition | 67 | | S13‑S14 | 10.1097/MPG.0000000000002164 | Comparison group online |
| Continuing Medical Education Improves Physician Communication Skills and Increases Likelihood of Pediatric Vaccination: findings from the Pediatric Influenza Vaccination Optimization Trial (PIVOT)—II | Fisher, W. A.; Gilca, V.; Murti, M.; Orth, A.; Garfield, H.; Roumeliotis, P.; Rampakakis, E.; Brown, V.; Yaremko, J.; Van Buynder, P.; et al., | 2023 | Vaccines | 11 | 1 | | 10.3390/vaccines11010017 | No knowledge, skills, or clinical outcome |
| Continuing education curriculum improved primary care physicians knowledge, competence and confidence on treatment of heart failure with reduced ejection fraction | Thevathasan, L.; Sendaydiego, A.; Ashley, N.; Chen, P.; Schoonheim, P. | 2021 | European Journal of Heart Failure | 23 | SUPPL 2 | 294 | https://dx.doi.org/10.1002/ejhf.2297 | Not randomised |
| Cost-effectiveness of educational outreach to primary care nurses to increase tuberculosis case detection and improve respiratory care: economic evaluation alongside a randomised trial | Fairall, Lara; Bachmann, Max O.; Zwarenstein, Merrick; Bateman, Eric D.; Niessen, Louis W.; Lombard, Carl; Majara, Bosielo; English, René; Bheekie, Angeni; van Rensburg, Dingie; Mayers, Pat; Peters, Annatjie; Chapman, Ronald | 2010 | Tropical medicine & international health : TM & IH | 15 | 3 | 277-286 | 10.1111/j.1365-3156.2009.02455.x | Wrong intervention (no online learning) |
| Costs and Cost-Effectiveness of mCME Version 2.0: An SMS-Based Continuing Medical Education Program for HIV Clinicians in Vietnam | Sabin, Lora L.; Mesic, Aldina; Le, Bao Ngoc; Halim, Nafisa; Cao, Chi Thi Hue; Bonawitz, Rachael; Nguyen, Ha Viet; Larson, Anna; Nguyen, Tam Thi Thanh; Le, Anh Ngoc; Gill, Christopher J. | 2022 | Global health, science and practice | 10 | 4 | | 10.9745/GHSP-D-22-00008 | Not randomised |

| Title | Authors | Year | Journal | Volume | Issue | Pages | DOI | Exclusion Reason |
|---|---|---|---|---|---|---|---|---|
| Decreasing Over Screening and Treatment of Cervical Precancers in Young Women | Nct | 2014 | https://clinicaltrials.gov/show/NCT02270021 | | | | | No data in abstract, and no PDF available |
| Design and methods of the Echo WISELY (Will Inappropriate Scenarios for Echocardiography Lessen SignificantlY) study: An investigator-blinded randomized controlled trial of education and feedback intervention to reduce inappropriate echocardiograms | Bhatia, R. Sacha; Ivers, Noah; Yin, Cindy X.; Myers, Dorothy; Nesbitt, Gillian; Edwards, Jeremy; Yared, Kibar; Wadhera, Rishi; Wu, Justina C.; Wong, Brian; Hansen, Mark; Weinerman, Adina; Shadowitz, Steven; Johri, Amer; Farkouh, Michael; Thavendiranathan, Paaladinesh; Udell, Jacob A.; Rambihar, Sherryn; Chow, Chi-Ming; Hall, Judith; Thorpe, Kevin E.; Rakowski, Harry; Weiner, Rory B. | 2015 | American heart journal | 170 | 2 | 202-209 | 10.1016/j.ahj.2015.04.022 | No original data |
| Design of Behavioral Economic Applications to Geriatrics Leveraging Electronic Health Records (BEAGLE): A pragmatic cluster randomized controlled trial | Brown, Tiffany; Rowe, Theresa A.; Lee, Ji Young; Petito, Lucia C.; Chmiel, Ryan; Ciolino, Jody D.; Doctor, Jason N.; Fox, Craig R.; Goldstein, Noah J.; Kaiser, Darren; Linder, Jeffrey A.; Meeker, Daniella; Peprah, Yaw; Persell, Stephen D. | 2022 | Contemporary clinical trials | 112 | | 106649 | 10.1016/j.cct.2021.106649 | Comparison group online |
| Design of a bilevel clinical trial targeting adherence in heart failure patients and their providers: The Congestive Heart Failure Adherence Redesign Trial (CHART) | Mangla, Ashvarya; Doukky, Rami; Richardson, DeJuran; Avery, Elizabeth F.; Dawar, Rebecca; Calvin, James E., Jr.; Powell, Lynda H. | 2018 | American heart journal | 195 | | 139-150 | 10.1016/j.ahj.2017.09.016 | No data in abstract, and no PDF available |
| Design of a randomized clinical trial to improve rates of amblyopia detection in preschool aged children in primary care settings | Wall, Terry C.; Marsh-Tootle, Wendy L.; Crenshaw, Katie; Person, Sharina D.; Datla, Raju; Kristofco, Robert E.; Hartmann, E. Eugenie | 2011 | Contemporary clinical trials | 32 | 2 | 204-214 | 10.1016/j.cct.2010.10.009 | Comparison group online |
| Design of the BiRmingham Early Detection In untREated psyChosis Trial (REDIRECT): cluster randomised controlled trial of general practitioner education in detection of first episode psychosis [ISRCTN87898421] | Tait, Lynda; Lester, Helen; Birchwood, Max; Freemantle, Nick; Wilson, Sue | 2005 | BMC health services research | 5 | 1 | 19 | 10.1186/1472-6963-5-19 | Wrong intervention (no online learning) |

| Title | Authors | Year | Journal | Volume | Issue | Pages | DOI | Exclusion Reason |
|---|---|---|---|---|---|---|---|---|
| Design, development, and evaluation of an online virtual emergency department for training trauma teams | Youngblood, Patricia; Harter, Phillip M.; Srivastava, Sakti; Moffett, Shannon; Heinrichs, Wm LeRoy; Dev, Parvati | 2008 | Simulation in healthcare : journal of the Society for Simulation in Healthcare | 3 | 3 | 146-153 | 10.1097/SIH.0b013e31817bedf7 | Not healthcare professionals |
| Designing tailored Web-based instruction to improve practicing physicians' chlamydial screening rates | Casebeer, Linda; Allison, Jeroan; Spettell, Claire M. | 2002 | Academic medicine : journal of the Association of American Medical Colleges | 77 | 9 | 929 | 10.1097/00001888-200209000-00032 | No original data |
| Designing tailored Web-based instruction to improve practicing physicians' preventive practices | Casebeer, Linda L.; Strasser, Sheryl M.; Spettell, Claire M.; Wall, Terry C.; Weissman, Norman; Ray, Midge N.; Allison, Jeroan J. | 2003 | Journal of medical Internet research | 5 | 3 | e20 | 10.2196/jmir.5.3.e20 | Comparison group online |
| Detection and management of chronic kidney disease and diabetes with e-technology based intervention: analysis of the chronic disease early detection and improved management in primary care project (CD IMPACT] | Jones, J.; Lumsden, N.; Simons, K.; Fernando, S.; Neil, C.; Manski-Mankervis, J. A.; Hamblin, P.; Janus, E.; Nelson, C. | 2019 | Nephrology dialysis transplantation | 34 | | a27‐ | 10.1093/ndt/gfz096.FO058 | Wrong intervention (no online learning) |
| Developing and Evaluating a Continuous Education Program for Healthcare Assistants in Macao: A Cluster-Randomized Trial | Cheong, Pak-Leng; Hsu, Nanly | 2021 | International journal of environmental research and public health | 18 | 9 | | 10.3390/ijerph18094990 | Not healthcare professionals |
| Developing and evaluating interventions to reduce inappropriate prescribing by general practitioners of antibiotics for upper respiratory tract infections: a randomised controlled trial to compare paper-based and web-based modelling experiments | Treweek, Shaun; Ricketts, Ian W.; Francis, Jillian; Eccles, Martin; Bonetti, Debbie; Pitts, Nigel B.; Maclennan, Graeme; Sullivan, Frank; Jones, Claire; Weal, Mark; Barnett, Karen | 2011 | Implementation science : IS | 6 | | 16 | 10.1186/1748-5908-6-16 | Duplicate |
| Developing and evaluating live CME sessions for family physicians on cancer and heredity: an RCT | Ntr | 2012 | https://trialsearch.who.int/Trial2.aspx?TrialID=NTR3323 | | | | | No original data |
| Developing educational program for middle-level public health nurses to create a new program which based on the community health needs: randomized control trial(Second time) | Umin | 2018 | https://trialsearch.who.int/Trial2.aspx?TrialID=JPRN-UMIN000032176 | | | | | No data in abstract, and no PDF available |
| Development and Feasibility Study of Educational Program for Midwives, Nurses, and Public Health Nurses Providing Preconception Care: a Pilot Randomized Controlled Trial | Jprn, Umin | 2023 | https://trialsearch.who.int/Trial2.aspx?TrialID=JPRN-UMIN000051089 | | | | | No data in abstract, and no PDF available |

| Title | Authors | Year | Journal | Volume | Issue | Pages | DOI | Exclusion Reason |
|---|---|---|---|---|---|---|---|---|
| Development and Validation of an Online Independent Training Program for TOR-BSST© Dysphagia Screeners | Nct | 2022 | https://clinicaltrials.gov/show/NCT05379699 | | | | | No data in abstract, and no PDF available |
| Development and evaluation of a pedagogical tool to improve understanding of a quality checklist: a randomised controlled trial | Fourcade, Lola; Boutron, Isabelle; Moher, David; Ronceray, Lucie; Baron, Gabriel; Ravaud, Philippe | 2007 | PLoS clinical trials | 2 | 5 | e22 | 10.1371/journal.pctr.0020022 | Not healthcare professionals |
| Development and evaluation of an online course aiming to train nurses in brief motivational interventions | Isrctn | 2019 | https://trialsearch.who.int/Trial2.aspx?TrialID=ISRCTN32603572 | | | | | No original data |
| Development and randomized controlled trial evaluation of E-learning trainings for professionals | König, Elisa; Maier, Anna; Fegert, Jörg Michael; Hoffmann, Ulrike | 2020 | Archives of public health = Archives belges de sante publique | 78 | 1 | 122 | 10.1186/s13690-020-00465-4 | No knowledge, skills, or clinical outcome |
| Development and randomized-controlled evaluation of a web-based training in evidence-based trauma therapy | Sansen, Lisa M.; Saupe, Laura B.; Steidl, Annika; Fegert, Jörg M.; Hoffmann, Ulrike; Neuner, Frank | 2020 | Professional Psychology: Research and Practice | 51 | 2 | 115-124 | 10.1037/pro000026210.1037/pro0000262.supp (Supplemental) | Not healthcare professionals |
| Development of HIV-assist, an online, educational, clinical decision support tool to guide patient-centered ARV regimen selection | Maddali, M.; Li, J.; Shah, M. | 2018 | Open Forum Infectious Diseases | 5 | Supplement 1 | S404-S405 | https://dx.doi.org/10.1093/ofid/ofy210.1157 | Not randomised |
| Development of a Tailored, Complex Intervention for Clinical Reflection and Communication about Suspected Urinary Tract Infections in Nursing Home Residents | Arnold, Sif H.; Olesen, Julie A.; Jensen, Jette N.; Bjerrum, Lars; Holm, Anne; Kousgaard, Marius B. | 2020 | Antibiotics (Basel, Switzerland) | 9 | 6 | | 10.3390/antibiotics9060360 | Not randomised |
| Development of a Web-Based Course to Maintain Skills in Nurses Trained to Screen for Dysphagia | Nct | 2007 | https://clinicaltrials.gov/show/NCT00570557 | | | | | No data in abstract, and no PDF available |
| Development of a comprehensive surgical pathology website for teaching and self-assessment at the resident and clinical practice level | Wu, C.; Rogers, D.; Olson, G.; Cerilli, L. | 2011 | Laboratory Investigation | 91 | SUPPL. 1 | 133A | | Comparison group online |
| Development of needs assessment and pilot education program for delivery of men's health care via nurse practitioners | Zappavigna, C.; Shamloul, R.; Cagiannos, I.; Gerridzen, R.; Bella, A. J. | 2010 | Journal of Men's Health | 7 | 3 | 329 | https://dx.doi.org/10.1016/j.jomh.2010.09.150 | No data in abstract, and no PDF available |
| Development, acceptability and uptake of an on-line communication skills education program targeting challenging conversations for oncology health professionals related to identifying and responding to anxiety and depression | Shaw, Joanne; Allison, Karen; Cuddy, Jessica; Lindsay, Toni; Grimison, Peter; Shepherd, Heather; Butow, Phyllis; Shaw, Tim; Baychek, Kate; Kelly, Brian | 2022 | BMC health services research | 22 | 1 | 132 | 10.1186/s12913-022-07521-5 | Not randomised |

| Title | Authors | Year | Journal | Volume | Issue | Pages | DOI | Exclusion Reason |
|---|---|---|---|---|---|---|---|---|
| Development, acceptability and uptake of an online anxiety and depression education program for oncology health professionals (ADAPT program) | Shaw, J.; Allison, K.; Cuddy, J.; Lindsay, T.; Grimison, P.; Shepherd, H.; Butow, P.; Shaw, T.; Baychek, K.; Kelly, B. | 2021 | Asia-Pacific Journal of Clinical Oncology | 17 | SUPPL 9 | 154 | https://dx.doi.org/10.1111/ajco.13716 | Not randomised |
| Development, feasibility and acceptability of an online course to teach primary palliative care skills to hepatology providers | Verma, M.; Denofrio, J.; Ramchandran, K.; Taddei, T. H.; Volk, M.; Navarro, V. J. | 2020 | Hepatology (Baltimore, Md.) | 72 | 1 SUPPL | 1156A-1157A | 10.1002/hep.31579 | Not randomised |
| Disseminating behavioural activation for depression via online training: preliminary steps | Hubley, Sam; Woodcock, Eric A.; Dimeff, Linda A.; Dimidjian, Sona | 2015 | Behavioural and cognitive psychotherapy | 43 | 2 | 224-238 | 10.1017/S1352465813000842 | Comparison group online |
| Does accuracy of v lead electrode placement differ based on gender of patient: results of the practical use of the latest standards of electrocardiography (pulse) trial | Davis, L. L.; Funk, M.; Fennie, K. P.; May, J. L.; Stephens, K.; Drew, B. J. | 2016 | Circulation | 134 | | | | Duplicate |
| Does an online diabetes education program for health care professionals improve inpatient diabetes care? | Actrn | 2017 | https://trialsearch.who.int/Trial2.aspx?TrialID=ACTRN12617000762358 | | | | | No knowledge, skills, or clinical outcome |
| Does an outcome-based approach to continuing medical education improve physicians' competences in rational prescribing? | Esmaily, Hamideh M.; Savage, Carl; Vahidi, Rezagoli; Amini, Abolghasem; Dastgiri, Saeed; Hult, Hakan; Dahlgren, Lars Owe; Wahlstrom, Rolf | 2009 | Medical teacher | 31 | 11 | e500-e506 | 10.3109/01421590902803096 | Wrong intervention (no online learning) |
| Does awareness of unconscious associations enhance learning about healthcare disparities? | Sabin, J.; Van Schaik, E.; Lynch, E.; Stoner, S. | 2010 | American Journal of Epidemiology | 171 | SUPPL. 11 | S129 | https://dx.doi.org/10.1093/aje/kwq151 | No original data |
| Does educating nurses with ventilator-associated pneumonia prevention guidelines improve their compliance? | Aloush, Sami M. | 2017 | American journal of infection control | 45 | 9 | 969-973 | 10.1016/j.ajic.2017.04.009 | Wrong intervention (no online learning) |
| Does online professional development for physiotherapists enhance clinical practice and patient outcomes? A mixed methods evaluation | Actrn | 2022 | https://trialsearch.who.int/Trial2.aspx?TrialID=ACTRN12622000123741 | | | | | No original data |
| Drug Exposure Feedback and Education for Nurses' Safety | Nct | 2014 | https://clinicaltrials.gov/show/NCT02283164 | | | | | No original data |
| E-Learning Training to Improve Pediatric Parenteral Nutrition Practice: A Pilot Study in Two University Hospitals | Petit, Laetitia-Marie; Le Pape, Pauline; Delestras, Stephanie; Nguyen, Christina; Marchand, Valerie; Belli, Dominique; Bonnabry, Pascal; Bajwa, Nadia; Fonzo-Christe, Caroline | 2020 | JPEN. Journal of parenteral and enteral nutrition | 44 | 6 | 1089-1095 | 10.1002/jpen.1730 | Not randomised |

| Title | Authors | Year | Journal | Volume | Issue | Pages | DOI | Exclusion Reason |
|---|---|---|---|---|---|---|---|---|
| E-learn CT angiography: A virtual training ground prepares for stroke imaging | Havsteen, I.; Christensen, A. F.; Nielsen, J. K.; Krieger, D.; Christensen, L.; Christensen, H. | 2011 | Stroke | 42 | 3 | e281 | https://dx.doi.org/10.1161/STR.0b013e3182074d9b | Not randomised |
| E-learning courses in epilepsy--concept, evaluation, and experience with the e-learning course "genetics of epilepsies" | Wehrs, Verena Hézser- V.; Pfäfflin, Margarete; May, Theodor W. | 2007 | Epilepsia | 48 | 5 | 872-879 | 10.1111/j.1528-1167.2007.01029.x | Not randomised |
| E-learning education for ultrasound-guided peripheral blocks. A prospective study evaluating individual increment in learning curves during a one-month trial period | Hansen, C.; Worm, B.; Dam, M.; Haase, N.; Poulsen, T.; Bendtsen, T.; Borglum, J. | 2015 | Regional Anesthesia and Pain Medicine | 40 | 5 SUPPL. 1 | e110 | https://dx.doi.org/10.1097/AAP.0000000000000308 | No data in abstract, and no PDF available |
| E-learning for early diagnosis of cerebral palsy: a randomised controlled trial evaluating the effectiveness of physician e-learning interventions on cerebral palsy diagnostic skills, behaviours and practice | Actrn | 2022 | https://trialsearch.who.int/Trial2.aspx?TrialID=ACTRN12622000184774 | | | | | No original data |
| E-learning versus workshops to teach critical appraisal to health professionals: a randomised controlled equivalence study | Isrctn | 2006 | https://trialsearch.who.int/Trial2.aspx?TrialID=ISRCTN47874687 | | | | | No data in abstract, and no PDF available |
| Early identification of sepsis in adults in primary care: A pilot project | Chaney, Heide L. | 2018 | | 79 | | | | Wrong intervention (no online learning) |
| Early intensification with fixed-ratio combination of basal insulin and GLP-1 receptor agonist in T2DM: Impact of online education on primary care physician knowledge and competence | Trier, J.; Griffith, G.; Ampudia-Blasco Javier, F.; McCarthy, R. | 2019 | Diabetes Technology and Therapeutics | 21 | Supplement 1 | A129 | https://dx.doi.org/10.1089/dia.2019.2525.abstracts | Not randomised |
| Educating Nurses About Venous Thromboembolism (VTE) Prevention | Nct | 2014 | https://clinicaltrials.gov/show/NCT02301793 | | | | | Comparison group online |
| Educating Providers About Lung Screening | Nct | 2021 | https://clinicaltrials.gov/show/NCT05064046 | | | | | No original data |
| Educating intensive care unit nurses to use central venous catheter infection prevention guidelines: effectiveness of an educational course | Aloush, Sami | 2018 | Journal of research in nursing : JRN | 23 | 5 | 406-413 | 10.1177/1744987118762992 | Wrong intervention (no online learning) |
| Education and Training Competences in Thoracic Ultrasound | Nct | 2018 | https://clinicaltrials.gov/show/NCT03728491 | | | | | No original data |
| Educational Efficacy Assessment of a Serious Game to Teach Insulin Therapy to Primary Care Physicians | Nct | 2012 | https://clinicaltrials.gov/show/NCT01759953 | | | | | No original data |

| Title | Authors | Year | Journal | Volume | Issue | Pages | DOI | Exclusion Reason |
|---|---|---|---|---|---|---|---|---|
| Educational Intervention to Reduce Outpatient Inappropriate Transthoracic Echocardiograms | Nct | 2013 | https://clinicaltrials.gov/show/NCT01944202 | | | | | Wrong intervention (no online learning) |
| Educational Video's Impact on Knowledge Regarding Cervical Cancer Screening | Nct, | 2023 | https://clinicaltrials.gov/show/NCT05756192 | | | | | Not healthcare professionals |
| Educational intervention to promote the screening of tuberculosis in primary care: randomized clinical trial with assigned clusters | Monegal, A. R. | 2007 | FMC formacion medica continuada en atencion primaria | 14 | 9 | 598- | | Duplicate |
| Educational program for middle-level public health nurses to develop new health services regarding community health needs: protocol for a randomized controlled trial | Yoshioka-Maeda, Kyoko; Katayama, Takafumi; Shiomi, Misa; Hosoya, Noriko | 2018 | BMC nursing | 17 | | 18 | 10.1186/s12912-018-0287-x | No original data |
| Educational program for nursing staff on management of resident-to-resident elder mistreatment (R-REM) | Actrn | 2017 | http://www.who.int/trialsearch/Trial2.aspx?TrialID=ACTRN12617001618347 | | | | | No original data |
| Educational videos for practitioners attending Baby Friendly Hospital Initiative workshops supporting breastfeeding positioning, attachment and hand expression skills: Effects on knowledge and confidence | Wallace, Louise M.; Ma, Yuanying; Qiu, Li Qian; Dunn, Orla M. | 2018 | Nurse education in practice | 31 | | Jul-13 | 10.1016/j.nepr.2018.04.005 | Wrong intervention (no online learning) |
| Effect evaluation of competency-based education (CBE) combined with multi-disciplinary team (MDT) teaching mode in respiratory rehabilitation nursing teaching : A randomized controlled trial | Feng, P.; Wu, J.; Jin, Z.; Cui, J.; Zhang, S.; He, L.; Zhao, H. | 2024 | Nurse education in practice | 76 | | 103896 | 10.1016/j.nepr.2024.103896 | Wrong intervention (no online learning) |
| Effect of Educational Intervention on the Rate of Rarely Appropriate Outpatient Echocardiograms Ordered by Attending Academic Cardiologists: A Randomized Clinical Trial | Dudzinski, David M.; Bhatia, R. Sacha; Mi, Michael Y.; Isselbacher, Eric M.; Picard, Michael H.; Weiner, Rory B. | 2016 | JAMA cardiology | 1 | 7 | 805-812 | 10.1001/jamacardio.2016.2232 | Wrong intervention (no online learning) |
| Effect of Interventional Educational Programs on Intensive Care Nurses' Perception, Knowledge, Attitude, and Practice About Physical Restraints: A Pre-/Postclinical Trial | Ahmadi, Mohamad; Bagheri-Saweh, Mohammad Iraj; Nouri, Bijan; Mohamadamini, Omid; Valiee, Sina | 2019 | Critical care nursing quarterly | 42 | 1 | 106-116 | 10.1097/CNQ.0000000000000244 | Not randomised |
| Effect of Metrics-Based Simulation Training to Proficiency on Procedure Quality and Errors Among Novice Cardiac Device Implanters: the IMPROF Randomized Trial | Mascheroni, J.; Stockburger, M.; Patwala, A.; Mont, L.; Rao, A.; Retzlaff, H.; Garweg, C.; Verbelen, T.; Gallagher, A. G. | 2023 | JAMA network open | 6 | 8 | e2322750 | 10.1001/jamanetworkopen.2023.22750 | No comparison or control |

| Title | Authors | Year | Journal | Volume | Issue | Pages | DOI | Exclusion Reason |
|---|---|---|---|---|---|---|---|---|
| Effect of Monkeypox Nano-Teaching Sessions versus Self-Learning on Nurses' Knowledge, Attitude, and Confidence in Disease Diagnosis and Management | Ibrahim, A. M.; Abd El-kader, R. G.; Ibrahim, A. A. E.; Kishk, D. M. A. | 2024 | International journal of Africa nursing sciences | 20 | | | 10.1016/j.ijans.2024.100713 | Wrong intervention (no online learning) |
| Effect of Web-based diabetes training program on diabetes-related knowledge, attitudes, and skills of health professionals: A randomized controlled trial | Karahan Okuroğlu, Gülten; Ecevit Alpar, Şule | 2019 | Japan journal of nursing science : JJNS | 16 | 2 | 184-193 | 10.1111/jjns.12228 | Not enough data for analysis |
| Effect of a Community-Based Medical Oncology Depression Screening Program on Behavioral Health Referrals Among Patients With Breast Cancer: A Randomized Clinical Trial | Hahn, Erin E.; Munoz-Plaza, Corrine E.; Pounds, Dana; Lyons, Lindsay Joe; Lee, Janet S.; Shen, Ernest; Hong, Benjamin D.; La Cava, Shannon; Brasfield, Farah M.; Durna, Lara N.; Kwan, Karen W.; Beard, David B.; Ferreira, Alexander; Padmanabhan, Aswini; Gould, Michael K. | 2022 | JAMA | 327 | 1 | 41-49 | 10.1001/jama.2021.22596 | Wrong intervention (no online learning) |
| Effect of a Peer Comparison and Educational Intervention on Medical Test Conversation Quality: a Randomized Clinical Trial | Ganguli, I.; Mulligan, K. L.; Chant, E. D.; Lipsitz, S.; Simmons, L.; Sepucha, K.; Rudin, R. S. | 2023 | JAMA network open | 6 | 11 | e2342464 | 10.1001/jamanetworkopen.2023.42464 | Not healthcare professionals |
| Effect of a Virtual Reality Simulation Modality on Registered Nurse Knowledge and Behavior Related to Clostridioides difficile Prevention: an Experimental, Cluster Randomized Controlled Trial | Phillips, J. M.; Harper, M. G.; Brecht, M. L.; DeVon, H. A. | 2024 | Journal for nurses in professional development | | | | 10.1097/NND.0000000000001031 | Comparison group online |
| Effect of a contact-based education intervention on reducing stigma among community health and care staff in Beijing, China: Pilot randomized controlled study | Zhang, Wufang; Henderson, Claire; Magnusdottir, Erla; Chen, Weiran; Ma, Ning; Ma, Hong; Thornicroft, Graham | 2022 | Asian journal of psychiatry | 73 | | 103096 | 10.1016/j.ajp.2022.103096 | Wrong intervention (no online learning) |
| Effect of a web-based course for pediatricians on communication about developmental screening | Marceau, L.; Sices, L.; Ranganathan, G.; Coleman, J.; Zuckerman, B. | 2018 | Pediatrics | 141 | 1 | | 10.1542/peds.141.1-MeetingAbstract.28 | No data in abstract, and no PDF available |
| Effect of a web-based curriculum on primary care practice: The basic skin cancer triage trial | Markova, A.; Weinstock, M. A.; Risica, P.; Shaikh, W.; Kirtania, U.; Ombao, H. | 2012 | Journal of Investigative Dermatology | 132 | SUPPL. 1 | S42 | https://dx.doi.org/10.1038/jid.2012.82 | Comparison group online |

| Title | Authors | Year | Journal | Volume | Issue | Pages | DOI | Exclusion Reason |
|---|---|---|---|---|---|---|---|---|
| Effect of a web-based curriculum on primary care practice: basic skin cancer triage trial | Markova, Alina; Weinstock, Martin A.; Risica, Patricia; Kirtania, Usree; Shaikh, Waqas; Ombao, Hernando; Chambers, Christopher V.; Kabango, Martin L.; Kallail, James K.; Post, Douglas | 2013 | Family medicine | 45 | 8 | 558-568 | | Duplicate |
| Effect of an Evidence-based mHealth Intervention on Cancer Pain Outcomes among People Admitted to an Inpatient Palliative Care Unit: a Wait-listed Randomised Controlled Trial | Phillips, J. L.; Heneka, N.; Lovell, M.; Lam, L.; Boyle, F.; Shaw, T. | 2023 | Palliative medicine | 37 | 1 | 83 | 10.1177/02692163231172891 | Wrong intervention (no online learning) |
| Effect of an emergency department education and empowerment intervention on uncontrolled hypertension in a predominately minority population: The AHEAD2 randomized clinical pilot trial | Prendergast, Heather; Del Rios, Marina; Durazo-Arvizu, Ramon; Escobar-Schulz, Sandra; Heinert, Sara; Jackson, Maya; Gimbar, Renee Petzel; Daviglus, Martha; Lara, Brenda; Khosla, Shaveta | 2021 | Journal of the American College of Emergency Physicians open | 2 | 2 | e12386 | 10.1002/emp2.12386 | Not healthcare professionals |
| Effect of comprehensive oncogenetics training interventions for general practitioners, evaluated at multiple performance levels | Houwink, Elisa J. F.; Muijtjens, Arno M. M.; van Teeffelen, Sarah R.; Henneman, Lidewij; Rethans, Jan Joost; Jacobi, Florijn; van der Jagt, Liesbeth; Stirbu, Irina; van Luijk, Scheltus J.; Stumpel, Connie T. R. M.; Meijers-Heijboer, Hanne E.; van der Vleuten, Cees; Cornel, Martina C.; Dinant, Geert Jan | 2015 | PloS one | 10 | 4 | e0122648 | 10.1371/journal.pone.0122648 | Not randomised |
| Effect of distributing an evidence-based guideline for prevention of osteoporosis on health education programs in municipal health centers: a randomized controlled trial | Nakatani, Yoshimi; Tamaki, Junko; Komatsu, Misa; Iki, Masayuki; Kajita, Etsuko | 2012 | Journal of epidemiology | 22 | 2 | 103-112 | 10.2188/jea.je20110036 | Wrong intervention (no online learning) |
| Effect of patient-specific ratings vs conventional guidelines on investigation decisions in angina: Appropriateness of Referral and Investigation in Angina (ARIA) Trial | Junghans, Cornelia; Feder, Gene; Timmis, Adam D.; Eldridge, Sandra; Sekhri, Neha; Black, Nick; Shekelle, Paul; Hemingway, Harry | 2007 | Archives of internal medicine | 167 | 2 | 195-202 | 10.1001/archinte.167.2.195 | Comparison group online |

| Title | Authors | Year | Journal | Volume | Issue | Pages | DOI | Exclusion Reason |
|---|---|---|---|---|---|---|---|---|
| Effect of the Virtual Simulation Teaching Software of Free Position Childbirth in Midwife Continuing Education: randomized Controlled Trial | ChiCtr | 2020 | https://trialsearch.who.int/Trial2.aspx?TrialID=ChiCTR2000041384 | | | | | No data in abstract, and no PDF available |
| Effect on the process of care of an active strategy to implement clinical guidelines on physiotherapy for low back pain: a cluster randomised controlled trial | Bekkering, G. E.; Hendriks, H. J. M.; van Tulder, M. W.; Knol, D. L.; Hoeijenbos, M.; Oostendorp, R. A. B.; Bouter, L. M. | 2005 | Quality & safety in health care | 14 | 2 | 107-112 | 10.1136/qshc.2003.009357 | Wrong intervention (no online learning) |
| Effective continuing education for breast disease: A randomized trial comparing home study and workshop formats | Young, C.; Chart, P.; Franssen, E.; Tipping, J.; Morris, B.; Davis, D. | 1998 | Journal of Continuing Education in the Health Professions | 18 | 2 | 86-92 | 10.1002/chp.1340180204 | Wrong intervention (no online learning) |
| Effectiveness Of A Multicomponent Implementation Strategy On Increasing Uptake Of USPSTF Hypertension Screening Recommendations In A Primary Care Network: the Embrace Cluster Randomized Trial | Kronish, I. M.; Phillips, E.; Carter, E.; Alcantara, C.; Schwartz, J. E.; Razon, D. T.; Serafini, M. A.; Flatow, J.; Sanchez, J.; Shimbo, D.; et al. | 2022 | Hypertension | 79 | | | 10.1161/hyp.79.suppl_1.P222 | Wrong intervention (no online learning) |
| Effectiveness of Evidence-Based Practice (EBP) Education on Emergency Nurses' EBP Attitudes, Knowledge, Self-Efficacy, Skills, and Behavior: A Randomized Controlled Trial | Koota, Elina; Kääriäinen, Maria; Kyngäs, Helvi; Lääperi, Mitja; Melender, Hanna-Leena | 2021 | Worldviews on evidence-based nursing | 18 | 1 | 23-32 | 10.1111/wvn.12485 | Comparison group online |
| Effectiveness of Individual Feedback and Coaching on Shared Decision-making Consultations in Oncology Care: Protocol for a Randomized Clinical Trial | van Veenendaal, Haske; Peters, Loes J.; Ubbink, Dirk T.; Stubenrouch, Fabienne E.; Stiggelbout, Anne M.; Brand, Paul Lp; Vreugdenhil, Gerard; Hilders, Carina Gjm | 2022 | JMIR research protocols | 11 | 4 | e35543 | 10.2196/35543 | No original data |
| Effectiveness of Interactive Voice Response for COVID-19 Vaccination Training in the Democratic Republic of the Congo | Nct | 2021 | https://clinicaltrials.gov/show/NCT05107479 | | | | | No original data |
| Effectiveness of Multimedia-Based Learning on the Improvement of Knowledge, Attitude, and Behavioral Intention toward COVID-19 Prevention among Nurse Aides in Taiwan: A Parallel-Interventional Study | Hsu, Yi-Min; Chang, Ting-Shan; Chu, Chien-Lun; Hung, Shu-Wen; Wu, Chih-Jung; Yeh, Tzu-Pei; Wang, Jiun-Yi | 2022 | Healthcare (Basel, Switzerland) | 10 | 7 | | 10.3390/healthcare10071206 | Not healthcare professionals |
| Effectiveness of a Game-Based Mobile App for Educating Intensive Critical Care Specialist Nurses in Extracorporeal Membrane Oxygenation Pipeline Preflushing: Quasi-Experimental Trial | Wang, Z.; Gu, R.; Wang, J.; Gai, Y.; Lin, H.; Zhang, Y.; Li, Q.; Sun, T.; Wei, L. | 2023 | JMIR Serious Games | 11 | 1 | | 10.2196/43181 | Not randomised |

| Title | Authors | Year | Journal | Volume | Issue | Pages | DOI | Exclusion Reason |
|---|---|---|---|---|---|---|---|---|
| Effectiveness of a dysphagia e-learning among nurses: a randomized controlled trial | Hendrick, Y.; Roos, C.; Beeckman, A. S. | 2019 | Dysphagia | 34 | 5 | 817- | 10.1007/s00455-019-10009-w | No original data |
| Effectiveness of a multifaceted intervention to improve emergency department care of low back pain: a stepped-wedge, cluster-randomised trial | Coombs, D. M.; Machado, G. C.; Richards, B.; Needs, C.; Buchbinder, R.; Harris, I. A.; Howard, K.; McCaffery, K.; Billot, L.; Edwards, J.; Rogan, E.; Facer, R.; Li, Q.; Maher, C. G. | 2021 | BMJ Qual Saf | 30 | 10 | 825-835 | 10.1136/bmjqs-2020-012337 | Wrong intervention (no online learning) |
| Effectiveness of a serious game for medical education on insulin therapy for diabetes: randomized controlled trial | Diehhl, L. A.; De Souza, R. M.; Gordan, P. A.; Esteves, R. Z.; Coelho, I. C. M. | 2015 | Diabetology & metabolic syndrome | 7 | | 71- | | Not enough data for analysis |
| Effectiveness of a sexual health care training to enhance psychiatric nurses' knowledge, attitude, and self-efficacy: A quasi-experimental study in southern Taiwan | Lu, Mei-Jou; Li, Jin-Biau; Wu, Chia-Yi; Huong, Pham Thi Thu; Hsu, Pei-Chen; Chang, Chiou-Rong | 2024 | Journal of the American Psychiatric Nurses Association | 30 | 1 | 17-29 | 10.1177/10783903211045733 | Wrong intervention (no online learning) |
| Effectiveness of a strategy that uses educational games to implement clinical practice guidelines among Spanish residents of family and community medicine (e-EDUCAGUIA project): a clinical trial by clusters | Del Cura-González, Isabel; López-Rodríguez, Juan A.; Sanz-Cuesta, Teresa; Rodríguez-Barrientos, Ricardo; Martín-Fernández, Jesús; Ariza-Cardiel, Gloria; Polentinos-Castro, Elena; Román-Crespo, Begoña; Escortell-Mayor, Esperanza; Rico-Blázquez, Milagros; Hernández-Santiago, Virginia; Azcoaga-Lorenzo, Amaya; Ojeda-Ruiz, Elena; González-González, Ana I.; Ávila-Tomas, José F.; Barrio-Cortés, Jaime; Molero-García, José M.; Ferrer-Peña, Raul; Tello-Bernabé, María Eugenia; Trujillo-Martín, Mar | 2016 | Implementation science : IS | 11 | | 71 | 10.1186/s13012-016-0425-3 | No original data |

| Title | Authors | Year | Journal | Volume | Issue | Pages | DOI | Exclusion Reason |
|---|---|---|---|---|---|---|---|---|
| Effectiveness of a tailored intervention to reduce antibiotics for urinary tract infections in nursing home residents: a cluster, randomised controlled trial | Arnold, Sif Helene; Nygaard Jensen, Jette; Bjerrum, Lars; Siersma, Volkert; Winther Bang, Christine; Brostrøm Kousgaard, Marius; Holm, Anne | 2021 | The Lancet. Infectious diseases | 21 | 11 | 1549-1556 | 10.1016/S1473-3099(21)00001-3 | Wrong intervention (no online learning) |
| Effectiveness of a training intervention to improve the management of vertigo in primary care: a multicentre cluster-randomised trial, VERTAP | Patiño, Jenniffer Elizabeth Pérez; Moreno, José Lluís Ballvé; Matos, Yolanda Rando; Ortega, Jesús Almeda; Puértolas, Oriol Cunillera; Muñoz, Ricard Carrillo; Balboa, Iván Villar; Compta, Xavier González; Agudelo, Olga Lucía Arias; Muñoz, Sebastiá Calero; Rodríguez, Vanessa Monforte; Cortes, Anna Navarro; Rodríguez, Eva Peguero | 2022 | Trials | 23 | 1 | 608 | 10.1186/s13063-022-06548-7 | No original data |
| Effectiveness of a web-based education program to improve vaccine storage conditions in primary care (Keep Cool): study protocol for a randomized controlled trial | Thielmann, Anika; Viehmann, Anja; Weltermann, Birgitta M. | 2015 | Trials | 16 | | 301 | 10.1186/s13063-015-0824-9 | No original data |
| Effectiveness of a web‐based learning program for promoting local healthcare planning competencies | Yoshioka‐Maeda, Kyoko; Katayama, Takafumi; Fujii, Hitoshi; Shiomi, Misa; Hosoya, Noriko; Mayama, Tatsushi | 2023 | Public Health Nursing | 40 | 5 | 685-695 | 10.1111/phn.13229 | No knowledge, skills, or clinical outcome |
| Effectiveness of aan education program on nurses' knowledge toward prevention of orthopedic wound infection in Baghdad teaching hospitals | Mahdi, Z. S.; Ahmed, S. A. | 2018 | Indian journal of public health research and development | 9 | 8 | 1123‐1128 | 10.5958/0976-5506.2018.00881.1 | Wrong intervention (no online learning) |
| Effectiveness of an Educational Program for Japanese Nurses to Work with Foreign Nurses: a Randomized Controlled Trial | Jprn, Umin | 2023 | https://trialsearch.who.int/Trial2.aspx?TrialID=JPRN-UMIN000051083 | | | | | No data in abstract, and no PDF available |
| Effectiveness of an Interdisciplinary Health Promotion Educational Program in Improving the Quality of Life of Individuals with Fibromyalgia in Brazil: a Randomized Clinical Trial of Amigos De Fibro | Antunes, M.; Loures, F. C. N.; Schmitt, A.; Frutos-Bernal, E.; Martin-Nogueras, A. M.; Pasqual Marques, A. | 2023 | Annals of the Rheumatic Diseases | 82 | Supplement 1 | 693 | https://dx.doi.org/10.1136/annrheumdis-2023-eular.736 | Not healthcare clinical practice |

| Title | Authors | Year | Journal | Volume | Issue | Pages | DOI | Exclusion Reason |
|---|---|---|---|---|---|---|---|---|
| Effectiveness of an Intervention Supporting Shared Decision Making for Destination Therapy Left Ventricular Assist Device: The DECIDE-LVAD Randomized Clinical Trial | Allen, Larry A.; McIlvennan, Colleen K.; Thompson, Jocelyn S.; Dunlay, Shannon M.; LaRue, Shane J.; Lewis, Eldrin F.; Patel, Chetan B.; Blue, Laura; Fairclough, Diane L.; Leister, Erin C.; Glasgow, Russell E.; Cleveland, Joseph C., Jr.; Phillips, Clifford; Baldridge, Vicie; Walsh, Mary Norine; Matlock, Daniel D. | 2018 | JAMA internal medicine | 178 | 4 | 520-529 | 10.1001/jamainternmed.2017.8713 | Not healthcare professionals |
| Effectiveness of an Intervention on Vaccine Hesitancy Among Pediatric Nurses and Pediatricians | Nct, | 2024 | https://clinicaltrials.gov/ct2/show/NCT06489236 | | | | | Not healthcare clinical practice |
| Effectiveness of an Online Educational Module in Improving Evidence-Based Practice Skills of Practicing Registered Nurses | Moore, Lora | 2017 | Worldviews on evidence-based nursing | 14 | 5 | 358-366 | 10.1111/wvn.12214 | No knowledge, skills, or clinical outcome |
| Effectiveness of an animation case-based learning approach to improve frontline nurses' knowledge and attitude on vital sign monitoring towards detecting clinical deterioration: a multi-site, pilot randomised controlled trial | Lin, Y. P.; Liaw, S. Y.; Chua, W. L.; Mok, W. Q. | 2015 | Annals of the academy of medicine singapore | 44 | 10 | S238- | | No data in abstract, and no PDF available |
| Effectiveness of an e-book in enhancing knowledge, coping behaviors and preventive strategies for sexual harassment prevention among new nurses: A randomized controlled study | Chang, T. S.; Tzeng, Y. L.; Teng, Y. K.; Chen, C. H. | 2025 | Nurse education in practice | 82 | | 104198 | https://dx.doi.org/10.1016/j.nepr.2024.104198 | Comparison group online |
| Effectiveness of an educational intervention on improving knowledge level of Chinese registered nurses on prevention of falls in hospitalized older people--a randomized controlled trial | Liu, Hui; Shen, Jun; Xiao, Lily Dongxia | 2012 | Nurse education today | 32 | 6 | 695-702 | 10.1016/j.nedt.2011.09.009 | Wrong intervention (no online learning) |
| Effectiveness of an educational program for mid-level Japanese public health nurses to improve program planning competencies: A preliminary randomized control trial | Yoshioka-Maeda, Kyoko; Shiomi, Misa; Katayama, Takafumi; Hosoya, Noriko; Kuroda, Mariko | 2019 | Public health nursing (Boston, Mass.) | 36 | 3 | 388-400 | 10.1111/phn.12580 | Not healthcare clinical practice |
| Effectiveness of an educational program on improving healthcare providers' knowledge of acute stroke: A randomized block design study | Rababah, Jehad A.; Al-Hammouri, Mohammed M.; AlNsour, Esra'a | 2021 | World journal of emergency medicine | 12 | 2 | 93-98 | 10.5847/wjem.j.1920-8642.2021.02.002 | Wrong intervention (no online learning) |

| Title | Authors | Year | Journal | Volume | Issue | Pages | DOI | Exclusion Reason |
|---|---|---|---|---|---|---|---|---|
| Effectiveness of an edutainment video teaching standard precautions - A randomized controlled evaluation study | Wolfensberger, A.; Anagnostopoulos, A.; Clack, L.; Meier, M. T.; Kuster, S. P.; Sax, H. | 2019 | Antimicrobial Resistance and Infection Control | 8 | 1 | | 10.1186/s13756-019-0531-5 | Intervention <10 mins |
| Effectiveness of an ergonomic training with exercise program for work-related musculoskeletal disorders among hemodialysis nurses: A pilot randomized control trial | Lee, M. J.; Wang, C. J.; Chang, J. H. | 2024 | Journal of Safety Research | 91 | | 481-491 | 10.1016/j.jsr.2024.09.007 | Not healthcare clinical practice |
| Effectiveness of an integrated pain management program on older persons and staff in nursing homes | Tse, M. | 2011 | European Journal of Pain Supplements | 5 | 1 | 245 | https://dx.doi.org/10.1016/S1754-3207%2811%2970848-3 | Wrong intervention (no online learning) |
| Effectiveness of continuing medical education and feedback to altering diabetes at a population level. A randomised controlled trial | Actrn | 2011 | http://www.who.int/trialsearch/Trial2.aspx?TrialID=ACTRN12611000553976 | | | | | No original data |
| Effectiveness of educational interventions to improve the quality use of medicines in Australian general practice for people with gout and older adults prescribed antidepressants | Actrn, | 2024 | https://trialsearch.who.int/Trial2.aspx?TrialID=ACTRN12624000989549 | | | | | No data in abstract, and no PDF available |
| Effectiveness of educational outreach visits compared with usual guideline dissemination to improve family physician prescribing-an 18-month open cluster-randomized trial | Pinto, Daniel; Heleno, Bruno; Rodrigues, David S.; Papoila, Ana Luísa; Santos, Isabel; Caetano, Pedro A. | 2018 | Implementation science : IS | 13 | 1 | 120 | 10.1186/s13012-018-0810-1 | Wrong intervention (no online learning) |
| Effectiveness of immersive teaching strategies on pressure injury: Impact on nurses' knowledge, attitudes and self-efficacy – A partially randomized participant preference (PRPP) controlled trial | Chao, W. Y.; Wu, Y. L.; Hsu, M. Y.; Chu, C. L. | 2025 | Nurse Education in Practice | 82 | | | 10.1016/j.nepr.2024.104237 | Comparison group online |
| Effectiveness of online medical education on clinical decision-making in hypoparathyroidism management | Larkin, A.; Chatterjee, P.; Badal, K.; Cusano, N. E. | 2016 | Endocrine Reviews | 37 | 2 Supplement 1 | | https://dx.doi.org/10.1210/endo-meetings.2016.BCHVD.8.SUN-325 | Not randomised |
| Effectiveness of online training in improving primary care doctors' competency in brief tobacco interventions: A cluster-randomized controlled trial of WHO modules in Delta State, Nigeria | Moeteke, Nnamdi Stephen; Oyibo, Patrick; Ochei, Oboratare; Ntaji, Maureen Iru; Awunor, Nyemike Simeon; Adeyemi, Mitchell Oritsewino; Enemuwe, Ibobo Mike; Agbatutu, Eseoghene; Adesoye, Oluwaseun Opeyemi | 2024 | PLoS ONE | 19 | 2 | | 10.1371/journal.pone.0292027 | No knowledge, skills, or clinical outcome |

| Title | Authors | Year | Journal | Volume | Issue | Pages | DOI | Exclusion Reason |
|---|---|---|---|---|---|---|---|---|
| Effectiveness of online versus in-person structured training program on arterial blood gas, electrolytes, and ventilatory management of critically ill patients | Jain, Gaurav; Gupta, Bhavna; Gupta, Priyanka; Panda, Sagarika; Sharma, Sameer; Rao, Shalinee | 2021 | Acute and critical care | 36 | 1 | 54-61 | 10.4266/acc.2020.00759 | Not randomised |
| Effectiveness of providing evidence-based practice education with workplace support for changing health professional's decision-making and outcomes of care: an evaluator blinded randomised controlled trial | Novak, I.; Campbell, L.; McIntyre, S. | 2011 | Developmental medicine and child neurology | 53 | | 60‐61 | 10.1111/j.1469-8749.2011.04112.x | Not randomised |
| Effectiveness of the Pediatric Nursing Excellence Model on Nurses' Knowledge and Practice in Pediatric Orthopedic Surgery Care: A Randomized Controlled Trial | Ramadan, O. M. E.; Hafiz, A. H.; Elsharkawy, N. B.; Katooa, N. E.; Abunar, A.; Abdelaziz, E. M.; Baraka, S. I. M.; Shaban, M.; Baraka, N. I. M. | 2024 | Children | 11 | 12 | | 10.3390/children11121457 | Wrong intervention (no online learning) |
| Effectiveness of the tailored EBP training program for Filipino physiotherapists: a randomised controlled trial | Dizon, Janine Margarita; Grimmer-Somers, Karen; Kumar, Saravana | 2011 | BMC medical education | 11 | | 14 | 10.1186/1472-6920-11-14 | Wrong intervention (no online learning) |
| Effectiveness of video assisted teaching programme on knowledge regarding practice of body mechanics among staff nurses in selected hosptals, Moradabad | Thomas, A.; Chithra, K.; Nageshwar, V. | 2017 | Indian journal of public health research and development | 8 | 2 | 39‐42 | 10.5958/0976-5506.2017.00079.1 | No data in abstract, and no PDF available |
| Effects evaluation of humanistic care abilities for new recruited nurses based on the blending instructional teaching model | Fan, Z.; Hanif, Mhbm; Hai, T. | 2019 | Basic & clinical pharmacology & toxicology | 125 | | 217‐ | 10.1111/bcpt.13266 | No data in abstract, and no PDF available |
| Effects of COPD guideline training on physicians' awareness, knowledge and readiness to Implement guidelines: a cluster randomized controlled trial | Alsubaiei, M. E.; Frith, P. A.; Cafarella, P. A.; Quinn, S.; Moamary, A.; McEvoy, D.; Effing, T. W. | 2017 | Respirology (Carlton, Vic.) | 22 | Suppl 2 | 142 [TP‐070] | | No original data |
| Effects of Cross-Training on Medical Teams' Teamwork and Collaboration: Use of Simulation | Hedges, Ashley R.; Johnson, Heather J.; Kobulinsky, Lawrence R.; Estock, Jamie L.; Eibling, David; Seybert, Amy L. | 2019 | Pharmacy (Basel, Switzerland) | 7 | 1 | | 10.3390/pharmacy7010013 | Comparison group online |
| Effects of Education on the Use of Personal Protective Equipment for Reduction of Contamination: A Randomized Trial | Yeon, Jeong Hwa; Shin, Yong Soon | 2020 | SAGE open nursing | 6 | | 2.37796E+15 | 10.1177/2377960820940621 | Wrong intervention (no online learning) |

| Title | Authors | Year | Journal | Volume | Issue | Pages | DOI | Exclusion Reason |
|---|---|---|---|---|---|---|---|---|
| Effects of a Multicomponent Restraint Reduction Program for Korean Nursing Home Staff | Kong, Eun-Hi; Song, Eunjin; Evans, Lois K. | 2017 | Journal of nursing scholarship : an official publication of Sigma Theta Tau International Honor Society of Nursing | 49 | 3 | 325-335 | 10.1111/jnu.12296 | Not healthcare professionals |
| Effects of a multi-component educational intervention on nurses' knowledge and adherence to standard precautions in intensive care units | Gomarverdi, Shiva; Khatiban, Mahnaz; Bikmoradi, Ali; Soltanian, Ali Reza | 2019 | Journal of infection prevention | 20 | 2 | 83-90 | 10.1177/1757177419830780 | Wrong intervention (no online learning) |
| Effects of a restraint minimization program on staff knowledge, attitudes, and practice: a cluster randomized trial | Pellfolk, Tony J. E.; Gustafson, Yngve; Bucht, Gösta; Karlsson, Stig | 2010 | Journal of the American Geriatrics Society | 58 | 1 | 62-69 | 10.1111/j.1532-5415.2009.02629.x | Wrong intervention (no online learning) |
| Effects of a simulation-based education programme on delirium care for critical care nurses: A randomized controlled trial | Ho, Mu-Hsing; Yu, Lee-Fen; Lin, Pu-Hung; Chang, Hui-Chen Rita; Traynor, Victoria; Huang, Wen-Cheng; Montayre, Jed; Chen, Kee-Hsin | 2021 | Journal of advanced nursing | 77 | 8 | 3483-3493 | 10.1111/jan.14938 | Comparison group online |
| Effects of a web-based education program for nurses using medical malpractice cases: A randomized controlled trial | Lim, Haena; Yi, Yeojin | 2021 | Nurse education today | 104 | | 104997 | 10.1016/j.nedt.2021.104997 | Not healthcare clinical practice |
| Effects of an educational intervention for general practitioners in adolescent health care principles: a randomized controlled study | Sanci, L. A.; Coffey, C. M.; Veit, F. C.; Carr-Gregg, M.; Patton, G. C.; Bowes, G.; Day, N. | 2000 | The Western journal of medicine | 172 | 3 | 157-163 | 10.1136/ewjm.172.3.157 | Wrong intervention (no online learning) |
| Effects of clinical-practice guideline and practice-based education on detection and outcome of depression in primary care: Hampshire depression project randomised controlled trial | Tammann, T. | 2000 | Praxis | 89 | 42 | 1721 | | Wrong intervention (no online learning) |
| Effects of distance learning on clinical management of LUTS in primary care: a randomised trial | Wolters, René; Wensing, Michel; Klomp, Maarten; Lagro-Jansen, Toine; Weel, Chris van; Grol, Richard | 2005 | Patient education and counseling | 59 | 2 | 212-218 | 10.1016/j.pec.2004.11.009 | Wrong intervention (no online learning) |
| Effects of gamification in advanced life support training for clinical nurses: a cluster randomized controlled trial | Kim, K.; Choi, D.; Shim, H.; Lee, C. A. | 2024 | Nurse education today | 140 | | 106263 | 10.1016/j.nedt.2024.106263 | Wrong intervention (no online learning) |
| Effects of mutual structured feedback on nurses performance | Shahvaran, R.; Rezaee, R.; Bazrafkan, L. | 2018 | Journal of Advanced Pharmacy Education and Research | 8 | | 44-48 | | Wrong intervention (no online learning) |

| Title | Authors | Year | Journal | Volume | Issue | Pages | DOI | Exclusion Reason |
|---|---|---|---|---|---|---|---|---|
| Effects of perceived educational support on usage of an Internet Nursing Reference Center | Marshall, Brenda L.; Sabbagh, Lia | 2015 | The Journal of Continuing Education in Nursing | 46 | 4 | 161-168 | 10.3928/00220124-20150320-01 | Not healthcare clinical practice |
| Efficacy of a training intervention on the quality of practitioners' decision support for patients deciding about place of care at the end of life: A randomized control trial: Study protocol | Murray, Mary Ann; O'Connor, Annette; Stacey, Dawn; Wilson, Keith G. | 2008 | BMC palliative care | 7 | | 4 | 10.1186/1472-684X-7-4 | No original data |
| Efficacy of a web-based intervention to improve and sustain knowledge and screening for amblyopia in primary care settings | Marsh-Tootle, Wendy L.; McGwin, Gerald; Kohler, Connie L.; Kristofco, Robert E.; Datla, Raju V.; Wall, Terry C. | 2011 | Investigative ophthalmology & visual science | 52 | 10 | 7160-7167 | 10.1167/iovs.10-6566 | Comparison group online |
| Efficacy of an educational Web site for educating physicians about bioterrorism | Chung, Sarita; Mandl, Kenneth D.; Shannon, Michael; Fleisher, Gary R. | 2004 | Academic emergency medicine : official journal of the Society for Academic Emergency Medicine | 11 | 2 | 143-148 | | Wrong intervention (no online learning) |
| Efficacy of didactic vs experiential learning | Saraswat, A.; Dominguez, E. P.; Watson Ii, W. D.; Moreland, J.; Elliott, J. O.; Bach, J. A. | 2015 | Journal of the American College of Surgeons | 221 | 4 SUPPL. 1 | S52 | | Wrong intervention (no online learning) |
| Efficacy of educational video game versus traditional educational apps at improving physician decision making in trauma triage: randomized controlled trial | Mohan, D.; Farris, C.; Fischhoff, B.; Rosengart, M. R.; Angus, D. C.; Yealy, D. M.; Wallace, D. J.; Barnato, A. E. | 2017 | BMJ (Clinical research ed.) | 359 | | j5416 | 10.1136/bmj.j5416 | Comparison group online |
| Efficacy of teaching clinical clerks and residents how to fill out the form 1 of the mental health act using an e-learning module | Garside, S.; Levinson, A.; Kuziora, S.; Bay, M.; Norman, G. | 2009 | | 2009-January | | 130-135 | | No comparison or control |
| Efforts to Improve Colonoscopy Efficiency by Reduction of the Non-Neoplastic Polypectomy Rate in a Colorectal Cancer Screening Program | Berger, D.; Jakate, S.; Esteban, M.; Godbee, M.; Losurdo, J.; Keshavarzian, A.; Melson, J. E. | 2020 | Gastrointestinal endoscopy | 91 | 6 | AB577-AB578 | 10.1016/j.gie.2020.03.3345 | Wrong intervention (no online learning) |

| Title | Authors | Year | Journal | Volume | Issue | Pages | DOI | Exclusion Reason |
|---|---|---|---|---|---|---|---|---|
| Elearning to improve parenteral nutrition skills in pediatric: pilot study in 2 university's hospital | Petit, L. M.; Pape, P. L.; Bajwa, N.; Kanavaki, I.; Garzoni, L.; McLin, V.; Fonzo-Christe, C.; Bonabry, P.; Belli, D. | 2017 | Journal of pediatric gastroenterology and nutrition | Conference: 50th Annual Meeting of the European Society for Paediatric Gastroenterology, Hepatology and Nutrition, ESPGHAN 2017. Czech Republic. 64 | pp 1001 | | 10.1097/01.mpg.0000516381.25680.b4 | Duplicate |
| Electronic physician notifications to improve guideline-based anticoagulation in atrial fibrillation: a randomized controlled trial | Ashburner, Jeffrey M.; Atlas, Steven J.; Khurshid, Shaan; Weng, Lu-Chen; Hulme, Olivia L.; Chang, Yuchiao; Singer, Daniel E.; Ellinor, Patrick T.; Lubitz, Steven A. | 2018 | Journal of general internal medicine | 33 | 12 | 2070-2077 | 10.1007/s11606-018-4612-6 | Wrong intervention (no online learning) |
| Embedding Learning in a Learning Health Care System to Improve Clinical Practice | McEvoy, Matthew D.; Dear, Mary Lynn; Buie, Reagan; Fowler, Leslie C.; Miller, Bonnie; Fleming, Geoffrey M.; Moore, Don; Rice, Todd W.; Bernard, Gordon R.; Lindsell, Christopher J. | 2021 | Academic medicine : journal of the Association of American Medical Colleges | 96 | 9 | 1311-1314 | 10.1097/ACM.0000000000003969 | Comparison group online |
| Empathy training for resident physicians: a randomized controlled trial of a neuroscience-informed curriculum | Riess, Helen; Kelley, John M.; Bailey, Robert W.; Dunn, Emily J.; Phillips, Margot | 2012 | Journal of general internal medicine | 27 | 10 | 1280-1286 | 10.1007/s11606-012-2063-z | Wrong intervention (no online learning) |

| Title | Authors | Year | Journal | Volume | Issue | Pages | DOI | Exclusion Reason |
|---|---|---|---|---|---|---|---|---|
| Empowering hospital-associated infection prevention and control: a quasi-experimental study on the effect of scenario-based simulation training | Lee, S. H.; Yang, I. S. | 2024 | Nurse education in practice | 76 | | 103936 | 10.1016/j.nepr.2024.103936 | Wrong intervention (no online learning) |
| Empowerment of nurses for integrating clients' religion/spirituality into clinical practice: outcomes of an online training program | Amiri, Hasan; Farokhzadian, Jamileh; Tirgari, Batool | 2021 | BMC nursing | 20 | 1 | 210 | 10.1186/s12912-021-00723-y | Not healthcare clinical practice |
| Empowerment of nurses' knowledge and skills on violence against women with mental illness: A randomized controlled trail | Vijayalakshmi, Poreddi; Gandhi, Sailaxmi; Nikhil Reddy, S. S.; Palaniappan, Marimuthu; BadaMath, Suresh | 2021 | Archives of psychiatric nursing | 35 | 3 | 261-266 | 10.1016/j.apnu.2021.03.005 | Wrong intervention (no online learning) |
| Enhancing Islamic based nurses' competencies in disaster response in Banda Aceh hospital, Indonesia | Actrn | 2020 | https://trialsearch.who.int/Trial2.aspx?TrialID=ACTRN12620001289909 | | | | | Wrong intervention (no online learning) |
| Enhancing motivation with the "virtual" supervisory role: a randomized trial | Wingo, Majken T.; Thomas, Kris G.; Thompson, Warren G.; Cook, David A. | 2015 | BMC medical education | 15 | | 76 | 10.1186/s12909-015-0348-8 | Comparison group online |
| Establishing the Clinical Utility of First StepDx PLUS and NextStepDx PLUS Study | Nct | 2015 | https://clinicaltrials.gov/show/NCT02414438 | | | | | No data in abstract, and no PDF available |
| Evaluating a Web-Based Ventilator Management Educational Program for Clinicians (The Lung Injury Knowledge Network [LINK] Study) | Nct | 2007 | https://clinicaltrials.gov/show/NCT00542737 | | | | | Not randomised |
| Evaluating a hybrid web-based basic genetics course for health professionals | Wallen, Gwenyth R.; Cusack, Georgie; Parada, Suzan; Miller-Davis, Claiborne; Cartledge, Tannia; Yates, Jan | 2011 | Nurse education today | 31 | 6 | 638-642 | 10.1016/j.nedt.2010.11.001 | Not randomised |
| Evaluating an Online Intervention to Improve Provider Management of Prenatal Depression: Randomized Control Trial | Leiferman, Jenn A.; Lee-Winn, Angela E.; Lacy, Rachael; Paulson, James F. | 2022 | Women's health issues : official publication of the Jacobs Institute of Women's Health | | | | 10.1016/j.whi.2022.08.009 | Not enough data for analysis |
| Evaluating an Online Intervention to Improve Provider Management of Prenatal Depression: a Randomized Controlled Trial | Leiferman, J. A.; Lee-Winn, A. E.; Lacy, R.; Paulson, J. F. | 2023 | Women's health issues | 33 | 2 | 175‐181 | 10.1016/j.whi.2022.08.009 | Not healthcare professionals |
| Evaluating an educational intervention of central venous catheter maintenance among oncology nurses | Bubb, T. N. | 2015 | American journal of infection control | 43 | 6 | S37‐ | | Wrong intervention (no online learning) |

| Title | Authors | Year | Journal | Volume | Issue | Pages | DOI | Exclusion Reason |
|---|---|---|---|---|---|---|---|---|
| Evaluating impact of a multi-dimensional education programme on perceived performance of primary care professionals in diabetes care | Parekh, Sanjoti; Bush, Robert; Cook, Susan; Grant, Phillipa | 2015 | Primary health care research & development | 16 | 6 | 589-596 | 10.1017/S1463423615000195 | Not randomised |
| Evaluating online continuing medical education seminars: evidence for improving clinical practices | Weston, Christine M.; Sciamanna, Christopher N.; Nash, David B. | 2008 | American journal of medical quality : the official journal of the American College of Medical Quality | 23 | 6 | 475-483 | 10.1177/1062860608325266 | Comparison group online |
| Evaluating the Effectiveness of I-NEED Program: Improving Nurses' Detection and Management of Elder Abuse and Neglect-A 6-Month Prospective Study | Mohd Mydin, Fadzilah Hanum; Wan Yuen, Choo; Othman, Sajaratulnisah; Mohd Hairi, Noran Naqiah; Mohd Hairi, Farizah; Ali, Zainudin; Abdul Aziz, Suriyati | 2022 | Journal of interpersonal violence | 37 | 1-Feb | NP719-NP741 | 10.1177/0886260520918580 | Wrong intervention (no online learning) |
| Evaluating the effectiveness of the cardiovascular assessment screening program with nurse practitioners and patients: results of a cluster randomised controlled trial | Bruneau, J.; Moralejo, D.; Parsons, K. | 2024 | BMC primary care | 25 | 1 | 185 | 10.1186/s12875-024-02432-2 | Wrong intervention (no online learning) |
| Evaluating the impact of an evidence-based medicine educational intervention on primary care doctors' attitudes, knowledge and clinical behaviour: a controlled trial and before and after study | Shuval, Kerem; Berkovits, Eldar; Netzer, Doron; Hekselman, Igal; Linn, Shai; Brezis, Mayer; Reis, Shmuel | 2007 | Journal of evaluation in clinical practice | 13 | 4 | 581-598 | 10.1111/j.1365-2753.2007.00859.x | Wrong intervention (no online learning) |
| Evaluating the implementation of the active life improving health behavior change program "BCP-VAMOS" in primary health care: Protocol of a pragmatic randomized controlled trial using the RE-AIM and CFIR frameworks | Konrad, Lisandra Maria; Ribeiro, Cezar Grontowski; Maciel, Elaine Cristina; Tomicki, Camila; Brito, Fabiana Almeida; Almeida, Fabio Araujo; Benedetti, Tânia Rosane Bertoldo | 2022 | Frontiers in public health | 10 | | 726021 | 10.3389/fpubh.2022.726021 | No original data |
| Evaluating the role of omega-3 fatty acids in dyslipidemia: Success of online CME | Spyropoulos, J.; Marko, J. | 2017 | Journal of Clinical Lipidology | 11 | 3 | 830-831 | | Not randomised |
| Evaluating video-reflexive methods to improve infection prevention and use of personal protective equipment in Australian hospitals | Wyer, M.; Barratt, R.; Hor, S.; Gilbert, L. | 2019 | Infection, disease and health | 24 | | S4- | 10.1016/j.idh.2019.09.013 | Wrong intervention (no online learning) |
| Evaluation of Adaptive Feedback in a Smartphone-Based Serious Game on Health Care Providers' Knowledge Gain in Neonatal Emergency Care: Protocol for a Randomized Controlled Trial | Tuti, Timothy; Winters, Niall; Muinga, Naomi; Wanyama, Conrad; English, Mike; Paton, Chris | 2019 | JMIR research protocols | 8 | 7 | e13034 | 10.2196/13034 | Comparison group online |

| Title | Authors | Year | Journal | Volume | Issue | Pages | DOI | Exclusion Reason |
|---|---|---|---|---|---|---|---|---|
| Evaluation of An Online Intervention In Improving General Practitioners' Practice In Prostate Cancer Screening | Nct | 2018 | https://clinicaltrials.gov/show/NCT03633214 | | | | | No data in abstract, and no PDF available |
| Evaluation of Case Management to Improve the Outpatient Care of Alcohol-related Disorders | Nct | 2006 | https://clinicaltrials.gov/show/NCT00314067 | | | | | Comparison group online |
| Evaluation of IMproving Palliative care Education and Training Using Simulation in Dementia (IMPETUS-D) a staff simulation training intervention to improve palliative care of people with advanced dementia living in nursing homes: a cluster randomised cont | Tropea, Joanne; Nestel, Debra; Johnson, Christina; Hayes, Barbara J.; Hutchinson, Anastasia F.; Brand, Caroline; Le, Brian H.; Blackberry, Irene; Caplan, Gideon A.; Bicknell, Ross; Hepworth, Graham; Lim, Wen K. | 2022 | BMC geriatrics | 22 | 1 | 127 | 10.1186/s12877-022-02809-x | Not healthcare professionals |
| Evaluation of Online Training Tools in Pediatric Resuscitation | Nct | 2018 | https://clinicaltrials.gov/show/NCT03640520 | | | | | No data in abstract, and no PDF available |
| Evaluation of a Web-Based ADHD Awareness Training in Primary Care: Pilot Randomized Controlled Trial With Nested Interviews | French, Blandine; Hall, Charlotte; Perez Vallejos, Elvira; Sayal, Kapil; Daley, David | 2020 | JMIR medical education | 6 | 2 | e19871 | 10.2196/19871 | Comparison group online |
| Evaluation of a Web-based learning system for skills in removing personal protective equipment for highly infectious diseases-A randomized controlled trial | Sato, S.; Iijima, S. | 2023 | Infection control and hospital epidemiology | 44 | 7 | 1131-1136 | 10.1017/ice.2022.219 | Not healthcare clinical practice |
| Evaluation of a computer based package on electrocardiography | Devitt, P.; Worthley, S.; Palmer, E.; Cehic, D. | 1998 | Australian and New Zealand journal of medicine | 28 | 4 | 432-435 | 10.1111/j.1445-5994.1998.tb02076.x | Wrong intervention (no online learning) |
| Evaluation of a national Bright Futures oral health curriculum for pediatric residents | Bernstein, Henry H.; Dhepyasuwan, Niramol; Connors, Kara; Volkan, Kevin; Serwint, Janet R. | 2013 | Academic pediatrics | 13 | 2 | 133-139 | 10.1016/j.acap.2012.10.010 | Comparison group online |
| Evaluation of a nurse-led dementia education and knowledge translation programme in primary care: A cluster randomized controlled trial | Wang, Yao; Xiao, Lily Dongxia; Ullah, Shahid; He, Guo-Ping; De Bellis, Anita | 2017 | Nurse education today | 49 | | 1-Jul | 10.1016/j.nedt.2016.10.016 | Wrong intervention (no online learning) |
| Evaluation of a one-hour asynchronous video training for eating disorder screening and referral in U.S. Pediatric Primary Care: A pilot study | Raffoul, Amanda; Vitagliano, Julia A.; Sarda, Vishnudas; Chan, Charmaine; Chwa, Cindy; Ferreira, Katelyn B.; Gooding, Holly C.; Forman, Sara F.; Austin, S. Bryn | 2022 | The International journal of eating disorders | 55 | 9 | 1245-1251 | 10.1002/eat.23766 | Comparison group online |

| Title | Authors | Year | Journal | Volume | Issue | Pages | DOI | Exclusion Reason |
|---|---|---|---|---|---|---|---|---|
| Evaluation of a women's safe shelter experience to teach internal medicine residents about intimate partner violence. A randomized controlled trial | Brienza, Rebecca S.; Whitman, Laura; Ladouceur, Lynnea; Green, Michael L. | 2005 | Journal of general internal medicine | 20 | 6 | 536-540 | 10.1111/j.1525-1497.2005.0100.x | Wrong intervention (no online learning) |
| Evaluation of an Internet-based decision-support system for applying the ATS/CDC guidelines for tuberculosis preventive therapy | Dayton, C. S.; Ferguson, J. S.; Hornick, D. B.; Peterson, M. W. | 2000 | Medical decision making : an international journal of the Society for Medical Decision Making | 20 | 1 | 1-Jun | 10.1177/0272989X0002000101 | Wrong intervention (no online learning) |
| Evaluation of an educational online intervention aiming to improve primary care physicians' awareness of addressing social determinants of health in clinical care in Saudi Arabia | Isrctn | 2021 | https://trialsearch.who.int/Trial2.aspx?TrialID=ISRCTN14600984 | | | | | No original data |
| Evaluation of an online Diabetes Needs Assessment Tool (DNAT) for health professionals: a randomised controlled trial | Schroter, Sara; Jenkins, Dean; Playle, Rebecca; Walsh, Kieran; Probert, Courtenay; Kellner, Thomas; Arnhofer, Gerhard; Owens, David | 2009 | Trials | 10 | | 63 | 10.1186/1745-6215-10-63 | Comparison group online |
| Evaluation of an online interactive Diabetes Needs Assessment Tool (DNAT) versus online self-directed learning: a randomised controlled trial | Schroter, Sara; Jenkins, Richard D.; Playle, Rebecca A.; Walsh, Kieran M.; Probert, Courtenay; Kellner, Thomas; Arnhofer, Gerhard; Owens, David R. | 2011 | BMC medical education | 11 | | 35 | 10.1186/1472-6920-11-35 | Comparison group online |
| Evaluation of benefit to patients of training mental health professionals in suicide guidelines: cluster randomised trial | de Beurs, Derek P.; de Groot, Marieke H.; de Keijser, Jos; van Duijn, Erik; de Winter, Remco F. P.; Kerkhof, Ad J. F. M. | 2016 | The British journal of psychiatry : the journal of mental science | 208 | 5 | 477-483 | 10.1192/bjp.bp.114.156208 | Not healthcare clinical practice |
| Evaluation of online training on the prevention of venous thromboembolism | Wolpin, Seth; Lee, Jung-Ah; Glenny, Robb W.; Wittkowsky, Ann K.; Wolf, Fredric M.; Zierler, Brenda K. | 2011 | Vascular and endovascular surgery | 45 | 2 | 146-156 | 10.1177/1538574410391281 | Not randomised |
| Evaluation of primary care physicians' competence in selective skin tumour triage after short versus long dermoscopy training: a randomized non-inferiority trial | Harkemanne, E.; Legrand, C.; Sawadogo, K.; van Maanen, A.; Vossaert, K.; Argenziano, G.; Braun, R.; Thomas, L.; Baeck, M.; Tromme, I. | 2023 | Journal of the European Academy of Dermatology and Venereology : JEADV | | | | 10.1111/jdv.19087 | Comparison group online |
| Evaluation of the Effectiveness of Psychoeducational Training Given to Nurses Working in a Psychiatric Clinic | Nct, | 2024 | https://clinicaltrials.gov/ct2/show/NCT06388798 | | | | | No data in abstract, and no PDF available |

| Title | Authors | Year | Journal | Volume | Issue | Pages | DOI | Exclusion Reason |
|---|---|---|---|---|---|---|---|---|
| Evaluation of the e-learning "Training for Occupational health professionals To Involve Significant others" (TOTIS) | Nl | 2020 | https://trialsearch.who.int/Trial2.aspx?TrialID=NL8744 | | | | | No original data |
| Evaluation of the effectiveness of an educational intervention in community pharmacists to improve pharmaceutical care in upper respiratory infections, including flu, colds, sore throat and sinusitis | Isrctn | 2018 | https://trialsearch.who.int/Trial2.aspx?TrialID=ISRCTN89439892 | | | | | Not randomised |
| Evaluation of the effects of the Global Initiative for Chronic Obstructive Lung Disease (GOLD)-guidelines training on knowledge and implementation of these guidelines by physicians who are providing COPD-care in the eastern province of Saudi Arabia: clust | Actrn | 2014 | https://trialsearch.who.int/Trial2.aspx?TrialID=ACTRN12614000741684 | | | | | No original data |
| Evaluation of the self management of osteoarthritis and low back pain through activity and skills physiotherapist e-learning training programme compared to face to face training | Hurley, D. A.; Keogh, A.; Hall, A.; Richmond, H.; Magdalinski, T.; Matthews, J. | 2018 | Osteoarthritis and cartilage | Conference: 2018 Osteoarthritis Research Society International, OARSI World Congress. United Kingdom. 26 | Supplement 1 | S269-S270 | | Not randomised |
| Evidence of improved knowledge and skills among hematologists/oncologists participating in online CME-certified activities | Willis, L.; Van Laar, E. S.; Abair, T.; Whitney, M.; Costello, C.; Krishnan, A. Y.; Lonial, S.; Kurtin, S.; Mikhael, J.; Landgren, O. | 2021 | Blood | 138 | SUPPL 1 | 4958 | https://dx.doi.org/10.1182/blood-2021-145375 | Not randomised |
| Evidence-based interventions in dementia: A pragmatic cluster-randomised trial of an educational intervention to promote earlier recognition and response to dementia in primary care (EVIDEM-ED) | Iliffe, Steve; Wilcock, Jane; Griffin, Mark; Jain, Priya; Thuné-Boyle, Ingela; Koch, Tamar; Lefford, Frances | 2010 | Trials | 11 | | 13 | 10.1186/1745-6215-11-13 | No original data |

| Title | Authors | Year | Journal | Volume | Issue | Pages | DOI | Exclusion Reason |
|---|---|---|---|---|---|---|---|---|
| Evidence-based practice for the athletic training profession | Welch, Cailee Elizabeth | 2013 | | 73 | | | | Duplicate |
| Examining the effect of teaching method and learning style on work performance for practicing home care clinicians | Caulfield, Johnette Lynn | 2002 | | 62 | | 3688-3688 | | No knowledge, skills, or clinical outcome |
| Examining the usefulness of extended digitally driven training programs for grass root level health workers | Ctri | 2019 | https://trialsearch.who.int/Trial2.aspx?TrialID=CTRI/2019/12/022245 | | | | | No data in abstract, and no PDF available |
| Experience of blended learning paradigm in teaching benign paroxysmal positional vertigo (BPPV). A randomized controlled trial | Bashir, K.; Mohammed Elmoheen, A.; Eltawagny, M.; Anjum, S.; Ibrahim, A.; Thomas, S. | 2019 | EMA - emergency medicine australasia | 31 | | 44- | 10.1111/1742-6723.13240 | Wrong intervention (no online learning) |
| Exposing clinicians to exposure: a randomized controlled dissemination trial of exposure therapy for anxiety disorders | Harned, Melanie S.; Dimeff, Linda A.; Woodcock, Eric A.; Kelly, Tim; Zavertnik, Jake; Contreras, Ignacio; Danner, Sankirtana M. | 2014 | Behavior therapy | 45 | 6 | 731-744 | 10.1016/j.beth.2014.04.005 | Comparison group online |
| Facebook as a Novel Tool for Continuous Professional Education on Dementia: Pilot Randomized Controlled Trial | Chan, Windy Sy; Leung, Angela Ym | 2020 | Journal of medical Internet research | 22 | 6 | e16772 | 10.2196/16772 | Comparison group online |
| Facilitating Adaptive Expertise in Learning Computed Tomography: a Multicenter Randomized Controlled Trial | Aliaga, L.; Bavolek, R. A.; Cooper, B. L.; Mariorenzi, A.; Ahn, J.; Kraut, A.; Duong, D.; Gisondi, M. A. | 2023 | Academic Emergency Medicine | 30 | | 10 | 10.1111/acem.14718 | Comparison group online |
| Facilitating Alcohol Screening and Treatment (FAST) | Nct | 2020 | https://clinicaltrials.gov/show/NCT04303676 | | | | | No original data |
| Facilitating the adoption of microalbuminuria (mau) screening among type II diabetic patients in primary care: preliminary results of a randomized educational intervention trial | Naimark, D. M. J.; Bott, M. T.; Tobe, S. W. | 2002 | Journal of the American Society of Nephrology : JASN | 13 | September, Program & Abstracts | 638a | | No data in abstract, and no PDF available |
| Feasibility and acceptability of e-learning to upskill diabetes educators in supporting people experiencing diabetes distress: a pilot randomised controlled trial | Halliday, Jennifer A.; Russell-Green, Sienna; Hagger, Virginia; O, Eric; Morris, Ann; Sturt, Jackie; Speight, Jane; Hendrieckx, Christel | 2022 | BMC medical education | 22 | 1 | 768 | 10.1186/s12909-022-03821-w | Not enough data for analysis |

| Title | Authors | Year | Journal | Volume | Issue | Pages | DOI | Exclusion Reason |
|---|---|---|---|---|---|---|---|---|
| Feasibility of implementing a web-based education program in geriatric pain and depression for home health care nurses | Brody, Abraham A.; Groce-Wofford, Trinity M. | 2013 | Home Health Care Management & Practice | 25 | 6 | 274-278 | 10.1177/1084822313494785 | Not enough data for analysis |
| Feedback in primary care can improve the prescribing of hormone replacement therapy in women with a history of hysterectomy | McCartney, P.; Macdowall, W.; Thorogood, M. | 2001 | British journal of clinical governance | 6 | 1 | 17‐21 | | Wrong intervention (no online learning) |
| Forensic nursing science knowledge and competency: the use of simulation | Drake, Stacy A.; Langford, Rae; Young, Anne; Ayers, Constance | 2015 | Critical care nursing quarterly | 38 | 1 | 81-88 | 10.1097/CNQ.0000000000000045 | Not healthcare professionals |
| Future Health Today: a cluster randomised controlled trial of quality improvement activities in general practice | Actrn | 2020 | https://trialsearch.who.int/Trial2.aspx?TrialID=ACTRN12620000993998 | | | | | No original data |
| GIANT: general Practitioner Implementation in Asia of Normoglycaemic Targets | Nct | 2007 | https://clinicaltrials.gov/show/NCT00499824 | | | | | Duplicate |
| Game-based versus traditional case-based learning: comparing effectiveness in stroke continuing medical education | Telner, Deanna; Bujas-Bobanovic, Maja; Chan, David; Chester, Bob; Marlow, Bernard; Meuser, James; Rothman, Arthur; Harvey, Bart | 2010 | Canadian family physician Medecin de famille canadien | 56 | 9 | e345-e351 | | Wrong intervention (no online learning) |
| General Practice Optimising Structured MOnitoring To Improve Clinical outcomes in Type 2 Diabetes: an individually randomised trial of the effect of retrospective continuous glucose monitoring (rCGM) for people with type 2 diabetes in general practice on | Actrn | 2016 | https://trialsearch.who.int/Trial2.aspx?TrialID=ACTRN12616001372471 | | | | | No original data |
| GenetiKiT: evaluation of an Educational Intervention on the Delivery of Genetics Services by Family Physicians | Nct | 2006 | https://clinicaltrials.gov/show/NCT00295529 | | | | | No original data |
| GenetiKit: a randomized controlled trial to enhance delivery of genetics services by family physicians | Carroll, June C.; Wilson, Brenda J.; Allanson, Judith; Grimshaw, Jeremy; Blaine, Sean M.; Meschino, Wendy S.; Permaul, Joanne A.; Graham, Ian D. | 2011 | Family practice | 28 | 6 | 615-623 | 10.1093/fampra/cmr040 | Wrong intervention (no online learning) |
| Glucose Control Using 1,5-AG Testing | Nct | 2018 | https://clinicaltrials.gov/show/NCT03765164 | | | | | Comparison group online |
| Guideline implementation in the Canadian chiropractic setting: a pilot cluster randomized controlled trial and parallel study | Dhopte, Prakash; French, Simon D.; Quon, Jeffrey A.; Owens, Heather; Bussières, André | 2019 | Chiropractic & manual therapies | 27 | | 31 | 10.1186/s12998-019-0253-z | No knowledge, skills, or clinical outcome |

| Title | Authors | Year | Journal | Volume | Issue | Pages | DOI | Exclusion Reason |
|---|---|---|---|---|---|---|---|---|
| Guideline recommended therapies in HFrEF, online medical education improves cardiologists' knowledge and competence | Lionel Thevathasan, L.; Miller, R.; Ashley, N.; Schoonheim, P. | 2019 | European Journal of Heart Failure | 21 | Supplement 1 | 143 | https://dx.doi.org/10.1002/ejhf.1488 | Not randomised |
| Headsmart Jordan: The Impact of an Hour of Zoom Lecture about Pediatric Brain Tumors Symptomatology on the Knowledge of the Health Care Providers | Amayiri, N.; Walker, D.; Ammar, K.; Bouffet, E. | 2021 | Pediatric Blood and Cancer | 68 | SUPPL 5 | | https://dx.doi.org/10.1002/pbc.29349 | Not randomised |
| Health improvement and prevention study (HIPS) - evaluation of an intervention to prevent vascular disease in general practice | Fanaian, Mahnaz; Laws, Rachel A.; Passey, Megan; McKenzie, Suzanne; Wan, Qing; Davies, Gawaine Powell; Lyle, David; Harris, Mark F. | 2010 | BMC family practice | 11 | | 57 | 10.1186/1471-2296-11-57 | No original data |
| Health literacy, advocacy and cardiovascular disease risk education by resident physicians for patients using a medical app: development, implementation and evaluation | Mehta, R. M. | 2014 | Canadian journal of cardiology | 30 | 10 | S214- | | No original data |
| Health workers learned brief intervention online through nextge-nu.org and deliver them effectively | Musau, A.; Mokaya, A. G.; Tele, A.; Ndetei, D. M.; Frank, E.; Clair, V.; Mutiso, V. | 2016 | Journal of addiction medicine | 10 | 3 | E17- | 10.1097/ADM.0000000000000224 | No knowledge, skills, or clinical outcome |
| Healthy Heart Africa-Kenya: A 12-Month Prospective Evaluation of Program Impact on Health Care Providers' Knowledge and Treatment of Hypertension | Ogola, Elijah N.; Okello, Francis O.; Herr, Jane L.; Macgregor-Skinner, Elizabeth; Mulvaney, Ashling; Yonga, Gerald | 2019 | Global heart | 14 | 1 | 61-70 | 10.1016/j.gheart.2019.02.002 | Wrong intervention (no online learning) |
| Helping Hands: Using Augmented Reality to Provide Remote Guidance to Health Professionals | Mather, Carey; Barnett, Tony; Broucek, Vlasti; Saunders, Annette; Grattidge, Darren; Huang, Weidong | 2017 | Studies in health technology and informatics | 241 | | 57-62 | | Not healthcare professionals |
| How can we improve bone health in the long-term care setting? Lessons from the vidos study | Kennedy, C.; Papaioannou, A.; Ioannidis, G.; Giangregorio, L.; Marr, S.; Morin, S.; Thabane, L.; Josse, R.; Crilly, R.; Pickard, L.; et al. | 2011 | Journal of bone and mineral research | 26 | | | | Duplicate |

| Title | Authors | Year | Journal | Volume | Issue | Pages | DOI | Exclusion Reason |
|---|---|---|---|---|---|---|---|---|
| Hypertension Improvement Project (HIP): study protocol and implementation challenges | Dolor, Rowena J.; Yancy, William S., Jr.; Owen, William F.; Matchar, David B.; Samsa, Gregory P.; Pollak, Kathryn I.; Lin, Pao-Hwa; Ard, Jamy D.; Prempeh, Maxwell; McGuire, Heather L.; Batch, Bryan C.; Fan, William; Svetkey, Laura P. | 2009 | Trials | 10 | | 13 | 10.1186/1745-6215-10-13 | No original data |
| I-NEED: improving Nurses? dEtection and managEment of elDer abuse and neglect | Isrctn | 2014 | https://trialsearch.who.int/Trial2.aspx?TrialID=ISRCTN47326902 | | | | | No original data |
| ISE to Support Constipation Management | Nct | 2016 | https://clinicaltrials.gov/show/NCT02977858 | | | | | No original data |
| Immersive virtual reality demonstrates improved and efficient surgical skill acquisition in senior orthopedic residents: a prospective blinded randomized controlled trial | Goel, D.; Lohre, R.; Bois, A.; Athwal, G. | 2019 | CMAJ. Canadian medical association journal | 62 | 6 | S216- | 10.1503/cjs.020119 | Wrong intervention (no online learning) |
| Immersive virtual reality: a novel and cool way to learn congenital heart defects | Gharpure, V.; Cormier, S.; Ganbote, T.; Shetty, I. | 2019 | Critical care medicine | 47 | 1 | | | Wrong intervention (no online learning) |
| Impact of a 16-community trial to promote judicious antibiotic use in Massachusetts | Finkelstein, Jonathan A.; Huang, Susan S.; Kleinman, Ken; Rifas-Shiman, Sheryl L.; Stille, Christopher J.; Daniel, James; Schiff, Nancy; Steingard, Ron; Soumerai, Stephen B.; Ross-Degnan, Dennis; Goldmann, Donald; Platt, Richard | 2008 | Pediatrics | 121 | 1 | e15-e23 | 10.1542/peds.2007-0819 | Not healthcare professionals |
| Impact of a Multi-modal Multi-professional Intervention on the Nurse on Antibiotics Prescribing in EHPAD Retirement Home | Nct | 2019 | https://clinicaltrials.gov/show/NCT03994523 | | | | | No original data |
| Impact of a Multifaceted Intervention Among Primary Care Physicians on Sickness Certification | Romani, Maya; Assaf, Georges; Mahfoud, Mirna; Hoteit, Reem; Saab, Basem Roberto | 2022 | The Journal of continuing education in the health professions | 42 | 3 | e121-e124 | 10.1097/CEH.0000000000000428 | Not randomised |
| Impact of a Serious Game on the Intention to Change Infection Prevention and Control Practices in Nursing Homes During the COVID-19 Pandemic: Protocol for a Web-Based Randomized Controlled Trial | Suppan, Laurent; Abbas, Mohamed; Catho, Gaud; Stuby, Loric; Regard, Simon; Harbarth, Stephan; Achab, Sophia; Suppan, Mélanie | 2020 | JMIR research protocols | 9 | 12 | e25595 | 10.2196/25595 | No original data |

| Title | Authors | Year | Journal | Volume | Issue | Pages | DOI | Exclusion Reason |
|---|---|---|---|---|---|---|---|---|
| Impact of a Simulation-based Training Curriculum of Non-technical Skills on Colonoscopy Performance | Nct | 2016 | https://clinicaltrials.gov/show/NCT02877420 | | | | | Comparison group online |
| Impact of a drug allergy education course for non-specialists: findings from ADAPT—A randomized crossover trial | Lucas, M.; Mak, H. W. F.; Lee, J. T. Y.; Kulkarni, R.; Chan, S. S. C.; Li, P. H. | 2024 | Allergy | | | | 10.1111/all.16270 | Comparison group online |
| Impact of a multifaceted intervention on cholesterol management in primary care practices: guideline adherence for heart health randomized trial | Bertoni, Alain G.; Bonds, Denise E.; Chen, Haiying; Hogan, Patricia; Crago, Lenore; Rosenberger, Erica; Barham, Ann Hiott; Clinch, C. Randall; Goff, David C., Jr. | 2009 | Archives of internal medicine | 169 | 7 | 678-686 | 10.1001/archinternmed.2009.44 | Wrong intervention (no online learning) |
| Impact of a new teaching model on the fine cosmetic suturing operation and quantitative assessment of the training effect on plastic surgeons | Xu, Y.; Xing, J.; Wulan, H.; Guo, L. | 2023 | Chinese journal of plastic and reconstructive surgery | 5 | 1 | 20‑24 | 10.1016/j.cjprs.2022.09.014 | Wrong intervention (no online learning) |
| Impact of a pain management program on nurses' knowledge and attitude toward pain in United Arab Emirates: Experimental-four Solomon group design | Salim, Nezar Ahmed; Tuffaha, Mohammed Ghassan; Brant, Jeannine M. | 2020 | Applied nursing research : ANR | 54 | | 151314 | 10.1016/j.apnr.2020.151314 | Wrong intervention (no online learning) |
| Impact of an Educational Program on Improving Nurses' Management of Fever: An Experimental Study | Hsiao, Bi-Hung; Tzeng, Ya-Ling; Lee, Kwo-Chen; Lu, Shu-Hua; Lin, Yun-Ping | 2022 | Healthcare (Basel, Switzerland) | 10 | 6 | | 10.3390/healthcare10061135 | Wrong intervention (no online learning) |
| Impact of an online educational activity on oncologists' knowledge regarding the latest developments in the treatment of uveal melanoma | Dorkhom, N.; Peters, K.; Nathan, P.; Carvajal, R. D. | 2023 | ESMO Open | 8 | 1 Supplement 3 | 101074 | https://dx.doi.org/10.1016/j.esmoop.2023.101074 | Not randomised |
| Impact of eLearning course on nurses' professional competence in seclusion and restraint practices: 9-month follow-up results of a randomized controlled study (ISRCTN32869544) | Kontio, R.; Hätönen, H.; Joffe, G.; Pitkänen, A.; Lahti, M.; Välimäki, M. | 2013 | Journal of psychiatric and mental health nursing | 20 | 5 | 411-418 | 10.1111/j.1365-2850.2012.01933.x | Not healthcare clinical practice |
| Impact of eLearning course on nurses' professional competence in seclusion and restraint practices: a randomized controlled study (ISRCTN32869544) | Kontio, R.; Lahti, M.; Pitkänen, A.; Joffe, G.; Putkonen, H.; Hätönen, H.; Katajisto, J.; Välimäki, M. | 2011 | Journal of psychiatric and mental health nursing | 18 | 9 | 813-821 | 10.1111/j.1365-2850.2011.01729.x | Duplicate |
| Impact of guideline implementation on patient care: a cluster RCT | Mettes, T. G.; van der Sanden, W. J. M.; Bronkhorst, E.; Grol, R. P. T. M.; Wensing, M.; Plasschaert, A. J. M. | 2010 | Journal of dental research | 89 | 1 | 71-76 | 10.1177/0022034509350971 | Comparison group online |

| Title | Authors | Year | Journal | Volume | Issue | Pages | DOI | Exclusion Reason |
|---|---|---|---|---|---|---|---|---|
| Impact of pharmacists' training on the knowledge of pain and associated risks: results of the OPTYMED 2 study | Attal, N.; Clairaz-Mahiou, B.; Louis, P.; Annenkova, A.; Milon, J. Y.; Bismut, H.; Perrot, S. | 2024 | Presse medicale open | 5 | | | 10.1016/j.lpmope.2024.100050 | Not healthcare professionals |
| Impact of rheumatoid arthritis disease activity test on clinical practice | Peabody, John W.; Strand, Vibeke; Shimkhada, Riti; Lee, Rachel; Chernoff, David | 2013 | PloS one | 8 | 5 | e63215 | 10.1371/journal.pone.0063215 | Comparison group online |
| Impact of tele-advice on community nurses' knowledge of venous leg ulcer care | Ameen, Jamal; Coll, Anne Marie; Peters, Melanie | 2005 | Journal of advanced nursing | 50 | 6 | 583-594 | 10.1111/j.1365-2648.2005.03442.x | Wrong intervention (no online learning) |
| Impact of the CAPTURE COPD Screening Tool in US Primary Care: A Cluster Randomized Trial | Martinez, F. J.; Yawn, B. P.; Angulo, D.; Lopez, C.; Murray, S.; Mannino, D.; Anderson, S.; Dolor, R.; Elder, N.; Joo, M.; Khan, I.; Knox, L. M.; Meldrum, C.; Peters, E.; Spino, C.; Tapp, H.; Thomashow, B.; Zittleman, L.; Brown, R.; Make, B.; Han, M. K. | 2024 | American Journal of Respiratory and Critical Care Medicine | | | | https://dx.doi.org/10.1164/rccm.202405-0921OC | Wrong intervention (no online learning) |
| Impact of the Save the Shame! Game on Advanced Life Support Knowledge | Nct, | 2023 | https://clinicaltrials.gov/show/NCT05692375 | | | | | No data in abstract, and no PDF available |
| Impact of using a validated delirium screening tool, with or without a pharmacist and nurseled delirium education program, on the ability of nurses to recognize delirium in a surgicaltrauma ICU | Lin, A.; Russell, B.; Devlin, J.; Norton, H.; Evans, S.; Gesin, G. | 2010 | Critical Care Medicine | 38 | SUPPL. 12 | A10 | https://dx.doi.org/10.1097/01.ccm.0000390903.16849.8c | Not randomised |
| Impact of ward-based education opportunities and protected study days for 'diabetes nurse champions' on attendance andperception ofknowledgeandconfidence in managing inpatient diabetes issues | Skivington, C.; Robertson, M.; Cleland, S. | 2020 | Diabetic medicine | 37 | SUPPL 1 | 92- | 10.1111/dme.14245 | Wrong intervention (no online learning) |
| Impact of web-based learning for health program planning competency, knowledge and skills among mid-level public health nurses: A randomized controlled trial | Yoshioka-Maeda, Kyoko; Shiomi, Misa; Katayama, Takafumi; Hosoya, Noriko | 2019 | Public health nursing (Boston, Mass.) | 36 | 6 | 836-846 | 10.1111/phn.12642 | Not healthcare clinical practice |
| Implementation and Evaluation of Bright Futures Curriculum Within CORNET Continuity Practice | Nct | 2008 | https://clinicaltrials.gov/show/NCT00658489 | | | | | No original data |

| Title | Authors | Year | Journal | Volume | Issue | Pages | DOI | Exclusion Reason |
| --- | --- | --- | --- | --- | --- | --- | --- | --- |
| Implementation into practice: The development, enhancement and delivery of an online training programme to support clinicians in the replication of the back skills training (best) programme in practice | Richmond, H.; Hall, A. M.; Copsey, B.; Jones, G.; Williamson, E.; Hansen, Z.; Lamb, S. E. | 2017 | Trials | 18 | Supplement 1 | | https://dx.doi.org/10.1186/s13063-017-1902-y | Not randomised |
| Implementation of Practice Standards for ECG Monitoring | Nct | 2011 | https://clinicaltrials.gov/show/NCT01269736 | | | | | No original data |
| Implementation of a Multi-Modal Training Program for the Management of Comorbid Mental Disorders in Drug and Alcohol Settings: Pathways to Comorbidity Care (PCC) | Louie, Eva; Morley, Kirsten C.; Giannopoulos, Vicki; Uribe, Gabriela; Wood, Katie; Marel, Christina; Mills, Katherine L.; Teesson, Maree; Edwards, Michael; Childs, Steven; Rogers, David; Dunlop, Adrian; Baillie, Andrew; Haber, Paul S. | 2021 | Journal of dual diagnosis | 17 | 4 | 304-312 | 10.1080/15504263.2021.1984152 | Not randomised |
| Implementation of a new clinical practice guideline regarding pain management during childhood vaccine injections | Chan, Samson; Pielak, Karen; McIntyre, Cheryl; Deeter, Brittany; Taddio, Anna | 2013 | Paediatrics & child health | 18 | 7 | 367-372 | | Wrong intervention (no online learning) |
| Implementation of clinical guidelines on physical therapy for patients with low back pain: randomized trial comparing patient outcomes after a standard and active implementation strategy | Bekkering, Geertruida E.; van Tulder, Maurits W.; Hendriks, Erik J. M.; Koopmanschap, Marc A.; Knol, Dirk L.; Bouter, Lex M.; Oostendorp, Rob A. B. | 2005 | Physical therapy | 85 | 6 | 544-555 | | Wrong intervention (no online learning) |
| Implementation of the NHLBI integrated guidelines for cardiovascular health and risk reduction in children and adolescents: rationale and study design for young hearts, strong starts, a cluster-randomized trial targeting body mass index, blood pressure, a | LaBresh, Kenneth A.; Lazorick, Suzanne; Ariza, Adolfo J.; Furberg, Robert D.; Whetstone, Lauren; Hobbs, Connie; de Jesus, Janet; Bender, Randall H.; Salinas, Ilse G.; Binns, Helen J. | 2014 | Contemporary clinical trials | 37 | 1 | 98-105 | 10.1016/j.cct.2013.11.011 | Wrong intervention (no online learning) |
| Implementation of the best intervention: a group cognitive behavioural approach for patients with low back pain | Richmond, H.; Hansen, Z.; Davies, D.; Williamson, E.; Lamb, S. | 2015 | Physiotherapy (united kingdom) | 101 | | eS815-eS816 | 10.1016/j.physio.2015.03.3702 | No original data |
| Implementing the assessment for rehabilitation tool: A mixed methods, cluster randomised implementation trial | Lynch, E. A.; Luker, J. A.; Cadilhac, D. A.; Hillier, S. L. | 2015 | International Journal of Stroke | 10 | SUPPL. 3 | 33 | https://dx.doi.org/10.1111/ijs.12584 | Wrong intervention (no online learning) |

| Title | Authors | Year | Journal | Volume | Issue | Pages | DOI | Exclusion Reason |
|---|---|---|---|---|---|---|---|---|
| Implementing the mobile continuing medical education (mCME) project in Vietnam: making it work and sharing lessons learned | Bonawitz, Rachael; Bird, Liat; Le, Ngoc Bao; Nguyen, Viet Ha; Halim, Nafisa; Williams, Anna Larson; Sabin, Lora; Gill, Christopher J. | 2019 | MHealth | 5 | | 7 | 10.21037/mhealth.2019.02.01 | No original data |
| Improved Complex Skill Acquisition by Immersive Virtual Reality Training: A Randomized Controlled Trial | Lohre, Ryan; Bois, Aaron J.; Athwal, George S.; Goel, Danny P. | 2020 | The Journal of bone and joint surgery. American volume | 102 | 6 | e26 | 10.2106/JBJS.19.00982 | Wrong intervention (no online learning) |
| Improved electrocardiographic monitoring practices in the practical use of the latest standards of electrocardiography (PULSE) trial | Funk, M.; Fennie, K. P.; May, J. L.; Stephens, K.; Feder, S. L.; Chang, P. S.; Drew, B. J. | 2013 | Circulation | 128 | 22 SUPPL. 1 | | | Duplicate |
| Improvement in emergency physician stroke thrombolysis knowledge: the increasing tPA stroke treatment through interventional behavior change tactics (INSTINCT) trial | Meurer, W. J.; Frederiksen, S. M.; Xu, Z.; Kade, A. M.; Morgenstern, L. B.; Haan, M. N.; Kalbfleisch, J. D.; Scott, P. A. | 2011 | Stroke | 42 | 3 | e147- | 10.1161/STR.0b013e3182074d9b | Wrong intervention (no online learning) |
| Improvement of hypertension management by structured physician education and feedback system: cluster randomized trial | Lüders, Stefan; Schrader, Joachim; Schmieder, Roland E.; Smolka, Wenefrieda; Wegscheider, Karl; Bestehorn, Kurt | 2010 | European journal of cardiovascular prevention and rehabilitation : official journal of the European Society of Cardiology, Working Groups on Epidemiology & Prevention and Cardiac Rehabilitation and Exercise Physiology | 17 | 3 | 271-279 | 10.1097/HJR.0b013e328330be62 | Wrong intervention (no online learning) |
| Improving Health Professional Recognition and Response to Child Maltreatment and Intimate Partner Violence: protocol for Two Mixed Methods Pilot Randomized Controlled Trials | Kimber, M.; Baker-Sullivan, E.; Stewart, D. E.; Vanstone, M. | 2024 | JMIR research protocols | 13 | | e50864 | 10.2196/50864 | Comparison group online |
| Improving IUD counseling among internal medicine residents: a randomized control multi-site educational trial | Hirsch, H. D.; McNamara, M.; Spencer, A.; Batur, P.; Spanos, P. | 2016 | Journal of general internal medicine | 31 | Suppl. 2 | S827-S828 | 10.1007/s11606-016-3657-7 | Wrong intervention (no online learning) |
| Improving adherence to acute low back pain guideline recommendations with chiropractors and physiotherapists: the ALIGN cluster randomised controlled trial | French, Simon D.; O'Connor, Denise A.; Green, Sally E.; Page, Matthew J.; Mortimer, Duncan S.; Turner, Simon L.; Walker, Bruce F.; Keating, Jennifer L.; Grimshaw, Jeremy M.; Michie, Susan; Francis, Jill J.; McKenzie, Joanne E. | 2022 | Trials | 23 | 1 | 142 | 10.1186/s13063-022-06053-x | Wrong intervention (no online learning) |

| Title | Authors | Year | Journal | Volume | Issue | Pages | DOI | Exclusion Reason |
|---|---|---|---|---|---|---|---|---|
| Improving antibiotic prescribing quality by an intervention embedded in the primary care practice accreditation: the ARTI4 randomized trial | van der Velden, Alike W.; Kuyvenhoven, Marijke M.; Verheij, Theo J. M. | 2016 | The Journal of antimicrobial chemotherapy | 71 | 1 | 257-263 | 10.1093/jac/dkv328 | Comparison group online |
| Improving appropriate use of omega-3 fatty acids in primary care: Success of online cme | Spyropoulos, J.; Boutsalis, G.; Anderson, D. | 2019 | Circulation | 140 | Supplement 1 | | https://dx.doi.org/10.1161/circ.140.suppl_1.16351 | Not randomised |
| Improving care after myocardial infarction using a 2-year internet-delivered intervention: the Department of Veterans Affairs myocardial infarction-plus cluster-randomized trial | Levine, Deborah A.; Funkhouser, Ellen M.; Houston, Thomas K.; Gerald, Joe K.; Johnson-Roe, Nancy; Allison, Jeroan J.; Richman, Joshua; Kiefe, Catarina I. | 2011 | Archives of internal medicine | 171 | 21 | 1910-1917 | 10.1001/archinternmed.2011.498 | Comparison group online |
| Improving drug dose calculation skills among nurses: does choice of didactic method matter? A randomized controlled trial | Simonsen, B. O.; Daehlin, G. K.; Johansson, I.; Farup, P. G. | 2011 | International journal of clinical pharmacy | 33 | 2 | 339- | 10.1007/s11096-011-9481-6 | Duplicate |
| Improving knowledge on vaccine storage management in general practices: Learning effectiveness of an online-based program | Thielmann, Anika; Puth, Marie-Therese; Weltermann, Birgitta | 2020 | Vaccine | 38 | 47 | 7551-7557 | 10.1016/j.vaccine.2020.09.049 | Not randomised |
| Improving management of diabetic dyslipidemia: Effect of online CME | Spyropoulos, J. S.; Lacouture, M.; Chatterjee, P. | 2016 | Diabetes | 65 | Supplement 1 | A176 | https://dx.doi.org/10.2337/db16-652-860 | Not randomised |
| Improving on-line skills and knowledge. A randomized trial of teaching rural physicians to use on-line medical information | Kronick, Jonathan; Blake, Catherine; Munoz, Eeva; Heilbrunn, Lila; Dunikowski, Lynn; Milne, William Kenneth | 2003 | Canadian family physician Medecin de famille canadien | 49 | | 312-317 | | Wrong intervention (no online learning) |
| Improving rehabilitation assessment and referral practices for patients with stroke in Australia. A mixed methods cluster-randomized implementation trial | Lynch, E.; Luker, J. A.; Cadilhac, D. A.; Fryer, C. E.; Hillier, S. L. | 2015 | International Journal of Stroke | 10 | SUPPL. 2 | 178 | https://dx.doi.org/10.1111/ijs.12479 | Duplicate |
| Improving resident's skills in the management of circulatory shock with a knowledge-based e-learning tool | Riaño, David; Real, Francis; Alonso, Jose Ramon | 2018 | International journal of medical informatics | 113 | | 49-55 | 10.1016/j.ijmedinf.2018.02.006 | Not enough data for analysis |
| Improving safety of blood administration at Alfred health | Akers, C.; Miller, K.; Whitehead, S.; Magrin, G.; Davis, A. | 2012 | Transfusion Medicine | 22 | 3 | 230 | https://dx.doi.org/10.1111/j.1365-3148.2012.01160.x | No original data |

| Title | Authors | Year | Journal | Volume | Issue | Pages | DOI | Exclusion Reason |
|---|---|---|---|---|---|---|---|---|
| Improving the impact of didactic resident training with online spaced education | Gyorki, David E.; Shaw, Tim; Nicholson, James; Baker, Caroline; Pitcher, Meron; Skandarajah, Anita; Segelov, Eva; Mann, G. Bruce | 2013 | ANZ journal of surgery | 83 | 6 | 477-480 | 10.1111/ans.12166 | Wrong intervention (no online learning) |
| Improving triage, treatment and transfer of patients with stroke in Emergency Departments: the T3 trial | Middleton, S.; Considine, J.; Dale, S.; Cheung, N. W.; McInnes, E.; Levi, C.; D'Este, C.; Cadilhac, D.; Grimshaw, J.; Fitzgerald, M. | 2018 | EMA - emergency medicine australasia | 30 | | 12- | 10.1111/1742-6723.12961 | Wrong intervention (no online learning) |
| Improving vaccination uptake among older adults (= 60 years) | Drks | 2022 | https://trialsearch.who.int/Trial2.aspx?TrialID=DRKS00027252 | | | | | No data in abstract, and no PDF available |
| Increased Knowledge of Non-Stimulant Clinical Data and Mechanisms Accompanied by Increased Confidence of Therapeutic Selection for ADHD Among Psychiatrists Following Participation in an Online Medical Education Program | Finnegan, T.; Wright, C. W.; Lubarda, J.; O'Neal, W.; Busse, G.; Cutler, A. J. | 2021 | Annals of Clinical Psychiatry | 33 | 3 SUPPL | 20-21 | | Not randomised |
| Increased hand hygiene compliance in nursing homes after a multimodal intervention: A cluster randomized controlled trial (HANDSOME) | Teesing, Gwen R.; Erasmus, Vicki; Nieboer, Daan; Petrignani, Mariska; Koopmans, Marion P. G.; Vos, Margreet C.; Verduijn-Leenman, Annette; Schols, Jos M. G. A.; Richardus, Jan H.; Voeten, Helene A. C. M. | 2020 | Infection control and hospital epidemiology | 41 | 10 | 1169-1177 | 10.1017/ice.2020.319 | Not healthcare professionals |
| Increasing Access to Evidence-based Treatments for Depression | Nct | 2020 | https://clinicaltrials.gov/show/NCT04619615 | | | | | No data in abstract, and no PDF available |
| Increasing Primary Care Physician Colorectal Cancer Screening Rates | Nct | 2009 | https://clinicaltrials.gov/show/NCT00955344 | | | | | No original data |
| Increasing students' and new professionals' knowledge of child sexual abuse outcomes: an evaluation of an online intervention | Theimer, Kate | 2020 | | 81 | | | | Not randomised |

| Title | Authors | Year | Journal | Volume | Issue | Pages | DOI | Exclusion Reason |
|---|---|---|---|---|---|---|---|---|
| Increasing the quantity and quality of searching for current best evidence to answer clinical questions: protocol and intervention design of the MacPLUS FS Factorial Randomized Controlled Trials | Agoritsas, Thomas; Iserman, Emma; Hobson, Nicholas; Cohen, Natasha; Cohen, Adam; Roshanov, Pavel S.; Perez, Miguel; Cotoi, Chris; Parrish, Rick; Pullenayegum, Eleanor; Wilczynski, Nancy L.; Iorio, Alfonso; Haynes, R. Brian | 2014 | Implementation science : IS | 9 | | 125 | 10.1186/s13012-014-0125-9 | No original data |
| Influencing Perceptions and Referrals by Primary Care Providers to Complementary Integrative Health | Adan, F.; Dusek, J. | 2021 | European journal of integrative medicine | 48 | | | 10.1016/j.eujim.2021.101912 | Not randomised |
| Innovative web-based multimedia curriculum improves cardiac examination competency of residents | Criley, Jasminka M.; Keiner, Jennifer; Boker, John R.; Criley, Stuart R.; Warde, Carole M. | 2008 | Journal of hospital medicine | 3 | 2 | 124-133 | 10.1002/jhm.287 | Not randomised |
| Integrating Simulation-Based Education and Precision Teaching to Improve Physicians' Performance of Lumbar Puncture in Clinical Practice | Reid-McDermott, B.; Lydon, S.; O'Connor, P.; De Bhulbh, A.; Hanahoe, B.; Byrne, D. | 2019 | BMJ simulation and technology enhanced learning | 5 | | A66-A67 | 10.1136/bmjstel-2019-aspihconf.123 | Not randomised |
| Integrative medicine: implementation and evaluation of a professional development program using experiential learning and conceptual change teaching approaches | Hewson, Mariana G.; Copeland, H. Liesel; Mascha, Edward; Arrigain, Susana; Topol, Eric; Fox, Joan E. B. | 2006 | Patient education and counseling | 62 | 1 | 5-Dec | 10.1016/j.pec.2006.03.005 | Wrong intervention (no online learning) |
| Interactive CME improves primary care physicians'knowledge and confidence regarding T2D and HF management | Larkin, A.; Healy, C.; Le, A. | 2018 | Endocrine Reviews | 39 | 2 Supplement 1 | | | Not randomised |
| Interactive Spaced Education improves clinicians' screening for prostate cancer: a multi-institutional randomized controlled trial | Kerfoot, B. P.; Lawler, E. V.; Sokolovskaya, G.; Gagnon, D.; Conlin, P. R. | 2009 | Journal of urology | 181 | 4 | 190- | | No original data |
| Interactive Spaced Education to Optimize Hypertension Management | Nct | 2009 | https://clinicaltrials.gov/show/NCT00904007 | | | | | Comparison group online |
| Interactive Spaced Online Education in Pediatric Trauma | Shenoi, R.; Rubalcava, D.; Naik-Mathuria, B.; Sloas, H. A.; Delemos, D.; Xu, L.; Mendez, D. | 2016 | SAGE Open | 6 | 2 | | 10.1177/2158244016653167 | Comparison group online |
| Interactive spaced education to assess and improve knowledge of clinical practice guidelines: a randomized controlled trial | Kerfoot, B. Price; Kearney, Michael C.; Connelly, Donna; Ritchey, Michael L. | 2009 | Annals of surgery | 249 | 5 | 744-749 | 10.1097/SLA.0b013e31819f6db8 | Comparison group online |

| Title | Authors | Year | Journal | Volume | Issue | Pages | DOI | Exclusion Reason |
|---|---|---|---|---|---|---|---|---|
| Interactive workshops increase chlamydia testing in primary care--a controlled study | McNulty, Cliodna Am; Thomas, Michael; Bowen, Joanne; Buckley, Charles; Charlett, Andre; Gelb, David; Foy, Chris; Sloss, John; Smellie, Stuart | 2008 | Family practice | 25 | 4 | 279-286 | 10.1093/fampra/cmn032 | Wrong intervention (no online learning) |
| Internet Intervention to Improve Rural Diabetes Care | Nct | 2006 | https://clinicaltrials.gov/show/NCT00403091 | | | | | Comparison group online |
| Internet training for nurse aides to prevent resident aggression | Irvine, A. Blair; Bourgeois, Michelle; Billow, Molly; Seeley, John R. | 2007 | Journal of the American Medical Directors Association | 8 | 8 | 519-526 | 10.1016/j.jamda.2007.05.002 | Not healthcare clinical practice |
| Investigating the effect of education based on PRECEDE-PROCEED model on the preventive behaviors of musculoskeletal disorders in a group of nurses | Rakhshani, T.; Limouchi, Z.; Daneshmandi, H.; Kamyab, A.; Jeihooni, A. K. | 2024 | Frontiers in public health | 12 | | 1371684 | 10.3389/fpubh.2024.1371684 | Wrong intervention (no online learning) |
| Investigation and standardization of a neurology department's protocols for the ability to recognize, triage and treat chemical nerve agent attacks | Wright, P.; Thirunavuukarasu, S.; Rush, S.; Molmenti, C.; Coffield, E. | 2017 | Neurology | 88 | 16 | | | No original data |
| Is chronic breathlessness less recognized and treated compared with chronic pain? A case-based randomised controlled trial | Ekstrom, M. P.; Sandberg, J.; Currow, D.; Ahmadi, Z. | 2018 | American journal of respiratory and critical care medicine | 197 | Meeting Abstracts | | | Wrong intervention (no online learning) |
| Keeping oncologists current with CDK4/6 inhibitors in HR+ breast cancer: The impact of online education | Parikh, K.; Tanzola, M.; Peters, P. M.; Cristofanilli, M. | 2020 | Journal of Clinical Oncology | 38 | 15 | | https://dx.doi.org/10.1200/JCO.2020.38.15-suppl.e13044 | Not randomised |
| Knowledge and health care resource allocation: CME/CPD course guidelines-based efficacy | Braido, F.; Comaschi, M.; Valle, I.; Delgado, L.; Coccini, A.; Guerreras, P.; Stagi, E.; Canonica, G. W. | 2012 | European annals of allergy and clinical immunology | 44 | 5 | 193-199 | | Not enough data for analysis |
| Knowledge transfer and retention of simulation-based learning for neurosurgical instruments: a randomised trial of perioperative nurses | Clarke, David B.; Galilee, Alena I.; Kureshi, Nelofar; Hong, Murray; Fenerty, Lynne; D'Arcy, Ryan C. N. | 2020 | BMJ simulation & technology enhanced learning | 7 | 3 | 146-153 | 10.1136/bmjstel-2019-000576 | Comparison group online |
| Late Breaking Abstract - Training thoracic ultrasound skills: a multicentre, blinded, randomized controlled trial of simulation-based training versus training on healthy figurants | Pietersen, P. I.; Jorgensen, R.; Graumann, O.; Konge, L.; Skaarup, S. H.; Schultz, H. H. L.; Laursen, C. B. | 2019 | European respiratory journal | 54 | | | 10.1183/13993003.congress-2019.OA5341 | Comparison group online |
| Learning and retention from an online tutorial among resident physicians | Bell, Douglas S.; Higa, Jerilyn; Mangione, Carol M. | 2007 | AMIA ... Annual Symposium proceedings. AMIA Symposium | | | 870 | | Comparison group online |

| Title | Authors | Year | Journal | Volume | Issue | Pages | DOI | Exclusion Reason |
|---|---|---|---|---|---|---|---|---|
| Learning with computerized guidelines in general practice?: A randomized controlled trial | Butzlaff, M.; Vollmar, H. C.; Floer, B.; Koneczny, N.; Isfort, J.; Lange, S. | 2004 | Family practice | 21 | 2 | 183-188 | 10.1093/fampra/cmh214 | Wrong intervention (no online learning) |
| Lessons Learned From a Practice-Based, Multisite Intervention Study With Nurse Participants | Friese, Christopher R.; Mendelsohn-Victor, Kari; Ginex, Pamela; McMahon, Carol M.; Fauer, Alex J.; McCullagh, Marjorie C. | 2017 | Journal of nursing scholarship : an official publication of Sigma Theta Tau International Honor Society of Nursing | 49 | 2 | 194-201 | 10.1111/jnu.12279 | Comparison group online |
| Long-term impact of four different strategies for delivering an on-line curriculum about herbs and other dietary supplements | Beal, Tiffany; Kemper, Kathi J.; Gardiner, Paula; Woods, Charles | 2006 | BMC medical education | 6 | | 39 | 10.1186/1472-6920-6-39 | Comparison group online |
| Longitudinal effects of single-dose simulation education with structured debriefing and verbal feedback on endotracheal suctioning knowledge and skills: A randomized controlled trial | Jansson, Miia M.; Syrjälä, Hannu P.; Ohtonen, Pasi P.; Meriläinen, Merja H.; Kyngäs, Helvi A.; Ala-Kokko, Tero I. | 2017 | American journal of infection control | 45 | 1 | 83-85 | 10.1016/j.ajic.2016.05.032 | Wrong intervention (no online learning) |
| Lung Cancer Screening Decision Aid Designed for a Primary Care Setting: A Randomized Clinical Trial | Schapira, M. M.; Hubbard, R. A.; Whittle, J.; Vachani, A.; Kaminstein, D.; Chhatre, S.; Rodriguez, K. L.; Bastian, L. A.; Kravetz, J. D.; Asan, O.; Prigge, J. M.; Meline, J.; Schrand, S.; Ibarra, J. V.; Dye, D. A.; Rieder, J. B.; Frempong, J. O.; Fraenkel, L. | 2023 | JAMA Network Open | 6 | 8 | E2330452 | https://dx.doi.org/10.1001/jamanetworkopen.2023.30452 | Not healthcare professionals |
| MOOC learners' engagement with two variants of virtual patients: A randomised trial | Stathakarou, N.; Scully, M. L.; Kononowicz, A. A.; Henningsohn, L.; Zary, N.; McGrath, C. | 2018 | Education Sciences | 8 | 2 | | 10.3390/educsci8020044 | Not healthcare professionals |
| Make a Journal Feel as if it Was Mailed Back From the Future | Nct | 2015 | https://clinicaltrials.gov/show/NCT02508051 | | | | | No original data |
| Measuring and Improving Evidence-Based Patient Care Using a Web-Based Gamified Approach in Primary Care (QualityIQ): Randomized Controlled Trial | Burgon, Trever; Casebeer, Linda; Aasen, Holly; Valdenor, Czarlota; Tamondong-Lachica, Diana; de Belen, Enrico; Paculdo, David; Peabody, John | 2021 | Journal of medical Internet research | 23 | 12 | e31042 | 10.2196/31042 | Comparison group online |
| Medical outcomes in acls knowledge (MOCK): the effect of standardized resident acls simulation training | Han, J.; Trammell, A.; Finklea, J.; Udoji, T.; Dressler, D.; Honig, E.; Abraham, P.; Fisher, M.; Ander, D.; Martin, G.; et al. | 2011 | Chest | 140 | 4 MEETING ABSTRACT | | 10.1378/chest.1109051 | Wrong intervention (no online learning) |

| Title | Authors | Year | Journal | Volume | Issue | Pages | DOI | Exclusion Reason |
|---|---|---|---|---|---|---|---|---|
| Mental Vitality at Work: A mental health module for Workers' Health Surveillance can improve work functioning | Ketelaar, S.; Nieuwenhuijsen, K.; Gärtner, F.; Bolier, L.; Smeets, O.; Van Dijk, F.; Sluiter, J. | 2013 | Tijdschrift voor Bedrijfs- en Verzekeringsgeneeskunde | 21 | 9 | 406-411 | 10.1007/s12498-013-0190-z | Not healthcare clinical practice |
| Mental health training for nursing home staff using computer-based interactive video: a 6-month randomized trial | Rosen, Jules; Mulsant, Benoit H.; Kollar, Marcia; Kastango, Kari B.; Mazumdar, Sati; Fox, Debra | 2002 | Journal of the American Medical Directors Association | 3 | 5 | 291-296 | 10.1097/01.JAM.0000027201.90817.21 | Wrong intervention (no online learning) |
| Mobile Learning to Improve Clinician's Ability to Break Bad News | Nct | 2019 | https://clinicaltrials.gov/show/NCT03804918 | | | | | No original data |
| Modifying dyspepsia management in primary care: a cluster randomised controlled trial of educational outreach compared with passive guideline dissemination | Banait, Gurvinder; Sibbald, Bonnie; Thompson, David; Summerton, Chris; Hann, Mark; Talbot, Stuart | 2003 | The British journal of general practice : the journal of the Royal College of General Practitioners | 53 | 487 | 94-100 | | Wrong intervention (no online learning) |
| Motivating HIV Providers in Vietnam to Learn: A Mixed-Methods Analysis of a Mobile Health Continuing Medical Education Intervention | Larson Williams, Anna; Hawkins, Andrew; Sabin, Lora; Halim, Nafisa; Le Ngoc, Bao; Nguyen, Viet Ha; Nguyen, Tam; Bonawitz, Rachael; Gill, Christopher | 2019 | JMIR medical education | 5 | 1 | e12058 | 10.2196/12058 | Not randomised |
| Multi-institutional validation and assessment of training modalities in robotic surgery (the mars project) | Raison, N.; Ahmed, K.; Aydin, A.; Mottrie, A.; Van Der Poel, H.; Dasgupta, P. | 2016 | Journal of urology | 195 | 4 | e117- | | Wrong intervention (no online learning) |
| Multicentre Randomised Controlled Trial to Assess the Impact of Online Training on Ct Head Interpretation Performance: The Simulation Training for Emergency Department Imaging 2 (Stedi2) Trial | Novak, A.; Ather, S.; Martinez, J.; Baron, T.; Triscott, S.; Davies, M.; Gulati, D.; Shashikala, R.; Wilson, S.; Keating, L. | 2023 | Emergency medicine journal | 40 | 12 | 872-873 | 10.1136/emj-2023-RCEM.23 | Not enough data for analysis |
| Multicentre study protocol comparing standard NRP to deveLoped Educational Modules for Resuscitation of Neonates in the Delivery Room with Congenital Heart Disease (LEARN-CHD) | Levy, P.; Thomas, A. R.; Law, B. H. Y.; Joynt, C.; Gupta, R.; Elshenawy, S.; Reed, D.; Pavlek, L. R.; Shepherd, J.; Gowda, S.; Johnson, B. A.; Ball, M.; Ali, N. | 2023 | BMJ Open | 13 | 4 | | 10.1136/bmjopen-2022-067391 | Comparison group online |

| Title | Authors | Year | Journal | Volume | Issue | Pages | DOI | Exclusion Reason |
|---|---|---|---|---|---|---|---|---|
| Multicomponent Internet continuing medical education to promote chlamydia screening | Allison, Jeroan J.; Kiefe, Catarina I.; Wall, Terry; Casebeer, Linda; Ray, Midge N.; Spettell, Claire M.; Hook, Edward W., 3rd; Oh, M. Kim; Person, Sharina D.; Weissman, Norman W. | 2005 | American journal of preventive medicine | 28 | 3 | 285-290 | 10.1016/j.amepre.2004.12.013 | Comparison group online |
| Multisite Single-Blinded Randomized Control Study of Transfer and Retention of Knowledge and Skill Between Nurses Using Simulation and Online Self-Study Module | Rutherford-Hemming, Tonya; Kelsey, Nichole C.; Grenig, Deanna L.; Feliciano, Michelle; Simko, Leslie; Henrich, Christina M. | 2016 | Simulation in healthcare : journal of the Society for Simulation in Healthcare | 11 | 4 | 264-270 | 10.1097/SIH.0000000000000168 | Wrong intervention (no online learning) |
| Nurse-led Antimicrobial Stewardship Intervention to Increase Antibiotic Appropriateness in Residential Aged Care Facilities | Nct | 2019 | https://clinicaltrials.gov/show/NCT03941509 | | | | | No original data |
| Nurses' Knowledge and Skills After Use of an Augmented Reality App for Advanced Cardiac Life Support Training: Randomized Controlled Trial | Sun, W. N.; Hsieh, M. C.; Wang, W. F. | 2024 | Journal of Medical Internet Research | 26 | | e57327 | https://dx.doi.org/10.2196/57327 | Not randomised |
| Nursing Home Characteristics Associated With Implementation of an Advance Care Planning Video Intervention | Loomer, Lacey; McCreedy, Ellen; Belanger, Emmanuelle; Palmer, Jennifer A.; Mitchell, Susan L.; Volandes, Angelo E.; Mor, Vincent | 2019 | Journal of the American Medical Directors Association | 20 | 7 | 804 | 10.1016/j.jamda.2019.01.133 | Not healthcare professionals |
| Online Continuing Medical Education Program Improves Knowledge among Neurologists and Primary Care Physicians Regarding the Management of Opioid Withdrawal Syndrome | Finnegan, T.; Murray, C.; Lubarda, J.; Kosten, T. | 2019 | Postgraduate Medicine | 131 | SUPPL 1 | 114 | https://dx.doi.org/10.1080/00325481.2019.1655695 | Not randomised |
| Online Medical Education Improves Knowledge of Data on Appropriate and Timely Use of Influenza Antiviral Medications to Patients at High Risk for Influenza-Related Complications and Morbidity | Armagan, A.; Smith, R. | 2021 | Open Forum Infectious Diseases | 8 | SUPPL 1 | S577-S578 | https://dx.doi.org/10.1093/ofid/ofab466.1166 | Not randomised |
| Online Medical Education Improves Knowledge of Recent Data in Epilepsy and Related Clinical Applicability Among General Neurologists and Epileptologists | Finnegan, T.; Merlo, K.; Lubarda, J.; Ferrari, L.; Krauss, G.; Sperling, M. | 2022 | Neurology | 98 | 18 SUPPL | | | Not randomised |
| Online Regional Anesthesia Resources - Are They Effective? | Nct | 2022 | https://clinicaltrials.gov/show/NCT05290974 | | | | | No original data |

| Title | Authors | Year | Journal | Volume | Issue | Pages | DOI | Exclusion Reason |
|---|---|---|---|---|---|---|---|---|
| Online Training & Certification for Competency in Dementia Friendly Hospital Care | Nct | 2019 | https://clinicaltrials.gov/show/NCT04182282 | | | | | No original data |
| Online Training Program Model for Effective Management of Nursing Services in Times of Crisis Such as Pandemic | Nct | 2021 | https://clinicaltrials.gov/show/NCT04778995 | | | | | Not healthcare clinical practice |
| Online cme successful at improving nephrologist understanding of emerging class to treat anemia associated with CKD | Larkin, A.; Anderson, D. R.; Boutsalis, G. | 2020 | Journal of the American Society of Nephrology | 31 | | 139 | https://dx.doi.org/10.1681/asn.2019040398 | Not randomised |
| Online cognitive behavioural therapy training for therapists: Outcomes, acceptability, and impact of support | Bennett-Levy, James; Hawkins, Russell; Perry, Helen; Cromarty, Paul; Mills, Jeremy | 2012 | Australian Psychologist | 47 | 3 | 174-182 | 10.1111/j.1742-9544.2012.00089.x | Comparison group online |
| Online education boosts clinician knowledge about emerging therapies for patients with systemic sclerosis-associated interstitial lung disease | Rohani-Montez, C.; Calle, M.; Allen, C.; Hoffmann-Vold, A. M.; Distler, O. | 2020 | Annals of the Rheumatic Diseases | 79 | SUPPL 1 | 1927 | https://dx.doi.org/10.1136/annrheumdis-2020-eular.457 | Not randomised |
| Online education significantly improved rheumatologists' understanding and interpretation of comparative treatment data for As | Stan, A.; Bell, E.; Schoonheim, P.; Baraliakos, X. | 2019 | Annals of the Rheumatic Diseases | 78 | Supplement 2 | 1449 | https://dx.doi.org/10.1136/annrheumdis-2019-eular.2997 | Not randomised |
| Online education yields significant gains in rheumatologists' knowledge of psoriatic disease | Stan, A.; Calle, M.; Schoonheim, P.; Gottlieb, A. B. | 2021 | Annals of the Rheumatic Diseases | 80 | SUPPL 1 | 1448-1449 | https://dx.doi.org/10.1136/annrheumdis-2021-eular.1569 | Not randomised |
| Online education yields significant gains in rheumatologists' knowledge of the role of enthesitis in the diagnosis and management of psa | Stan, A.; Calle, M.; Schoonheim, P.; Mease, P. J. | 2020 | Annals of the Rheumatic Diseases | 79 | SUPPL 1 | 570 | https://dx.doi.org/10.1136/annrheumdis-2020-eular.3137 | Not randomised |
| Online education yields significant gains in rheumatologists' knowledge of the role of jak inhibitors in the management of ra | Bell, E.; Calle, M.; Van Vollenhoven, R. | 2020 | Annals of the Rheumatic Diseases | 79 | SUPPL 1 | 1925 | https://dx.doi.org/10.1136/annrheumdis-2020-eular.2294 | Not randomised |
| Online medical education improves knowledge of both risk factors for chronification and clinical trial data in migraine | Finnegan, T.; Ullman, S. | 2018 | Headache | 58 | Supplement 2 | 158-159 | | Not randomised |
| Online training resources to aid therapeutic radiographers in engaging in conversations about physical activity and diet: A mixed methods study | Pallin, N. D.; Webb, J.; Brown, L.; Woznitza, N.; Stewart-Lord, A.; Charlesworth, L.; Beeken, R. J.; Fisher, A. | 2022 | Radiography (London, England : 1995) | 28 | 1 | 124-132 | 10.1016/j.radi.2021.09.004 | Comparison group online |

| Title | Authors | Year | Journal | Volume | Issue | Pages | DOI | Exclusion Reason |
|---|---|---|---|---|---|---|---|---|
| Optimal Use of HPV Vaccine Use in Primary Care | Nct | 2019 | https://clinicaltrials.gov/show/NCT04180462 | | | | | No original data |
| Optimizing delivery of health care interventions | Isrctn | 2012 | https://trialsearch.who.int/Trial2.aspx?TrialID=ISRCTN21998507 | | | | | Duplicate |
| Optimizing educational video through comparative trials in clinical environments | Aronson, I. D.; Plass, J. L.; Bania, T. C. | 2012 | Educational Technology Research and Development | 60 | 3 | 469-482 | 10.1007/s11423-011-9231-4 | Not healthcare professionals |
| Optimizing guideline adherence of rheumatologists | Ntr | 2014 | https://trialsearch.who.int/Trial2.aspx?TrialID=NTR4449 | | | | | No original data |
| Osteoporosis improvement: a large-scale randomized controlled trial of patient and primary care physician education | Solomon, Daniel H.; Katz, Jeffrey N.; Finkelstein, Joel S.; Polinski, Jennifer M.; Stedman, Margaret; Brookhart, M. Alan; Arnold, Marilyn; Gauthier, Suzanne; Avorn, Jerry | 2007 | Journal of bone and mineral research : the official journal of the American Society for Bone and Mineral Research | 22 | 11 | 1808-1815 | 10.1359/jbmr.070717 | Wrong intervention (no online learning) |
| Outcomes from the trial implementation of a multidisciplinary online learning program in rural mental health emergency care | Hills, D. J.; Robinson, T.; Kelly, B.; Heathcote, S. | 2010 | Education for health (Abingdon, England) | 23 | 1 | 351 | | Not randomised |
| Overcoming Therapeutic Inertia in Multiple Sclerosis Care: A Pilot Randomized Trial Applying the Traffic Light System in Medical Education | Saposnik, Gustavo; Maurino, Jorge; Sempere, Angel P.; Terzaghi, Maria A.; Ruff, Christian C.; Mamdani, Muhammad; Tobler, Philippe N.; Montalban, Xavier | 2017 | Frontiers in neurology | 8 | | 430 | 10.3389/fneur.2017.00430 | Intervention <10 mins |
| Overcoming barriers to disseminating exposure therapies for anxiety disorders: a pilot randomized controlled trial of training methods | Harned, Melanie S.; Dimeff, Linda A.; Woodcock, Eric A.; Skutch, Julie M. | 2011 | Journal of anxiety disorders | 25 | 2 | 155-163 | 10.1016/j.janxdis.2010.08.015 | Comparison group online |
| P.0090 Continuing medical education curriculum improves psychiatrists' knowledge, competence and confidence on management of major depressive disorder | Thevathasan, L.; Fairley, L.; Phillips, C.; Chen, P.; Demyttenaere, K.; Vieta, E.; Falkai, P.; Goodwin, G. | 2021 | European Neuropsychopharmacology | 53 | Supplement 1 | S63-S64 | https://dx.doi.org/10.1016/j.euroneuro.2021.10.090 | Not randomised |
| PTSD Training for PCPs in a Virtual World | Nct | 2019 | https://clinicaltrials.gov/show/NCT03898271 | | | | | Comparison group online |
| Pain Management Algorithms for Implementing Best Practices in Nursing Homes: Results of a Randomized Controlled Trial | Ersek, Mary; Neradilek, Moni Blazej; Herr, Keela; Jablonski, Anita; Polissar, Nayak; Du Pen, Anna | 2016 | Journal of the American Medical Directors Association | 17 | 4 | 348-356 | 10.1016/j.jamda.2016.01.001 | Wrong intervention (no online learning) |

| Title | Authors | Year | Journal | Volume | Issue | Pages | DOI | Exclusion Reason |
|---|---|---|---|---|---|---|---|---|
| Patient and Provider Web-Based Decision Support for Breast Cancer Chemoprevention: A Randomized Controlled Trial | Crew, Katherine D.; Bhatkhande, Gauri; Silverman, Thomas; Amenta, Jacquelyn; Jones, Tarsha; McGuinness, Julia E.; Mata, Jennie; Guzman, Ashlee; He, Ting; Dimond, Jill; Tsai, Wei-Yann; Kukafka, Rita | 2022 | Cancer prevention research (Philadelphia, Pa.) | 15 | 10 | 689-700 | 10.1158/1940-6207.CAPR-22-0013 | Comparison group online |
| Patterns and rates of use of an evidence-based practice intranet resource for allied health professionals: a randomised controlled trial | Campbell, L.; Novak, I.; McIntyre, S.; Dickson, A. | 2010 | Developmental medicine and child neurology | 52 | | 31‑ | 10.1111/j.1469-8749.2009.03594.x | Wrong intervention (no online learning) |
| Pediatric Code Cart Training: can Augmented Reality Improve Pediatric Readiness for Emergency Medicine Residents | Roszczynialski, K.; Lee, M.; Wang, Y.; Darji, H.; Schertzer, K.; Jones, J.; Pokrajac, N.; Krzyzaniak, S. | 2024 | Annals of Emergency Medicine | 84 | 4 | S16‑S17 | 10.1016/j.annemergmed.2024.08.043 | Comparison group online |
| Peer-to-Peer JXTA Architecture for Continuing Mobile Medical Education Incorporated in Rural Public Health Centers | Rajasekaran, Rajkumar; Iyengar, Nallani Chackravatula Sriman Narayana | 2013 | Osong public health and research perspectives | 4 | 2 | 99-106 | 10.1016/j.phrp.2013.03.004 | Not randomised |
| Phase II Motivational Interviewing: an Experiential Online Training Tool | Nct | 2014 | https://clinicaltrials.gov/show/NCT02264327 | | | | | No data in abstract, and no PDF available |
| Physician Versus Nonphysician Instruction: Evaluating an Expert Curriculum-Competent Facilitator Model for Simulation-Based Central Venous Catheter Training | Musits, Andrew N.; Phrampus, Paul E.; Lutz, John W.; Bear, Todd M.; Maximous, Stephanie I.; Mrkva, Andrew J.; O’Donnell, John M. | 2019 | Simulation in healthcare : journal of the Society for Simulation in Healthcare | 14 | 4 | 228-234 | 10.1097/SIH.0000000000000374 | Not healthcare professionals |
| Pilot Study of an Online Parent-Training Course for Disruptive Behavior with Live Remote Coaching for Practitioners | Ortiz, Camilo; Vidair, Hilary; Acri, Mary; Chacko, Anil; Kobak, Kenneth | 2020 | Professional psychology, research and practice | 51 | 2 | 125-133 | 10.1037/pro0000286 | Not randomised |
| Pilot testing of a new approach to improving the prescribing of many drugs for older people who live in their own home and are cared for by general practitioners | Isrctn | 2019 | https://trialsearch.who.int/Trial2.aspx?TrialID=ISRCTN41009897 | | | | | Duplicate |
| Portable Video Media Versus Standard Verbal Communication in Surgical Information Delivery to Nurses: A Prospective Multicenter, Randomized Controlled Crossover Trial | Kam, Jonathan; Ainsworth, Hannah; Handmer, Marcus; Louie-Johnsun, Mark; Winter, Matthew | 2016 | Worldviews on evidence-based nursing | 13 | 5 | 363-370 | 10.1111/wvn.12162 | Wrong intervention (no online learning) |

| Title | Authors | Year | Journal | Volume | Issue | Pages | DOI | Exclusion Reason |
|---|---|---|---|---|---|---|---|---|
| Potential unintended consequences due to Medicare's "no pay for errors" rule? A randomized controlled trial of an educational intervention with internal medicine residents | Mookherjee, Somnath; Vidyarthi, Arpana R.; Ranji, Sumant R.; Maselli, Judy; Wachter, Robert M.; Baron, Robert B. | 2010 | Journal of general internal medicine | 25 | 10 | 1097-1101 | 10.1007/s11606-010-1395-9 | Wrong intervention (no online learning) |
| Pragmatic Implementation of Online Obesity Treatment and Maintenance Interventions in Primary Care: a Randomized Clinical Trial | Thomas, J. G.; Panza, E.; Goldstein, C. M.; Hayes, J. F.; Benedict, N.; O'Leary, K.; Wing, R. R. | 2024 | JAMA Internal Medicine | 184 | 5 | 502-509 | 10.1001/jamainternmed.2023.8438 | Not healthcare professionals |
| Preventing Early Dialysis Starts | Nct | 2014 | https://clinicaltrials.gov/show/NCT02183987 | | | | | Wrong intervention (no online learning) |
| Preventing disease through opportunistic, rapid engagement by primary care teams using behaviour change counselling (PRE-EMPT): protocol for a general practice-based cluster randomised trial | Spanou, Clio; Simpson, Sharon A.; Hood, Kerry; Edwards, Adrian; Cohen, David; Rollnick, Stephen; Carter, Ben; McCambridge, Jim; Moore, Laurence; Randell, Elizabeth; Pickles, Timothy; Smith, Christine; Lane, Claire; Wood, Fiona; Thornton, Hazel; Butler, Chris C. | 2010 | BMC family practice | 11 | | 69 | 10.1186/1471-2296-11-69 | No knowledge, skills, or clinical outcome |
| Process of developing an interdisciplinary obstetric anaesthesia simulation-based education course with involvement of six health-care professions | Ekelund, K.; Albrechtsen, C. K.; Ostergaard, D.; Sorensen, J. L. | 2013 | Acta Anaesthesiologica Scandinavica, Supplement | 57 | SUPPL. 120 | 8 | https://dx.doi.org/10.1111/aas.12153 | No data in abstract, and no PDF available |
| Project to implement an adequate pain management in nursing homes | Drks | 2016 | https://trialsearch.who.int/Trial2.aspx?TrialID=DRKS00011062 | | | | | Not healthcare professionals |
| Promoting Adherence to Influenza Vaccination Recommendations in Pediatric Practice | Werk, Lloyd N.; Diaz, Maria Carmen; Cadilla, Adriana; Franciosi, James P.; Hossain, Md Jobayer | 2019 | Journal of primary care & community health | 10 | | 2.15013E+15 | 10.1177/2150132719853061 | Not enough data for analysis |
| Promoting Hand Hygiene Compliance: PSYGIENE—a Cluster-Randomized Controlled Trial of Tailored Interventions | von Lengerke, Thomas; Lutze, Bettina; Krauth, Christian; Lange, Karin; Stahmeyer, Jona Theodor; Chaberny, Iris Freya | 2017 | Deutsches Arzteblatt international | 114 | 3 | 29-36 | 10.3238/arztebl.2017.0029 | Wrong intervention (no online learning) |

| Title | Authors | Year | Journal | Volume | Issue | Pages | DOI | Exclusion Reason |
|---|---|---|---|---|---|---|---|---|
| Promoting Informed Decisions About Colorectal Cancer Screening in Older Adults (PRIMED Study): a Physician Cluster Randomized Trial | Sepucha, K.; Han, P. K. J.; Chang, Y.; Atlas, S. J.; Korsen, N.; Leavitt, L.; Lee, V.; Percac-Lima, S.; Mancini, B.; Richter, J.; Scharnetzki, E.; Siegel, L. C.; Valentine, K. D.; Fairfield, K. M.; Simmons, L. H. | 2023 | Journal of General Internal Medicine | 38 | 2 | 406 | https://dx.doi.org/10.1007/s11606-022-07738-4 | Not healthcare professionals |
| Promoting Positive Care Interactions (PPCI) in Assisted Living | Nct | 2022 | https://clinicaltrials.gov/show/NCT05618834 | | | | | No original data |
| Promoting evidence-based best practices for hip fracture prevention in residential aged care | Scherer, S. C.; Jennings, C.; Rule, J.; Smeaton, M.; Farrell, M. J.; Garratt, S. A.; Flicker, L.; Davis, I.; Wark, J. D. | 2006 | Australasian Journal on Ageing | 25 | 4 | 185-190 | 10.1111/j.1741-6612.2006.00182.x | Wrong intervention (no online learning) |
| Promoting oral cancer examination to primary care providers at medical clinics in nebraska | Wee, Alvin G. | 2015 | | 76 | | | | Duplicate |
| Promoting rational antibiotic therapy in German primary care | Isrctn | 2022 | https://trialsearch.who.int/Trial2.aspx?TrialID=ISRCTN95468513 | | | | | No original data |
| Promotion of urine microalbuminuria screening among primary care physicians: a randomized, controlled, educational intervention trial | Naimark, D. M. J.; Bott, M. T.; Tobe, S. W.; Reznick, R. K.; David, D. | 2001 | Journal of the American Society of Nephrology : JASN | 12 | Program & Abstracts | 231A | | No data in abstract, and no PDF available |
| Prospective study comparing virtual reality to live demonstration for neonatal intubation education | O'Sullivan, D.; Bosley, C.; Barwise, A.; Stetson, R.; Dong, Y.; Mavis, S.; Bellamkonda, V.; Colby, C.; Pickering, B. | 2022 | Critical care medicine | 50 | 1 SUPPL | 511- | 10.1097/01.ccm.0000810428.57493.2b | Wrong intervention (no online learning) |
| Protocol for a phase III pragmatic stepped wedge cluster randomised controlled trial comparing the effectiveness and cost-effectiveness of screening and guidelines with, versus without, implementation strategies for improving pain in adults with cancer at | Luckett, Tim; Phillips, Jane; Agar, Meera; Lam, Lawrence; Davidson, Patricia M.; McCaffrey, Nicola; Boyle, Frances; Shaw, Tim; Currow, David C.; Read, Alison; Hosie, Annmarie; Lovell, Melanie | 2018 | BMC health services research | 18 | 1 | 558 | 10.1186/s12913-018-3318-0 | No original data |
| Protocol for comparing two training approaches for primary care professionals implementing the Safe Environment for Every Kid (SEEK) model | Dubowitz, Howard; Saldana, Lisa; Magder, Laurence A.; Palinkas, Lawrence A.; Landsverk, John A.; Belanger, Rose L.; Nwosu, Ugonna S. | 2020 | Implementation science communications | 1 | | 78 | 10.1186/s43058-020-00059-9 | No original data |

| Title | Authors | Year | Journal | Volume | Issue | Pages | DOI | Exclusion Reason |
|---|---|---|---|---|---|---|---|---|
| Protocol for the Quick Clinical study: a randomised controlled trial to assess the impact of an online evidence retrieval system on decision-making in general practice | Coiera, Enrico; Magrabi, Farah; Westbrook, Johanna I.; Kidd, Michael R.; Day, Richard O. | 2006 | BMC medical informatics and decision making | 6 | | 33 | 10.1186/1472-6947-6-33 | No original data |
| RCT of Intensive, Brief, and Control Indigenous Cultural Safety Training Interventions for Health Care Providers | Nct, | 2023 | https://clinicaltrials.gov/show/NCT05890144 | | | | | Not healthcare clinical practice |
| REDIRECT: cluster randomised controlled trial of GP training in first-episode psychosis | Lester, Helen; Birchwood, Max; Freemantle, Nick; Michail, Maria; Tait, Lynda | 2009 | The British journal of general practice : the journal of the Royal College of General Practitioners | 59 | 563 | e183-e190 | 10.3399/bjgp09X420851 | Wrong intervention (no online learning) |
| Randomised trial of an internet-based evidence-based medicine continuing education intervention | Isrctn | 2006 | https://trialsearch.who.int/Trial2.aspx?TrialID=ISRCTN00946598 | | | | | No data in abstract, and no PDF available |
| Randomized Controlled Trial of an Intervention to Improve Nurses' Hazardous Drug Handling | Friese, Christopher R.; Yang, James; Mendelsohn-Victor, Kari; McCullagh, Marjorie | 2019 | Oncology nursing forum | 46 | 2 | 248-256 | 10.1188/19.ONF.248-256 | Comparison group online |
| Randomized controlled trial comparing four strategies for delivering e-curriculum to health care professionals [ISRCTN88148532] | Kemper, Kathi J.; Gardiner, Paula; Gobble, Jessica; Mitra, Ananda; Woods, Charles | 2006 | BMC medical education | 6 | | 2 | 10.1186/1472-6920-6-2 | Comparison group online |
| Randomized trial comparing Internet-based training in cognitive behavioural therapy theory, assessment and formulation to delayed-training control | Rakovshik, Sarah G.; McManus, Freda; Westbrook, David; Kholmogorova, Alla B.; Garanian, Natalya G.; Zvereva, Natalya V.; Ougrin, Dennis | 2013 | Behaviour research and therapy | 51 | 6 | 231-239 | 10.1016/j.brat.2013.01.009 | Not healthcare professionals |
| Randomized trial of HIV-assist versus guidelines for art selection by trainees | Ramirez, J.; Maddali, M.; Nematollahi, S.; Li, J. Z.; Shah, M. | 2020 | Topics in antiviral medicine | 28 | 1 | 424- | | Wrong intervention (no online learning) |
| Randomized trial of a web-based nurse education intervention to increase discussion of clinical trials | Margevicius, Seunghee; Daly, Barbara; Schluchter, Mark; Flocke, Susan; Manne, Sharon; Surdam, Jessica; Fulton, Sarah; Meropol, Neal J. | 2021 | Contemporary clinical trials communications | 22 | | 100789 | 10.1016/j.conctc.2021.100789 | Comparison group online |
| Randomized, controlled trial of the effectiveness of simulation education: A 24-month follow-up study in a clinical setting | Jansson, Miia M.; Syrjälä, Hannu P.; Ohtonen, Pasi P.; Meriläinen, Merja H.; Kyngäs, Helvi A.; Ala-Kokko, Tero I. | 2016 | American journal of infection control | 44 | 4 | 387-393 | 10.1016/j.ajic.2015.10.035 | Wrong intervention (no online learning) |

| Title | Authors | Year | Journal | Volume | Issue | Pages | DOI | Exclusion Reason |
|---|---|---|---|---|---|---|---|---|
| Rationale and evidence for emerging antibodydrug conjugates in gynecological cancers: Effect of online education on clinician knowledge | Fisher, G.; Furedy, A.; Vandenbroucque, J.; Concin, N. | 2020 | International Journal of Gynecological Cancer | 30 | SUPPL 4 | A10-A 11 | https://dx.doi.org/10.1136/ijgc-2020-ESGO.22 | Not randomised |
| Ready for SDM - The effect of a digital training module for clinicians supporting the implementation of shared decision-making | Isrctn, | 2023 | https://trialsearch.who.int/Trial2.aspx?TrialID=ISRCTN74230763 | | | | | No data in abstract, and no PDF available |
| Reducing the risk for cardiovascular events in diabetic dyslipidemia: Effect of online CME | Spyropoulos, J.; Boutsalis, G.; Shin, P. C.; LaCouture, M. | 2018 | Clinical Cardiology | 41 | Supplement 1 | 8 | https://dx.doi.org/10.1002/clc.23056 | Not randomised |
| Remotely Versus Locally Facilitated Simulation-based Training in Management of the Deteriorating Patient by Newly Graduated Health Professionals: A Controlled Trial | Christensen, Margrethe Duch; Rieger, Kathryn; Tan, Shane; Dieckmann, Peter; Østergaard, Doris; Watterson, Leonie M. | 2015 | Simulation in healthcare : journal of the Society for Simulation in Healthcare | 10 | 6 | 352-359 | 10.1097/SIH.0000000000000123 | Not randomised |
| Renal Transplantation Nurse Training on Patient-Centered Care Competence and Counseling Skills | Nct, | 2023 | https://clinicaltrials.gov/ct2/show/NCT06015984 | | | | | No data in abstract, and no PDF available |
| Results from the CLUES study: a cluster randomized trial for the evaluation of cardiovascular guideline implementation in primary care in Spain | Etxeberria, Arritxu; Alcorta, Idoia; Pérez, Itziar; Emparanza, Jose Ignacio; Ruiz de Velasco, Elena; Iglesias, Maria Teresa; Rotaeche, Rafael | 2018 | BMC health services research | 18 | 1 | 93 | 10.1186/s12913-018-2863-x | Wrong intervention (no online learning) |
| Rethinking Radiology: An Active Learning Curriculum for Head Computed Tomography Interpretation | Aliaga, Leonardo; Clarke, Samuel Owen | 2022 | The western journal of emergency medicine | 23 | 1 | 47-51 | 10.5811/westjem.2021.10.53665 | Comparison group online |
| Rhythm Control in Atrial Fibrillation: Online Cme Improves Cardiologists' Knowledge, Competence, and Confidence | Harris, M.; Amara, W.; LaCouture, M.; Guedj, P. | 2023 | Journal of the American College of Cardiology | 81 | 8 Supplement | 89 | https://dx.doi.org/10.1016/S0735-1097%2823%2900533-8 | Not enough data for analysis |
| Rhythm control strategy in recent atrial fibrillation: An online educational intervention improved physician knowledge, competence, and confidence | Harris, M.; Amara, W.; Chang, L. F.; Lacouture, M.; Guedj, P. | 2021 | Circulation | 144 | SUPPL 1 | | https://dx.doi.org/10.1161/circ.144.suppl_1.12074 | Not randomised |
| SEEK: dissemination and Implementation | Nct | 2018 | https://clinicaltrials.gov/show/NCT03642327 | | | | | No original data |
| SISTAQUIT (Supporting Indigenous Smokers to Assist Quitting) The SISTAQUIT trial compares usual care to health provider training in culturally-appropriate smoking cessation care for pregnant Aboriginal and/or Torres Strait Islander women | Actrn | 2018 | https://trialsearch.who.int/Trial2.aspx?TrialID=ACTRN12618000972224 | | | | | No original data |

| Title | Authors | Year | Journal | Volume | Issue | Pages | DOI | Exclusion Reason |
|---|---|---|---|---|---|---|---|---|
| SWOG 1904: Cluster-randomized controlled trial of patient and provider decision support to increase chemoprevention informed choice among women with atypical hyperplasia or lobular carcinoma in situ (MiCHOICE) | Crew, K. D.; Anderson, G.; Arnold, K.; Stieb, A.; Amenta, J. N.; Law, C.; Sandoval-Leon, A.; Colonna, S.; King, T.; Mangino, D.; Pruthi, S.; Perdekamp, M. G.; Braun-Inglis, C.; Krisher, S.; Yee, L.; Bertoni, D.; Seaward, S.; Wisinski, K. B.; Floyd, J.; Zarwan, C.; Ballinger, T. J.; VanderWalde, L.; Ross, M. M.; Steen, P.; Lo, S.; Conlin, A.; Yost, K.; Ellerton, J.; Lin, E.; Pederson, H. J.; Sardesai, S.; Jernigan, C.; Hershman, D.; Neuhouser, M. L.; Arun, B. K.; Kukafka, R. | 2023 | Cancer Research | 83 | 5 Supplement | | https://dx.doi.org/10.1158/1538-7445.SABCS22-OT1-15-01 | No data in abstract, and no PDF available |
| Safety quest: a novel web-based game to teach QI and patient safety | Shieh, L.; Evans, K.; Phadke, A.; Katznelson, L. | 2018 | Journal of hospital medicine | 13 | 4 | | | No data in abstract, and no PDF available |
| Secondary fracture prevention in home health care: initial results from a group randomized trial | Kilgore, M.; Outman, R.; Saag, K.; Locher, J.; Allison, J.; Curtis, J. | 2010 | Journal of bone and mineral research | 25 | | S209‐S210 | 10.1002/jbmr.5650251303 | Wrong intervention (no online learning) |
| Shared Decision Making: prostate Cancer Screening | Nct | 2005 | https://clinicaltrials.gov/show/NCT00207649 | | | | | No original data |
| Shared decision making, contextualized | Ferrer, Robert L.; Gill, James M. | 2013 | Annals of Family Medicine | 11 | 4 | 303-305 | 10.1370/afm.1551 | No original data |
| Simulated provider training improves diabetes management | Sperl-Hillen, J. A.; O'Connor, P.; Rush, W.; Ekstrom, H.; Appana, D.; Fernandes, O.; Asche, S. | 2013 | Journal of diabetes science and technology | 7 | 1 | A130 | | Not enough data for analysis |
| Simulated training improves provider knowledge and confidence in managing diabetes | Sperl-Hillen, J.; O'Connor, P.; Rush, W.; Asche, S.; Ekstrom, H.; Fernandes, O.; Rudge, A.; Appana, D.; Amundson, J.; Johnson, P. | 2012 | Diabetes | 61 | | A313‐A314 | 10.2337/db12-836-1328 | No knowledge, skills, or clinical outcome |
| Simulation Improves Procedural Protocol Adherence During Central Venous Catheter Placement: A Randomized Controlled Trial | Peltan, Ithan D.; Shiga, Takashi; Gordon, James A.; Currier, Paul F. | 2015 | Simulation in healthcare : journal of the Society for Simulation in Healthcare | 10 | 5 | 270-276 | 10.1097/SIH.0000000000000096 | Comparison group online |

| Title | Authors | Year | Journal | Volume | Issue | Pages | DOI | Exclusion Reason |
|---|---|---|---|---|---|---|---|---|
| Simulation Versus Interactive Mobile Learning for Teaching Extracorporeal Membrane Oxygenation to Clinicians: A Randomized Trial | Gannon, Whitney D.; Stokes, John W.; Pugh, Meredith E.; Bacchetta, Matthew; Benson, Clayne; Casey, Jonathan D.; Craig, Lynne; Semler, Matthew W.; Shah, Ashish S.; Troutt, Ashley; Rice, Todd W. | 2022 | Critical care medicine | 50 | 5 | e415-e425 | 10.1097/CCM.0000000000005376 | Wrong intervention (no online learning) |
| Simulation education as a single intervention does not improve hand hygiene practices: A randomized controlled follow-up study | Jansson, Miia M.; Syrjälä, Hannu P.; Ohtonen, Pasi P.; Meriläinen, Merja H.; Kyngäs, Helvi A.; Ala-Kokko, Tero I. | 2016 | American journal of infection control | 44 | 6 | 625-630 | 10.1016/j.ajic.2015.12.030 | Wrong intervention (no online learning) |
| Simulation in continuing education: Improving evidence-based decisions for rheumatoid arthritis management | Mehta, N.; Warters, M.; Blevins, D. | 2014 | Arthritis and Rheumatology | 66 | SUPPL. 10 | S873 | https://dx.doi.org/10.1002/art.38914 | Not randomised |
| Simulation training: Stressful but useful | Mahoney, L.; Fredette, M.; Bong, C.; Weinstock, P.; Lightdale, J. | 2009 | Journal of Pediatric Gastroenterology and Nutrition | 49 | SUPPL. 1 | E83 | https://dx.doi.org/10.1097/MPG.0b013e3181c64b4a | No knowledge, skills, or clinical outcome |
| Simulation-Based Education for Staff Managing Aggression and Externalizing Behaviors in Children With Autism Spectrum Disorder in the Hospital Setting: Pilot and Feasibility Study Protocol for a Cluster Randomized Controlled Trial | Mitchell, Marijke Jane; Newall, Fiona Helen; Sokol, Jennifer; Williams, Katrina Jane | 2020 | JMIR research protocols | 9 | 6 | e18105 | 10.2196/18105 | No data in abstract, and no PDF available |
| Simulation-based training for burr hole surgery and surgical instrument recognition | Clarke, D. B.; Hong, M.; Kureshi, N.; Fenerty, L.; Thibault-Halman, G.; D'Arcy, R. | 2017 | CMAJ. Canadian Medical Association Journal | 60 | 3 Supplement 2 | S18-S19 | https://dx.doi.org/10.1503/cjs.005617 | Comparison group online |
| Simulation-based training program effect on pediatric nurses' knowledge and performance regarding heel-prick during newborn blood screening test | Asiri, Abdulaziz; Almowafy, Abeer A.; Moursy, Shimaa M.; Abd-Elhay, Hanan A.; Ahmed, Shimaa Abdelrahim Khalaf; Abdelrahem, Aml S.; Seif, Marim T. Abo; Ahmed, Faransa A. | 2025 | BMC nursing | 24 | 1 | 110 | https://dx.doi.org/10.1186/s12912-024-02657-7 | Wrong intervention (no online learning) |
| Small talk big difference: results of a randomised controlled trial of an online educational intervention to improve the quality and rate of primary-care referrals to weight management programmes for patients with type 2 diabetes and obesity | Logue, J. | 2019 | Obesity facts | 12 | | 31- | 10.1159/000489691 | No original data |

| Title | Authors | Year | Journal | Volume | Issue | Pages | DOI | Exclusion Reason |
|---|---|---|---|---|---|---|---|---|
| Spaced Education to Optimize Prostate Cancer Screening | Nct | 2010 | https://clinicaltrials.gov/show/NCT01168323 | | | | | No original data |
| Speed and accuracy of a point of care web-based knowledge resource for clinicians: a controlled crossover trial | Cook, David A.; Enders, Felicity; Linderbaum, Jane A.; Zwart, Dale; Lloyd, Farrell J. | 2014 | Interactive journal of medical research | 3 | 1 | e7 | 10.2196/ijmr.2811 | Not randomised |
| Stereoscopic virtual reality does not improve knowledge acquisition of congenital heart disease | Patel, Neil; Costa, Anthony; Sanders, Stephen P.; Ezon, David | 2021 | The international journal of cardiovascular imaging | 37 | 7 | 2283-2290 | 10.1007/s10554-021-02191-6 | Comparison group online |
| Strengthening health visitors' breastfeeding support: Results from a cluster randomised study | Rossau, H. K.; Nilsson, I. M. S.; Gadeberg, A. K.; Forman, J. L.; Strandberg-Larsen, K.; Nielsen, J.; Villadsen, S. F. | 2024 | Nurse Education in Practice | 78 | | | 10.1016/j.nepr.2024.104033 | Comparison group online |
| Stroke Clinical Coding and Documentation Education Program | Actrn | 2022 | https://trialsearch.who.int/Trial2.aspx?TrialID=ACTRN12622000575730 | | | | | No original data |
| Study protocol for training providers in private practice in family-based treatment for adolescents with anorexia nervosa: A randomized controlled feasibility trial | Citron, Kyra; Johnson, Madelyn; Matheson, Brittany E.; Onipede, Z. Ayotola; Yang, Hyun-Joon; Bohon, Cara; Le Grange, Daniel; Lock, James | 2022 | Contemporary clinical trials | 120 | | 106889 | 10.1016/j.cct.2022.106889 | No original data |
| Study protocol: Randomized controlled trial of web-based decision support tools for high-risk women and healthcare providers to increase breast cancer chemoprevention | Crew, Katherine D.; Silverman, Thomas B.; Vanegas, Alejandro; Trivedi, Meghna S.; Dimond, Jill; Mata, Jennie; Sin, Margaret; Jones, Tarsha; Terry, Mary Beth; Tsai, Wei-Yann; Kukafka, Rita | 2019 | Contemporary clinical trials communications | 16 | | 100433 | 10.1016/j.conctc.2019.100433 | No data in abstract, and no PDF available |
| Success of online curriculum-based education in the management of multiple myeloma and continuing gaps among hematologists/oncologists | Van Laar, E. S.; Landgren, O. | 2015 | Blood | 126 | 23 | 3320 | | Not randomised |
| Suicide Prevention Training for PC Providers-in-training | Nct | 2016 | https://clinicaltrials.gov/show/NCT02996344 | | | | | No original data |

| Title | Authors | Year | Journal | Volume | Issue | Pages | DOI | Exclusion Reason |
|---|---|---|---|---|---|---|---|---|
| System-integrated technology-enabled model of care to improve the health of stroke patients in rural China: protocol for SINEMA—a cluster-randomized controlled trial | Gong, E.; Gu, W.; Sun, C.; Turner, E. L.; Zhou, Y.; Li, Z.; Bettger, J. P.; Oldenburg, B.; Amaya-Burns, A.; Wang, Y.; Xu, L. Q.; Yao, J.; Dong, D.; Xu, Z.; Li, C.; Hou, M.; Yan, L. L. | 2019 | American Heart Journal | 207 | | 27-39 | 10.1016/j.ahj.2018.08.015 | No knowledge, skills, or clinical outcome |
| TENSE-study | Ntr | 2012 | https://trialsearch.who.int/Trial2.aspx?TrialID=NTR3620 | | | | | Not healthcare clinical practice |
| Tailored educational intervention for primary care to improve the management of dementia: the EVIDEM-ED cluster randomized controlled trial | Wilcock, J.; Iliffe, S.; Griffin, M.; Jain, P.; Thune-Boyle, I.; Lefford, F.; Rapp, D. | 2013 | Trials | 14 | | 397 | 10.1186/1745-6215-14-397 | Wrong intervention (no online learning) |
| Tailored, online education on comparative effectiveness studies in rheumatoid arthritis: Success in improving knowledge and clinical decisions | Mehta, N.; McCardell, E.; Geissel, K. | 2015 | Arthritis and Rheumatology | 67 | SUPPL. 10 | | https://dx.doi.org/10.1002/art.39448 | Not randomised |
| Targeted interventions to prevent transitioning from acute to chronic low back pain in high-risk patients: development and delivery of a pragmatic training course of psychologically informed physical therapy for the TARGET trial | Beneciuk, Jason M.; George, Steven Z.; Greco, Carol M.; Schneider, Michael J.; Wegener, Stephen T.; Saper, Robert B.; Delitto, Anthony | 2019 | Trials | 20 | 1 | 256 | 10.1186/s13063-019-3350-3 | Not randomised |
| Teaching Important Basic EEG Patterns of Bedside Electroencephalography to Critical Care Staffs: A Prospective Multicenter Study | Legriel, S.; Jacq, G.; Lalloz, A.; Geri, G.; Mahaux, P.; Bruel, C.; Brochon, S.; Zuber, B.; Andre, C.; Dervin, K.; Holleville, M.; Cariou, A. | 2021 | Neurocritical Care | 34 | 1 | 144-153 | https://dx.doi.org/10.1007/s12028-020-01010-5 | Not healthcare professionals |
| Teaching Primary Care Genetics: A Randomized Controlled Trial Comparison | Telner, Deanna; Carroll, June C.; Regehr, Glenn; Tabak, Diana; Semotiuk, Kara; Freeman, Risa | 2017 | Family medicine | 49 | 6 | 443-450 | | Not enough data for analysis |
| Teaching breast cancer screening via text messages as part of continuing education for working nurses: a case-control study | Alipour, Sadaf; Jannat, Forouzandeh; Hosseini, Ladan | 2014 | Asian Pacific journal of cancer prevention : APJCP | 15 | 14 | 5607-5609 | 10.7314/apjcp.2014.15.14.5607 | Wrong intervention (no online learning) |
| Teaching the National Institutes of Health Stroke Scale to Paramedics (E-Learning vs Video): Randomized Controlled Trial | Koka, Avinash; Suppan, Laurent; Cottet, Philippe; Carrera, Emmanuel; Stuby, Loric; Suppan, Mélanie | 2020 | Journal of medical Internet research | 22 | 6 | e18358 | 10.2196/18358 | Comparison group online |

| Title | Authors | Year | Journal | Volume | Issue | Pages | DOI | Exclusion Reason |
|---|---|---|---|---|---|---|---|---|
| Technology-enabled academic detailing: computer-mediated education between pharmacists and physicians for evidence-based prescribing | Ho, Kendall; Nguyen, Anne; Jarvis-Selinger, Sandra; Novak Lauscher, Helen; Cressman, Céline; Zibrik, Lindsay | 2013 | International journal of medical informatics | 82 | 9 | 762-771 | 10.1016/j.ijmedinf.2013.04.011 | Not randomised |
| Teleconferenced educational detailing: diabetes education for primary care physicians | Harris, Stewart B.; Leiter, Lawrence A.; Webster-Bogaert, Susan; Van, Daphne M.; O'Neill, Colleen | 2005 | The Journal of continuing education in the health professions | 25 | 2 | 87-97 | 10.1002/chp.13 | Wrong intervention (no online learning) |
| Testing Test-Enhanced Continuing Medical Education: A Randomized Controlled Trial | Feldman, Mark; Fernando, Oshan; Wan, Michelle; Martimianakis, Maria Athina; Kulasegaram, Kulamakan | 2018 | Academic medicine : journal of the Association of American Medical Colleges | 93 | 11S Association of American Medical Colleges Learn Serve Lead: Proceedings of the 57th Annual Research in Medical Education Sessions | S30-S36 | 10.1097/ACM.0000000000002377 | Wrong intervention (no online learning) |
| Testing a Novel Deliberate Practice Intervention to Improve Diagnostic Reasoning in Trauma Triage: a Pilot Randomized Clinical Trial | Mohan, D.; Elmer, J.; Arnold, R. M.; Forsythe, R. M.; Fischhoff, B.; Rak, K.; Barnes, J. L.; White, D. B. | 2023 | JAMA network open | 6 | 5 | e2313569 | 10.1001/jamanetworkopen.2023.13569 | No knowledge, skills, or clinical outcome |
| The ADAPT Program to support the management of anxiety and depression in adult cancer patients: a cluster randomised trial to evaluate different implementation strategies | Actrn | 2017 | https://trialsearch.who.int/Trial2.aspx?TrialID=ACTRN12617000411347 | | | | | No knowledge, skills, or clinical outcome |
| The Additional Value of an E-Mail to Inform Healthcare Professionals of a Drug Safety Issue: A Randomized Controlled Trial in the Netherlands | Piening, Sigrid; de Graeff, Pieter A.; Straus, Sabine M. J. M.; Haaijer-Ruskamp, Flora M.; Mol, Peter G. M. | 2013 | Drug safety | 36 | 9 | 723-731 | 10.1007/s40264-013-0079-x | Wrong intervention (no online learning) |

| Title | Authors | Year | Journal | Volume | Issue | Pages | DOI | Exclusion Reason |
|---|---|---|---|---|---|---|---|---|
| The CKD-DETECT study: An RCT aimed at improving intention to initiate a kidney health check in Australian practice nurses | Sinclair, Peter M.; Kable, Ashly; Levett-Jones, Tracy; Holder, Carl; Oldmeadow, Christopher J. | 2019 | Journal of clinical nursing | 28 | 15-16 | 2745-2759 | 10.1111/jocn.14882 | Comparison group online |
| The Dash to Educate on Nash: Does Medical Education Improve Physician Knowledge? | Lubarda, J.; Smith, S.; Johnson, K. | 2019 | Gastroenterology | 156 | 6 Supplement 1 | S-1236 | https://dx.doi.org/10.1016/S0016-5085%2819%2940087-5 | Not randomised |
| The Effect of Education on Knowledge, attitude, and washing and sterilization Behavior of Surgery tools among Operating room staff | Irct2012080410488N | 2012 | https://trialsearch.who.int/Trial2.aspx?TrialID=IRCT2012080410488N1 | | | | | No data in abstract, and no PDF available |
| The Effect of Education on Nurses' Knowledge About High-flow Nasal Cannula Oxygen Therapy | Nct | 2022 | https://clinicaltrials.gov/show/NCT05362279 | | | | | No original data |
| The Effect of an Education Module to Reduce Weight Bias Among Healthcare Professionals in a Private Hospital Setting | Nct | 2021 | https://clinicaltrials.gov/show/NCT04741113 | | | | | Duplicate |
| The Effect of an Education Module to Reduce Weight Bias among Medical Centers Employees: A Randomized Controlled Trial | Sherf-Dagan, Shiri; Kessler, Yafit; Mardy-Tilbor, Limor; Raziel, Asnat; Sakran, Nasser; Boaz, Mona; Kaufman-Shriqui, Vered | 2022 | Obesity facts | 15 | 3 | 384-394 | 10.1159/000521856 | Not healthcare professionals |
| The Effectiveness of a Computer-Tailored E-Learning Program for Practice Nurses to Improve Their Adherence to Smoking Cessation Counseling Guidelines: Randomized Controlled Trial | de Ruijter, Dennis; Candel, Math; Smit, Eline Suzanne; de Vries, Hein; Hoving, Ciska | 2018 | Journal of medical Internet research | 20 | 5 | e193 | 10.2196/jmir.9276 | No knowledge, skills, or clinical outcome |
| The First Aid Skill Simulation Training of Nurses of General Wards | Nct | 2016 | https://clinicaltrials.gov/show/NCT02995785 | | | | | No original data |
| The Influence Of Online Continuing Medical Education On Disparities In Diagnosis And Treatment Of Heart Failure In Women | del Nido, E. L.; Turell, W.; Drexel, C.; Januzzi, J. L. | 2022 | Journal of Cardiac Failure | 28 | 5 Supplement | S33-S34 | https://dx.doi.org/10.1016/j.cardfail.2022.03.090 | Not randomised |
| The MacArthur Foundation Depression Education Program for Primary Care Physicians: background and rationale | Cole, S.; Raju, M.; Barrett, J.; Gerrity, M.; Dietrich, A. | 2000 | General hospital psychiatry | 22 | 5 | 299-358 | 10.1016/s0163-8343(00)80007-9 | No original data |
| The Quality in Acute Stroke Care (QASC) Australia Trial: national Translation of Fever, Sugar, Swallow (FeSS) Protocols | Actrn | 2022 | https://trialsearch.who.int/Trial2.aspx?TrialID=ACTRN12622000028707 | | | | | No original data |
| The Study Guide Cluster Randomized Control Trial | Nct | 2017 | https://clinicaltrials.gov/show/NCT03312218 | | | | | No original data |

| Title | Authors | Year | Journal | Volume | Issue | Pages | DOI | Exclusion Reason |
|---|---|---|---|---|---|---|---|---|
| The additional value of an e-mail to inform healthcare professionals of a drug safety issue | Piening, S.; De Graeff, P. A.; Straus, Smjm; Haaijer-Ruskamp, F. M.; Mol, P. G. M. | 2013 | Pharmacoepidemiology and drug safety | 22 | | 246- | 10.1002/pds.3512 | Wrong intervention (no online learning) |
| The computer-based drug and alcohol training assessment in Kenya | Clair, V.; Mutiso, V.; Musau, A.; Frank, E.; Ndetei, D. | 2017 | Drug and alcohol dependence | Conference: 2016 Annual Meeting of the College on Problems of Drug Dependence, CPDD 2016. United States. 171 | pp e42-e43 | | 10.1016/j.drugalcdep.2016.08.129 | No data in abstract, and no PDF available |
| The educational impact of web-based and face-to-face patient deterioration simulation programs: An interventional trial | Chung, Catherine; Cooper, Simon J.; Cant, Robyn P.; Connell, Cliff; McKay, Angela; Kinsman, Leigh; Gazula, Swapnali; Boyle, Jayne; Cameron, Amanda; Cash, Penny; Evans, Lisa; Kim, Jeong-Ah; Masud, Rana; McInnes, Denise; Norman, Lisa; Penz, Erika; Rotter, Thomas; Tanti, Erin; Breakspear, Tom | 2018 | Nurse education today | 64 | | 93-98 | 10.1016/j.nedt.2018.01.037 | Not randomised |
| The effect of a complementary e-learning course on implementation of a quality improvement project regarding care for elderly patients: a stepped wedge trial | Van de Steeg, Lotte; Langelaan, Maaike; Ijkema, Roelie; Wagner, Cordula | 2012 | Implementation science : IS | 7 | | 13 | 10.1186/1748-5908-7-13 | No original data |
| The effect of a multifaceted educational intervention on allied health clinicians' outcome measurement behaviours | Bowman, Julia A. | 2018 | | 75 | | | | Wrong intervention (no online learning) |

| Title | Authors | Year | Journal | Volume | Issue | Pages | DOI | Exclusion Reason |
|---|---|---|---|---|---|---|---|---|
| The effect of a simulation-based mastery learning intervention on pediatric interns procedural skills performance: a multicenter randomized trial | Kessler, D. O.; Arteaga, G.; Foltin, J.; Haubner, L.; Kamdar, G.; Krantz, A.; Lindower, J.; Miller, M.; O'Malley, S.; Petrescu, M.; Pusic, M. V.; Rocker, J.; Shah, N.; Strother, C.; Tilt, L.; Weinberg, E.; Auerbach, M. | 2010 | Pediatric Emergency Care | 26 | 9 | 703-704 | https://dx.doi.org/10.1097/PEC.0b013e3181f3469c | Wrong intervention (no online learning) |
| The effect of an educational video intervention on knowledge of obesity and weight bias in dietetic interns: A mixed methods analysis | Isom, Kellene A. | 2021 | | 82 | | | | Wrong intervention (no online learning) |
| The effect of educational intervention on the knowledge and attitude of intensive care nurses in the prevention of pressure ulcers | Karimian, Mohamad; Khalighi, Ebrahim; Salimi, Ebrahim; Borji, Milad; Tarjoman, Asma; Mahmoudi, Yosof | 2020 | The International journal of risk & safety in medicine | 31 | 2 | 89-95 | 10.3233/JRS-191038 | Wrong intervention (no online learning) |
| The effect of evidence-based nursing education on nurses' clinical decision making: A randomized controlled trial | Ghodsi Astan, Parisa; Goli, Rasoul; Hemmati Maslakpak, Masumeh; Rasouli, Javad; Alilu, Leyla | 2022 | Health science reports | 5 | 5 | e837 | 10.1002/hsr2.837 | Wrong intervention (no online learning) |
| The effect of the SAFE or SORRY? programme on patient safety knowledge of nurses in hospitals and nursing homes: a cluster randomised trial | van Gaal, Betsie G. I.; Schoonhoven, Lisette; Vloet, Lilian C. M.; Mintjes, Joke A. J.; Borm, George F.; Koopmans, Raymond T. C. M.; van Achterberg, Theo | 2010 | International journal of nursing studies | 47 | 9 | 1117-1125 | 10.1016/j.ijnurstu.2010.02.001 | Wrong intervention (no online learning) |
| The effectiveness of generic emails versus a remote knowledge broker to integrate mood management into a smoking cessation programme in team-based primary care: a cluster randomised trial | Minian, N.; Ahad, S.; Ivanova, A.; Veldhuizen, S.; Zawertailo, L.; Ravindran, A.; de Oliveira, C.; Baliunas, D.; Mulder, C.; Bolbocean, C.; et al. | 2021 | Implementation science | 16 | 1 | 30 | 10.1186/s13012-021-01091-6 | No knowledge, skills, or clinical outcome |
| The effectiveness of guideline implementation strategies on improving antipsychotic medication management for schizophrenia | Owen, Richard R.; Hudson, Teresa; Thrush, Carol; Thapa, Purushottam; Armitage, Tracey; Landes, Reid D. | 2008 | Medical care | 46 | 7 | 686-691 | 10.1097/MLR.0b013e3181653d43 | Wrong intervention (no online learning) |
| The effectiveness of the Ethics Quarter intervention on the ethical activity profile of nurse managers: A randomized controlled trial | Laukkanen, Laura; Suhonen, Riitta; Poikkeus, Tarja; Löyttyniemi, Eliisa; Leino-Kilpi, Helena | 2022 | Journal of nursing management | 30 | 7 | 2126-2137 | 10.1111/jonm.13411 | No knowledge, skills, or clinical outcome |

| Title | Authors | Year | Journal | Volume | Issue | Pages | DOI | Exclusion Reason |
|---|---|---|---|---|---|---|---|---|
| The effectiveness of video training in improving intensive care nurses' knowledge about brain death identification | Deniz, İsmail; Ayhan, Hatice | 2022 | Nursing in critical care | | | | 10.1111/nicc.12863 | Wrong intervention (no online learning) |
| The effectiveness of video training in improving intensive care nurses' knowledge about brain death identification | Deniz, I.; Ayhan, H. | 2024 | Nursing in critical care | 29 | 1 | 80‐89 | 10.1111/nicc.12863 | Wrong intervention (no online learning) |
| The effects of education about depression in primary care | Thompson, C. | 1999 | | | | | | Wrong intervention (no online learning) |
| The effects of various instructional methods on retention of knowledge about pressure ulcers among critical care and medical-surgical nurses | Cox, Jill; Roche, Sharon; Van Wynen, Elizabeth | 2011 | Journal of continuing education in nursing | 42 | 2 | 71-78 | 10.3928/00220124-20100802-03 | Wrong intervention (no online learning) |
| The evolving role of parp inhibitors in newly diagnosed advanced ovarian cancer: The effect of online education on clinician knowledge, competence and confidence | Fisher, G.; Furedy, A.; Vandenbroucque, J.; Monk, B. | 2020 | International Journal of Gynecological Cancer | 30 | SUPPL 4 | A65-A 66 | https://dx.doi.org/10.1136/ijgc-2020-ESGO.117 | Not randomised |
| The impact of Professional Boundaries for Health Professionals (PBHP) training on knowledge, comfort, experience, and ethical decision-making: a longitudinal randomized controlled trial | Fronek, Patricia; Kendall, Melissa B. | 2017 | Disability and rehabilitation | 39 | 24 | 2522-2 529 | 10.1080/09638288.2016.1236152 | Wrong intervention (no online learning) |
| The impact of Stress Management and Resailience Training (SMART) on academic physicians during the implementation of a new Health Information System: An exploratory randomized controlled trial | Spilg, Edward G.; Kuk, Hanna; Ananny, Lesley; McNeill, Kylie; LeBlanc, Vicki; Bauer, Brent A.; Sood, Amit; Wells, Philip S. | 2022 | PloS one | 17 | 4 | e02672 40 | 10.1371/journal.pone.0267240 | Not healthcare clinical practice |
| The impact of a training programme incorporating the conceptual framework of the International Classification of Functioning (ICF) on behaviour regarding interprofessional practice in Rwandan health professionals: A cluster randomized control trial | Sagahutu, Jean Baptiste; Kagwiza, Jeanne; Cilliers, Francois; Jelsma, Jennifer | 2020 | PloS one | 15 | 2 | e02262 47 | 10.1371/journal.pone.0226247 | Wrong intervention (no online learning) |
| The impact of an educational program on nurses' shared decision making attitudes: A randomized controlled trial | Hsu, Hsiu-Chin; Lin, Mei-Hsiang | 2022 | Applied nursing research : ANR | 65 | | 151587 | 10.1016/j.apnr.2022.151587 | Wrong intervention (no online learning) |
| The impact of delirium education through e-learning on outcomes in patients and nurses: an intervention trial | Isrctn | 2017 | https://trialsearch.who.int/Trial2.aspx?TrialID=ISRCTN82293702 | | | | | Not randomised |
| The impact of educational interventions on primary health care workers' knowledge of occupational exposure to blood or body fluids | Krishnan, Prassana; Dick, Finlay; Murphy, Elizabeth | 2007 | Occupational medicine (Oxford, England) | 57 | 2 | 98-103 | 10.1093/occmed/kql126 | Wrong intervention (no online learning) |

| Title | Authors | Year | Journal | Volume | Issue | Pages | DOI | Exclusion Reason |
|---|---|---|---|---|---|---|---|---|
| The implementation and effectiveness of Brief Physical Activity Counselling by Physiotherapists (BEHAVIOUR) on levels of patient physical activity | Actrn | 2021 | https://trialsearch.who.int/Trial2.aspx?TrialID=ACTRN12621000194864 | | | | | No original data |
| The importance of involving healthcare professionals in the production of neurodiversity healthcare training | French, B. | 2023 | European Psychiatry | 66 | Supplement 1 | S19 | https://dx.doi.org/10.1192/j.eurpsy.2023.77 | Not randomised |
| The improving care in chronic obstructive lung disease study: CAROL improving processes of care and quality of life of COPD patients in primary care: study protocol for a randomized controlled trial | Steurer-Stey, Claudia; Markun, Stefan; Lana, Kaba Dalla; Frei, Anja; Held, Ulrike; Wensing, Michel; Rosemann, Thomas | 2014 | Trials | 15 | | 96 | 10.1186/1745-6215-15-96 | Wrong intervention (no online learning) |
| The mCME Project: A Randomized Controlled Trial of an SMS-Based Continuing Medical Education Intervention for Improving Medical Knowledge among Vietnamese Community Based Physicians' Assistants | Gill, Christopher J.; Le Ngoc, Bao; Halim, Nafisa; Nguyen Viet, Ha; Larson Williams, Anna; Nguyen Van, Tan; McNabb, Marion; Tran Thi Ngoc, Lien; Falconer, Ariel; An Phan Ha, Hai; Rohr, Julia; Hoang, Hai; Michiel, James; Nguyen Thi Thanh, Tam; Bird, Liat; Pham Vu, Hoang; Yeshitla, Mahlet; Ha Van, Nhu; Sabin, Lora | 2016 | PloS one | 11 | 11 | e0166293 | 10.1371/journal.pone.0166293 | Wrong intervention (no online learning) |
| The potential counter effect of COVID-19 outbreak on an antimicrobial agents prescribing educational intervention | Yasein, Nada; Shroukh, Wejdan; Barghouti, Farihan; Hassanin, Omayma; Yousef, Hala; AlSmairat, Maram; Al Hiary, Ghadeer; AlFayoumi, Farah | 2021 | Journal of infection in developing countries | 15 | 11 | 1653-1660 | 10.3855/jidc.15213 | Wrong intervention (no online learning) |
| The practical use of the latest standards of electrocardiography (PULSE) trial: nursing-focused intervention improves nurses' knowledge and quality of ECG monitoring | Funk, M.; Fennie, K. P.; Stephens, K.; May, J. L.; Winkler, C.; Chang, P. S.; Feder, S.; Knudson, K.; Bovino, L. R.; Turkman, Y.; et al. | 2014 | Circulation | 130 | | | | Duplicate |
| The praecox program: evaluation of an online educational program to improve neonatal palliative care practice | Kain, V. J.; Pritchard, M. A.; Yates, P.; Curtis, M.; Fraser, R. | 2016 | Journal of paediatrics and child health | 52 | | 30‐ | 10.1111/jpc.13194 | Not enough data for analysis |

| Title | Authors | Year | Journal | Volume | Issue | Pages | DOI | Exclusion Reason |
|---|---|---|---|---|---|---|---|---|
| The role of online resources to support patients and health professionals, how well used or useful are they? | Shaw, J. M.; Shepherd, H. L.; Butow, P.; Davies, F.; Cuddy, J.; Harris, M.; Kirsten, L.; Kelly, B. | 2022 | Asia-Pacific Journal of Clinical Oncology | 18 | Supplement 3 | 76 | https://dx.doi.org/10.1111/ajco.13868 | Duplicate |
| The study design and rationale of the randomized controlled trial: translating COPD guidelines into primary care practice | Parker, Donna R.; Eaton, Charles B.; Ahern, David K.; Roberts, Mary B.; Rafferty, Caitlin; Goldman, Roberta E.; McCool, F. Dennis; Wroblewski, Joseph | 2013 | BMC family practice | 14 | | 56 | 10.1186/1471-2296-14-56 | No original data |
| The virtual rheumatology clinic: Virtual patients for resident education in rheumatology | Marston, B. A.; Siegel, D.; Anandarajah, A. P.; Lang, V. | 2016 | Arthritis and Rheumatology | 68 | Supplement 10 | 1466-1467 | https://dx.doi.org/10.1002/art.39977 | Not randomised |
| Traditional Versus Simulation Resident Surgical Laparoscopic Salpingectomy Training: A Randomized Controlled Trial | Patel, Nima R.; Makai, Gretchen E.; Sloan, Nancy L.; Della Badia, Carl R. | 2016 | Journal of minimally invasive gynecology | 23 | 3 | 372-377 | 10.1016/j.jmig.2015.11.005 | Wrong intervention (no online learning) |
| Training family physicians and primary care nurses improves diagnostic assessment of dementia: results of a randomized controlled trial | Perry, M. | 2011 | Alzheimer's & dementia | 7 | 4 | S498‐ | 10.1016/j.jalz.2011.05.2393 | Wrong intervention (no online learning) |
| Training for occupational health physicians to involve significant others in the return-to-work process of workers with chronic diseases: a randomized controlled trial | Snippen, Nicole C.; de Vries, Haitze J.; Hagedoorn, Mariët; Brouwer, Sandra | 2022 | Disability and rehabilitation | | | 1-Nov | 10.1080/09638288.2022.2107091 | Not healthcare clinical practice |
| Training pediatric fellows in palliative care: a comparison of simulation-based training and didactic education | Brock, K.; Cohen, H.; Sourkes, B.; Good, J.; Halamek, L. | 2017 | Pediatric blood and cancer. Conference: 30th annual meeting of the american society of pediatric hematology/oncology, ASPHO 2017. Canada | 64 | | S65‐S66 | 10.1002/pbc.26591 | Not randomised |
| Training pediatric health care providers in prevention of dental decay: results from a randomized controlled trial | Slade, Gary D.; Rozier, R. Gary; Zeldin, Leslie P.; Margolis, Peter A. | 2007 | BMC health services research | 7 | | 176 | 10.1186/1472-6963-7-176 | Wrong intervention (no online learning) |
| Training providers in nighttime postural care intervention: a randomized control trial | Hutson, J. | 2019 | Developmental medicine and child neurology | 61 | | 93‐ | 10.1111/dmcn.14353 | Not healthcare professionals |
| Translating The GOLD COPD Guidelines Into Primary Care Practice | Nct | 2010 | https://clinicaltrials.gov/show/NCT01237561 | | | | | No data in abstract, and no PDF available |

| Title | Authors | Year | Journal | Volume | Issue | Pages | DOI | Exclusion Reason |
|---|---|---|---|---|---|---|---|---|
| Translating research into practice: Protocol for a community-engaged, stepped wedge randomized trial to reduce disparities in breast cancer treatment through a regional patient navigation collaborative | Battaglia, Tracy A.; Freund, Karen M.; Haas, Jennifer S.; Casanova, Nicole; Bak, Sharon; Cabral, Howard; Freedman, Rachel A.; White, Karen Burns; Lemon, Stephenie C. | 2020 | Contemporary clinical trials | 93 | | 106007 | 10.1016/j.cct.2020.106007 | No original data |
| Type 2 diabetes quality improvement CME: Impact on physician knowledge | Larkin, A.; Healy, C.; Le, A. | 2016 | Endocrine Reviews | 37 | 2 Supplement 1 | | https://dx.doi.org/10.1210/endo-meetings.2016.EHDE.1.SUN-757 | Not randomised |
| Understanding obesity and weight loss interventions: Effect of online education on physician knowledge and confidence | Trier, J.; Kushner, R. F.; McCarthy, R. | 2018 | Obesity Facts | 11 | Supplement 1 | 124-125 | https://dx.doi.org/10.1159/000489691 | Not randomised |
| Use of Resources and Method of Proctoring During the NBCRNA Continued Professional Certification Assessment: Analysis of Outcomes | Spence, D.; Ward, R.; Wooden, S.; Browne, M.; Song, H.; Hawkins, R.; Wojnakowski, M. | 2019 | Journal of Nursing Regulation | 10 | 3 | 37-46 | 10.1016/S2155-8256(19)30147-4 | Wrong intervention (no online learning) |
| Use of an algorithm-based electronic application in the management of penicillin allergies | Vijayaraghavan, N.; Otome, O. | 2023 | Open Forum Infectious Diseases | 10 | | S573 | 10.1093/ofid/ofad500.1113 | Wrong intervention (no online learning) |
| Use of an internet-based instruction module in teaching complex clinical subject matter to physicians-in-training | Stasek, J. E.; Way, D. P.; Hurtubise, L.; Nagel, R.; Hasbrouck, C.; Hudson, A. | 2010 | American journal of respiratory and critical care medicine | 181 | 1 Meeting Abstracts | | | Not healthcare professionals |
| Use of the Smartphone App WhatsApp as an E-Learning Method for Medical Residents: Multicenter Controlled Randomized Trial | Clavier, Thomas; Ramen, Julie; Dureuil, Bertrand; Veber, Benoit; Hanouz, Jean-Luc; Dupont, Hervé; Lebuffe, Gilles; Besnier, Emmanuel; Compere, Vincent | 2019 | JMIR mHealth and uHealth | 7 | 4 | e12825 | 10.2196/12825 | Comparison group online |
| Using Effective Provider-Patient Communication to Improve Cancer Screening Among Low Literacy Patients | Nct | 2011 | https://clinicaltrials.gov/show/NCT01361035 | | | | | Duplicate |

| Title | Authors | Year | Journal | Volume | Issue | Pages | DOI | Exclusion Reason |
|---|---|---|---|---|---|---|---|---|
| Using Mobile Virtual Reality Simulation to Prepare for In-Person Helping Babies Breathe Training: Secondary Analysis of a Randomized Controlled Trial (the eHBB/mHBS Trial) | Ezenwa, Beatrice Nkolika; Umoren, Rachel; Fajolu, Iretiola Bamikeolu; Hippe, Daniel S.; Bucher, Sherri; Purkayastha, Saptarshi; Okwako, Felicitas; Esamai, Fabian; Feltner, John B.; Olawuyi, Olubukola; Mmboga, Annet; Nafula, Mary Concepta; Paton, Chris; Ezeaka, Veronica Chinyere | 2022 | JMIR medical education | 8 | 3 | e37297 | 10.2196/37297 | Wrong intervention (no online learning) |
| Using a participatory action research approach to facilitate the implementation of the IADL profile in clinical practice | Bottari, C.; Kairy, D.; Shun, P. L. W.; Ouellet, C.; Magnan, C.; Poissant, L.; Dawson, D.; Swaine, B. | 2016 | Brain injury. Conference: 11th world congress on brain injury of the international brain injury association. Netherlands. Conference start: 20160302. Conference end: 20160305 | 30 | 5-6 | 593 | 10.3109/02699052.2016.1162060 | Not randomised |
| Using a theory-based, customized video game as an educational tool to improve physicians' trauma triage decisions: study protocol for a randomized cluster trial | Mohan, D.; Angus, D. C.; Chang, C. H.; Elmer, J.; Fischhoff, B.; Rak, K. J.; Barnes, J. L.; Peitzman, A. B.; White, D. B. | 2024 | Trials | 25 | 1 | 127 | 10.1186/s13063-024-07961-w | No data in abstract, and no PDF available |
| Using an Analysis of Behavior Change to Inform Effective Digital Intervention Design: How Did the PRIMIT Website Change Hand Hygiene Behavior Across 8993 Users? | Ainsworth, B.; Steele, M.; Stuart, B.; Joseph, J.; Miller, S.; Morrison, L.; Little, P.; Yardley, L. | 2017 | Annals of behavioral medicine : a publication of the Society of Behavioral Medicine | 51 | 3 | 423-431 | 10.1007/s12160-016-9866-9 | Not healthcare professionals |
| Using evidence-integrated e-learning to enhance case management continuing education for psychiatric nurses: a randomised controlled trial with follow-up | Liu, Wen- I.; Rong, Jiin-Ru; Liu, Chieh-Yu | 2014 | Nurse education today | 34 | 11 | 1361-1367 | 10.1016/j.nedt.2014.03.004 | Wrong intervention (no online learning) |
| Using goutpro to make medical trainees gout pros-a single blinded randomized control study | Ngo, L.; Miller, E.; Valen, P. A.; Duran, A. | 2017 | Arthritis & rheumatology | 69 | | | | Wrong intervention (no online learning) |
| Using people with aphasia to train health professionals in effective communication strategies over the internet | Actrn | 2016 | http://www.who.int/trialsearch/Trial2.aspx?TrialID=ACTRN12616000062426 | | | | | No original data |
| Using technology to improve access, quality and outcomes: tobacco treatment in mental health settings | Brunette, M. F.; Dzebisashvili, N.; Xie, H.; Akerman, S.; Pratt, S.; Ferron, J. C.; Bartels, S. | 2015 | Journal of mental health policy and economics | 18 | | S4-S5 | | No data in abstract, and no PDF available |

| Title | Authors | Year | Journal | Volume | Issue | Pages | DOI | Exclusion Reason |
|---|---|---|---|---|---|---|---|---|
| Using virtual patient simulations to prepare primary health care professionals to conduct substance use and mental health screening and brief intervention | Albright, Glenn; Bryan, Craig; Adam, Cyrille; McMillan, Jeremiah; Shockley, Kristen | 2018 | Journal of the American Psychiatric Nurses Association | 24 | 3 | 247-259 | 10.1177/1078390317719321 | Not enough data for analysis |
| VHA Clinicians and Bioterror Events: interactive Web-based Learning | Nct | 2005 | https://clinicaltrials.gov/show/NCT00123396 | | | | | Duplicate |
| Validation of an affordable and accessible alternative simulation technology | Auguste, T. C.; Benda, N. C.; Kellogg, K.; Fairbanks, R. | 2017 | Obstetrics and Gynecology | 129 | Supplement 1 | 43S | | Wrong intervention (no online learning) |
| Validation of an on-line independent training program for TOR-BSST© dysphagia screeners | Darling, S.; MaeBrae-Waller, A.; Casaubon, L. K.; Reichardt, B. | 2023 | Neurorehabilitation and neural repair | 37 | 5 | 47S | 10.1177/15459683231163223 | Comparison group online |
| Vermont Diabetes Information System | Nct | 2005 | https://clinicaltrials.gov/show/NCT00109369 | | | | | Wrong intervention (no online learning) |
| Video instruction increases accuracy in performance of nurse dysphagia screening | Reed, C.; Fraley, D.; Sharp, J.; Hernandez, A.; Proulx, J.; Kattan, A. C.; Williams, H.; Wilson, R. L. | 2013 | Stroke | 44 | 2 | | | Not randomised |
| Video strategies improved health professional knowledge across different contexts: a helix counterbalanced randomized controlled study | Sarkies, Mitchell N.; Maloney, Stephen; Symmons, Mark; Haines, Terry P. | 2019 | Journal of clinical epidemiology | 112 | | 1-Nov | 10.1016/j.jclinepi.2019.04.003 | Intervention <10 mins |
| Video-based learning versus traditional lecture-based learning for osteoporosis education: a randomized controlled trial | Chotiyarnwong, Pojchong; Boonnasa, Wararat; Chotiyarnwong, Chayaporn; Unnanuntana, Aasis | 2021 | Aging clinical and experimental research | 33 | 1 | 125-131 | 10.1007/s40520-020-01514-2 | Not healthcare professionals |
| Videotape-based decision aid for colon cancer screening. A randomized, controlled trial | Pignone, M.; Harris, R.; Kinsinger, L. | 2000 | Annals of internal medicine | 133 | 10 | 761-769 | 10.7326/0003-4819-133-10-200011210-00008 | Not healthcare professionals |
| Virtual Reality for Patient Education about Hypertension: A Randomized Pilot Study | Jiravska Godula, B.; Jiravsky, O.; Matheislova, G.; Kuriskova, V.; Valkova, A.; Puskasova, K.; Dokoupil, M.; Dvorakova, V.; Prifti, A.; Foral, D.; Jiravsky, F.; Hecko, J.; Hudec, M.; Neuwirth, R.; Miklik, R. | 2023 | Journal of Cardiovascular Development and Disease | 10 | 12 | 481 | https://dx.doi.org/10.3390/jcdd10120481 | Not healthcare professionals |
| Virtual Worlds Technology to Enhance Training for Primary Care Providers in Assessment and Management of Posttraumatic Stress Disorder Using Motivational Interviewing: pilot Randomized Controlled Trial | Manuel, J. K.; Purcell, N.; Abadjian, L.; Cardoos, S.; Yalch, M.; Hill, C.; McCarthy, B.; Bertenthal, D.; McGrath, S.; Seal, K. | 2023 | JMIR medical education | 9 | | e42862 | 10.2196/42862 | Comparison group online |

| Title | Authors | Year | Journal | Volume | Issue | Pages | DOI | Exclusion Reason |
|---|---|---|---|---|---|---|---|---|
| Virtual diabetes education improves resident physician knowledge and performance: a cluster-randomized trial | Sperl-Hillen, J.; O'Connor, P. J.; Ekstrom, H.; Asche, S.; Appana, D.; Johnson, P. E. | 2013 | Diabetes | 62 | | A172-A173 | 10.2337/db13-680-858 | Duplicate |
| Virtual patient simulation to improve nurses' relational skills in a continuing education context: a convergent mixed methods study | Rouleau, G.; Gagnon, M. P.; Côté, J.; Richard, L.; Chicoine, G.; Pelletier, J. | 2022 | BMC Nursing | 21 | 1 | | 10.1186/s12912-021-00740-x | Not randomised |
| Virtual reality simulation for critical pediatric airway management training | Putnam, Elizabeth M.; Rochlen, Lauryn R.; Alderink, Erik; Augé, James; Popov, Vitaliy; Levine, Robert; Tait, Alan R. | 2021 | Journal of clinical and translational research | 7 | 1 | 93-99 | | Comparison group online |
| Virtual reality skills training for health care professionals in alcohol screening and brief intervention | Fleming, Michael; Olsen, Dale; Stathes, Hilary; Boteler, Laura; Grossberg, Paul; Pfeifer, Judie; Schiro, Stephanie; Banning, Jane; Skochelak, Susan | 2009 | Journal of the American Board of Family Medicine : JABFM | 22 | 4 | 387-398 | 10.3122/jabfm.2009.04.080208 | Not healthcare professionals |
| Web application for assisting the management of patients with common cutaneous adverse drug reactions: a multi-center randomized controlled trial | Tctr, | 2023 | https://trialsearch.who.int/Trial2.aspx?TrialID=TCTR20230119006 | | | | | No data in abstract, and no PDF available |
| Web-Based Just-in-Time Information and Feedback on Antibiotic Use for Village Doctors in Rural Anhui, China: Randomized Controlled Trial | Shen, XingRong; Lu, Manman; Feng, Rui; Cheng, Jing; Chai, Jing; Xie, Maomao; Dong, Xuemeng; Jiang, Tao; Wang, Debin | 2018 | Journal of medical Internet research | 20 | 2 | e53 | 10.2196/jmir.8922 | Wrong intervention (no online learning) |
| Web-Based Learning in Residents' Continuity Clinics: A Randomized, Controlled Trial | Cook, David A.; Dupras, Denise M.; Thompson, Warren G.; Pankratz, V. Shane | 2005 | Academic Medicine | 80 | 1 | 90-97 | 10.1097/00001888-200501000-00022 | Comparison group online |
| Web-Based Respiratory Education About Tobacco and Health Phase II | Nct | 2012 | https://clinicaltrials.gov/show/NCT01642264 | | | | | Not enough data for analysis |
| Web-Based Tailored Educational Program in Improving Nurse Communication With Patients About Clinical Trials | Nct | 2014 | https://clinicaltrials.gov/show/NCT02129517 | | | | | No original data |
| Web-Based Training for Nurses on Shared Decision Making and Prenatal Screening for Down Syndrome: Protocol for a Randomized Controlled Trial | Poulin Herron, Alex; Agbadje, Titilayo Tatiana; Cote, Melissa; Djade, Codjo Djignefa; Roch, Geneviève; Rousseau, Francois; Légaré, France | 2020 | JMIR research protocols | 9 | 10 | e17878 | 10.2196/17878 | No original data |

| Title | Authors | Year | Journal | Volume | Issue | Pages | DOI | Exclusion Reason |
|---|---|---|---|---|---|---|---|---|
| Web-based Clinical Pedagogy Program to Enhance Nurse Preceptors' Teaching Competency | Nct | 2018 | https://clinicaltrials.gov/show/NCT03456297 | | | | | No original data |
| Web-based intervention modelling experiments: A way of exploring professional behaviour change interventions before a full-scale trial | Treweek, S.; Bonetti, D.; Barnett, K.; Eccles, M.; Francis, J.; Jones, C.; MacLennan, G.; Pitts, N.; Ricketts, I.; Sullivan, F.; Weal, M. | 2013 | Clinical Trials | 10 | SUPPL. 2 | S42-S43 | https://dx.doi.org/10.1177/1740774513497438 | Not randomised |
| Web-based training for primary care providers on screening, brief intervention, and referral to treatment (SBIRT) for alcohol, tobacco, and other drugs | Stoner, Susan A.; Mikko, A. Tasha; Carpenter, Kelly M. | 2014 | Journal of substance abuse treatment | 47 | 5 | 362-370 | 10.1016/j.jsat.2014.06.009 | Not enough data for analysis |
| What does recent data related to SGLT2 inhibitors mean for my patients? Effect of online medical education on physician knowledge and competence | Larkin, A.; LaCouture, M.; Le, A. | 2017 | Endocrine Reviews | 38 | 3 Supplement 1 | | | Not randomised |
| Young Hearts, Strong Starts | Nct | 2013 | https://clinicaltrials.gov/show/NCT01893593 | | | | | No original data |
| [Multimodal training of general practitioners--evaluation and knowledge increase within the framework of the dementia management initiative in general medicine (IDA)] | Vollmar, Horst Christian; Grässel, Elmar; Lauterberg, Jörg; Neubauer, Simone; Grossfeld-Schmitz, Maria; Koneczny, Nik; Schürer-Maly, Cornelia-Christine; Koch, Mitra; Ehlert, Norman; Holle, Rolf; Rieger, Monika A.; Butzlaff, Martin | 2007 | Zeitschrift fur arztliche Fortbildung und Qualitatssicherung | 101 | 1 | 27-34 | 10.1016/j.zgesun.2006.12.005 | Wrong intervention (no online learning) |
| [The effect of a scenario-based simulation communication course on improving the communication skills of nurses] | Huang, Ya-Hsuan; Hsieh, Suh-Ing; Hsu, Li-Ling | 2014 | Hu li za zhi The journal of nursing | 61 | 2 | 33-43 | 10.6224/JN.61.2.33 | Wrong intervention (no online learning) |
| a Randomized Controlled Trial Evaluating the Impact of an e-Learning Intervention Based on the Improving Well-Being and Health for People with Dementia (Wheld) Person-Centred Care Training Programme | McDermid, J.; Fossey, J.; Da Silva, M. V.; Ballard, C. | 2018 | Alzheimer's & dementia | 14 | 7 | P1406- | 10.1016/j.jalz.2018.06.2926 | Comparison group online |
| eLearning course may shorten the duration of mechanical restraint among psychiatric inpatients: a cluster-randomized trial | Kontio, Raija; Pitkänen, Anneli; Joffe, Grigori; Katajisto, Jouko; Välimäki, Maritta | 2014 | Nordic journal of psychiatry | 68 | 7 | 443-449 | 10.3109/08039488.2013.855254 | Not healthcare clinical practice |

| Title | Authors | Year | Journal | Volume | Issue | Pages | DOI | Exclusion Reason |
|---|---|---|---|---|---|---|---|---|
| eLearning improves allied health professionals' knowledge and confidence to manage medically unexplained chronic fatigue states: a randomized controlled trial | Jones, M. D.; Casson, S. M.; Barry, B. K.; Li, S. H.; Valenzuela, T.; Cassar, J.; Lamanna, C.; Lloyd, A. R.; Sandler, C. X. | 2023 | Journal of psychosomatic research | 173 | | 111462 | 10.1016/j.jpsychores.2023.111462 | Not enough data for analysis |
| mCME project V.2.0: randomised controlled trial of a revised SMS-based continuing medical education intervention among HIV clinicians in Vietnam | Gill, Christopher J.; Le, Ngoc Bao; Halim, Nafisa; Chi, Cao Thi Hue; Nguyen, Viet Ha; Bonawitz, Rachael; Hoang, Pham Vu; Nguyen, Hoang Long; Huong, Phan Thi Thu; Larson Williams, Anna; Le, Ngoc Anh; Sabin, Lora | 2018 | BMJ global health | 3 | 1 | e000632 | 10.1136/bmjgh-2017-000632 | Comparison group online |
