## Supplementary Files 1-7 for "Effects of online professional learning on healthcare professionals’ knowledge and skill acquisition: A systematic review and meta-analysis": Supplementary File 2_Inclusion Criteria.pdf

**Detailed Inclusion Criteria**

Studies were eligible for inclusion if they met the following criteria:

1) *Design*: Randomised controlled trials where healthcare professionals were assigned to online learning versus any non-online comparison (e.g., waitlist control, active control, or face-to-face learning). Quasi-experimental and non-randomized trials were excluded due to the higher risk of confounding [1].
2) *Participants*: Any qualified healthcare professionals (e.g., doctors, nurses, midwives, dentists, allied health professionals, psychologists, psychiatrists, pharmacists) working in a clinical or community healthcare setting. We excluded university students as they were not considered practising professionals.
3) *Intervention*: Any form of online professional learning. Aligned with previous research [2,3], we defined online learning as educational interventions with online multimedia elements (e.g., videos, video-conferencing) or active learning components (e.g., interactive websites, quizzes). We excluded educational materials that did not contain any active learning components, such as books, journal articles, static PDF documents, standalone clinical decision-making tools without training, and websites without learning support (e.g., websites for academic journals). Virtual Reality (VR) interventions were generally excluded, except when used as a training simulator in conjunction with an online learning module [e.g., 4]. We judged VR to be distinct from most other online learning, with different costs, benefits, and user experiences [5–7]. To ensure the interventions were substantive educational experiences rather than shorter online messaging (e.g., TikTok videos), the review included only interventions lasting 10

minutes or longer. This duration threshold was established to differentiate between brief ‘nudges’ from educational interventions designed to promote lasting behaviour change.

4) *Comparison*: We included studies that compared interventions against any other form of professional learning (i.e., workshops, lectures, simulations, handouts, textbook guidelines) or control (i.e., waitlist or no intervention).

5) *Outcome*: We included studies that reported measures of knowledge or skills relevant to patient care [8], assessed through standardised measures such as examination results, quiz performance, or skill demonstration in simulated settings. Studies that only included self-reported measures of knowledge, skill or professional confidence (e.g., participants' perceptions of their own abilities) were excluded, as were measures only indirectly related to patient care (e.g., fire safety training).
