## Supplementary Files 1-7 for "Effects of online professional learning on healthcare professionals’ knowledge and skill acquisition: A systematic review and meta-analysis": Supplementary File 3_Full Search Strategy.pdf

**Full Search Strategy**

***MEDLINE Complete***

( ( ( ( ABS ( nurs* OR "healthcare professional*" OR "health professional*" OR physician* OR "healthcare provider" OR "consultant*" OR clinician* OR "allied health" OR "medical staff" OR "primary care*" ) ) OR ( TITLE ( nurs* OR "healthcare professional*" OR "health professional*" OR physician* OR "healthcare provider" OR clinician* OR "allied health" OR "medical staff" OR "consultant*" OR "primary care*" ) ) OR ( MH ( "Nurses" OR "Physicians" OR "Medical Staff" OR “Allied Health Personnel”) ) AND

( ( ABS ( online OR education* OR internet OR e-learning OR web) ) OR ( TITLE ( online OR education* OR internet OR e-learning OR web) ) OR (MH ("Education, Distance") ) AND

( ( ABS (trial OR trials OR random* OR controlled OR RCT*) ) OR ( TITLE (trial OR trials OR random* OR controlled OR RCT*) ) OR (MH ("Random Allocation") ) AND

( ( ABS ( "guideline adherence" OR "reported change*" OR audit* OR screening OR "clinical behavior" OR "clinical behaviour*" OR "behavior* change*" OR "behaviour* change*" OR "clinical change*" OR "clinical practice" OR "nurse evaluation" OR "learning outcome*" OR "care skill*" OR knowledge ) ) OR ( TITLE ( "guideline adherence" OR "reported change*" OR audit* OR screening OR "clinical behavior" OR "clinical behaviour*" OR "behavior* change*" OR "behaviour* change*" OR "clinical change*" OR "clinical practice" OR "nurse evaluation" OR "learning outcome*" OR "care skill*" OR knowledge) ) OR (MH ( "Practice Guidelines as Topic"

OR "Guideline Adherence" OR "Nursing Audit" OR "Medical Audit" OR "Clinical Audit") ) ) )

***Scopus***

( ( ( ( ABS ( nurs* OR "healthcare professional*" OR "health professional*" OR physician* OR "healthcare provider" OR "consultant*" OR clinician* OR "allied health" OR "medical staff" OR "primary care*" ) ) OR ( TITLE ( nurs* OR "healthcare professional*" OR "health professional*" OR physician* OR "healthcare provider" OR clinician* OR "allied health" OR "medical staff" OR "consultant*" OR "primary care*" ) ) )

AND ( ( ABS ( online OR education* OR internet OR e-learning OR web ) ) OR ( TITLE ( online OR education* OR internet OR e-learning OR web ) ) ) AND

( ( ABS (trial OR trials OR random* OR controlled OR RCT* ) ) OR ( TITLE ( trial OR trials OR random* OR controlled OR RCT* ) ) ) AND

( ( ABS ( "guideline adherence" OR "reported change*" OR audit* OR screening OR "clinical behavior" OR "clinical behaviour*" OR "behavior* change*" OR "behaviour* change*" OR "clinical change*" OR "clinical practice" OR "nurse evaluation" OR "learning outcome*" OR "care skill*" OR knowledge ) ) OR ( TITLE ( "guideline adherence" OR "reported change*" OR audit* OR screening OR "clinical behavior" OR "clinical behaviour*" OR "behavior* change*" OR "behaviour* change*" OR "clinical change*" OR "clinical practice" OR "nurse evaluation" OR "learning outcome*" OR "care skill*" OR knowledge ) ) ) )

***Embase (Ovid)***

(nurs* OR "healthcare professional*" OR "health professional*" OR physician* OR "healthcare provider" OR "consultant*" OR clinician* OR "allied health" OR "medical staff" OR "primary care*").ab. OR (nurs* OR "healthcare professional*" OR "health professional*" OR physician* OR "healthcare provider" OR "consultant*" OR clinician* OR "allied health" OR "medical staff" OR "primary care*").ti. AND

(online OR education* OR internet OR e-learning OR web).ab OR (online OR education* OR internet OR e-learning OR web).ti. AND

(trial OR trials OR random* OR controlled OR RCT*).ab. OR (trial OR trials OR random* OR controlled OR RCT*).ti. AND

("guideline adherence" OR "reported change*" OR audit* OR screening OR "clinical behavior" OR "clinical behaviour*" OR "behavior* change*" OR "behaviour* change*" OR "clinical change*" OR "clinical practice" OR "nurse evaluation" OR "learning outcome*" OR "care skill*" OR knowledge).ab. OR ("guideline adherence" OR "reported change*" OR audit* OR screening OR "clinical behavior" OR "clinical behaviour*" OR "behavior* change*" OR "behaviour* change*" OR "clinical change*" OR "clinical practice" OR "nurse evaluation" OR "learning outcome*" OR "care skill*" OR knowledge).ti.

***Central Register of Controlled Trials***

( ( ( nurs* OR "healthcare professional*" OR "health professional*" OR physician* OR "healthcare provider" OR "consultant*" OR clinician* OR "allied health" OR "medical staff" OR "primary care*" ):ab) OR ( nurs* OR "healthcare professional*" OR "health professional*" OR physician* OR "healthcare provider" OR clinician* OR "allied health" OR "medical staff" OR "consultant*" OR "primary care*" ):ti )

AND ( ( online OR education* OR internet OR e-learning OR web ):ab) OR ( online OR education* OR internet OR e-learning OR web ):ti ) AND

( (trial OR trials OR random* OR controlled OR RCT* ):ab ) OR ( ( trial OR trials OR random* OR controlled OR RCT* ):ti ) AND

( ( "guideline adherence" OR "reported change*" OR audit* OR screening OR "clinical behavior" OR "clinical behaviour*" OR "behavior* change*" OR "behaviour* change*" OR "clinical change*" OR "clinical practice" OR "nurse evaluation" OR "learning outcome*" OR "care skill*" OR knowledge ):ab ) OR ( ( "guideline adherence" OR "reported change*" OR audit* OR screening OR "clinical behavior" OR "clinical behaviour*" OR "behavior* change*" OR "behaviour* change*" OR "clinical change*" OR "clinical practice" OR "nurse evaluation" OR "learning outcome*" OR "care skill*" OR knowledge ):ti ) )

***PsycINFO***

( ( ( ( AB ( nurs* OR "healthcare professional*" OR "health professional*" OR physician* OR "healthcare provider" OR "consultant*" OR clinician* OR "allied health" OR "medical staff" OR "primary care*" ) ) OR ( TI ( nurs* OR "healthcare professional*" OR "health professional*" OR physician* OR "healthcare provider" OR "consultant*" OR clinician* OR "allied health" OR "medical staff" OR "primary care*" ) ) ) OR (DE ( "Nurses" OR "Allied Health Personnel" OR "Physicians") ) AND

( ( AB ( online OR education* OR internet OR e-learning OR web ) ) OR ( TI (online OR education* OR internet OR e-learning OR web ) ) OR (DE ( "Electronic Learning" OR "Distance Education" ) ) AND

( ( AB (trial OR trials OR random* OR controlled OR RCT* ) ) OR ( TI ( trial OR trials OR random* OR controlled OR RCT* ) ) OR ( DE "Randomized Controlled Trials" ) )

AND

( ( AB ( "guideline adherence" OR "reported change*" OR audit* OR screening OR "clinical behavior" OR "clinical behaviour*" OR "behavior* change*" OR "behaviour* change*" OR "clinical change*" OR "clinical practice" OR "nurse evaluation" OR "learning outcome*" OR "care skill*" OR knowledge ) ) OR ( TI ( "guideline adherence" OR "reported change*" OR audit* OR screening OR "clinical behavior" OR "clinical behaviour*" OR "behavior* change*" OR "behaviour* change*" OR "clinical change*" OR "clinical practice" OR "nurse evaluation" OR "learning outcome*" OR "care skill*" OR knowledge ) ) OR (DE ("Evidence Based Practice" OR "Best Practices" OR "Clinical Audits") ) ) )
