## Supplementary Files 1-7 for "Effects of online professional learning on healthcare professionals’ knowledge and skill acquisition: A systematic review and meta-analysis": Supplementary File 7_Risk of Bias Summary Table.pdf

*Risk of Bias in Individual Studies (N = 171)*

| Author (Year) | Randomisation Process | Deviation from Intended Interventions | Missing Outcome Data | Measurement of Outcomes | Selection of the Reported Result | Overall Bias |
|---|---|---|---|---|---|---|
| Alvarado et al. 2023 | Low | Low | Low | Low | Some concerns | Some concerns |
| Arnold et al. 2013 | Low | High | High | Low | Some concerns | High |
| Badiei et al. 2016 | Some concerns | High | High | Low | Some concerns | High |
| Batchelor-Murphy et al. 2015 | Low | High | High | Low | Some concerns | High |
| Bell et al. 2000 | Some concerns | Low | Low | Some concerns | Some concerns | Some concerns |
| Bello et al. 2005 | Some concerns | Some concerns | Low | Low | Some concerns | Some concerns |
| Benda et al. 2020 | Some concerns | Some concerns | Low | Low | Some concerns | Some concerns |
| Benjamin et al. 2008 | Low | Some concerns | Low | Low | Some concerns | Some concerns |
| Bhattacharyya et al. 2021 | Low | Low | Low | Low | Some concerns | Some concerns |
| Bochicchio et al. 2006 | High | Some concerns | Low | Low | Some concerns | High |
| Bonabi et al. 2019 | Some concerns | Some concerns | High | Low | Some concerns | High |
| Brateanu et al. 2019 | Some concerns | Low | Low | Low | Some concerns | Some concerns |
| Bredesen et al. 2016 | Low | Some concerns | Some concerns | Low | Some concerns | Some concerns |
| Carrick et al. 2017 | Some concerns | Some concerns | Low | Low | Low | Some concerns |
| Carstensen et al. 2023 | Low | High | Low | Low | Some concerns | High |
| Chaisriya et al. 2024 | Low | Low | Low | Low | Some concerns | Some concerns |
| Chang et al. 2024 | Some concerns | High | Some concerns | Some concerns | Some concerns | High |
| Chenkin et al. 2008 | Some concerns | Low | Low | Low | Some concerns | Some concerns |
| Chisholm et al. 2025 | Low | Some concerns | Low | Low | Low | Some concerns |
| Chuang et al. 2022 | Some concerns | High | Some concerns | Low | Some concerns | High |
| Clancy et al. 2016 | High | Some concerns | High | Low | Some concerns | High |
| Connolly et al. 2014 | Some concerns | Some concerns | Some concerns | High | Some concerns | High |
| Cullinan et al. 2017 | Some concerns | High | Some concerns | Low | Some concerns | High |
| Demir et al. 2024 | Some concerns | High | Some concerns | Some concerns | Some concerns | High |
| Diehl et al. 2017 | High | High | High | Low | Some concerns | High |

| Author (Year) | Randomisation Process | Deviation from Intended Interventions | Missing Outcome Data | Measurement of Outcomes | Selection of the Reported Result | Overall Bias |
|---|---|---|---|---|---|---|
| Dimeff et al. 2009 | Some concerns | Low | Low | High | Some concerns | High |
| Edrich et al. 2016 | Some concerns | Some concerns | Low | Some concerns | Some concerns | Some concerns |
| Ehrenreich-May et al. 2016 | Some concerns | Low | Low | Some concerns | Some concerns | Some concerns |
| El-Helou et al. 2025 | Low | Some concerns | Some concerns | Low | Some concerns | Some concerns |
| Elgie et al. 2010 | Some concerns | Some concerns | High | Low | Some concerns | High |
| Elzeky et al. 2022 | Low | Some concerns | Low | Low | Some concerns | Some concerns |
| Farokhzadian et al. 2023 | Low | Low | Low | Low | Some concerns | Some concerns |
| Fordis et al. 2005 | Low | High | High | Low | Some concerns | High |
| Gallegos et al. 2021 | Some concerns | Some concerns | Low | Some concerns | Some concerns | Some concerns |
| Garcia-Rodriguez et al. 2016 | Some concerns | Some concerns | Low | Low | Some concerns | Some concerns |
| Gerbert et al. 2002 | Some concerns | Some concerns | Low | Some concerns | Some concerns | Some concerns |
| Gordon et al. 2011 | Low | Some concerns | Some concerns | Low | Some concerns | Some concerns |
| Green et al. 2020 | Some concerns | Some concerns | Some concerns | Some concerns | Some concerns | Some concerns |
| Guden et al. 2024 | Low | High | High | Some concerns | Some concerns | High |
| Halms et al. 2024 | Low | Some concerns | Low | Some concerns | Low | Some concerns |
| Haney et al. 2012 | High | Some concerns | Low | Low | Some concerns | High |
| Harris et al. 2008 | Some concerns | High | Some concerns | Some concerns | Some concerns | High |
| Harris et al. 2013 | Some concerns | Some concerns | Some concerns | Low | Some concerns | Some concerns |
| Hayer et al. 2022 | Some concerns | Some concerns | Low | Low | Some concerns | Some concerns |
| Heard et al. 2017 | Low | High | Some concerns | Low | Some concerns | High |
| Heidarian et al. 2025 | High | Low | Some concerns | Low | Some concerns | High |
| Horiuchi et al. 2009 | Low | High | High | Low | Some concerns | High |
| Houwink et al. 2014 | Some concerns | Some concerns | Some concerns | Low | Some concerns | Some concerns |
| Hoysted et al. 2019 | Low | Low | Low | Low | Some concerns | Some concerns |
| Hsieh et al. 2021 | Low | Some concerns | Low | Low | Some concerns | Some concerns |
| Huang et al. 2019 | Some concerns | Some concerns | Low | Some concerns | Some concerns | Some concerns |
| Huang et al. 2021 | Low | Low | Low | Low | Some concerns | Some concerns |
| Hugenholtz et al 2008 | High | Some concerns | Low | Some concerns | Some concerns | Some concerns |
| Hurtado et al. 2023 | High | High | Some concerns | Low | Some concerns | High |
| Hussein et al. 2022 | Low | High | Low | Low | Some concerns | High |

| Author (Year) | Randomisation Process | Deviation from Intended Interventions | Missing Outcome Data | Measurement of Outcomes | Selection of the Reported Result | Overall Bias |
|---|---|---|---|---|---|---|
| Jain et al. 2010 | Low | Some concerns | Low | Low | Some concerns | Some concerns |
| Jalali et al. 2022 | High | High | Low | Low | Some concerns | High |
| Jaunay et al. 2019 | Low | High | Some concerns | Low | Low | High |
| Jensen et al. 2009 | High | Low | Some concerns | Low | Some concerns | High |
| Johannsen et al. 2023 | Some concerns | Some concerns | Low | Some concerns | Low | Some concerns |
| Kang et al. 2022 | Low | High | Low | Low | Some concerns | High |
| Karvinen & Reed 2022 | Low | Low | High | Some concerns | Some concerns | High |
| Karvinen et al. 2017 | Low | Some concerns | Low | Low | Some concerns | Some concerns |
| Kato et al. 2017 | Low | Low | Low | Low | Some concerns | Some concerns |
| Keleekai et al. 2016 | Some concerns | High | Low | Low | Some concerns | High |
| Kemper et al. 2002 | Some concerns | Low | Low | Low | Some concerns | Some concerns |
| Kerfoot et al. 2009 | Some concerns | Some concerns | Some concerns | Some concerns | Some concerns | Some concerns |
| Khoshbaten et al. 2014 | Some concerns | Some concerns | Low | Low | Some concerns | Some concerns |
| KhoshnoodiFar et al. 2019 | Some concerns | Some concerns | Some concerns | Low | Some concerns | Some concerns |
| Kim & Hwang, 2022 | Some concerns | Low | Low | Low | Some concerns | Some concerns |
| Kim & Shin. 2013 | Some concerns | Some concerns | Low | Low | Some concerns | Some concerns |
| Kim et al. 2023 | Low | Low | Low | Low | Some concerns | Some concerns |
| Kimemia, J. 2018 | Low | Some concerns | Low | Low | Some concerns | Some concerns |
| Koeppen et al. 2024 | Low | Low | Some concerns | Some concerns | Low | Some concerns |
| Krishnamachari et al. 2018 | Some concerns | Low | Low | Low | Some concerns | Some concerns |
| Le et al. 2010 | High | High | High | Some concerns | Some concerns | High |
| Lee & Choi 2024 | High | High | Some concerns | Low | Some concerns | High |
| Leiva-Fernández et al. 2020 | Some concerns | Low | Low | Some concerns | Some concerns | Some concerns |
| Lhbibani et al. 2022 | Some concerns | Some concerns | Low | Low | Some concerns | Some concerns |
| Li A. 2022 | Some concerns | High | High | Some concerns | Some concerns | High |
| Liaw et al. 2015 | Some concerns | Low | Some concerns | Low | Some concerns | Some concerns |
| Liaw et al. 2016 | Some concerns | Low | Some concerns | Low | Some concerns | Some concerns |
| Liaw et al. 2017 | Low | Some concerns | Low | Low | Some concerns | Some concerns |
| Liu et al. 2020 | Low | Low | Low | Low | Some concerns | Some concerns |
| Los et al. 2023 | Some concerns | High | High | Some concerns | Some concerns | High |

| Author (Year) | Randomisation Process | Deviation from Intended Interventions | Missing Outcome Data | Measurement of Outcomes | Selection of the Reported Result | Overall Bias |
|---|---|---|---|---|---|---|
| Lyon et al. 2022 | Some concerns | High | Low | Low | High | High |
| Maertens et al. 2017 | Low | High | Some concerns | Low | Some concerns | High |
| Mallon et al. 2022 | Some concerns | Low | Some concerns | Low | Some concerns | Some concerns |
| Maloney et al. 2011 | Low | High | High | Some concerns | Some concerns | Some concerns |
| Marriott et al. 2023 | Some concerns | Low | Low | Some concerns | Some concerns | Some concerns |
| Maruyama et al. 2022 | Low | Low | Some concerns | Low | Some concerns | Some concerns |
| Maruyama et al., 2022 | High | Low | High | Low | Some concerns | High |
| McCrow et al. 2014 | High | Low | Low | Low | Some concerns | High |
| Mesquita et al. 2019 | Some concerns | Low | Low | Some concerns | Some concerns | Some concerns |
| Mohan et al. 2018 | Low | Low | Low | Low | Some concerns | Some concerns |
| Monti & Perreault 2020 | Low | Some concerns | Low | Low | Some concerns | Some concerns |
| Mun and Hwang 2020 | Low | Low | Low | Low | Some concerns | Some concerns |
| Murray et al. 2010 | Low | High | High | Low | High | High |
| NajafiGhezeljeh et al. 2019 | Some concerns | Some concerns | Low | Low | Some concerns | Some concerns |
| Nakamura et al. 2022 | Low | Low | High | Low | Some concerns | High |
| Ng et al. 2022 | Some concerns | Some concerns | High | Low | Some concerns | High |
| Nixon et al. 2019 | Some concerns | Low | Low | Low | Some concerns | Some concerns |
| Nyberg et al. 2022 | Low | Low | Low | Low | Some concerns | Some concerns |
| Ozawa et al. 2022 | Some concerns | High | Some concerns | High | Some concerns | High |
| Padalino et al. 2007 | High | Some concerns | Low | Some concerns | High | High |
| Pahud et al. 2020 | Some concerns | Some concerns | High | Low | Some concerns | High |
| Passos et al. 2022 | Some concerns | High | High | Low | Some concerns | High |
| Peabody et al. 2017 | Some concerns | High | High | Low | Some concerns | High |
| Pelayo et al. 2011 | Low | Low | Low | Low | Some concerns | Some concerns |
| Pelayo-Alvarez et al. 2013 | Low | Some concerns | Some concerns | Low | High | High |
| Perkins et al. 2012 | Low | Low | Low | Some concerns | Some concerns | Some concerns |
| Petit et al. 2019 | Some concerns | Low | Low | Low | Some concerns | Some concerns |
| Philbrook et al. 2010 | Some concerns | High | Low | High | Some concerns | High |
| Phuangngoenmak et al. 2019 | Some concerns | Some concerns | Low | Low | Some concerns | Some concerns |
| Platz et al. 2010 | High | High | High | Some concerns | Some concerns | High |

| Author (Year) | Randomisation Process | Deviation from Intended Interventions | Missing Outcome Data | Measurement of Outcomes | Selection of the Reported Result | Overall Bias |
|---|---|---|---|---|---|---|
| Richmond et al. 2016 | Low | Some concerns | Low | Low | Some concerns | Some concerns |
| Robinson et al. 2018 | Some concerns | Some concerns | Low | Low | Some concerns | Some concerns |
| Sabaghnejad et al. 2024 | High | Some concerns | Some concerns | Some concerns | Some concerns | High |
| Sadegh Madani et al. 2023 | Some concerns | Some concerns | Some concerns | Low | Some concerns | Some concerns |
| Saunders et al. 2016 | High | Low | Low | Low | Some concerns | High |
| Scheetz et al. 2019 | Some concerns | Some concerns | High | Low | Some concerns | High |
| Schulman et al. 2012 | Some concerns | Some concerns | High | Low | Some concerns | High |
| Schumacher 2009 | Low | High | Some concerns | Low | Some concerns | High |
| Serwetnyk et al. 2013 | Some concerns | Some concerns | Some concerns | High | High | High |
| Sheen et al. 2008 | Some concerns | High | High | High | Some concerns | High |
| Siebert et al. 2022 | Low | Low | Low | Some concerns | Some concerns | Some concerns |
| Silab et al. 2024 | Low | Low | Low | Low | Some concerns | Some concerns |
| Silva et al. 2016 | Some concerns | High | High | Low | Some concerns | High |
| Simonsen et al. 2014 | Some concerns | Some concerns | Some concerns | Low | Some concerns | Some concerns |
| Smeekens et al. 2011 | Low | Low | Low | Low | Some concerns | Some concerns |
| Snippen et al. 2023 | High | Low | Low | Low | Some concerns | High |
| Soon et al. 2020 | Some concerns | Some concerns | Some concerns | Low | Some concerns | Some concerns |
| Soper 2017 | Some concerns | Some concerns | Low | Low | Some concerns | Some concerns |
| Sperl-Hillen et al. 2014 | Low | High | High | Low | Some concerns | High |
| Stacey et al. 2006 | Low | Some concerns | Low | Low | Some concerns | Some concerns |
| Steele et al. 2013 | Some concerns | High | High | Low | High | High |
| Stewart et al. 2005 | High | Some concerns | Some concerns | Low | Some concerns | High |
| Takeuchi et al. 2023 | Low | High | High | Low | Some concerns | High |
| Tang et al. 2024 | Low | High | High | Low | Some concerns | High |
| Tassi, A. 2017 | Low | Some concerns | High | Low | Some concerns | High |
| Taylor et al. 2013 | Low | High | High | High | Some concerns | High |
| Taylor et al. 2020 | Some concerns | High | High | Low | Some concerns | High |
| Thai et al. 2020 | Some concerns | Some concerns | Low | Low | Some concerns | High |
| Thompson et al. 2016 | Some concerns | Low | Low | Low | Some concerns | Some concerns |
| Toole et al. 2013 | Low | High | High | Low | Some concerns | High |

| Author (Year) | Randomisation Process | Deviation from Intended Interventions | Missing Outcome Data | Measurement of Outcomes | Selection of the Reported Result | Overall Bias |
|---|---|---|---|---|---|---|
| Triola et al. 2006 | Some concerns | High | High | Low | Some concerns | High |
| Van Stiphout et al. 2018 | Low | Some concerns | Low | Low | Some concerns | Some concerns |
| Velasco et al. 2015 | Some concerns | High | High | Low | Some concerns | High |
| Viguier et al. 2015 | Low | Low | High | Low | Some concerns | High |
| Vollmar et al. 2010 | Low | Some concerns | Some concerns | Low | Low | Some concerns |
| Wadey et al. 2022 | Some concerns | Low | Some concerns | Some concerns | Some concerns | Some concerns |
| Wakefield et al. 2014 | Some concerns | Some concerns | High | Low | Some concerns | High |
| Walker et al. 2020 | Some concerns | Some concerns | Low | Low | Some concerns | Some concerns |
| Wang et al. 2017 | Some concerns | Some concerns | Low | Low | Some concerns | Some concerns |
| Wang et al. 2024 | Some concerns | Some concerns | Some concerns | High | High | High |
| Wanty et al. 2023 | Some concerns | Some concerns | High | Low | Some concerns | High |
| Weaver et al. 2016 | Some concerns | Some concerns | Low | Low | Some concerns | High |
| Wee et al. 2016 | Low | Low | Some concerns | Low | Some concerns | Some concerns |
| Welch et al. 2014 | Some concerns | Some concerns | Some concerns | Low | Some concerns | Some concerns |
| Westmoreland et al. 2010 | Some concerns | High | High | Low | Some concerns | High |
| Wilkes et al. 2017 | Some concerns | High | High | Low | Some concerns | High |
| Williams et al. 2021 | Some concerns | High | High | Low | Some concerns | High |
| Worm 2013 | Some concerns | Some concerns | Low | Low | Some concerns | Some concerns |
| Wright 2008 | High | High | High | Low | Some concerns | High |
| Yao et al. 2015 | High | High | High | Low | Some concerns | High |
| Yoo et al. 2019 | Some concerns | High | High | Low | Some concerns | High |
| Zarifsanaiey et al. 2022 | Low | Low | Low | Low | Some concerns | Some concerns |
| Zhang et al. 2024 | Low | Some concerns | Low | Low | Some concerns | Some concerns |
| Zhao et al. 2024 | Low | Some concerns | Low | Low | Some concerns | Some concerns |
| das Graças Silva Matsubara et al. 2016 | Some concerns | High | Low | Low | Some concerns | High |
| de Gannes et al. 2004 | Some concerns | High | High | Low | Some concerns | High |
