## Supplementary figures and images for "Effects of online professional learning on healthcare professionals’ knowledge and skill acquisition: A systematic review and meta-analysis"

### Supplementary File 5 learning_combined_design_moderator_plot_control.pdf

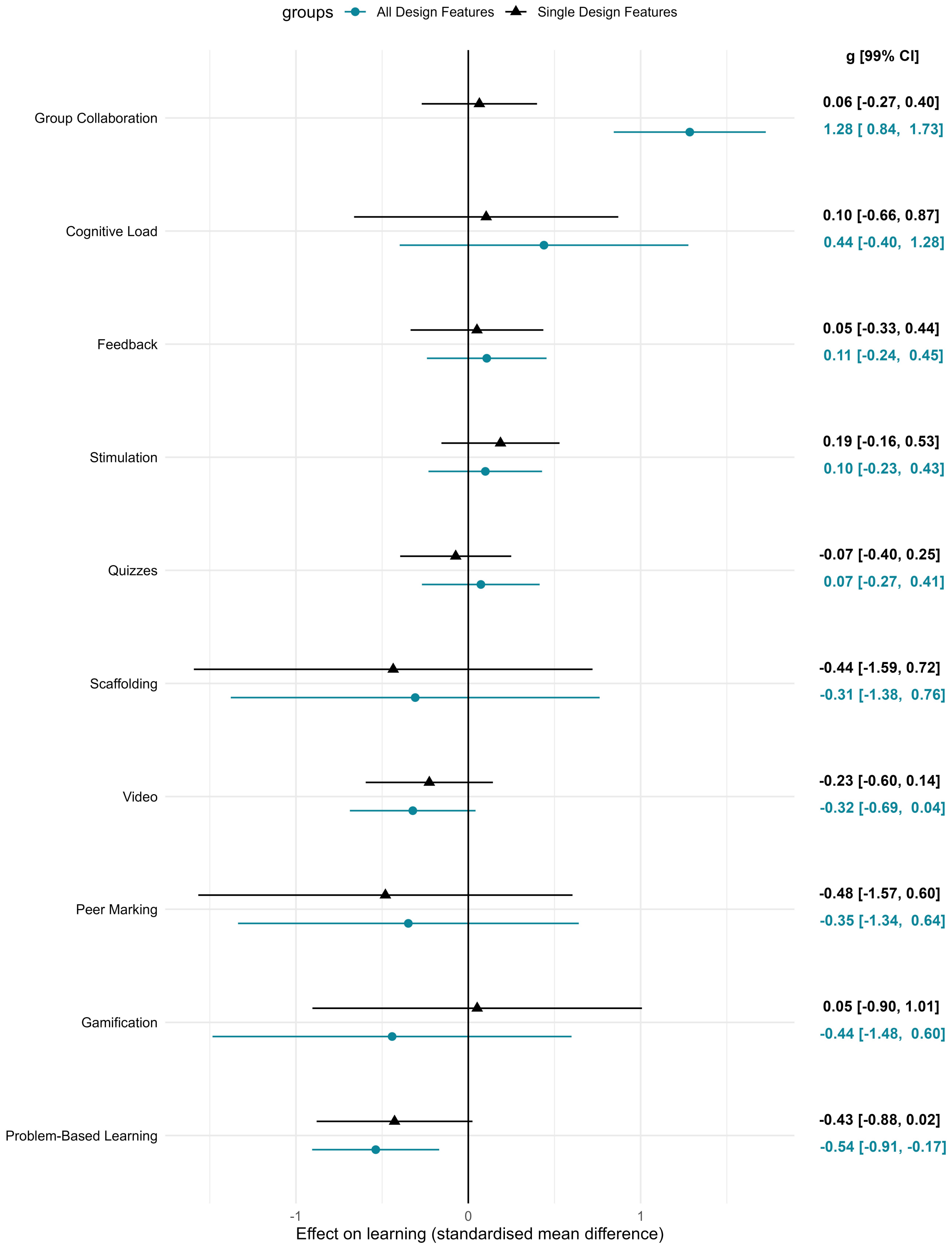

### Supplementary File 6 learning_combined_design_moderator_plot_f2f.pdf

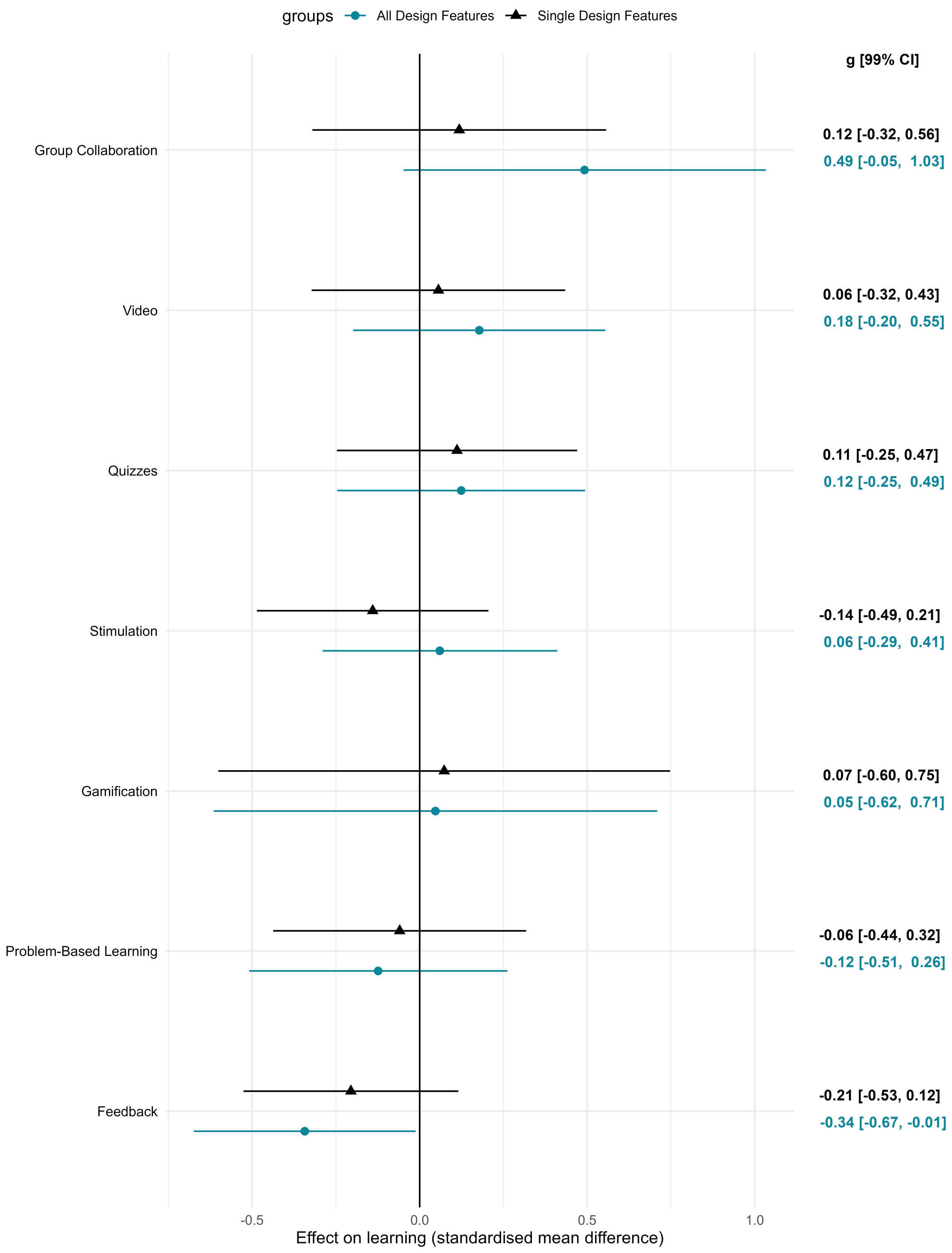
